# Risk of new diagnoses and exacerbations of chronic conditions after SARS-CoV-2 infection: a systematic review update

**DOI:** 10.1101/2025.05.13.25326692

**Authors:** Lindsay A. Gaudet, Jennifer Pillay, Dianne Zakaria, Sabrina Saba, Ben Vandermeer, Maria Tan, Lisa Hartling

**Affiliations:** Alberta Research Center for Health Evidence, University of Alberta, Edmonton, Alberta, Canada; LifespanChronic Diseases and Conditions Division, Centre for Surveillance and Applied Research, Health Promotion and Chronic Disease Prevention Branch, Public Health Agency of Canada, Ottawa, Ontario, Canada; Epidemiology Coordinating and Research Centre, Faculty of Medicine & Dentistry, University of Alberta, Edmonton, Alberta, Canada

**Author notes:** Corresponding author: Lisa Hartling, PhD, Professor, Department of Pediatrics, Director, Alberta Research Centre for Health Evidence University of Alberta, 4-472 Edmonton Clinic Health Academy 11405-87 Avenue, Edmonton, AB, T6G 1C9.

**Keywords:** chronic disease, COVID-19, Disease Progression, SARS-CoV-2, Symptom Flare Up, systematic review

## Abstract

The large number of individuals infected by severe acute respiratory syndrome-coronavirus-2 (SARS-CoV-2) necessitates estimation of the future healthcare burdens. We updated a systematic review examining associations between SARS-CoV-2 infection and the incidence of new diagnoses and exacerbations of chronic conditions. Updated searches were run September 4, 2024 in MEDLINE and EMBASE for observational studies utilizing a control group, addressing confounding by sex and comorbidities, and reporting age-stratified data for ≥1 chronic condition category (n=12) or condition type (n=46) of interest. Two human reviewers screened 50% of identified titles/abstracts, after which DistillerAI acted as second reviewer. Two human reviewers assessed full-texts of relevant studies for eligibility based on *a priori* criteria. One reviewer extracted data and assessed risk of bias; a second reviewer verified results data and risk of bias assessments. Pooled hazard ratios (HRs) were estimated with inverse-variance weighting, and two reviewers used GRADE to assess certainty in our conclusions of little-to-no association (i.e., HR 0.75 to 1.25), small-to-moderate association (i.e., HR 0.51 to 0.74 or 1.26 to 1.99), or large association (i.e., HR ≤0.50 or ≥2.00). We identified 46 new studies and brought forward 23 studies from the original review. After SARS-CoV-2 infection, there is probably increased risk of new diagnoses for several chronic conditions, especially in adults (18-64 years) and older adults (≥65 years). Most findings are based on data from earlier pandemic periods; their relevance to contemporary populations is uncertain due to differences in vaccination rates, implemented public health measures, and circulating variants of concern.

## INTRODUCTION

Since its emergence in December of 2019, the severe acute respiratory syndrome corona virus-2 (SARS-CoV-2) has infected hundreds of millions of people worldwide in a global pandemic. Early in the pandemic, the potential long-term consequences of SARS-CoV-2 infection were raised,^1^ leading to concerns about the future burdens of SARS-CoV-2 infection. Evaluations of whether SARS-CoV-2 infection may lead to increased risk of long-term sequelae is important to understanding how SARS-CoV-2 may impact the future burden of health outcomes on populations and healthcare resources.

We previously conducted a systematic review examining new diagnoses and exacerbations of a range of chronic conditions after SARS-CoV-2 infection.^2^ From studies conducted early in the pandemic (i.e., 2020-2022), we observed increased risks of new diagnoses for some chronic conditions in some age groups (e.g., cardiovascular conditions in adults ≥65 years old). However, we found few studies reporting on individuals <18 years old and individuals with severe SARS-CoV-2 infection (i.e., hospitalized during the acute phase of infection). In light of evolving contextual factors such as increased vaccination coverage, new variants of predominance, and easing of public health measures, and with the hopes of filling gaps identified by our previous synthesis, we undertook an update of our review to provide more current evidence on the associations between SARS-CoV-2 infection and subsequent new diagnoses or exacerbations of chronic conditions to support informed policy and decision making for the current SARS-CoV-2 context.

## OBJECTIVE

Our objective was to update our previous review examining associations between SARS-CoV-2 infection and the incidence of new diagnoses or exacerbations of chronic conditions, stratified by age and severity of infection.

## METHODS

The eligibility criteria and methods for this update were defined *a priori* with input from disease surveillance experts at the Public Health Agency of Canada. The protocol was prospectively registered on PROSPERO (CRD42024585278). This review has been reported according to the Preferred Reporting Items for Systematic reviews and Meta-Analyses (PRISMA) statement.^3^

### Study eligibility

We included prospective or retrospective observational studies carried out in high-income Organisation for Economic Co-operation and Development member countries, comparing individuals with suspected or confirmed SARS-CoV-2 infection (exposed) to those without (controls), and published in English or French. To be eligible, studies had to report on severity of SARS-CoV-2 infection (e.g., hospitalization status), account for potential confounding by at least sex and two or more comorbidities (e.g., by matching, using propensity scores, or multivariable regression), and report outcomes of interest stratified by age groups to allow for analysis and summarization of findings by key life-course stages: <18 years old, 18-64 years old, and ≥65 years old. Age groups similar to these (i.e., within ∼3 years of the 18-year cut-off and ∼5 years for the 65-year cut-off) were included and analyzed within the most appropriate age group. We included studies of people with either confirmed (e.g., by laboratory testing) or suspected (e.g., physician diagnosed) SARS-CoV-2 infection. We did not require control groups to test negative for SARS-CoV-2 and hospitalized patients without SARS-CoV-2 or individuals with other respiratory infections (e.g., influenza) could serve as controls.

Before study screening was concluded, we obtained approval for a protocol deviation for studies reporting on exacerbations of pre-existing chronic conditions to waive the eligibility criteria requiring adjustment for comorbidities. In place of adjustment for two or more comorbidities, in these studies we required authors to account for, at minimum, sex and two potential confounders related to either SARS-CoV-2 infection or disease status of the chronic condition under investigation.

### Conditions of interest

At the protocol stage, categories of conditions of interest included: autoimmune conditions, cardiovascular diseases, chronic kidney disease, diabetes, inflammatory bowel diseases, mental illness, musculoskeletal disorders, neurological conditions, respiratory diseases, and stroke. Additional categories for fibromyalgia and sleep disorder were created during data extraction; fibromyalgia and sleep disorders were originally flagged as conditions of interest under the neurological and mental illness conditions categories, respectively; the decision to create new categories to enhance interpretation of the findings was made in consultation with disease surveillance experts.

Because of lack of evidence for association in the literature, cancers were dropped from the conditions of interest. Similarly, within the category of mental illness, alcohol and substance use disorders were deemed no longer of interest. Lastly, we deprioritized outcome data that were either: i) a composite of cardiovascular disorders and stroke; or ii) a composite of new diagnoses and exacerbations within the same category or condition type, as sufficient data on relevant individual conditions/outcomes were identified.

We also consulted with disease surveillance experts at the Public Health Agency of Canada to chart out individual conditions (“condition types”) reported by included studies within each category (e.g., dementia within the category of neurological conditions). Because of the limited clinical relevance, we did not synthesize dementia/cognitive impairment outcomes in individuals <18 years old. We attempted to only include studies reporting on diagnoses of chronic conditions, which we defined as a new occurrence of disease documented by a healthcare provider in medical records; however, there may not have been standard diagnostic testing performed in all cases.

### Search Strategy and Study Selection

Searches developed by an information specialist were run in Medline and Embase on September 4, 2024. Searches combined concepts for SARS-CoV-2 infection, post-acute/follow-up, review outcomes (e.g., incidence), and categories and conditions of interest; complete search strategies are available in Appendix A. Searches for conditions included in the original review were limited to records published on or after July 1, 2022. Searches for newly added conditions of interest (i.e., autoimmune conditions and inflammatory bowel diseases) were limited to records published on or after January 1, 2020, in line with the previous review. Duplicate records were removed in Endnote (v. 20.3, Clarivate Analytics, Philadelphia, PA) and unique records uploaded to DistillerSR (Evidence Partners, Ottawa, Canada) for screening. We also screened the references of included studies and relevant systematic reviews for potentially eligible studies; the supplemental searching was completed on September 20, 2024.

Unique records identified by the search strategies were reviewed in a two-stage process, first by title and abstract (screening) and then by full-text (selection). During screening, DistillerSR’s machine learning feature, DistillerAI, was enabled. DistillerAI learns from human reviewers’ inclusion decisions to assign a likelihood score (0 to 1, with values closer to 1 indicating higher likelihood of inclusion) to each unscreened record and prioritizes the most relevant records for screening by the human reviewers (i.e., the most relevant records are screened first).^4^ When a threshold likelihood score for inclusion is applied, DistillerAI can also act as a second reviewer with high sensitivity and specificity.^5, 6^ Thus, two human reviewers independently screened the first 50% of records, after which DistillerAI acted as the second reviewer for the remaining records using a likelihood threshold of 0.7 (the lowest score assigned to a study included by a human reviewer).

Using standardized forms, four human reviewers piloted the eligibility criteria and screening form on 200 titles and abstracts. In order to efficiently train the DistillerAI tool to identify relevant records, the 200 titles and abstracts used for piloting consisted of a purposefully selected sample of 100 records from the previous review and a random sample of 100 new records. The selection form, for use with full-text records, was piloted using studies identified as potentially eligible from the 100 new records in the screening pilot (n=13). The purposefully selected records from the previous review used for piloting and AI training are not counted within the PRISMA flow diagram.

After screening, attempts were made to retrieve the full texts of all potentially relevant records. Two human reviewers independently reviewed all retrieved full-texts and came to consensus, with adjudication by a review lead or other reviewer (e.g., statistician) when necessary.

### Data Extraction

We used structured forms developed in Microsoft Excel (v. 2019, Microsoft Corporation, Redmond, WA) for the previous review for data extraction and risk of bias (ROB) assessments. No changes to the forms were required and reviewers involved in this update piloted both forms during the initial review; therefore, additional piloting was not undertaken. One reviewer extracted study characteristics and results; a second reviewer verified the results data for accuracy and completeness. Findings from the most adjusted model available were prioritized in all cases. We also revisited previously included studies to ensure data on new conditions of interest were extracted.

Studies with overlapping populations were not automatically excluded, and we used data for any outcomes/strata that did not overlap between studies. When one or more studies reported similar outcomes for the same population with significant overlap in time period, we prioritized data from the study reporting the outcome most applicable to our review. If studies reported on outcomes of similar relevance, we took data with the longer follow-up time, or longest enrolment period when duration of follow-up was similar.

### Risk of Bias

For ROB, one reviewer assessed each study and another reviewer verified their assessments. We used the Joanna Briggs Institute (JBI) critical appraisal checklist for cohort studies.^7^ We specifically considered in our assessment the validity of SARS-CoV-2 infection confirmation. Studies restricting their COVID-19 cases to confirmed infections (i.e., detection of SARS-CoV-2 via RT-PCR or antigen test, including use of ICD-10 code U07.1 - COVID-19, virus identified) were considered at lower ROB than those allowing for suspected cases. We also had concerns when a study did not censor control participants who may have contracted SARS-CoV-2 during the follow-up period. Additionally, we had concerns about ROB in outcome ascertainment when >20% of participants were followed-up for <6 months (or <12 months for studies reporting on autoimmune disorders and mental illness, due to the complexity in diagnosing these conditions). We assigned an overall ROB rating (low, moderate or high) based on the number of questions answered “No” for each study (0 for low, 1 for moderate, 2 for high). Final ROB assessments were incorporated into our certainty of evidence assessments guided by the Grading of Recommendations, Assessment, Development and Evaluation (GRADE) approach (see below).^8^

### Synthesis

We estimated random-effects pooled hazard ratios (HRs) and generated forest plots for each category of chronic conditions and condition types within categories, by age category and SARS-CoV-2 infection care setting (inpatient vs. outpatient/mixed) using inverse variance weighting in Review Manager (RevMan; v5.4, The Cochrane Collaboration, 2020). Since new diagnosis of a condition can only occur once, we considered HRs and incidence rate ratios (IRRs) interchangeable. If a HR or IRR was not available, we also considered risk ratios and, when cumulative incidence was <10%, odds ratios. Data not appropriate for meta-analysis were synthesized descriptively.

For studies reporting data for multiple diagnoses within a condition type (e.g., tachycardia and ventricular arrhythmia would both fall under the condition type “arrhythmias/dysrhythmias”), we estimated an average weighted by the inverse of the variance to give more weight to results with more precise estimates. We also used this process when a study reported multiple individual conditions within a category if the study did not also report a suitable composite outcome for the category. When a study-reported age strata extended past the review age category cut-off (e.g., to include data from a 50-69 year age group in the 18-64 year age category), the study was reweighted by dividing the standard error by the square root of the proportion of the study age stratum falling within the review age category (e.g., standard error/((15/20)^0.5^) for the provided example).

For all categories and condition types with at least low certainty of some association (i.e., small-to-moderate: HR 0.51 to 0.74 or 1.26 to 1.99; or large: HR ≤ 0.50 or ≥ 2.00), we estimated the excess incidence (and 95% confidence intervals) per 1000 people over 6 months by multiplying the pooled estimate (and 95% confidence limits) of the adjusted relative effect by an estimate of the control (non-SARS-CoV-2 infected) event rate. We used a hierarchy to identify the most relevant data to use for the control event rate. If at least one study reported a composite outcome (e.g., any cardiovascular event) within a category, we used that study’s reported control group incidence rate for that condition category. When a category had no directly reported composite incidence, we looked at the individual conditions in that category. Where conditions within a category are linked or sometimes co-occur, the condition with the highest incidence stood as a conservative estimate for the category. When two or more conditions within a category were considered by disease surveillance experts not to generally co-occur (e.g., myoneural junction/muscle disease and Parkinson’s disease), we summed their incidences as an estimate of the control event rate. When multiple studies reported a control event rate for a category or condition, we took an average weighted by sample size. We converted all control event rates to a standard 6-month period, which was most representative of follow-up duration in the included studies, e.g., a 1-year incidence rate was halved to estimate the incidence over 6 months.

We planned to explore statistical and clinical (e.g., clinical definitions, populations, etc.) heterogeneity through stratified secondary analyses based on:

1. confirmed SARS-CoV-2 exposure (>90% of cases identified by real-time polymerase chain reaction or antigen test results, including use of ICD-10 code U07.1) vs. others;
2. short-term (1 to <12 months after SARS-CoV-2 infection) vs. longer-term (≥12 months after SARS-CoV-2 infection) maximum follow-up;
3. predominant SARS-CoV-2 variant (Omicron lineage vs. others).

Stratified analyses were limited to outcomes with ≥2 studies in each group; subgroup differences were assessed using the Chi^2^ test in RevMan. When SARS-CoV-2 variant was not reported, we categorized studies based on the dominant SARS-CoV-2 variant in that country during the study’s reported time period of SARS-CoV-2 infections. We referred to Our World in Data for country-level data on variants of predominance and assigned start of the Omicron period for that country to the first day that Omicron was reported to account for ≥80% of cases.^9^

Lastly, we narratively summarized any within-study time-varying effects or variant-based sub-group analyses reported by included studies, regardless of whether these comparisons were stratified by age.

### Certainty Assessments using GRADE

To assess certainty in the evidence around new diagnoses, two reviewers reached consensus through discussion in relation to our thresholds of effect for the relative effects for each outcome, guided by GRADE.^8, 10^ Because this review examines prognosis after SARS-CoV-2 infection (i.e. association not causation), we started the evidence at high certainty^11^ and rated down to moderate, low, or very low certainty based on five domains: within-study ROB, indirectness, inconsistency, imprecision, and reporting biases. We rated down by 0, 1 or 2 levels depending on the seriousness of the concerns in each domain, i.e., how much the domain appeared to impact the conclusions. Additionally, we did not rate down if some of our concerns in one domain likely stemmed from issues in another domain; for example, we did not rate down for inconsistency if differences in estimates across studies appeared to be related to ROB (for which we rated down). For all new diagnosis outcomes, we considered a relative effect of 0.75-1.25 as little-to-no association, 0.51-0.74 and 1.26-1.99 as small-to-moderate, and ≤0.50 or ≥2.00 as large. We rated the conclusions for which we had the greatest certainty. For example, if we had concerns in a domain about a large association, but did not have concerns about a small-to-moderate association, we assessed our certainty in a small-to-moderate association. In this report, we use language consistent with GRADE, i.e., “there is”, “there is probably”, and “there may be”, to indicate high, moderate, and low certainty, respectively, in statements drawing conclusions about the findings.^12^

To assess concerns due to ROB, we rated down when ≥50% of weight came from studies assessed at high ROB and our conclusions changed about an association when high ROB studies were removed from the pooled estimate. We did not rate down when the point estimates of all studies showed the same direction of association or when removing high ROB studies did not change our conclusions about the association.

We considered rating down for inconsistency when: there was concerning variation in study estimates around our thresholds (i.e., when roughly ≥20% or ≥40% of an estimate’s weight came from studies with point estimates substantially outside our thresholds, for serious or very serious concerns, respectively) that was not explained by subgroup analysis or other domains; the weight of studies outside of the threshold about which we drew conclusions was near to but <20% (or <40%, for very serious concerns) and the studies outside that threshold showed associations in the opposite direction (or large associations in either direction for outcomes otherwise showing little-to-no difference); or when an estimate based on a single study substantially disagreed with other estimates for the same category or condition type. For studies reporting multiple age groups within this reviews’ age strata of interest, concerns about inconsistency compared with other studies in an analysis were based on a within-study average across reported age groups. In studies reporting on distinct geographic populations (i.e., different countries), these were considered separately due to assumed differences in public health measures, health systems, and available data, etc.

For indirectness, we primarily considered rating down when ≥50% of the estimate weight came from pre-Omicron populations, or from one or two specific diagnoses (when applicable). In outcomes showing an association, we had concerns about applicability to a contemporary population (i.e., highly vaccinated and exposed primarily to Omicron variant SARS-CoV-2) of findings from studies conducted early in the pandemic. Therefore, we rated down when the majority (≥50%) of the weight contributing to an estimate was from pre-Omicron data and there was no evidence, either from sensitivity analyses of Omicron-specific data or within-study variant comparisons, indicating a similar direction of association between variants. We did not have serious concerns about indirectness, and did not rate down, when ≥50% of the weight of an estimate came from studies spanning both time periods. We also did not rate down for indirectness if our conclusion was little-to-no association; we assumed new associations between Omicron and conditions of interest that were not observed earlier in the pandemic were unlikely. We also had concerns when a category (or condition type encompassing multiple diagnoses) estimate relied substantially on data from a single diagnosis or limited age group.

For imprecision, we rated down once each time one of the limits of the 95% CI extended across an effect threshold (i.e., from small-to-moderate association into little-to-no difference or vice versa). We did not rate down for imprecision when we rated down for inconsistency in outcomes with ≥2 studies (i.e., when a wide 95% CI would reflect inconsistency vs. imprecision).

To assess reporting biases, we compared outcomes specified in each report’s methods section (and protocol if available) with the outcomes reported in the results section. Outcomes specified in the methods but not reported in the results (“missing”) were considered concerning.

## RESULTS

The PRISMA flow diagram for screening is available in Figure 1. We screened 4,143 unique database records from our search update and 19 additional relevant records identified from other sources. We included 46 new studies and brought forward data from 23 of the 25 studies from the original review. Two studies included in the original review^13, 14^ and five newly identified eligible studies^15–19^ were superseded by studies reporting new or better data in an overlapping population.

**FIGURE 1.**
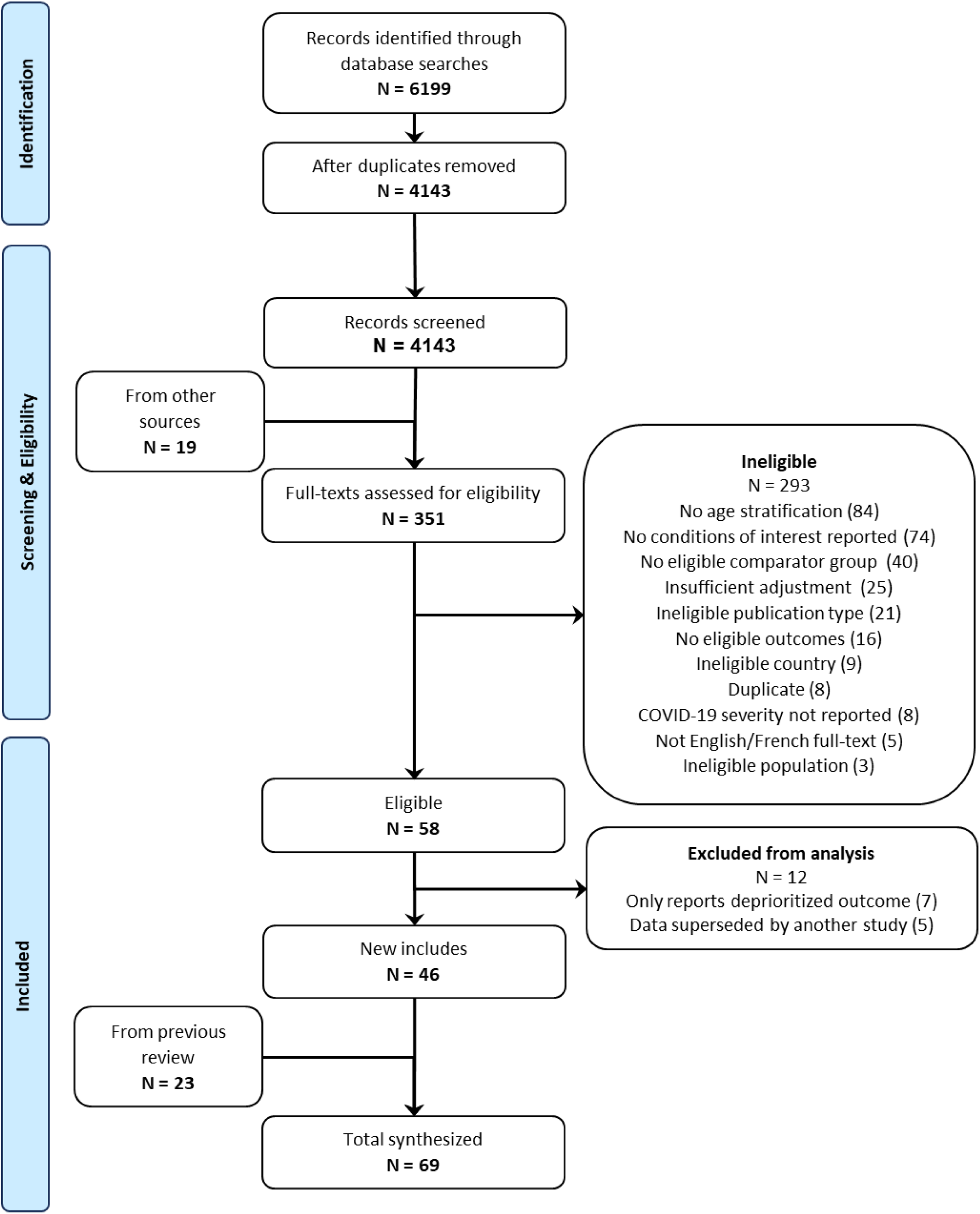
FLOW OF RECORDS THROUGH SEARCH AND SELECTION PROCESS FOR A SYSTEMATIC REVIEW UPDATE ON NEW DIAGNOSES AND EXACERBATIONS OF CHRONIC CONDITIONS AFTER SARS-COV-2 INFECTION.

### Study characteristics

Citations for and study details of included studies are available in Appendix B and Table S1, respectively. Overall, we included 69 studies from 12 countries in our analyses: United States (27), United Kingdom (10), Germany (8), Denmark (5), Korea (5), Sweden (3), Canada (2), Italy (2), Japan (2), Austria (1), Israel (1), and Spain (1), and 2 multi-national studies. Included studies primarily (58/69, 84%) used retrospective cohort study designs; 9 studies used prospective cohort designs, and 2 studies were self-controlled case series. Regardless of study design, most studies (63/69, 91%) utilized a concurrent comparator group. Twenty-one studies (30%) reported on patients <18 years old, 52 studies (75%) reported on patients 18-64 years old, and 55 studies (80%) reported on patients ≥65 years old. The median (IQR) proportion of female patients was 51.3% (49% to 56%). In terms of case confirmation, 71% (49/69) required evidence of a positive laboratory test (or applicable ICD-10 code) to categorize patients in the SARS-CoV-2 infected group. Confirmatory testing was required less often for control groups (34/69 studies, 49%).

Sixteen studies (23%) reported on new diagnosis of chronic conditions after hospitalization with SARS-CoV-2 by age category. Across studies reporting a measure of central tendency for follow-up duration (n=31), the median follow-up duration was 244 days (∼8 months). Among all studies, the median longest possible follow-up was 15 months (range: 2-42 months). Forty-nine studies (71%) reported data limited to pre-Omicron time periods and only 3 studies (4%) reported outcomes specific to the Omicron period.^20–22^ Sixty-two studies (90%) reported on new diagnoses; 7 studies (10%) reported data on exacerbations after SARS-CoV-2 infection of pre-existing chronic conditions.^23–29^

Figure 2 summarizes ROB assessments; Table S2 presents per-study details. Just over half of the studies (35/69, 51%) were assessed at moderate ROB, 10 (14%) at low ROB, and 24 (35%) at high ROB. The most frequent ROB concern was the potential for misclassification, largely due to differential exposure ascertainment methods between groups, e.g., from not confirming the absence of exposure with negative tests in the control group.

**FIGURE 2.**
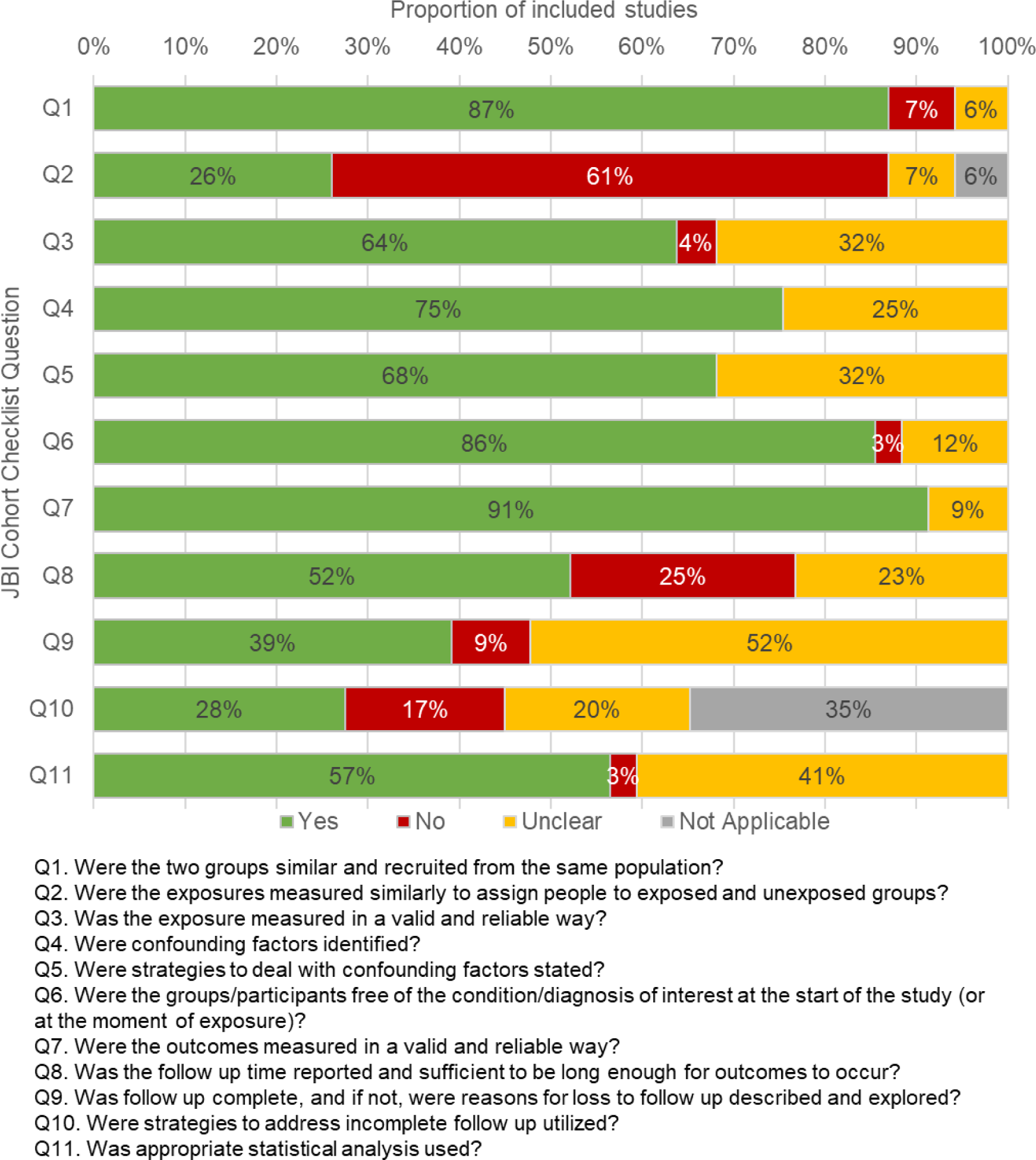
SUMMARY OF RISK OF BIAS ASSESSED BY THE JOHANNA-BRIGGS INSTITUTE (JBI) COHORT CHECKLIST FOR STUDIES INCLUDED IN A SYSTEMATIC REVIEW OF SARS-COV-2 INFECTION AND CHRONIC CONDITIONS.

### New diagnoses

Table 1 summarizes associated relative effects and their certainty for our primary analysis by age strata and acute SARS-CoV-2 care context (inpatients vs. outpatients/mixed). We do not report directions of effect for outcomes in which we had very low certainty; for transparency, effect estimates for outcomes with very low certainty are available in Table S3. Additionally, the table does not include inpatients <18 years old because data were reported for only one outcome (overall mental illness) in this group, which is presented in the Table 1 footnotes. Detailed findings for the primary analysis on new diagnoses, including explanations for certainty ratings, estimated excess incidence, and forest plots, are presented in Table S3 and Appendix C, respectively. Over half (107/206; 52%) of reported outcomes were rated down for indirectness (limited applicability) to the current context because they were informed primarily by data on pre-Omicron variants. We had no serious concerns in the reporting bias domain for any outcome and no serious concerns about missing studies or small study effects. Only findings of moderate or high certainty are reported below.

**TABLE 1.**
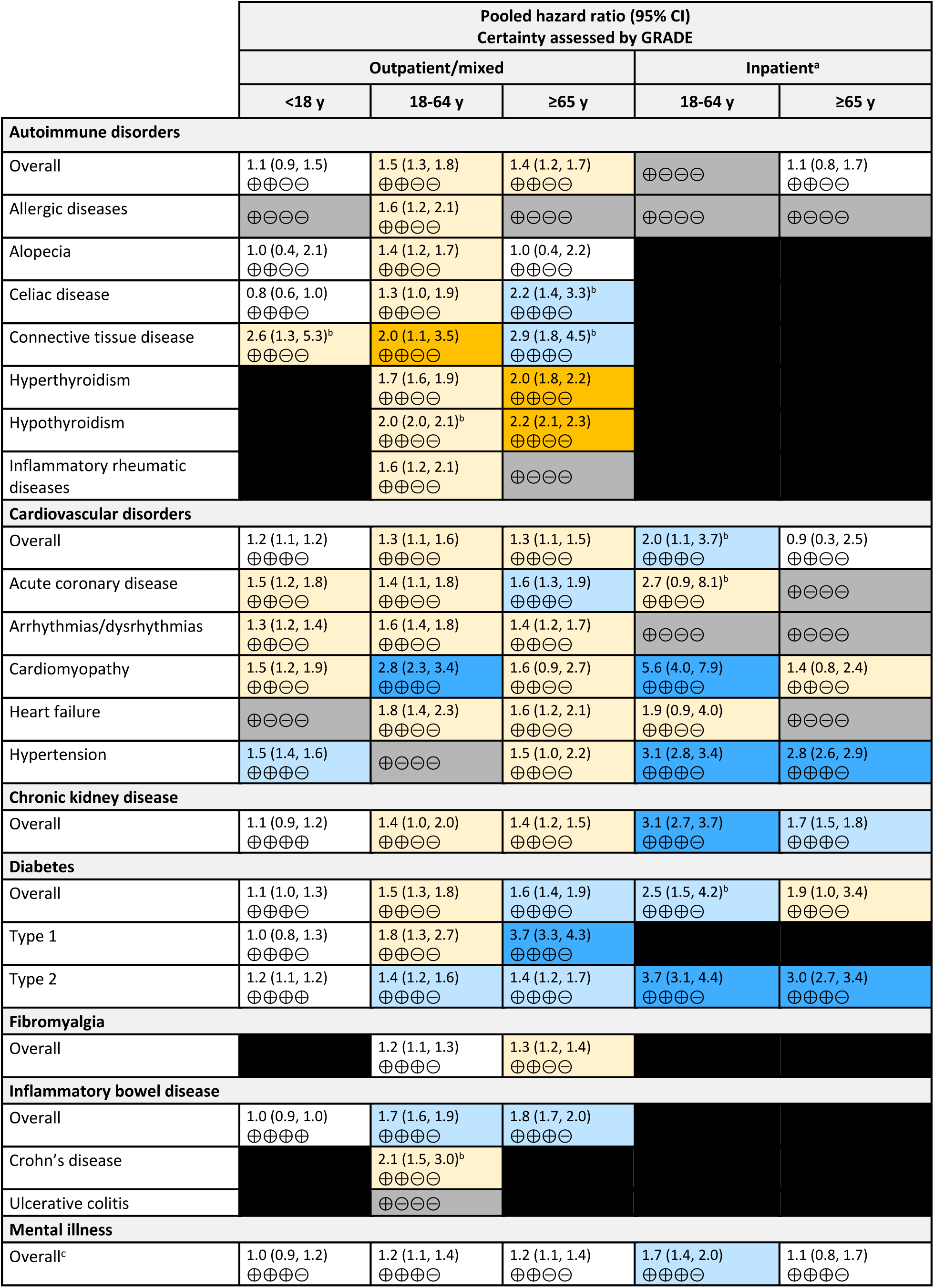
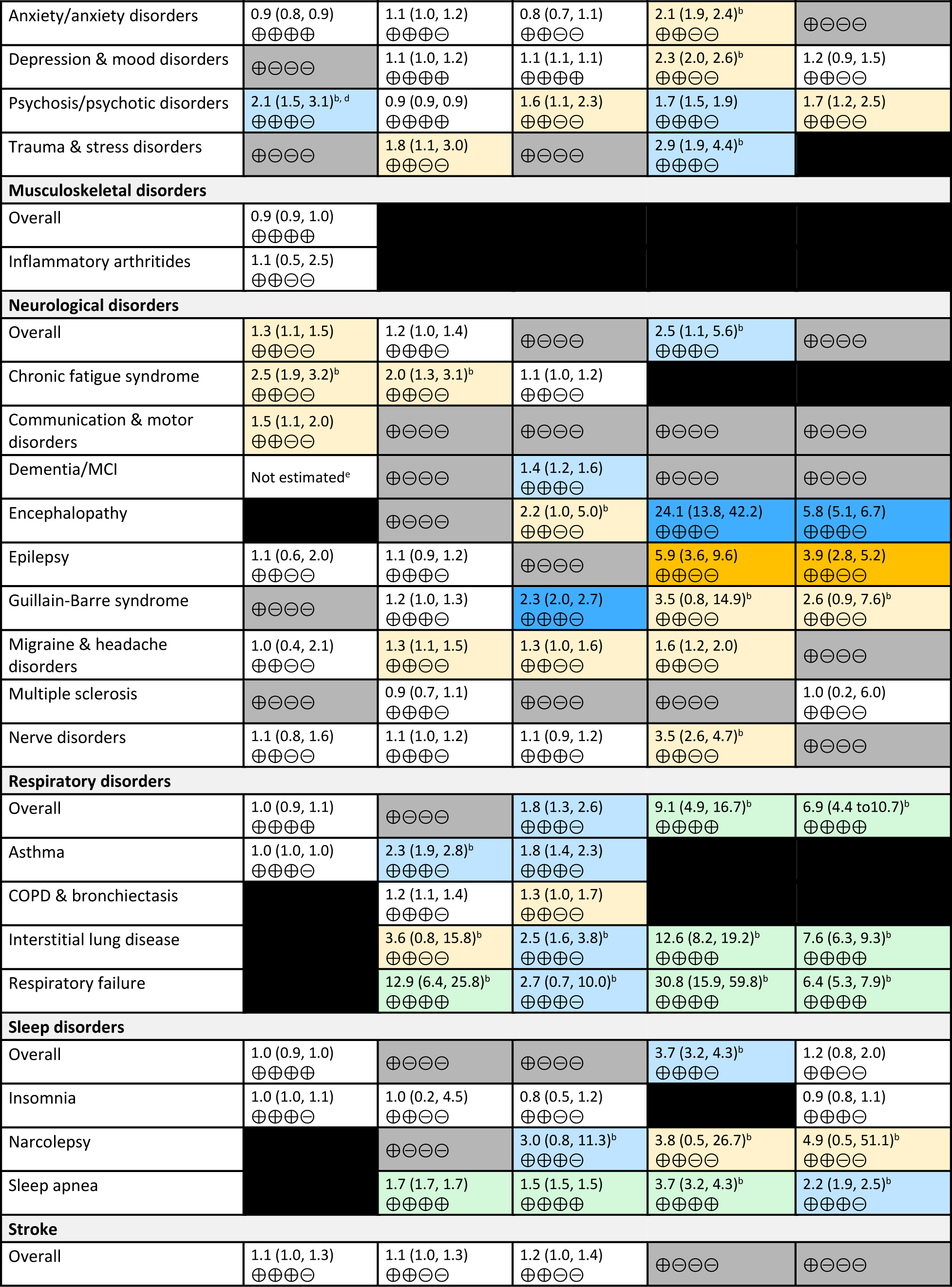
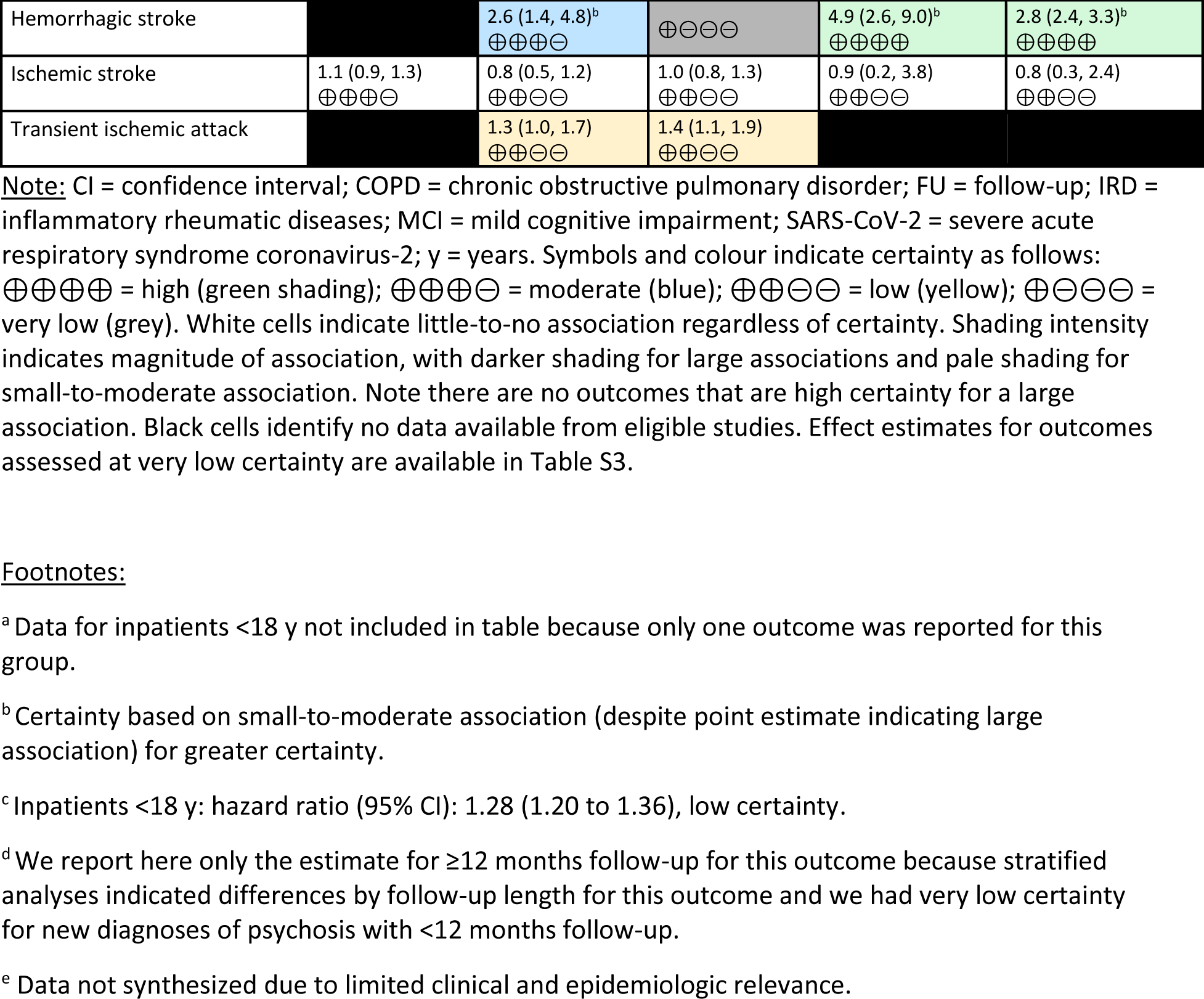
MAGNITUDE AND CERTAINTY OF EVIDENCE OF RELATIVE EFFECTS BETWEEN SARS-COV-2 INFECTION AND NEW DIAGNOSES OF CHRONIC CONDITIONS, BY CARE SETTING AND AGE GROUP.

Among <18-year-olds receiving outpatient/mixed care, there is (high certainty) little-to-no association between SARS-CoV-2 infection and subsequent new diagnoses of: type 2 diabetes, chronic kidney disease, inflammatory bowel disease, musculoskeletal disorders, anxiety/anxiety disorders, overall respiratory disorders, and overall sleep disorders. There is probably (moderate certainty) a small-to-moderate increase in new diagnoses of hypertension, and psychosis/psychotic disorders ≥12 months after infection.

For outpatients/mixed care patients 18-64 years old, there is an association between SARS-CoV-2 infection and small-to-moderate increases in respiratory failure and sleep apnea. For this group there is little-to-no association for depression and mood disorders and psychosis/psychotic disorder. There is probably a large increase in new diagnoses of cardiomyopathy and small-to-moderate increases in type 2 diabetes, inflammatory bowel disease, asthma, and hemorrhagic stroke.

Among 18-64-year-olds who received inpatient care for SARS-CoV-2 infection, there are small-to-moderate increases in subsequent new diagnoses of overall respiratory disorders, interstitial lung disease, respiratory failure, sleep apnea and hemorrhagic stroke. In this group, there are probably large increases in subsequent new diagnoses of cardiomyopathy, hypertension, chronic kidney disease, type 2 diabetes, and encephalopathy, and small-to-moderate increases in overall cardiovascular disorders, overall diabetes, overall mental illness, psychosis/psychotic disorders, trauma and stress disorders, overall neurological disorders, and overall sleep disorders.

After SARS-CoV-2 infection in ≥65-year-olds receiving outpatient/mixed care, there is a small-to-moderate increase in new diagnoses of sleep apnea, and little-to-no association between SARS-CoV-2 infection and subsequent new diagnoses of depression and mood disorders. Further, there are probably large increases in new diagnoses of type 1 diabetes and Guillain-Barre syndrome, and small-to-moderate increases in celiac disease, connective tissue disease, acute coronary disease, overall diabetes, type 2 diabetes, inflammatory bowel disease, dementia/mild cognitive impairment, overall respiratory disorders, asthma, respiratory failure, interstitial lung disease, and narcolepsy.

Among ≥65-year-olds hospitalized for SARS-CoV-2 infection, there are small-to-moderate increases in subsequent new diagnoses of overall respiratory disorders, interstitial lung disease, respiratory failure, and hemorrhagic stroke. There are probably large increases in new diagnoses of hypertension, type 2 diabetes, and encephalopathy; small-to-moderate increases in chronic kidney disease and sleep apnea.

#### Stratified secondary analyses

Stratified secondary analyses, including forest plots, by case confirmation and follow-up timing are presented in Appendices D and E. None of the 3 studies reporting Omicron-specific data reported on the same outcomes, thus we were unable to carry out planned stratified secondary analyses by variant.

Overall, heterogeneity based on case confirmation was limited; however, statistically significant differences leading to a difference in conclusions were observed in 4 outcomes among the >64-year-old age group: overall cardiovascular disorders, overall mental illness, and overall sleep disorders in the outpatient/mixed care setting; and overall mental illness in the inpatient care setting. More specifically, studies based on confirmed cases showed smaller associations between SARS-CoV-2 infection and new diagnoses compared with others; other outcomes also mirrored this trend, although without statistical significance in the differences between groups. Although this finding is methodologically important to consider for future studies, we chose not to GRADE these sub-groups separately because the clinical relevance and interpretation of the differences in risk between patients with a confirmed SARS-CoV-2 infection documented in health records compared with those diagnosed based on symptoms or at-home testing not captured in health records is unclear.

No differences in association by longest possible follow-up (<12 months vs. ≥12 months) were observed in outcomes reported by ≥2 studies per subgroup, with the exception of psychosis/psychotic disorders in outpatients/mixed patients <18 years old, for which there was a larger association with longer follow-up. When assessing certainty in the evidence for these subgroups individually, we had very low certainty for new diagnoses of psychosis with <12 months follow-up (not shown in Table 1), due to very serious concerns about inconsistency and serious concerns around indirectness. We had moderate certainty in the evidence around a small-to-moderate association (HR: 2.14; 95% CI: 1.46, 3.14, downgraded from a large association for greater certainty) of new diagnosis of psychosis ≥12 months after SARS-CoV-2 infection.

#### Within-study analyses

Eleven studies reported occurrence of new diagnoses across time or by variant of concern (Table 2).^20–22, 28, 30–36^

**TABLE 2.** SUMMARY OF TIME-VARYING AND VARIANT EFFECTS ON NEW DIAGNOSES AND EXACERBATIONS OF CHRONIC CONDITIONS AFTER SARS-COV-2 INFECTION.

| Study | Age group<br>Acute SARS-CoV-2 care | Conditions reported | Time varying effects | Effects across Variants |
| --- | --- | --- | --- | --- |
| **New diagnoses** |  |  |  |  |
| **Katsoularis 2023<sup>30</sup>** | 18-64 y, ≥65 y<br>Mixed | Cardiovascular (atrial tachycardias, paroxysmal supraventricular tachycardias, bradyarrhythmia) | In individuals ≤60 years old, the IRR for atrial tachycardia was significantly increased in the first 60 days after COVID-19 diagnosis compared to the control period. The IRR declined and became non-significant at 61-90 and 91-180 days. The IRR for paroxysmal supraventricular tachycardia was significantly elevated at 31-60 days after COVID-19, and remained significantly elevated at 91-180 days. Although elevated during the first 30 days of infection, the IRR for bradyarrhythmia was not significantly elevated in the post-acute period.<br>In individuals >60 years old; IRRs for atrial tachycardia, paroxysmal supraventricular tachycardias, and bradyarrhythmia in the post-acute phase were highest at 31 to 60 days after diagnosis, declining and becoming non-significant over time. | Not applicable (pre-Omicron) |
| **Mizrahi 2023<sup>31</sup>** | <18 y, 18-64 y, ≥65 y<br>Outpatient | Many | The HRs for most conditions in most age groups were not significantly different between 30-180 days and 180-360 days after infection.<br>In 12-18 year-olds, the HR for hypertension was significantly elevated during 30-180 days after infection, and subsequently decreased 180-360 days after infection.<br>For 41-60 year-olds, risk of cardiac arrhythmias, celiac disease, diseases of the nervous system, were higher for COVID-19 patients compared to controls 30-180 days after infection, but this difference disappeared by 180-360 days after infection.<br>In older adults (>60 years) the HR for ischemic stroke and TIA indicated decreased risk for COVID-19 patients compared to controls 30-180 days after infection, but not 180-360 days after infection. Risks of depression and diabetes mellitus were similar between groups 30-180 days after infection, but increased for COVID-19 patients compared to controls during the period 180-360 days after infection. Risk of ischemic heart disease were similar between COVID-19 and control groups at 30-180 days after infection, but the HR decreased for COVID-19 patients compared to controls during the 180-360 day period after infection. | Not applicable (pre-Omicron) |
| **Murata 2024<sup>20</sup>** | ≥65 y<br>Mixed | Mental illness (overall psychiatric disorders; anxiety disorders; mood disorders, psychotic disorders);<br>Neurological disorders (organic mental disorders [dementia])<br>Sleep disorders (Insomnia) | None reported | Non-hospitalized COVID-19 cases compared to individuals with other respiratory infections were at significantly lower risk of overall psychiatric disorders, anxiety disorders, and insomnia during the Omicron periods, but not during Alpha or Delta periods.<br>Hospitalized COVID-19 cases were also at lower risk of overall psychiatric disorders and anxiety disorders compared to other respiratory tract infections during Omicron periods, but not during Alpha or Delta periods. |
| **Noorzae 2023<sup>21</sup>** | <18 y<br>Mixed | Diabetes (type 1) | The HRs for type 1 diabetes diagnosis were similar for the time periods from 30 days to 6 months and >6 months after testing positive for SARS-CoV-2. | No differences in HRs were observed between periods of variant predominance. |
| **Rezel-Potts 2022<sup>32</sup>** | 18-64 y<br>Mixed | Cardiovascular disease<br>Diabetes | For both cardiovascular diseases and diabetes, IRR was highest at 4-7 weeks after infection, decreasing over time to RR ~1 by 24 weeks after infection. | Not applicable (pre-Omicron) |
| **Taquet 2022<sup>33</sup>** | <18 y, 18-64 y, ≥65 y<br>Mixed | Many | Outcomes fell into three categories:<br>1) within 2 years, HRs have returned to baseline (e.g., mood disorder, anxiety disorder, and ischemic stroke) and cumulative incidence equalizes between cohorts;<br>2) within 2 years HRs have returned to baseline but equal cumulative incidence was not reached;<br>3) HRs remained greater than 1 at the end of the follow-up period and new diagnoses are being made more frequently after COVID-19 diagnosis than after a diagnosis of another respiratory infection up to 2 years after the index event. These different risk trajectories were broadly similar in children, adults, and older adults. | Risk profiles differed across variants:<br>Alpha: 6-mos HRs did not notably change from before to after emergence of Alpha.<br>Delta: Increased 6-month HRs of anxiety disorders, insomnia, cognitive deficit, epilepsy or seizures, and ischemic stroke, but a lower risk of dementia, were observed in those diagnosed after the emergence of the delta variant compared to those diagnosed before.<br>Omicron: After Omicron, patients were at an increased risk (over 140 days of follow-up) of dementia, mood disorders, and nerve, nerve root, and plexus disorders. After emergence of Omicron, risk was not significantly increased for most other reported outcomes, including anxiety disorders and epilepsy or seizures. |
| **Taylor 2024<sup>34</sup>** | 18-64 y, ≥65 y<br>Mixed | Diabetes (type 2) | HRs in the pre-vaccination cohorts for all age groups were significantly increased from 5-52 weeks but not 53-102 weeks after infection, except for 60-79-year-olds for whom the HR remained significantly increased at 53-103 weeks after infection. | Not applicable (pre-Omicron) |
| **Valdivieso-Martinez 2024<sup>22</sup>** | <18 y<br>Mixed | Mental illness (major depressive disorder, anxiety disorder, adjustment disorder)<br>Neurological disorders (migraine)<br>Sleep disorder (insomnia)<br>Cardiovascular disorders (tachycardia) | None reported | The incidences of major depressive disorder, anxiety disorder, adjustment disorder, and migraine were observed to be higher among pre-Omicron cases than in unvaccinated Omicron cases, although these differences were not quantified statistically. |
| **Yang 2024<sup>35</sup>** | ≥65y<br>Outpatient | Cardiovascular disorders (acute myocardial infarction, abnormality of heart rhythm, atrial fibrillation and flutter, cardiac arrhythmia, heart failure, ischemic heart disease)<br>Stroke (all stroke, acute ischemic stroke, hemorrhagic stroke) | Across cardiovascular conditions, time-varying HRs sharply decreased from 1 to 4 months, stabilizing thereafter.<br>For stroke outcomes, time-varying HRs showed approximately linear declines between 1 and 6 months. After 6 months, the time-varying HRs for stroke outcomes generally stabilized. | Not applicable (pre-Omicron) |
| **Zhang-James 2024<sup>36</sup>** | <18 y<br>Mixed | Mental illness (new mental disorder diagnosis) | None reported | The 2-year probability of having any mental disorder diagnosis was higher for patients infected during the Omicron variant period than during Alpha or Delta periods. Similarly, the HR was significantly higher for Omicron variants than for Alpha and Delta variants. |
| **Exacerbations** |  |  |  |  |
| **Prahalad 2023<sup>28</sup>** | <18 y<br>Mixed | Diabetes (type 1) | At 90 days after cohort entry, glycated hemoglobin (HbA1c) values in COVID-19 cases did not increase at a rate different than in controls. However, beyond 180 days, HbA1c values in COVID-19 cases declined more rapidly than in the control cohort (p=0.024). This indicates a sustained increase in blood glucose after COVID-19 with a return to baseline (or even slightly lower) following recovery, potentially attributed to adjustments in insulin dosing in response to the sustained post-infection increase in blood glucose. | In patients 5-20 years old with pre-existing type 1 diabetes, the hazard ratio in the Omicron period for the composite outcome of an ED visit or hospitalization for diabetic ketoacidosis or severe hypoglycemia was less than half the HR in the pre-Omicron period (0.64 vs. 1.62, for the Omicron and pre-Omicron periods, respectively). |

Abbreviations: COVID-19 = coronavirus disease 2019; HR = hazard ratio; IRR = incidence rate ratio; RR = relative risk; SARS-CoV-2 = severe acute respiratory syndrome coronavirus 2. Note: SARS-CoV-2 infection and COVID-19 are used interchangeably.

Eight studies reported on whether risk varied with time since SARS-CoV-2 infection.^21, 28, 30–35^ In general, risk of new diagnoses of chronic conditions appears to be highest in the time periods closest to infection, with the association moving towards the null by 6-12 months post-infection.

Six studies compared risks between Omicron-lineage variants and other SARS-CoV-2 variants.^20–22, 28, 33, 36^ Among patients aged <18 years, one study reported no difference between variants for the risk of new diagnoses of type 1 diabetes, while another study found that the risk of exacerbation of pre-existing type 1 diabetes was lower for Omicron cases than pre-Omicron cases.^21, 28^ Findings from 2 studies suggested that risks for neurological disorders, including migraine and epilepsy, were not increased in Omicron periods compared to pre-Omicron periods.^22, 33^ Evidence for mental illness was conflicting. For patients <18 years, 1 study reported the 2-year probability of any mental illness diagnosis was higher for patients infected during the Omicron period,^36^ while another observed that the incidences of major depressive disorder, anxiety disorder, and adjustment disorder were lower in Omicron cases than pre-Omicron cases (although these differences were not quantified statistically).^22^ Among both non-hospitalized and hospitalized patients ≥65 years old during Omicron, but not Alpha or Delta periods, SARS-CoV-2 cases were at lower risk of overall psychiatric disorders and anxiety disorders compared to concurrent controls with non-SARS-CoV-2 respiratory infections.^20^ In another study of patients aged 0 to 65+ years identified from both outpatient and inpatient settings, there was an increased risk of mood disorders, but not anxiety disorders, in Omicron versus pre-Omicron cases.^33^ Evidence for other condition categories was sparse. Differences observed between variants may be confounded by average severity of acute disease and mortality of each variant, as well as factors other than variant such as differences in access to health services, vaccination, reinfection, at-home testing, and public health measures in place at the time of infection across time periods.

### Exacerbations of chronic conditions

Seven studies reported on exacerbation of pre-existing chronic conditions after SARS-CoV-2 infection; Table 3 summarizes the findings from these studies.^23–29^ For most chronic conditions, evidence on disease exacerbations or relapse after SARS-CoV-2 infection is either inconclusive or lacking entirely. A study on outcomes in people with type 1 diabetes <18 years old suggested a trend towards worse outcomes (i.e., emergency department visit or hospitalization for diabetic ketoacidosis or severe hypoglycemia) following SARS-CoV-2 infection.^28^ A study of worsening diabetes (type 1 & type 2) in adults ≤65 years or >65 years old found no statistically significant difference in risks of nephropathy and neuropathy in either age group but a higher risk of end-stage renal disease in those >65 years old.^29^ In 1 study, SARS-CoV-2 infection was not associated with subsequent inflammatory bowel disease-related hospitalization or surgery.^24^ Both studies reporting on exacerbation/relapse of multiple sclerosis after SARS-CoV-2 infection reported no increased risk across all of several outcomes measured.^23, 26^ One study reporting on severe asthma exacerbations found risk was higher in patients with a severe SARS-CoV-2 infection compared with a matched control group, but not among patients with a non-severe SARS-CoV-2 infection.^25^ Finally, 1 study among people <18 years old with a range of chronic conditions found that healthcare utilization was higher in all care settings (e.g., outpatient, emergency department, inpatient) for those with a SARS-CoV-2 infection compared to those without.^17^

**TABLE 3.** SUMMARIZED FINDINGS ON THE RISK OF EXACERBATIONS OF CHRONIC CONDITIONS AFTER SARS-COV-2 INFECTION.

| Study | Age group(s)<br>Acute SARS-CoV-2 care | Conditions reported | Outcome(s) | Findings |
| --- | --- | --- | --- | --- |
| **Bsteh 2022<sup>23</sup>** | 18-64 y<br>Mixed | Neurological disorders (multiple sclerosis) | Relapse (acute central nervous system inflammatory demyelinating event with a duration of at least 24 hours and separated from the last relapse by at least 30 days)<br>Worsening disability (EDSS increase of $\geq 1.5$ points from a baseline score of 0, $\geq 1.0$ points from a baseline score of 1.0–5.5, or $\geq 0.5$ points from a baseline score of $>5.5$, sustained for at least 3 months) | 12 months after COVID-19 diagnosis in patients with multiple sclerosis, COVID-19 was not associated with increased odds of relapse (aOR: 1.1; 95% CI: 0.67, 2.4) or worsening disability (aOR: 0.96; 95% CI: 0.54, 2.1). |
| **Hong 2022<sup>24</sup>** | 18-64 y<br>Mixed | IBD | IBD-related hospitalization or surgery | COVID-19 was not associated with subsequent IBD-related hospitalization or surgery (aHR: 0.84; 95% CI: 0.44, 1.42). |
| **Lee 2024<sup>25</sup>** | 18-64 y, $\geq 65$ y<br>Mixed | Respiratory disorders (asthma) | Emergency room visits or hospitalizations related to asthma and the simultaneous presence of oral or intravenous corticosteroid use | The risk of severe exacerbations was higher in the COVID-19 cohort compared with the matched cohort (HR: 1.57; 95% CI: 1.06, 2.32). Subgroup analyses revealed that risk of severe exacerbation was elevated after severe COVID-19 (HR: 5.12; 95% CI: 3.27, 8.01) but not after non-severe COVID-19 (HR: 0.83; 95% CI: 0.51, 1.34) |
| **Moniti 2024<sup>26</sup>** | 18-64 y<br>Mixed | Neurological disorders (multiple sclerosis) | Disability worsening (EDSS score increase of $\geq 1.5$, 1.0 or 0.5 from baseline EDSS score of 0, $\leq 5.5$ or $\geq 6.0$, respectively, after a 3-month relapse-free period)<br>Occurrence of relapse<br>Change in disease-modifying therapy<br>New or enlarging brain T2-hyperintense lesions | 18-24 months after infection, rates of EDSS worsening (15% vs 11%, $p=1.00$), number of relapses (6% vs 5%, $p=1.00$), disease-modifying therapy change (7% vs 4%, $p=0.81$), and patients with new T2-lesions (9% vs 11%, $p=1.00$) were similar between those with COVID-19 and those without, respectively. |
| **Pajor 2022<sup>27</sup>** | $<18$ y<br>Mixed | Many | Healthcare utilization (Any inpatient, emergency department, or outpatient healthcare visit). | Across a range of chronic conditions, healthcare utilization in all care settings was higher in those with a COVID-19 diagnosis than those without, with a larger increase for inpatient visits than for emergency department or outpatient visits. |
| **Prahalad 2023<sup>28</sup>** | $<18$ y<br>Mixed | Diabetes (type 1) | Healthcare utilization (emergency department visit or hospitalization for diabetic ketoacidosis or severe hypoglycemia) | While a trend toward worse outcomes following COVID-19 in type 1 diabetes patients was observed (HR: 1.45; 95% CI: 0.97, 2.16), the findings were not statistically significant. |
| **Wan 2023b<sup>29</sup>** | 18-64 y, $\geq 65$ y<br>Mixed | Diabetes (type 1 or type 2) | Disease worsening (development of neuropathy, nephropathy, or end-stage renal disease) | Risk of nephropathy after COVID-19 was not significantly elevated in either those $>65$y (HR: 1.09; 95% CI: 0.50, 2.5) or $\leq 65$ y (HR: 1.2; 95% CI: 0.67, 1.90). Risk of neuropathy was also not elevated in either age group (HR: 1.7; 95% CI: 0.98, 3.50 and HR: 1.48; 95% CI: 0.75, 2.70 for $>65$y and $\leq 65$ y, respectively). Risk of end-stage renal disease after COVID-19 was significantly elevated for individuals $>65$y old (HR: 2.18; 95% CI: 1.0, 4.50) but not for those $\leq 65$ y (HR: 2.17; 95% CI: 0.59, 6.0) compared with uninfected individuals. |

Abbreviations: aOR = adjusted odds ratio; aHR = adjusted hazard ratio; CI = confidence interval; COVID-19 = coronavirus disease 2019; EDSS = Expanded Disability Status Scale; HR = hazard ratio; IBD = inflammatory bowel disease; SARS-CoV-2 = severe acute respiratory syndrome coronavirus 2, y=years. Note: SARS-CoV-2 infection and COVID-19 are used interchangeably.

## DISCUSSION

We updated a previous systematic review from 2023 examining associations between SARS-CoV-2 infection and subsequent new diagnoses or exacerbations of chronic conditions.^2^ Compared to the original review, this update adds 13 condition types in 4 additional categories of chronic conditions (autoimmune disorders, fibromyalgia, inflammatory bowel diseases, sleep disorders). Our comprehensive search strategy identified many new relevant studies, nearly tripling the number of included studies from 25 in the original review to 69 in this update. Our strict eligibility criteria, including a requirement that studies control for confounding by sex and comorbidities likely increased the overall quality of studies included in our review, while stratifying the analyses by age category and severity of acute SARS-CoV-2 infection (using a proxy of hospitalization) enables meaningful interpretation of the findings. Our findings suggest increased diagnoses of several chronic conditions in adults (18-64 y) and older adults (≥65 y) after SARS-CoV-2 infection. At least small associations for cardiovascular disorders, diabetes, and respiratory disorders were somewhat consistent across adult age categories and acute SARS-CoV-2 severities. In pediatric (<18 years old) patients, we had at least moderate certainty about little-to-no association for many outcomes in the outpatient/mixed care group, although many pediatric outcomes were reported by single studies. Additionally, only 7 studies reported on exacerbations of pre-existing chronic conditions after SARS-CoV-2 infection, all in outpatient or mixed care groups. Large, high-quality studies of the long-term impact of SARS-CoV-2 infection on pre-existing conditions are still lacking in all ages and in individuals hospitalized during acute SARS-CoV-2 infection.

Despite the many thousands of studies on SARS-CoV-2, gaps in the evidence persist for many areas of enquiry. Data are especially sparse for pediatric populations. Although the number of studies reporting outcomes in patients <18 years old increased from 10 in the initial review to 21 in this update, as already stated, many outcomes for the <18 years old group were reported by single studies. Despite this, there was some consistency across outcomes for <18-year-olds, many of which showed little-to-no association. Only 1 study reported on new diagnoses of chronic conditions after SARS-CoV-2 infection in pediatric patients who received hospital care, and eligible studies reporting on exacerbations of pre-existing chronic conditions in children and adolescents were also limited.

Age-stratified data from the latter half of the pandemic (i.e., since the rise of Omicron variants) were sparse in our review; data specific to the Omicron period was reported by only 3 studies. We are uncertain about the continued relevance of findings from earlier in the pandemic due to differences in vaccination rates, circulating variants of concern, and reinfections, all of which have been shown to impact risk of post-acute sequelae.^37–39^ Thus, the findings of this review likely reflect the impact of SARS-CoV-2 infection on people affected during the pre-Omicron pandemic period, but may less accurately reflect the impact from contemporary or future SARS-CoV-2 infections.

Finally, it is worth mentioning that although we assessed certainty in relative effects, rather than absolute effects, the magnitude of relative association alone does not present the entire picture. A condition showing a large association but with a low background incidence may be less burdensome than a condition showing a smaller association but with a higher background incidence. For example, we report, with moderate certainty, a large association (HR: 2.32, 95% CI: 2.00, 2.70) for Guillain-Barre syndrome in ≥65 year old outpatient/mixed patients, an increase of 1.8 additional diagnoses per 10,000 SARS-CoV-2 cases over 6 months (Table S3). On the other hand, in this same group, a small-to-moderate increase in overall diabetes (about which we also had moderate certainty) represents an increase of 144 additional diagnoses per 10,000 SARS-CoV-2 cases over 6 months (Table S3). We have tried not to over emphasize the importance of the relative effect sizes reported here, but it remains important for readers to consider the implications of our findings within the broader epidemiologic picture.

### Strengths and Limitations

As do all reviews, our work has both strengths and limitations. Our review is comprehensive, both in terms of the search strategy and the breadth of chronic conditions of interest. We evaluated the risk of bias in each study and systematically assessed our certainty in the evidence in order to emphasize more reliable findings. In addition to stratification by age and acute SARS-CoV-2 severity in our primary analysis, we carried out additional stratified analyses in order to tease out nuance within our findings. While we required eligible studies to utilize a non-SARS-CoV-2 comparator and use either design or analysis methods to account for confounding by sex and comorbidities, we did not require studies to limit control groups to individuals with a negative SARS-CoV-2 test, potentially leading to contamination with SARS-CoV-2 infected individuals in some control groups. Additionally, many of the meta-analyses do not account for other important confounders, such as vaccination status, recurrent SARS-CoV-2 infections, or differences in care-seeking behaviour between control and SARS-CoV-2 infected groups, which studies rarely reported or controlled for. Additional factors that may confound observed associations between SARS-CoV-2 infection and new diagnoses of chronic conditions, such as increased health care contacts and monitoring in infected individuals, are difficult to fully account for in observational study designs.

### Conclusions

After SARS-CoV-2 infection, there is probably an increased risk of new diagnoses for several, but not all chronic conditions, especially in adults (18-64 years) and older adults (≥65 years). While we observed at least a small (i.e., at least 26%) increase in new diagnoses of some chronic conditions after SARS-CoV-2 infection, the extent of increased risk causally attributable to the SARS-CoV-2 virus, versus other factors (e.g., detection bias due to increased care seeking by individuals with SARS-CoV-2 infection, differential vaccination rates) is unclear and the certainty in the evidence should be interpreted around conclusions of association, rather than causality. Finally, how the associations between SARS-CoV-2 infection and new chronic condition diagnoses change over time since infection or by variant of concern is still uncertain for many conditions.

## Data Availability

All data produced in the present study are available upon reasonable request to the authors.

## Abbreviations

CI: confidence interval
GRADE: Grading of Recommendations, Assessment, Development and Evaluation
HR: hazard ratio
ICD: international classification of diseases
IRR: incidence rate ratio
JBI: Johanna Briggs Institute
PRISMA: Preferred Reporting Items for Systematic Reviews and Meta-Analyses
ROB: risk of bias
SARS-CoV-2: severe acute respiratory syndrome-coronavirus-2

## Acknowledgements

This work was supported by the Public Health Agency of Canada under Contract no. 4600002510. Dr. Hartling is supported by a Canada Research Chair in Knowledge Synthesis and Translation, and is a Distinguished Researcher with the Stollery Science Lab supported by the Stollery Children’s Hospital Foundation. We thank the Chronic Disease surveillance experts at the Public Health Agency of Canada for their contributions to this work. The analyses, conclusions, opinions and statements expressed herein are solely those of the authors. No endorsement by the Public Health Agency of Canada is intended or inferred.

## Conflicts of Interest

The authors declare no conflicts.

## Notes

### Competing Interest Statement

The authors have declared no competing interest.

### Funding Statement

This work was supported by the Public Health Agency of Canada. No endorsement by the Public Health Agency of Canada is intended or inferred.

