## Supplementary material for "Risk of new diagnoses and exacerbations of chronic conditions after SARS-CoV-2 infection: a systematic review update": Suplemental file

#### Appendix A: Search strategies for Medline and Embase for a systematic review update on new diagnoses and exacerbations of chronic conditions after SARS-CoV-2 infection

**MEDLINE search strategy**

Database(s): Ovid MEDLINE(R) ALL 1946 to September 03, 2024

| # | Searches | Results |
|---|---|---|
| 1 | ("after covid*" or postcovid* or postcorona* or (post* adj (COVID or COVID-19 or COVID19 or coronavirus* or corona virus* or SARS-CoV-2 or SARS-CoV2 or SARSCoV-2))).ti,ab,kf. | 19208 |
| 2 | (("after-acute" or "after discharge" or "after hospital discharge" or "after-infection" or post-acute or postacute or post-discharg* or postdischarg* or postinfect* or post-infect* or post-viral or postviral or sequela* or survivor$1) adj5 (COVID or COVID-19 or COVID19 or coronavirus* or corona-virus* or SARS-CoV-2 or SARS-CoV2 or SARSCoV-2 or SARSCoV2)).ti,ab,kf. | 7825 |
| 3 | (("more than" or longer or "at least" or "after" or follow* or from or upward* or later* or over or prospectiv* or retrospectiv* or review* or longitud* or post or beyond) adj5 (day? or week? or wk or wks or month? or mo or mos or year? or yr or yrs)).ti,ab,kf. | 3500784 |
| 4 | (COVID or COVID-19 or COVID19 or coronavirus* or corona virus* or SARS-CoV-2 or SARS-CoV2 or SARSCoV-2 or SARSCoV2).ti. | 366200 |
| 5 | 3 and 4 | 47032 |
| 6 | 1 or 2 or 5 [AFTER COVID] | 63270 |
| 7 | Chronic Disease/ | 287241 |
| 8 | (Chronic adj3 (condition$1 or diagnos$2 or disease$1 or disorder* or failure or illness*)).ti,ab,kf. | 545095 |
| 9 | (("late sequela*" or (longterm or long-term)) adj3 (consequence* or damage or effect$1 or impact* or outcome* or result* or sequela*)).ti,ab,kf. | 286061 |
| 10 | exp Cardiovascular Disease/ | 2812841 |
| 11 | (((cardio* or cardia* or heart? or myocardi* or vascula* or atria* or ventric* or coronary) and (disease* or disorder* or condition* or syndrome* or illness* or abnormal* or dysfunction* or disturb* or event* or adverse* or inflam*)) or nstemi or stemi or cvd or fibrillation or "paroxysmal dyspnea" or arrhythmia* or dysrhythmia* or bradycard* or "brugada syndrome*" or "commotio cordis" or "long qt syndrome" or parasystole or "pre-excitation syndrome*" or tachycardia* or "conduction disturbance*").ti,ab,kf. | 1875729 |
| 12 | ("cardiovascular stroke*" or "heart attack*" or "heart infarct*" or "myocardial infarct*").ti,ab,kf. | 245846 |
| 13 | exp myocardial revascularization/ | 98726 |

| | | |
|---|---|---|
| 14 | "myocardial revasculari?ation".ti,ab,kf. | 5462 |
| 15 | Diabetes mellitus, type 1/ or Diabetes mellitus, type 2/ or Diabetic ketoacidosis/ or Prediabetic state/ | 265112 |
| 16 | (diabet* or "t1dm" or "t1d" or "t2dm" or "t2d" or "niddm" or insulin resistan* or prediabet* or dysglyc?emi* or hyperglyc?emi* or hypoglyc?emi* or hyper glyc?emi* or hypo glyc?emi* or "hyperosmolar state*" or ketoacidosis or (impaired fasting adj2 (glucose or sugar)) or ((glucose or carbohydrate*) adj3 (toler* or intoler*))).ti,ab,kf. | 961379 |
| 17 | exp Gout/ | 14560 |
| 18 | (gout* or cheiragra* or chiragra* or urate inflammation* or pseudogout* or (crystal* adj3 arthropath*)).ti,ab,kf. | 20541 |
| 19 | Joint Diseases/ or Rheumatic Diseases/ | 50724 |
| 20 | exp Osteoarthritis/ | 81869 |
| 21 | (arthros* or arthrit* or monarthrit* or monoarthrit* or coxitis* or sacroiliitis or osteoarthrit* or "polymyalgia rheumatica" or spondylosis* or spondylarthrosis or spondyloarthrosis).ti,ab,kf. | 358218 |
| 22 | exp Mental Disorders/ | 1506218 |
| 23 | (anxiety* or depression* or schizo* or psychotic* or psychosis or psychoses or amnesia* or delirium* or gender dysphoria* or paraphilia* or hallucination* or paranoid or paranoia or paraphrenia or sexual dysfunction* or hysteria or phobia? or agoraphobia or neurasthenia or hypochondriasis or hypochondria or hypochondriac or adjustment reaction*).ti,ab,kf. | 872740 |
| 24 | ((manic or mania or depress*) adj3 episode*).ti,ab,kf. | 16319 |
| 25 | ("anorexia nervosa" or bulimia or PTSD or ((bipolar* or eating or personality or mood* or conduct or impulse or dissociative or eating or neurotic or obsessive or compulsive or panic or phobic or neurocognitiv* or traumatic* or posttraumatic* or post-traumatic or stress* or neurocognitive* or amnestic* or behaviour* or behavior* or neurobehav* or delusion* or manic or mania or depress* or conversion or somatoform or gender identity or attention-deficit or hyperactivity or conduct or emotion* or tic or tics or sleep or sexual identity or dysthymic or alcohol-induced or drug-induced or social functioning) adj3 disorder*)).ti,ab,kf. | 359589 |
| 26 | ((mental* or psychiatric* or psychological* or neuropsych*) adj3 (disorder* or ill or illness* or diagnosis or diagnoses)).ti,ab,kf. | 217874 |
| 27 | ((intellectual* or developmental* or neurodevelopmental or learning) adj3 (disabilit* or disorder* or syndrome*)).ti,ab,kf. | 82430 |
| 28 | exp Nervous System Diseases/ | 2977886 |
| 29 | exp Dementia/ | 218576 |
| 30 | (epilep* or seizure* or "panayiotopoulos syndrome" or "rasmussen syndrome" or "alpers disease" or hypsarrhythmia or infantie spasm* or "lennox gastaut | 1273367 |

| | | |
|---|---|---|
| | syndrome*" or merrf syndrome* or myoclonus* or "nodding syndrome*" or lipofuscinosis or (sclerosis adj3 (multiple* or disseminate* or insular*)) or chariot disease* or parkinsonism* or hemiparkinsonism* or (parkinson* adj3 (syndrom* or disorder* or disease*)) or "paralysis agitans" or amnesia* or alzheimer* or aphasi* or "diffuse neurofibrillary tangle*" or "lobar degeneration*" or "acquired dyslexia" or alexia or cadasil or "cada sil" or "cerebral autosomal dominant arteriopathy" or "lewy body disease*" or (pick* adj1 (complex* or disease* or disorder* or syndrome*)) or huntington* or "juvenile chorea" or "chorea major" or "kluver bucy" or (mental* adj3 deteriorat*) or ((neuro* or neural* or cognitive* or cognition* or consciousness or sleep* or wake*) adj3 (impair* or side effect* or function* or disfunction* or dysfunction* or disorder* or degenerat* or disease*)) or "pseudo-dementia*" or pseudodementia* or "rett syndrome*" or senile* or tauopath* or tourette*).ti,ab,kf. | |
| 31 | exp Stroke/ | 184413 |
| 32 | ("cerebrovascular accident*" or "cerebro-vascular accident*" or stroke or strokes or ((brain* or cerebellar or cerebral or lacunar) adj2 infarct*)).ti,ab,kf. | 370242 |
| 33 | Attack, Transient Ischemic/ | 22282 |
| 34 | "transient ischemic attack*".ti,ab,kf. | 14992 |
| 35 | exp Pulmonary Disease, Chronic Obstructive/ | 70293 |
| 36 | ((chronic obstructive adj4 (pulmonary or airway? or lung?)) or chronic airflow obstruction* or copd or coad or aecopd or obstructive pulmonary or asthma* or emphysema* or (chronic adj3 (bronchitis or bronchus infection* or bronchi infection* or bronchus inflammation or bronchi inflammation))).ti,ab,kf. | 298563 |
| 37 | exp Renal Insufficiency, Chronic/ | 140432 |
| 38 | (((chronic kidney or chronic renal) adj2 (disease? or insufficienc* or failur?)) or CKD or chronic nephropath*).ti,ab,kf. | 122166 |
| 39 | (or/7-38) and (COVID or COVID-19 or COVID19 or coronavirus* or corona virus* or SARS-CoV-2 or SARS-CoV2 or SARSCoV-2 or SARSCoV2).ti. [CONDITIONS 1] | 89089 |
| 40 | 6 and 39 [AFTER COVID CONDITIONS 1] | 21958 |
| 41 | Inflammatory Bowel Diseases/ | 33584 |
| 42 | ("Chronic inflammatory bowel disease*" or IBD or "indeterminate colitis").ti,ab,kf. | 41543 |
| 43 | exp Colitis/ | 66122 |
| 44 | Enterocolitis/ | 2550 |
| 45 | (colitis or enterocolitis).ti,ab,kf. | 102610 |
| 46 | Crohn Disease/ | 46297 |
| 47 | "crohn* disease".ti,ab,kf. | 58777 |
| 48 | Irritable Bowel Syndrome/ | 10038 |

| | | |
|---|---|---|
| 49 | (("irritable bowel" or "irritable colon") adj syndrome*).ti,ab,kf. | 17520 |
| 50 | or/41-49 [GI] | 194168 |
| 51 | Graves Disease/ | 16473 |
| 52 | Hyperthyroidism/ | 27252 |
| 53 | (hyperthyroid* or "Graves disease" or "Grave's Disease" or "Basedow* Disease").ti,ab,kf. | 40024 |
| 54 | Hypothyroidism/ | 30004 |
| 55 | Hypothyroidism, Autoimmune.rs. [Supplementary Concept] | 115 |
| 56 | (Hypothyroid* or ((deficien* adj1 (thyroid or "Thyroid-Stimulating Hormone")) or "TSH Deficien*")).ti,ab,kf. | 43829 |
| 57 | Hashimoto Disease/ | 4485 |
| 58 | Thyroiditis, Autoimmune/ | 7466 |
| 59 | ("Autoimmune Thyroiditis" or (Hashimoto* adj (disease or thyroid*)) or "Lymphocytic Thyroiditis").ti,ab,kf. | 11944 |
| 60 | Lupus Erythematosus, Systemic/ | 63418 |
| 61 | ("Systemic Lupus Erythematosus" or (SLE and lupus)).ti,ab,kf. | 64426 |
| 62 | Psoriasis/ | 44295 |
| 63 | Arthritis, Psoriatic/ | 8748 |
| 64 | (Psoriasis or "psoriatric arthritis").ti,ab,kf. | 54176 |
| 65 | exp Vasculitis/ and Autoimmune Diseases/ | 1530 |
| 66 | Vasculitis, Central Nervous System/ | 1342 |
| 67 | Lupus Vasculitis, Central Nervous System/ | 1045 |
| 68 | Giant Cell Arteritis/ | 7845 |
| 69 | ("auto-immune vasculitis" or "autoimmune vasculitis" or (("auto-immune vascular" or "autoimmune vascular") adj (disease* or disorder* or inflammat*))).ti,ab,kf. | 323 |
| 70 | ("Primary Central Nervous System Vasculitis" or "Primary CNS Vasculitis" or "Giant cell arteritis" or vasculitis).ti,ab,kf. | 47633 |
| 71 | Vitiligo/ or vitiligo.ti,ab,kf. | 10128 |
| 72 | exp Skin Diseases/ and autoimmune.ti,ab,kf. | 20088 |
| 73 | (((skin or cutaneous or dermatological) adj3 (condition* or disorder* or disease*)) and autoimmune).ti,ab,kf. | 4760 |

| | | |
|---|---|---|
| 74 | or/51-73 [Other autoimmune] | 319271 |
| 75 | (50 or 74) and (COVID or COVID-19 or COVID19 or coronavirus* or corona virus* or SARS-CoV-2 or SARS-CoV2 or SARSCoV-2 or SARSCoV2).ti. [GI, Other Autoimmune] | 4231 |
| 76 | 6 and 75 [AFTER COVID CONDITIONS 2] | 1221 |
| 77 | *Incidence/ or *Prevalence/ or *Risk/ or *Odds Ratio/ | 6038 |
| 78 | (incidence or incident or prevalent or "logistic regression$" or "cox regression$" or odds or ((prevalence or rate or rates or risk or risks or hazard) adj5 (relative or ratio$ or difference$ or proportional))).ti,ab,kf. | 2418829 |
| 79 | ((diagno$2 or likelihood or likely) adj3 (develop* or elevated or high or higher or heightened or increase* or increasing)).ti,ab,kf. | 100794 |
| 80 | ((new or newly) adj3 (onset or diagnos$2 or problem$1)).ti,ab,kf. | 121906 |
| 81 | *Disease Progression/ or (exacerbat* or flare$1 or provoke$1 or provocation or trigger* or worsen*).ti,ab,kf. | 757302 |
| 82 | *Prognosis/ or (prognos?s or trajector*).ti,ab,kf. | 720133 |
| 83 | or/77-82 | 3805168 |
| 84 | Epidemiologic Methods/ | 31625 |
| 85 | exp Epidemiologic Studies/ | 3336658 |
| 86 | Observational Studies as Topic/ | 10015 |
| 87 | Clinical Studies as Topic/ | 842 |
| 88 | (Observational Study or Clinical Study).pt. | 167140 |
| 89 | (observational adj3 (study or studies or design or analysis or analyses)).ti,ab,kf. | 252938 |
| 90 | cohort*.ti,ab,kf. | 973933 |
| 91 | (prospective adj3 (study or studies or design or analysis or analyses)).ti,ab,kf. | 513863 |
| 92 | ((follow-up or followup) adj3 (study or studies or design or analysis or analyses)).ti,ab,kf. | 105239 |
| 93 | ((longitudinal or long-term or longterm) adj3 (study or studies or design or analysis or analyses or data)).ti,ab,kf. | 274425 |
| 94 | (retrospective adj3 (study or studies or design or analysis or analyses or data or review)).ti,ab,kf. | 738677 |
| 95 | (case-referent adj3 (study or studies or design or analysis or analyses)).ti,ab,kf. | 642 |
| 96 | (population adj3 (study or studies or analysis or analyses)).ti,ab,kf. | 254040 |
| 97 | (descriptive adj3 (study or studies or design or analysis or analyses)).ti,ab,kf. | 123941 |

| | | |
|---|---|---|
| 98 | ((multidimensional or multi-dimensional) adj3 (study or studies or design or analysis or analyses)).ti,ab,kf. | 5492 |
| 99 | ((cross-sectional or cross-sectional or crosssectional or crossectional) adj3 (study or studies or design or research or analysis or analyses or survey or findings)).ti,ab,kf. | 479195 |
| 100 | ((natural adj experiment) or (natural adj experiments)).ti,ab,kf. | 3799 |
| 101 | (quasi-experiment* or quasi-experiment*).ti,ab,kf. | 23405 |
| 102 | ((non-experiment$2 or nonexperiment$2) adj3 (study or studies or design or analys?s)).ti,ab,kf. | 1845 |
| 103 | (prevalence adj3 (study or studies or analys?s)).ti,ab,kf. | 54516 |
| 104 | Control groups/ | 2131 |
| 105 | Case-control studies/ | 338163 |
| 106 | ((case or group* or study or patient) adj3 (control$2 or compar* or match* or series)).ti,ab,kf. | 1736810 |
| 107 | Population surveillance/ | 63048 |
| 108 | (surveillance adj3 (continuous or continual or continued or data or ongoing or population or research or study)).ti,ab,kf. | 38167 |
| 109 | or/84-108 | 5701152 |
| 110 | 40 and 83 and 109 [AFTER COVID CONDITIONS 1 + IPRE + STUDY DESIGN APPLIED] | 5254 |
| 111 | 110 and (202207* or 202208* or 202209* or 202210* or 202211* or 202212* or 2023* or 2024*).dt,ez,da. [CONDITIONS 1 Update] | 3137 |
| 112 | 76 and 83 and 109 [AFTER COVID CONDITIONS 2 + IPRE + STUDY DESIGN APPLIED] | 246 |
| 113 | limit 112 to yr="2020 -Current" [CONDITIONS 2 date limit] | 246 |
| 114 | 111 or 113 [AFTER COVID CONDITIONS] | 3295 |
| 115 | (exp Animals/ or Models, Animal/ or Disease Models, Animal/) not Humans/ | 5254991 |
| 116 | ((animal or animals or canine* or dog or dogs or feline or hamster* or lamb or lambs or mice or monkey or monkeys or mouse or murine or pig or pigs or piglet* or porcine or primate* or rabbit* or rats or rat or rodent* or sheep* or veterinar*) not (human* or patient*)).mp. | 5156089 |
| 117 | 114 not (115 or 116) [ANIMAL-ONLY STUDIES REMOVED] | 3295 |
| 118 | (Case Reports.pt. or (case report? or case study or case presentation).ti.) not (review* or trial*).ti,ab,kf,hw. | 2216731 |
| 119 | (comment or editorial or news or newspaper article).pt. | 1758972 |
| 120 | 117 not (118 or 119) [CASE STUDIES, OPINION PIECES REMOVED] | 3238 |

| | | |
|---|---|---|
| 121 | 120 not pandemic.ti. | 2583 |
| 122 | remove duplicates from 121 | 2564 |

**EMBASE search strategy**

Database(s): Embase 1974 to 2024 September 03

| # | Searches | Results |
|---|---|---|
| 1 | ("after covid*" or postcovid* or postcorona* or (post* adj (COVID or COVID-19 or COVID19 or coronavirus* or corona virus* or SARS-CoV-2 or SARS-CoV2 or SARSCoV-2))).ti,ab,kw. | 27655 |
| 2 | (("after-acute" or "after discharge" or "after hospital discharge" or "after-infection" or post-acute or postacute or post-discharg* or postdischarg* or postinfect* or post-infect* or post-viral or postviral or sequela* or survivor$1) adj5 (COVID or COVID-19 or COVID19 or coronavirus* or corona-virus* or SARS-CoV-2 or SARS-CoV2 or SARSCoV-2 or SARSCoV2)).ti,ab. | 10079 |
| 3 | (("more than" or longer or "at least" or "after" or follow* or from or upward* or later* or over or prospectiv* or retrospectiv* or review* or longitud* or post or beyond) adj5 (day? or week? or wk or wks or month? or mo or mos or year? or yr or yrs)).ti,ab. | 5270209 |
| 4 | (COVID or COVID-19 or COVID19 or coronavirus* or corona virus* or SARS-CoV-2 or SARS-CoV2 or SARSCoV-2 or SARSCoV2).ti. | 416454 |
| 5 | 3 and 4 | 65209 |
| 6 | 1 or 2 or 5 [AFTER COVID] | 86367 |
| 7 | chronic disease/ | 212266 |
| 8 | (Chronic adj3 (condition$1 or diagnos$2 or disease$1 or disorder* or failure or illness*)).ti,ab. | 788825 |
| 9 | (("late sequela*" or (longterm or long-term)) adj3 (consequence* or damage or effect$1 or impact* or outcome* or result* or sequela*)).ti,ab. | 418195 |
| 10 | transient ischemic attack/ | 50341 |
| 11 | cardiovascular disease/ | 377950 |
| 12 | heart muscle revascularization/ | 40165 |
| 13 | cerebrovascular accident/ | 317620 |
| 14 | dementia/ | 158571 |
| 15 | insulin dependent diabetes mellitus/ or non insulin dependent diabetes mellitus/ or diabetic ketoacidosis/ or impaired glucose tolerance/ | 514311 |
| 16 | gout/ | 27501 |
| 17 | arthropathy/ or rheumatic disease/ | 77971 |
| 18 | osteoarthritis/ | 110915 |
| 19 | mental disease/ | 298430 |

| | | |
|---|---|---|
| 20 | neurologic disease/ | 172630 |
| 21 | chronic obstructive lung disease/ | 189364 |
| 22 | chronic kidney failure/ | 168033 |
| 23 | ("cardiovascular stroke*" or "heart attack*" or "heart infarct*" or "myocardial infarct*").ti,ab. | 348396 |
| 24 | "myocardial revasculari?ation".ti,ab. | 6739 |
| 25 | (((cardio* or cardia* or heart? or myocardi* or vascula* or atria* or ventric* or coronary) and (disease* or disorder* or condition* or syndrome* or illness* or abnormal* or dysfunction* or disturb* or event* or adverse* or inflam*)) or nstemi or stemi or cvd or fibrillation or paroxysmal dyspnea or arrhythmia* or dysrhythmia* or bradycard* or brugada syndrome* or commotio cordis or long qt syndrome or parasystole or pre-excitation syndrome* or tachycardia* or conduction disturbance*).ti,ab. | 2727172 |
| 26 | ("cerebrovascular accident*" or "cerebro-vascular accident*" or stroke or strokes or ((brain* or cerebellar or cerebral or lacunar) adj2 infarct*)).ti,ab. | 573961 |
| 27 | "transient ischemic attack*".ti,ab. | 23326 |
| 28 | (diabet* or "t1dm" or "t1d" or "t2dm" or "t2d" or "niddm" or insulin resistan* or prediabet* or dysglyc?emi* or hyperglyc?emi* or hypoglyc?emi* or hyper glyc?emi* or hypo glyc?emi* or "hyperosmolar state*" or ketoacidosis or (impaired fasting adj2 (glucose or sugar)) or ((glucose or carbohydrate*) adj3 (toler* or intoler*))).ti,ab. | 1421764 |
| 29 | ((chronic obstructive adj4 (pulmonary or airway? or lung?)) or chronic airflow obstruction* or copd or coad or aecopd or obstructive pulmonary or asthma* or emphysema* or (chronic adj3 (bronchitis or bronchus infection* or bronchi infection* or bronchus inflammation or bronchi inflammation))).ti,ab. | 440797 |
| 30 | (arthros* or arthrit* or monarthrit* or monoarthrit* or coxitis* or sacroiliitis or osteoarthrit* or "polymyalgia rheumatica" or spondylosis* or spondylarthrosis or spondyloarthrosis).ti,ab. | 496822 |
| 31 | (gout* or cheiragra* or chiragra* or urate inflammation* or pseudogout* or (crystal* adj3 arthropath*)).ti,ab. | 26783 |
| 32 | (epilep* or seizure* or "panayiotopoulos syndrome" or "rasmussen syndrome" or "alpers disease" or hypsarrhythmia or infantie spasm* or "lennox gastaut syndrome*" or merrf syndrome* or myoclonus* or "nodding syndrome*" or lipofuscinosis or (sclerosis adj3 (multiple* or disseminate* or insular*)) or chariot disease* or parkinsonism* or hemiparkinsonism* or (parkinson* adj3 (syndrom* or disorder* or disease*)) or "paralysis agitans" or amnesia* or alzheimer* or aphasi* or "diffuse neurofibrillary tangle*" or "lobar degeneration*" or "acquired dyslexia" or alexia or cadasil or "cada sil" or "cerebral autosomal dominant arteriopathy" or "lewy body disease*" or (pick* adj1 (complex* or disease* or disorder* or syndrome*)) or huntington* or "juvenile chorea" or "chorea major" or "kluver bucy" or (mental* adj3 deteriorat*) or ((neuro* or neural* or cognitive* or cognition* or consciousness or sleep* or wake*) adj3 (impair* or side effect* or | 1744271 |

| | | |
|---|---|---|
| | function* or disfunction* or dysfunction* or disorder* or degenerat* or disease*)) or "pseudo-dementia*" or pseudodementia* or "rett syndrome*" or senile* or tauopath* or tourette*).ti,ab. | |
| 33 | (((chronic kidney or chronic renal) adj2 (disease? or insufficienc* or failur?)) or CKD or chronic nephropath*).ti,ab. | 195752 |
| 34 | (anxiety* or depression* or schizo* or psychotic* or psychosis or psychoses or amnesia* or delirium* or gender dysphoria* or paraphilia* or hallucination* or paranoid or paranoia or paraphrenia or sexual dysfunction* or hysteria or phobia? or agoraphobia or neurasthenia or hypochondriasis or hypochondria or hypochondriac or adjustment reaction*).ti,ab. | 1146933 |
| 35 | ((manic or mania or depress*) adj3 episode*).ti,ab. | 24066 |
| 36 | ("anorexia nervosa" or bulimia or PTSD or ((bipolar* or eating or personality or mood* or conduct or impulse or dissociative or eating or neurotic or obsessive or compulsive or panic or phobic or neurocognitiv* or traumatic* or posttraumatic* or post-traumatic or stress* or neurocognitive* or amnestic* or behaviour* or behavior* or neurobehav* or delusion* or manic or mania or depress* or conversion or somatoform or gender identity or attention-deficit or hyperactivity or conduct or emotion* or tic or tics or sleep or sexual identity or dysthymic or alcohol-induced or drug-induced or social functioning) adj3 disorder*)).ti,ab. | 482640 |
| 37 | ((mental* or psychiatric* or psychological* or neuropsych*) adj3 (disorder* or ill or illness* or diagnosis or diagnoses)).ti,ab. | 279913 |
| 38 | ((intellectual* or developmental* or neurodevelopmental or learning) adj3 (disabilit* or disorder* or syndrome*)).ti,ab. | 107822 |
| 39 | (or/7-38) and (COVID or COVID-19 or COVID19 or coronavirus* or corona virus* or SARS-CoV-2 or SARS-CoV2 or SARSCoV-2 or SARSCoV2).ti. [CONDITIONS 1] | 108329 |
| 40 | 6 and 39 [AFTER COVID CONDITIONS 1] | 31839 |
| 41 | inflammatory bowel disease/ | 67985 |
| 42 | ("inflammatory bowel disease*" or (IBD and "inflammatory bowel")).ti,ab. | 113768 |
| 43 | colitis/ or collagenous colitis/ or enterocolitis/ or lymphocytic colitis/ or microscopic colitis/ or ulcerative colitis/ | 154993 |
| 44 | (colitis or enterocolitis).ti,ab. | 156485 |
| 45 | Crohn disease/ or "crohn* disease".ti,ab. | 127854 |
| 46 | irritable colon/ | 34206 |
| 47 | ("irritable bowel syndrome*" or "irritable colon syndrome*").ti,ab. | 27194 |
| 48 | or/41-47 [GI] | 346955 |
| 49 | autoimmune thyroiditis/ | 9511 |
| 50 | Graves disease/ | 25668 |

| | | |
|---|---|---|
| 51 | Hashimoto disease/ | 16189 |
| 52 | hyperthyroidism/ | 41404 |
| 53 | ("Autoimmune Thyroiditis" or (Hashimoto* adj (disease or thyroid*)) or "Lymphocytic Thyroiditis").ti,ab. | 16553 |
| 54 | (hyperthyroid* or "Graves disease" or "Grave's Disease" or "Basedow* Disease").ti,ab. | 46210 |
| 55 | hypothyroidism/ | 82066 |
| 56 | (Hypothyroid* or ((deficien* adj1 (thyroid or "Thyroid-Stimulating Hormone")) or "TSH Deficien*")).ti,ab. | 65161 |
| 57 | systemic lupus erythematosus/ | 118297 |
| 58 | ("Systemic Lupus Erythematosus" or (SLE and lupus)).ti,ab. | 94617 |
| 59 | psoriasis/ | 82554 |
| 60 | psoriatic arthritis/ | 33491 |
| 61 | (Psoriasis or "psoriatric arthritis").ti,ab. | 80376 |
| 62 | autoimmune vasculopathy/ | 106 |
| 63 | systemic autoimmune disease/ and vasculitis.ti,ab. | 63 |
| 64 | (central nervous system vasculitis/ or brain vasculitis/) and autoimmune.ti,ab. | 413 |
| 65 | ("auto-immune vasculitis" or "autoimmune vasculitis" or (("auto-immune vascular" or "autoimmune vascular") adj (disease* or disorder* or inflammat*))).ti,ab. | 557 |
| 66 | giant cell arteritis/ | 11233 |
| 67 | ("auto-immune vasculitis" or "autoimmune vasculitis" or (("auto-immune vascular" or "autoimmune vascular") adj (disease* or disorder* or inflammat*))).ti,ab. | 557 |
| 68 | ("Primary Central Nervous System Vasculitis" or "Primary CNS Vasculitis" or "Giant cell arteritis" or vasculitis).ti,ab. | 73313 |
| 69 | vitiligo/ or vitiligo.ti,ab. | 18340 |
| 70 | exp autoimmune skin disease/ | 27138 |
| 71 | (((skin or cutaneous or dermatological) adj3 (condition* or disorder* or disease*)) and autoimmune).ti,ab. | 7366 |
| 72 | or/49-71 [Other Autoimmune] | 511687 |
| 73 | (48 or 72) and (COVID or COVID-19 or COVID19 or coronavirus* or corona virus* or SARS-CoV-2 or SARS-CoV2 or SARSCoV-2 or SARSCoV2).ti. [GI, Other Autoimmune] | 9338 |

| | | |
|---|---|---|
| 74 | 6 and 73 [AFTER COVID CONDITIONS 2] | 3086 |
| 75 | *incidence/ or *prevalence/ or *risk/ or *odds ratio/ | 213569 |
| 76 | (incidence or incident or prevalent or "logistic regression$" or "cox regression$" or odds or ((prevalence or rate or rates or risk or risks or hazard) adj5 (relative or ratio$ or difference$ or proportional))).ti,ab,kw. | 3409989 |
| 77 | ((diagno$2 or likelihood or likely) adj3 (develop* or elevated or high or higher or heightened or increase* or increasing)).ti,ab,kw. | 144481 |
| 78 | ((new or newly) adj3 (onset or diagnos$2 or problem$1)).ti,ab,kw. | 219596 |
| 79 | Disease exacerbation/ or (exacerbat* or flare$1 or provoke$1 or provocation or trigger* or worsen*).ti,ab,kw. | 1192268 |
| 80 | prognosis/ | 703084 |
| 81 | *prognosis/ or (prognos?s or trajector*).ti,ab,kw. | 1047044 |
| 82 | or/75-81 [Risk] | 5787606 |
| 83 | observational study/ | 390647 |
| 84 | cohort analysis/ | 1215242 |
| 85 | longitudinal study/ | 220697 |
| 86 | follow up/ | 2242665 |
| 87 | retrospective study/ | 1679032 |
| 88 | exp case control study/ | 241713 |
| 89 | cross-sectional study/ | 661999 |
| 90 | quasi experimental study/ | 12971 |
| 91 | prospective study/ | 938949 |
| 92 | (observational adj3 (study or studies or design or analysis or analyses)).ti,ab,kw. | 390444 |
| 93 | cohort*.ti,ab,kw. | 1634727 |
| 94 | (prospective adj3 (study or studies or design or analysis or analyses)).ti,ab,kw. | 774220 |
| 95 | ((follow up or followup) adj3 (study or studies or design or analysis or analyses)).ti,ab,kw. | 151922 |
| 96 | ((longitudinal or long-term or longterm) adj3 (study or studies or design or analysis or analyses or data)).ti,ab,kw. | 371772 |
| 97 | (retrospective adj3 (study or studies or design or analysis or analyses or data or review)).ti,ab,kw. | 1223938 |
| 98 | (case-referent adj3 (study or studies or design or analysis or analyses)).ti,ab,kw. | 698 |

| | | |
|---|---|---|
| 99 | (population adj3 (study or studies or analysis or analyses)).ti,ab,kw. | 381209 |
| 100 | (descriptive adj3 (study or studies or design or analysis or analyses)).ti,ab,kw. | 186840 |
| 101 | ((multidimensional or (multi adj dimensional)) adj3 (study or studies or design or analysis or analyses)).ti,ab,kw. | 6276 |
| 102 | ((cross-sectional or cross-sectional or crosssectional or crossectional) adj3 (study or studies or design or research or analysis or analyses or survey or findings)).ti,ab,kw. | 624742 |
| 103 | ((natural adj experiment) or (natural adj experiments)).ti,ab,kw. | 3897 |
| 104 | (quasi-experiment* or quasi-experiment*).ti,ab,kw. | 28548 |
| 105 | ((non-experiment$2 or nonexperiment$2) adj3 (study or studies or design or analys?s)).ti,ab,kw. | 2456 |
| 106 | (prevalence adj3 (study or studies or analys?s)).ti,ab,kw. | 78356 |
| 107 | control group/ | 110806 |
| 108 | case control study/ | 223217 |
| 109 | ((case or group* or study or patient) adj3 (control$2 or compar* or match* or series)).ti,ab,kw. | 2498026 |
| 110 | population surveillance/ | 470 |
| 111 | (surveillance adj3 (continuous or continual or continued or data or ongoing or population or research or study)).ti,ab,kw. | 50082 |
| 112 | or/83-111 | 8426301 |
| 113 | 40 and 82 and 112 [AFTER COVID CONDITIONS 1 + IPRE + STUDY DESIGN APPLIED] | 9625 |
| 114 | limit 113 to dc=20220701-20240901 [CONDITIONS 1 Update] | 6095 |
| 115 | 74 and 82 and 112 [AFTER COVID CONDITIONS 1 + IPRE + STUDY DESIGN APPLIED] | 904 |
| 116 | limit 115 to yr="2020 -Current" [CONDITIONS 2 date limit] | 904 |
| 117 | 114 or 116 [AFTER COVID CONDITIONS] | 6641 |
| 118 | (exp animal/ or exp animal model/) not human/ | 5573278 |
| 119 | ((animal or animals or canine* or dog or dogs or feline or hamster* or lamb or lambs or mice or monkey or monkeys or mouse or murine or pig or pigs or piglet* or porcine or primate* or rabbit* or rats or rat or rodent* or sheep* or veterinar*) not (human* or patient*)).mp. | 5114241 |
| 120 | 117 not (118 or 119) [ANIMAL-ONLY STUDIES REMOVED] | 6636 |
| 121 | (Case Reports.pt. or (case report? or case study or case presentation).ti.) not (review* or trial*).ti,ab,kw,hw. | 378972 |
| 122 | (comment or editorial or news or newspaper article).pt. | 821218 |

| | | |
|---|---|---|
| 123 | (conference abstract* or conference paper*).pt. | 5998453 |
| 124 | 120 not (or/121-123) [CASE STUDIES, OPINION PIECES, CONFERENCE ABS REMOVED] | 4460 |
| 125 | 124 not pandemic.ti. | 3663 |
| 126 | remove duplicates from 125 | 3635 |

#### Appendix B. List of studies included in a systematic review update examining new diagnoses and exacerbations of chronic conditions after SARS-CoV-2 infection

Table S1. Study details of studies included in a systematic review update examining new diagnoses and exacerbations of chronic conditions after SARS-CoV-2 infection.

| **Author year**<br><br>**Country**<br><br>**Data source** | **Study design**<br><br>**Comparator timing**<br><br>**Test required for SARS-CoV-2/ comparator group** | **SARS-CoV-2 infection period**<br><br>**Acute infection care setting**<br><br>**% Hospitalized/ % ICU** | **Total sample size**<br><br>**Age range in years**<br><br>**% Female** | **Mean (SD) or median (IQR) follow-up**<br><br>**Outcome ascertainment timing** | **Outcomes** |
|---|---|---|---|---|---|
| Abel 2021[1]<br><br>United Kingdom<br><br>Clinical Practice Research Datalink Aurum | Retrospective cohort<br><br>Concurrent<br><br>Yes/No | Feb 2020-Dec 2020<br><br>Outpatient<br><br>NA | 11,923,105<br><br>16-80+<br><br>50% | 6.3 (4.0-9.3) weeks<br><br>Up to 10 months | Mental illness (composite of anxiety disorders, depression, psychosis) |
| Ayoubkhani 2021[2]<br><br>United Kingdom<br><br>Hospital Episode Statistics Admitted Patient Care and General Practice Extraction Service Data for Pandemic Planning and Research | Retrospective cohort<br><br>Concurrent<br><br>No/No | Jan 2020-Aug 2020<br><br>Inpatient<br><br>100%/10% | 95,560<br><br>0-70+<br><br>45% | Cov: 140 (50) days<br>Con: 153 (33) days<br><br>Up to 10 months after discharge | Chronic kidney disease; Diabetes (composite of type 1 & type 2); Respiratory disorders (NOS) |
| Battistoni 2024[3]<br><br>Italy<br><br>COoperativa di MEdicina GENerale | Retrospective cohort<br><br>Historical<br><br>NR/NA | Jan 2020-Dec 2022<br><br>Mixed<br><br>NR/NR | 63528<br><br>19-65+<br><br>55% | NR<br><br>Up to 3 years | Cardiovascular disorders (acute coronary disease, arrhythmias, heart failure); Stroke (NOS) |
| Bsteh 2022[4]<br><br>Austria<br><br>Austrian MS-COVID-19 registry | Retrospective cohort<br><br>Concurrent<br><br>Yes/NR | Jan 2020-Jun 2021<br><br>Mixed<br><br>10%/NR | 422<br><br>Mean (SD) COV 42.6 (12.2); con 43.4 (12.6)<br><br>69% | Minimum 6 months<br><br>Up to 24 months | Neurological disorders (multiple sclerosis; exacerbation) |

| Author year<br><br>Country<br><br>Data source | Study design<br><br>Comparator timing<br><br>Test required for SARS-CoV-2/ comparator group | SARS-CoV-2 infection period<br><br>Acute infection care setting<br><br>% Hospitalized/ % ICU | Total sample size<br><br>Age range in years<br><br>% Female | Mean (SD) or median (IQR) follow-up<br><br>Outcome ascertainment timing | Outcomes |
|---|---|---|---|---|---|
| Chang 2023[5]<br><br>USA<br><br>TriNetX US Collaborative Network | Retrospective cohort<br><br>Concurrent<br><br>Yes/No | Jan 2020-Dec 2021<br><br>Mixed<br><br>NR/NR | 3,814,479<br><br>18-65+<br><br>57% | NR<br><br>30 days to 6 months | Autoimmune disorders (connective tissue diseases, celiac, inflammatory rheumatic diseases); Diabetes (type 1); Inflammatory bowel diseases (NOS) |
| Chevinksy 2021[6]<br><br>USA<br><br>Premier Healthcare Database Special COVID-19 Release | Prospective cohort<br><br>Concurrent<br><br>No/NR | Mar 2020-Jun 2020<br><br>Mixed<br><br>11%/ NR | 305 (<18 y only)<br><br>0-17 (adult data not stratified by age) | NR<br><br>30 to 120 days | 31 different conditions across 7 categories. Reports “Children with COVID-19 were not more likely to experience new diagnoses than children without COVID-19.” Not included in meta-analysis, as no effect sizes or variances were reported and attempts to contact authors were unsuccessful. |
| Choi 2023[7]<br><br>Korea<br><br>Health Insurance Review and Assessment Service database | Retrospective cohort<br><br>Concurrent<br><br>NR/No | Jan2020-Sep 2021<br><br>Mixed<br><br>NR/NR | 1,392,720<br><br>19-89<br><br>50% | 12.5 months<br><br>30 days to 28 months | Diabetes (type 1) |
| Cohen 2022[8]<br><br>USA<br><br>UnitedHealth Group Clinical Research Database | Retrospective cohort<br><br>Concurrent<br><br>No/No | Jan 2020-Dec 2020<br><br>Mixed<br><br>13%/NR | 174,674<br><br>65+<br><br>58% | 78 (30-175) days<br><br>21 to 141 days | Autoimmune disorders (atopic dermatitis); Cardiovascular disorders (acute coronary disease, cardiogenic shock, cardiac arrhythmia, cardiomyopathy, congestive heart failure, coronary disease, hypertension, myocardial infarction, tachycardia); Chronic kidney disease; Diabetes (type 2); Mental illness (any mental health diagnosis, psychosis); Neurological disorders (dementia, encephalopathy, epilepsy Guillain-Barre syndrome, migraine, peripheral neuropathy); Respiratory disorders (chronic respiratory failure, interstitial lung disease); Sleep disorders (obstructive sleep apnea); Stroke (ischemic stroke) |

| Author year<br><br>Country<br><br>Data source | Study design<br><br>Comparator timing<br><br>Test required for SARS-CoV-2/ comparator group | SARS-CoV-2 infection period<br><br>Acute infection care setting<br><br>% Hospitalized/ % ICU | Total sample size<br><br>Age range in years<br><br>% Female | Mean (SD) or median (IQR) follow-up<br><br>Outcome ascertainment timing | Outcomes |
|---|---|---|---|---|---|
| Daugherty 2021[9]<br><br>USA<br><br>UnitedHealth Group Clinical Discovery Database | Retrospective cohort<br><br>Concurrent<br><br>No/No | Up to Oct 2020<br><br>Mixed<br><br>8%/1% | 533,172<br><br>18-65<br><br>53% | 87 (45-124) days<br><br>21 to 141 days | Autoimmune disorders (atopic dermatitis); Cardiovascular disorders (acute coronary disease, arrythmia, cardiogenic shock, cardiomyopathy, congestive heart failure, coronary disease, hypertension, myocardial infarction, tachycardia); Chronic kidney disease; Diabetes (type 2); Mental illness (any mental health diagnosis, anxiety, depression & mood disorders; psychosis); Neurological disorders (Alzheimer's disease, dementia, encephalopathy, Guillain-Barre syndrome, migraine, peripheral neuropathy, seizure); Respiratory disorders (chronic respiratory failure, interstitial lung disease); Sleep disorders (obstructive sleep apnea); Stroke (hemorrhagic stroke, ischemic stroke) |
| Donnachie 2022[10]<br><br>Germany<br><br>Bavarian Association of Statutory Health Insurance Physicians claims database | Retrospective cohort<br><br>Concurrent<br><br>Yes/Yes | Jan 2020-Jun 2021<br><br>Outpatient<br><br>NA | 454,649<br><br>0-90+<br><br>54% | NR<br><br>Up to 2 years | Mental illness (composite of anxiety & mood disorders); Neurological disorders (chronic fatigue syndrome, mild cognitive impairment) |
| Gollop 2024[11]<br><br>Germany<br><br>IQVIA Disease Analyzer database | Retrospective cohort<br><br>Concurrent<br><br>NR/No | Jan 2020-Nov 2021<br><br>Mixed<br><br>NR/NR | 16,258<br><br>65-81+<br><br>59% | Cov: 404 (140) days<br>Con: 407 (142) days<br><br>Up to 26 months | Neurological disorders (dementia) |

| Author year<br><br>Country<br><br>Data source | Study design<br><br>Comparator timing<br><br>Test required for SARS-CoV-2/ comparator group | SARS-CoV-2 infection period<br><br>Acute infection care setting<br><br>% Hospitalized/ % ICU | Total sample size<br><br>Age range in years<br><br>% Female | Mean (SD) or median (IQR) follow-up<br><br>Outcome ascertainment timing | Outcomes |
|---|---|---|---|---|---|
| Gordon 2024[12]<br><br>USA<br><br>Healthcare Integrated Research Database | Retrospective cohort<br><br>Concurrent<br><br>Yes/Yes | May 2020-Mar 2021<br><br>Mixed<br><br>1%/NR | 447,684<br><br>0-17<br><br>50% | NR<br><br>2 weeks to 6 months | Cardiovascular disorders (atherosclerotic cardiovascular disease, arrhythmias); Diabetes (NOS); Inflammatory bowel diseases (NOS); Mental illness (composite of anxiety disorders, mood disorders, ADHD, & SUD*); Respiratory disorders (NOS) |
| Gronkjaer 2023[13]<br><br>Denmark<br><br>Danish Civil Registration System | Retrospective cohort<br><br>Concurrent<br><br>Yes/Yes | Mar 2020-Dec2021<br><br>Mixed<br><br>2%/<1% | 4,888,615<br><br>0-80+<br><br>49% | NR<br><br>Up to 22 months | Neurological disorders (any neurological disorder, epilepsy, myopathy, nerve, nerve root & plexus disorders, neuromuscular disorders, polyneuropathy) |
| Ho 2021[14]<br><br>Scotland<br><br>The Community Health Index register, Electronic Communication of Surveillance in Scotland, Rapid Preliminary Inpatient Data, Scottish Morbidity Record 01, and death certificates | Self-controlled case series<br><br>Self<br><br>Yes/NA | Mar 2020-Oct 2020<br><br>Inpatient<br><br>100%/NR | 792<br><br>Myocardial infarction (MI): >75; Ischemic stroke (IS): median (IRQ) 82 (73-87)<br><br>MI: 40%; IS: 52%% | NR<br><br>28 to 56 days | Cardiovascular disorders (myocardial infarction); Stroke (ischemic stroke) |
| Hong 2022[15]<br><br>USA<br><br>New York Crohn's and Colitis Organization | Prospective cohort<br><br>Concurrent<br><br>Yes/Yes | Feb 2020-Dec 2020<br><br>Mixed<br><br>16%/NR | 502<br><br>Median (IQR) 39 (30-52)<br><br>55% | Cov: 358 (238-459) days<br>Con: 417 (304-495) days<br><br>Up to 22 months | Inflammatory bowel diseases (composite of Crohn's disease & ulcerative colitis; exacerbations) |
| Huang 2024[16]<br><br>USA*<br><br>TriNetX Research Network | Retrospective cohort<br><br>Concurrent<br><br>Yes/Yes | Jan 2020-Dec 2021<br><br>Mixed<br><br>NR/NR | 2,758,622<br><br>65+<br><br>52% | NR<br><br>Up to 12 months | Autoimmune disorders (thyrotoxicosis, hypothyroid) |

| Author year<br><br>Country<br><br>Data source | Study design<br><br>Comparator timing<br><br>Test required for SARS-CoV-2/ comparator group | SARS-CoV-2 infection period<br><br>Acute infection care setting<br><br>% Hospitalized/ % ICU | Total sample size<br><br>Age range in years<br><br>% Female | Mean (SD) or median (IQR) follow-up<br><br>Outcome ascertainment timing | Outcomes |
|---|---|---|---|---|---|
| Jacob 2022 [17]<br><br>Germany<br><br>IQVIA Disease Analyzer database | Retrospective cohort<br><br>Concurrent<br><br>No/No | Mar 2020-May 2021<br><br>Outpatient<br><br>NA | 112,700<br><br>18-70+<br><br>52% | NR<br><br>Up to 14 months | Mental illness (anxiety disorders, depression) |
| Katsoularis 2023[18]<br><br>Sweden<br><br>SmiNet | Retrospective cohort<br><br>Self<br><br>Yes/NA | Feb 2020-May 2021<br><br>Mixed<br><br>36%/10% | 6,703<br><br>18-85+<br><br>42% | NR<br><br>30 to 180 days | Cardiovascular disorders (arrhythmias) |
| Kim 2024[19]<br><br>South Korea, Japan<br><br>JMDC (Japan cohort) | Retrospective cohort<br><br>Concurrent<br><br>Yes/NR | Jan 2020-Dec 2021<br><br>Mixed<br><br>NR/NR | 225,313<br><br>20-60+<br><br>63% | Median, Cov: 11.3 months; Con: 12.1 months<br><br>30 days to 24 months | Autoimmune disorders (inflammatory rheumatic diseases) |
| Kompaniyets 2022[20]<br><br>USA<br><br>HealthVerity | Retrospective cohort<br><br>Concurrent<br><br>No/No | Mar 2020-Nov 2021<br><br>Mixed<br><br>NR/NR | 3,125,676<br><br>0-17<br><br>50% | NR<br><br>60 to 365 days | Cardiovascular disorders (cardiac dysrhythmias); Chronic kidney disease; Diabetes (type 1, type 2); Mental illness (anxiety disorders, mood disorder); Musculoskeletal disorders (muscle disorders); Neurological disorders (nervous system disorder); Respiratory disorders (asthma); Sleep disorders (composite NOS); Stroke (NOS) |
| Kyung 2024 [21]<br><br>South Korea, Japan<br><br>K-COV-N cohort (South Korea); JMDC (Japan) | Retrospective cohort<br><br>Concurrent<br><br>Yes/NR | Jan 2020-Dec 2021<br><br>Mixed<br><br>South Korea: NR/15%; Japan: NR/NR | 5,731,321<br><br>20-60+<br><br>South Korea: 49%; Japan 39% | NR<br><br>30 days to >6 months | Autoimmune disorders (alopecia) |

| Author year<br><br>Country<br><br>Data source | Study design<br><br>Comparator timing<br><br>Test required for SARS-CoV-2/ comparator group | SARS-CoV-2 infection period<br><br>Acute infection care setting<br><br>% Hospitalized/ % ICU | Total sample size<br><br>Age range in years<br><br>% Female | Mean (SD) or median (IQR) follow-up<br><br>Outcome ascertainment timing | Outcomes |
|---|---|---|---|---|---|
| Lam 2023[22]<br><br>UK<br><br>United Kingdom Biobank | Retrospective cohort<br><br>Concurrent<br><br>Yes/No | Mar 2020-May 2021<br><br>Mixed<br><br>NR/NR | 815,910<br><br>18-65+<br><br>55% | 243 (212-294) days<br><br>21 days to 32 months | Cardiovascular disorders (atrial fibrillation, coronary artery disease, heart failure, myocardial infarction); Chronic kidney disease (end-stage renal disease); Mental illness (anxiety, post-traumatic stress disorder, psychotic disorders); Stroke (NOS) |
| Lee 2023 [23]<br><br>Korea<br><br>Korean National Health Insurance claims-based database | Retrospective cohort<br><br>Historical<br><br>Yes/NA | Oct 2020-Sep 2021<br><br>Inpatient<br><br>100%/1% (mechanical ventilation) | 154,302<br><br>18-65+<br><br>51% | Cov: 169 (128–287) days; Con: 277 (142–376) days<br><br>30 days to 15 months | Cardiovascular disorders (cardiac arrest, congestive heart failure, dysrhythmia, myocardial infarction); Stroke (NOS) |
| Lee 2024 [24]<br><br>Korea<br><br>Korean National Health Insurance claims-based database | Retrospective cohort<br><br>Concurrent<br><br>Yes/No | Oct 2020-Dec 2021<br><br>Mixed<br><br>15%/NR | 21,416<br><br>20-70+<br><br>61% | 87 (33-205) days<br><br>14 days to 15 months | Respiratory disorders (asthma; exacerbations) |
| Lim 2023 [25]<br><br>Korea<br><br>Korean National Health Insurance claims-based database | Retrospective cohort<br><br>Concurrent<br><br>Yes/No | Oct 2020-Dec 2021<br><br>Mixed<br><br>NR/NR | 6,489,467<br><br>18-65+<br><br>50% | Cov: 119.7 (117.9) days; Con: 121.4 (118.7) days<br><br>Up to 15 months | Autoimmune disorders (Adult-onset still disease, alopecia areata, alopecia totalis, ankylosing spondylitis, Behcet’s disease, psoriasis, rheumatoid arthritis, sarcoidosis, Sjorgren’s syndrome, vitiligo); Cardiovascular disorders (congestive heart failure, myocardial infarction); Inflammatory bowel diseases (Crohn’s disease, ulcerative colitis); Stroke (NOS) |

| Author year<br><br>Country<br><br>Data source | Study design<br><br>Comparator timing<br><br>Test required for SARS-CoV-2/ comparator group | SARS-CoV-2 infection period<br><br>Acute infection care setting<br><br>% Hospitalized/ % ICU | Total sample size<br><br>Age range in years<br><br>% Female | Mean (SD) or median (IQR) follow-up<br><br>Outcome ascertainment timing | Outcomes |
|---|---|---|---|---|---|
| Lin 2024 [26]<br><br>USA<br><br>TriNetX US Collaborative Network | Retrospective cohort<br><br>Concurrent<br><br>Yes/Yes | Jan 2020-Jun 2022<br><br>Mixed<br><br>NR/NR | 3,867,758<br><br>65+<br><br>56% | NR<br><br>2 weeks to 12 months | Sleep disorders (obstructive sleep apnea) |
| Ma 2024 [27]<br><br>UK<br><br>UK Biobank database | Retrospective cohort<br><br>Concurrent<br><br>NR/NR | Jan 2020-Oct 2022<br><br>Mixed<br><br>7%/1% | 471,982<br><br>37-73<br><br>55% | 254 (184-366) days<br><br>30 days to 34 months | Inflammatory bowel diseases (NOS) |
| Meng 2024 [28]<br><br>UK<br><br>UK Biobank database | Retrospective cohort<br><br>Concurrent<br><br>NR/No | Jan 2020-Sep 2022<br><br>Mixed<br><br>7%/NR | 471,982<br><br>37-73<br><br>55% | 300 (184-364) days<br><br>30 days to 24 months | Respiratory disorders (asthma, bronchiectasis, chronic obstructive pulmonary disease, interstitial lung disease) |
| Mizrahi 2023 [29]<br><br>Israel<br><br>Maccabi Healthcare Services | Retrospective cohort<br><br>Concurrent<br><br>Yes/No | Mar 2020-Oct 2021<br><br>Outpatient<br><br>NA | 599,740<br><br>0-60+<br><br>51% | NR<br><br>1 to 12 months | Autoimmune disorders (celiac); Cardiovascular disorders (cardiac arrhythmias, congestive heart failure, hypertension, ischemic heart disease); Diabetes (composite of type 1 & type 2); Mental illness (anxiety, depression, psychosis); Neurological disorders (epilepsy, myoneural junction or muscle disease, nerve root and plexus disorders, neuropathies, trigeminal neuralgia); Respiratory disorders (pulmonary disease, respiratory failure); Sleep disorders (insomnia); Stroke (composite of ischemic stroke and TIA) |

| Author year<br><br>Country<br><br>Data source | Study design<br><br>Comparator timing<br><br>Test required for SARS-CoV-2/ comparator group | SARS-CoV-2 infection period<br><br>Acute infection care setting<br><br>% Hospitalized/ % ICU | Total sample size<br><br>Age range in years<br><br>% Female | Mean (SD) or median (IQR) follow-up<br><br>Outcome ascertainment timing | Outcomes |
|---|---|---|---|---|---|
| Moniti 2024 [30]<br><br>Italy<br><br>Multiple Sclerosis Center of the IRCCS San Raffaele Hospital | Prospective cohort<br><br>Concurrent<br><br>Yes/NR | Mar 2020-Mar 2021<br><br>Mixed<br><br>1%/NR | 322<br><br>Median (IQR) Cov: 41 (33; 49);<br>Con: 45 (35; 52)<br><br>56% | NR<br><br>Up to 24 months | Neurological disorders (multiple sclerosis; exacerbations) |
| Murata 2024 [31]<br><br>Japan<br><br>Longevity Improvement and Fair Evidence Study | Retrospective cohort<br><br>Concurrent<br><br>Yes/No | Mar 2021-Dec 2022<br><br>Mixed<br><br>15%/NR | 22,414<br><br>65+<br><br>54% | NR<br><br>Up to 3 months | Mental illness (overall psychiatric disorders, anxiety disorders, mood disorders, psychotic disorders); Neurological disorders (organic mental disorders); Sleep disorders (insomnia) |
| Naveed 2023 [32]<br><br>Canada<br><br>British Columbia COVID-19 Cohort | Retrospective cohort<br><br>Concurrent<br><br>Yes/Yes | Jan 2020-Dec 2021<br><br>Mixed<br><br>NR/NR | 629,935<br><br>18-65+<br><br>51% | 257 (102-356) days<br><br>30 days to 24 months | Diabetes (NOS) |
| Nersesjan 2023 [33]<br><br>Denmark<br><br>Danish Civil Registration System | Retrospective cohort<br><br>Concurrent<br><br>Yes/Yes | Mar 2020-Dec 2021<br><br>Mixed<br><br>NR/NR | 4,152,792<br><br>18-65+<br><br>50% | NR<br><br>Up to 24 months | Mental illness (any mental disorder) |

| Author year<br><br>Country<br><br>Data source | Study design<br><br>Comparator timing<br><br>Test required for SARS-CoV-2/ comparator group | SARS-CoV-2 infection period<br><br>Acute infection care setting<br><br>% Hospitalized/ % ICU | Total sample size<br><br>Age range in years<br><br>% Female | Mean (SD) or median (IQR) follow-up<br><br>Outcome ascertainment timing | Outcomes |
|---|---|---|---|---|---|
| Nielsen 2024 [34]<br><br>Denmark<br><br>Danish Civil Registration System | Retrospective cohort<br><br>Concurrent<br><br>Yes/No | Mar 2020-Jan 2023<br><br>Mixed<br><br>1%/NR | 3,239,008<br><br>12-70+<br><br>48% | NR<br><br>1 to 35 months | Mental illness (composite of ADHD, anxiety, autism spectrum disorder *, bipolar disorders, depression, eating disorders, personality disorders, schizophrenia spectrum disorders, stress-related disorders, SUD*); Neurological disorders (cognition & memory disorders, extrapyramidal and movement disorders, multiple sclerosis, peripheral system disorders, systematic atrophies affecting the central nervous system); Stroke (composite of hemorrhagic stroke, ischemic stroke, & TIA) |
| Noorzae 2023 [35]<br><br>Denmark<br><br>Danish Civil Registration System | Retrospective cohort<br><br>Concurrent<br><br>Yes/Yes | Mar 2020-Aug 2022<br><br>Mixed<br><br>NR/NR | 1,239,894<br><br>0-17<br><br>49% | NR<br><br>30 days to 30 months | Diabetes (type 1) |
| Oh 2024 [36]<br><br>South Korea, Japan, UK<br><br>National Health Insurance Service (Korean cohort); JDMC (Japanese cohort); UK Biobank (UK) | Retrospective cohort<br><br>Concurrent<br><br>Yes/No | Jan 2020-Dec 2021<br><br>Mixed<br><br>South Korea: 16%/NR; Japan: NR/NR | 3,377,185<br><br>20-60+<br><br>South Korea: 45%; Japan 33% | NR<br><br>30 days to 24 months | Autoimmune disorders (allergic rhinitis, atopic dermatitis, food allergy); Respiratory disorders (asthma) |

| Author year<br><br>Country<br><br>Data source | Study design<br><br>Comparator timing<br><br>Test required for SARS-CoV-2/ comparator group | SARS-CoV-2 infection period<br><br>Acute infection care setting<br><br>% Hospitalized/ % ICU | Total sample size<br><br>Age range in years<br><br>% Female | Mean (SD) or median (IQR) follow-up<br><br>Outcome ascertainment timing | Outcomes |
|---|---|---|---|---|---|
| Pajor 2022 [37]<br><br>USA<br><br>PEDSnet | Retrospective cohort<br><br>Concurrent<br><br>Yes/Yes | Mar 2020-Feb 2022<br><br>Mixed<br><br>5%/1% | 557,418<br><br>0-20<br><br>47% | Cov: 0.43 (0.15-1.11) years;<br>Con: 0.78 (0.38-1.27) years<br><br>Up to 24 months | Autoimmune disorders (autoinflammatory syndromes, immunity disorders; vitiligo); Cardiovascular disorders (heart failure, cardiomyopathy, pulmonary heart disease, coronary atherosclerosis); Chronic kidney disease (NOS); Mental illness (gender dysphoria); Neurological disorders (communication & motor disorders; epilepsy; myopathies); Respiratory disorders (chronic obstructive pulmonary disease & bronchiectasis); Stroke (cerebrovascular disease)(exacerbations) |
| Park 2021 [38]<br><br>Korea<br><br>The National Health Insurance Service database | Retrospective cohort<br><br>Concurrent<br><br>No/No | Jan 2020-Jun 2020<br><br>Mixed<br><br>NR/15% | 260,883<br><br>20-80+<br><br>54% | NR<br><br>Up to 12 months | Mental illness (mental illness) |
| Pietropaolo 2022 [39]<br><br>USA<br><br>TriNetX COVID-19 Research Network | Retrospective cohort<br><br>Concurrent<br><br>No/No | Jan 2020-Jun 2021<br><br>Mixed<br><br>NR/NR | 4,070,133<br><br>0-30<br><br>45% | NR<br><br>1 day to 18 months | Diabetes (type 1, type 2) |
| Prahalad 2023 [40]<br><br>USA<br><br>PEDSnet | Retrospective cohort<br><br>Concurrent<br><br>Yes/Yes | Mar 2020-Dec 2021<br><br>Mixed<br><br>5%/NR | 2,404<br><br>0-21<br><br>51% | Minimum 6 months<br><br>28 days to 26 months | Diabetes (type 1; exacerbations) |
| Qureshi 2022 [41]<br><br>USA<br><br>Cerner Real-World Data | Retrospective cohort<br><br>Concurrent<br><br>Yes/Yes | Up to Jun 2021<br><br>Inpatient<br><br>100%/6% | 20,806<br><br>0-70+<br><br>51% | 182 (113-277) days<br><br>30 days to 9 months | Neurological disorders (dementia) |

| Author year<br><br>Country<br><br>Data source | Study design<br><br>Comparator timing<br><br>Test required for SARS-CoV-2/ comparator group | SARS-CoV-2 infection period<br><br>Acute infection care setting<br><br>% Hospitalized/ % ICU | Total sample size<br><br>Age range in years<br><br>% Female | Mean (SD) or median (IQR) follow-up<br><br>Outcome ascertainment timing | Outcomes |
|---|---|---|---|---|---|
| Rahman 2024 [42]<br><br>USA<br><br>National COVID-19 Cohort Collaborative Data Enclave platform | Retrospective cohort<br><br>Concurrent<br><br>NR/No | Jan 2020-May 2023<br><br>Inpatient<br><br>100%/NR | 441,601<br><br>17-70<br><br>47% | NR<br><br>22 to 180 days | Mental illness (schizophrenia spectrum & psychotic disorders) |
| Rao 2022 [43]<br><br>USA<br><br>PEDSnet | Retrospective cohort<br><br>Concurrent<br><br>Yes/Yes | Mar 2020-Oct 2021<br><br>Mixed<br><br>NR/NR | 659,286<br><br>0-20<br><br>47% | Cov: 4.6 (0.7) months; Con: 4.7 (0.7) months<br><br>28 days to 6 months | Autoimmune disorders (myositis); Mental illness (mental health treatment); Neurological disorders (communication/motor disorders) |
| Rezel-Potts 2022 [44]<br><br>England<br><br>CPRD database | Prospective cohort<br><br>Concurrent<br><br>No/No | Feb 2020-Jan 2021<br><br>Mixed<br><br>NR/NR | 857,300<br><br>Median (IQR) 35 (22-50)<br><br>56% | NR<br><br>5 to 52 weeks | Cardiovascular disorders (atrial arrhythmias, heart failure, myocardial infarction & ischemic heart disease); Diabetes (type 1, type 2); Stroke (NOS) |
| Robertson 2023 [45]<br><br>Sweden<br><br>Swedish Military Service Register | Retrospective cohort<br><br>Historical<br><br>Yes/NA | Jan 2020-Jan 2022<br><br>Mixed<br><br>5%/NR | 622,764<br><br>37-70<br><br>0% | NR<br><br>60 days to 25 months | Respiratory disorders (NOS) |

| Author year<br><br>Country<br><br>Data source | Study design<br><br>Comparator timing<br><br>Test required for SARS-CoV-2/ comparator group | SARS-CoV-2 infection period<br><br>Acute infection care setting<br><br>% Hospitalized/ % ICU | Total sample size<br><br>Age range in years<br><br>% Female | Mean (SD) or median (IQR) follow-up<br><br>Outcome ascertainment timing | Outcomes |
|---|---|---|---|---|---|
| Roessler 2022 [46]<br><br>Germany<br><br>6 statutory health insurance organizations | Retrospective cohort<br><br>Concurrent<br><br>Yes/No | Until Jun 2020<br><br>Mixed<br><br>5%/2% | 71,700<br><br>0-80+<br><br>59% | Cov: 236 (44) days<br>Con: 254 (36) days<br><br>3 months to 340 days | Cardiovascular disease (cardiac arrhythmias, heart failure, heart murmurs, myocardial infarction, other cardiac arrhythmias); Mental illness (adjustment disorder, anxiety disorders, depressive disorders, emotional and behavioural disorders, obsessive-compulsive disorder); Neurological conditions (chronic fatigue syndrome, developmental delay, dyslexia, epilepsy, facial nerve paralysis, migraine, movement disorders, other coordination disorders/ataxia, speech & language disorders); Stroke (NOS) |
| Salah 2022 [47]<br><br>USA<br><br>National COVID-19 Cohort Collaborative Data Enclave platform | Retrospective cohort<br><br>Concurrent<br><br>NR/No | Mar 2020-Mar 2022<br><br>Inpatient<br><br>100%/NR | 587,330<br><br>Mean (SD) age: Cov: 51 (22);<br>Con: 46 (23)<br><br>55% | 367 (190-536) days<br><br>Up to 24 months | Cardiovascular disorders (heart failure) |
| Schmitt 2024 [48]<br><br>Germany<br><br>German routine healthcare data covering inpatient and outpatient care, diagnoses, prescriptions and demographic data | Retrospective cohort<br><br>Concurrent<br><br>Yes/NR | Jan 2020-Dec 2020<br><br>Mixed<br><br>NR/NR | 2,549,696<br><br>0-80+<br><br>NR | NR<br><br>3 to 24 months | Autoimmune disorders (atopic dermatitis) |

| Author year<br><br>Country<br><br>Data source | Study design<br><br>Comparator timing<br><br>Test required for SARS-CoV-2/ comparator group | SARS-CoV-2 infection period<br><br>Acute infection care setting<br><br>% Hospitalized/ % ICU | Total sample size<br><br>Age range in years<br><br>% Female | Mean (SD) or median (IQR) follow-up<br><br>Outcome ascertainment timing | Outcomes |
|---|---|---|---|---|---|
| Smith 2024 [49]<br><br>Germany<br><br>IQVIA Disease Analyzer database | Retrospective cohort<br><br>Concurrent<br><br>NR/No | Mar 2020-Dec 2021<br><br>Outpatient<br><br>NA | 123,472<br><br>16-60+<br><br>49% | NR<br><br>Up to 12 months | Mental illness (depression) |
| Taquet 2022 [50]<br><br>USA<br><br>TriNetX Analytics Network, TriNetX's US Collaborative Network | Retrospective cohort<br><br>Concurrent<br><br>Yes/Yes | Jan 2020-Apr 2022<br><br>Mixed<br><br>NR/NR | 2,568,874<br><br>0-65+<br><br>58% | 7.3 months<br><br>1 to 24 months | Mental illness (anxiety disorder, mood disorder, psychotic disorder); Neurological disorders (epilepsy, dementia Guillain-Barre syndrome, myoneural junction/muscle disease, nerve/nerve root/plexus disorder, parkinsonism); Stroke (ischemic stroke) |
| Tartof 2022 [51]<br><br>USA<br><br>Vaccine Safety Datalink | Retrospective cohort<br><br>Concurrent<br><br>Yes/Yes | Mar 2020-Nov 2020<br><br>Inpatient<br><br>100%/NR | 255,718<br><br>0-85+<br><br>54% | NR<br><br>14 days to 6 months | Autoimmune disorders (alopecia); cardiovascular disorders (arrhythmias); Diabetes (NOS); Mental illness (anxiety, psychosis); Sleep disorders (NOS); Stroke (NOS) |
| Taylor 2024 [52]<br><br>UK<br><br>OpenSAFELY-TPP | Retrospective cohort<br><br>Concurrent<br><br>NR/No | Jan 2020-Dec 2021<br><br>Mixed<br><br>NR/NR | 16,699,943<br><br>18-90+<br><br>51% | NR<br><br>5 weeks to 24 months | Diabetes (type 2) |
| Tesch 2023 [53]<br><br>Germany<br><br>Post-COVID-19 Monitoring in Routine Health Insurance Data consortium | Retrospective cohort<br><br>Concurrent<br><br>Yes/No | Jan 2020-Dec 2020<br><br>Mixed<br><br>8%/2% | 2,201,764<br><br>0-80+<br><br>57% | NR<br><br>3 to 15 months | Autoimmune disorders (composite of 41 conditions) |

| Author year<br><br>Country<br><br>Data source | Study design<br><br>Comparator timing<br><br>Test required for SARS-CoV-2/ comparator group | SARS-CoV-2 infection period<br><br>Acute infection care setting<br><br>% Hospitalized/ % ICU | Total sample size<br><br>Age range in years<br><br>% Female | Mean (SD) or median (IQR) follow-up<br><br>Outcome ascertainment timing | Outcomes |
|---|---|---|---|---|---|
| Valdivieso-Martinez 2024 [54]<br><br>Spain<br><br>Valencian Health Agency electronic health record system (i.e., ABUCASIS) | Retrospective cohort<br><br>Concurrent<br><br>Yes/NR | Dec 2021-Feb 2022<br><br>Mixed<br><br>1%/NR | 16,656<br><br>0-17<br><br>49% | 179 (10.6) days<br><br>30 to 180 days | Cardiovascular disorders (tachycardia); Mental illness (adjustment disorder, anxiety disorder, major depressive disorder); Neurological disorders (migraine); Sleep disorders (insomnia) |
| Velasquez-Garcia 2024 [55]<br><br>Canada<br><br>British Columbia COVID-19 Cohort | Retrospective cohort<br><br>Concurrent<br><br>Yes/Yes | Jan 2020-Dec2021<br><br>Mixed<br><br>NR/NR | 649,320<br><br>Median (IQR) 33 (25-42)<br><br>52% | 260 (103-359) days<br><br>30 days to 24 months | Cardiovascular disorders (myocardial infarction); Stroke (composite NOS, TIA) |
| Wan 2023a [56]<br><br>UK<br><br>UK Biobank database | Retrospective cohort<br><br>Concurrent<br><br>NR/NR | Mar 2020-May 2021<br><br>Mixed<br><br>NR/NR | 601,623<br><br>60-84<br><br>56% | NR<br><br>21 days to 24 months | Cardiovascular disorders (acute coronary disease, atrial fibrillation, arrhythmias, atrial flutter, coronary heart disease, heart failure); Inflammatory bowel diseases (inflammatory bowel disease, Crohn’s disease); Mental illness (adjustment disorder, anxiety, PTSD); Neurological conditions (neurocognitive decline); Sleep disorders (composite NOS); Stroke (composite NOS, TIA) |
| Wan 2023b [57]<br><br>UK<br><br>UK Biobank database | Retrospective cohort<br><br>Concurrent<br><br>NR/NR | Mar 2020-May 2021<br><br>Mixed<br><br>NR/NR | 88,358<br><br>Mean (SD): 67.99 (8.38)<br><br>42% | 244.5 days<br><br>21 days to 24 months | Diabetes (type 1 or type 2; exacerbations) |

| Author year<br><br>Country<br><br>Data source | Study design<br><br>Comparator timing<br><br>Test required for SARS-CoV-2/ comparator group | SARS-CoV-2 infection period<br><br>Acute infection care setting<br><br>% Hospitalized/ % ICU | Total sample size<br><br>Age range in years<br><br>% Female | Mean (SD) or median (IQR) follow-up<br><br>Outcome ascertainment timing | Outcomes |
|---|---|---|---|---|---|
| Wang 2022a [58]<br><br>USA<br><br>US Collaborative Network in TriNetX | Retrospective cohort<br><br>Concurrent<br><br>Yes/No | Feb 2020-May 2021<br><br>Mixed<br><br>NR/NR | 1,381,784<br><br>65-85+<br><br>57% | NR<br><br>Up to 360 days | Neurological disorders (Alzheimer's disease) |
| Wang 2022b [59]<br><br>USA<br><br>US Collaborative Network in TriNetX | Retrospective cohort<br><br>Concurrent<br><br>Yes/No | Up to Mar 2021<br><br>Mixed<br><br>NR/NR | 6,245,282<br><br>20-65+<br><br>54% | NR<br><br>30 days to 12 months | Cardiovascular disorders (acute coronary disease, angina, atrial fibrillation & flutter, bradycardia, cardiac arrest, cardiogenic shock, cardiomyopathy, heart failure, ischemic cardiomyopathy, myocardial infarction, tachycardia, ventricular arrhythmias); Stroke (composite NOS, TIA) |
| Westman 2022 [60]<br><br>Sweden<br><br>SmiNET national register | Prospective cohort<br><br>Historical<br><br>Yes/NA | Feb 2020-Dec 2021<br><br>Mixed<br><br>NR/1% | 2,446,113<br><br>0-100+<br><br>51% | NR<br><br>Up to 22 months | Neurological conditions (epilepsy) |
| Xie 2022a [61]<br><br>USA<br><br>US Department of Veterans Affairs | Prospective cohort<br><br>Concurrent<br><br>Yes/No | Mar 2020-Sept 2021<br><br>Mixed<br><br>8%/2% | 4,299,721<br><br>Mean (SD): 60.92 (17.02)<br><br>12% | Cov: 352 (244-406) days; Con: 352 (245-406) days<br><br>30 days to 12 months | Diabetes (NOS) |
| Xie 2022b [62]<br><br>USA<br><br>US Department of Veterans Affairs | Prospective cohort<br><br>Concurrent<br><br>Yes/No | Mar 2020-Jan 2021<br><br>Mixed<br><br>11%/4% | 5,791,407<br><br>Mean (SD): 61.42 (15.64)<br><br>10% | Cov: 347 (317-440) days Con: 348 (318-441) days<br><br>30 days to 12 months | Cardiovascular disorders (dysrhythmia, ischemic heart disease); Stroke (composite of stroke & TIA) |

| Author year<br><br>Country<br><br>Data source | Study design<br><br>Comparator timing<br><br>Test required for SARS-CoV-2/ comparator group | SARS-CoV-2 infection period<br><br>Acute infection care setting<br><br>% Hospitalized/ % ICU | Total sample size<br><br>Age range in years<br><br>% Female | Mean (SD) or median (IQR) follow-up<br><br>Outcome ascertainment timing | Outcomes |
|---|---|---|---|---|---|
| Xu 2022 [63]<br><br>USA<br><br>US Department of Veterans Affairs | Prospective cohort<br><br>Concurrent<br><br>Yes/No | Mar 2020-Jan 2021<br><br>Mixed<br><br>11%/4% | 5,792,863<br><br>Mean (SD): 61.42 (15.64)<br><br>10% | Cov: 408 (378-500) days<br>Con: 348 (318-441) days<br><br>31 days to 12 months | Mental illness (anxiety disorders, major depressive disorders, psychotic disorders, stress/adjustment disorders); Neurological disorders (any neurologic outcome, Alzheimer's disease, disorders of the peripheral nerves, epilepsy, extrapyramidal and movement disorders, migraine/headache disorders); Stroke (composite of cerebral venous thrombosis, hemorrhagic stroke, ischemic stroke & TIA) |
| Yang 2024 [64]<br><br>USA<br><br>Medicare Geographic Variation files | Retrospective cohort<br><br>Concurrent<br><br>Yes/NR | April 2020-April-2021<br><br>Outpatient<br><br>NA/NA | 1,888,742<br><br>66+<br><br>58% | 18.5 (16.5-20.5) months<br><br>Up to 26 months | Cardiovascular disorders (abnormality of heart rhythm, acute myocardial infarction, atrial fibrillation & flutter, cardiac arrhythmia, cardiomyopathy, heart failure, ischemic heart disease); Stroke (hemorrhagic stroke, ischemic stroke, TIA) |
| Zang 2023 [65]<br><br>USA<br><br>INSIGHT and OneFlorida+ Clinical Research Networks | Retrospective cohort<br><br>Concurrent<br><br>Yes/Yes | Mar 2020-Nov 2021<br><br>Mixed<br><br>NR/NR | 560,752<br><br>20-65+<br><br>60% | NR<br><br>31 to 180 days | Cardiovascular disorders (heart failure); Fibromyalgia; Mental illness (anxiety disorders); Neurological disorders (dementia, encephalopathy, myopathies, polyneuropathies); Respiratory disorders (COPD); Sleep disorders (composite NOS) |
| Zappacosta 2022 [66]<br><br>Germany<br><br>IQVIA Disease Analyzer database | Retrospective cohort<br><br>Concurrent<br><br>Yes/NR | Mar 2020-Jun 2021<br><br>Mixed<br><br>NR/NR | 117,808<br><br>18-70+<br><br>54% | 363.7 (17.1) days<br><br>Up to 22 months | Cardiovascular disorders (cardiovascular events); Stroke (cerebrovascular events) |
| Zarifkar 2022 [67]<br><br>Denmark<br><br>Electronic health records (EPIC, version 2021, Wisconsin, USA) | Prospective cohort<br><br>Concurrent<br><br>Yes/Yes | Feb 2020-Nov 2021<br><br>Mixed<br><br>19%/NR | 919,731<br><br>18-80+<br><br>57% | NR<br><br>Up to 12 months | Neurological disorders (Alzheimer’s disease, Guillain-Barre syndrome, multiple sclerosis, myasthenia gravis, Parkinson's disease); Sleep disorders (narcolepsy); Stroke (ischemic stroke) |

| Author year<br><br>Country<br><br>Data source | Study design<br><br>Comparator timing<br><br>Test required for SARS-CoV-2/ comparator group | SARS-CoV-2 infection period<br><br>Acute infection care setting<br><br>% Hospitalized/ % ICU | Total sample size<br><br>Age range in years<br><br>% Female | Mean (SD) or median (IQR) follow-up<br><br>Outcome ascertainment timing | Outcomes |
|---|---|---|---|---|---|
| Zhang 2024 [68]<br><br>USA<br><br>RECOVER consortium | Retrospective cohort<br><br>Concurrent<br><br>Yes/Yes | Mar 2020-Mar 2023<br><br>Mixed<br><br>NR/NR | 1,213,322<br><br>0-21<br><br>49% | NR<br><br>28 days to 42 months | Cardiovascular disorders (arrhythmias, cardiac arrest, cardiogenic shock, cardiomyopathy, hypertension, heart failure) |
| Zhang-James 2024 [69]<br><br>USA<br><br>US TriNetX Research Network | Retrospective cohort<br><br>Concurrent<br><br>NR/NR | Jan 2020-Dec 2023<br><br>Mixed<br><br>NR/NR | 579,306<br><br>0-21<br><br>Children: 48%; Adolescents: 54% | NR<br><br>Up to 2 years | Mental illness (new mental disorder diagnosis, mood disorders, non-mood psychotic disorder) |

Note: COVID-19 and SARS-CoV-2 infection are used interchangeably. ADHD = attention deficit/hyperactivity disorder; con = control/comparator group; COPD = chronic obstructive pulmonary disorder; cov = SARS-CoV-2 infected group; COVID-19 = corona virus disease 2019; ICU = intensive care unit; IQR = interquartile range; NA = not applicable; NOS = not otherwise specified; NR = not reported; PTSD = post-traumatic stress disorder; SARS-CoV-2 = severe acute respiratory syndrome corona virus-2; SD = standard deviation; SUD = substance use disorder; TIA = transient ischemic attack

Table S2. Risk of bias assessment according to JBI's Cohort Studies tool

| Study | Q1 | Q2 | Q3 | Q4 | Q5 | Q6 | Q7 | Q8 | Q9 | Q10 | Q11 | Overall risk of bias |
|---|---|---|---|---|---|---|---|---|---|---|---|---|
| **Abel 2021** | Y | N | U | Y | Y | Y | Y | U | U | U | Y | Moderate |
| **Ayoubkhani 2021** | N | N | Y | Y | Y | Y | Y | Y | Y | NA | Y | High |
| **Battistoni 2024** | N | NA | U | Y | Y | U | Y | U | Y | NA | Y | Moderate |
| **Bsteh 2022** | U | N | U | U | U | U | Y | Y | N | N | Y | High |
| **Chang 2023** | Y | U | Y | Y | Y | Y | Y | N | Y | NA | Y | Moderate |
| **Chevinsky 2021** | U | U | Y | Y | Y | Y | Y | N | U | U | U | Moderate |
| **Choi 2023** | Y | N | U | U | U | N | Y | Y | U | U | U | High |
| **Cohen 2022** | Y | N | Y | Y | Y | Y | Y | Y | Y | NA | Y | Moderate |
| **Daugherty 2021** | Y | N | Y | Y | Y | Y | Y | Y | Y | Y | Y | Moderate |
| **Donnachie 2022** | Y | Y | Y | Y | Y | Y | Y | Y | N | Y | N | High |
| **Gollop 2023** | Y | Y | U | Y | Y | Y | Y | N | Y | NA | Y | Moderate |
| **Gordon 2024** | Y | Y | Y | Y | Y | U | Y | U | Y | NA | U | Low |
| **Gronkjaer 2023** | Y | Y | Y | U | U | Y | Y | N | Y | NA | Y | Moderate |
| **Ho 2021** | N | NA | N | Y | Y | Y | Y | N | Y | NA | Y | High |
| **Hong 2022** | Y | Y | Y | U | U | Y | Y | Y | U | U | U | Low |
| **Huang 2024** | Y | Y | U | Y | U | Y | Y | N | U | N | U | High |
| **Jacob 2022** | Y | N | Y | Y | U | Y | Y | U | U | Y | U | Moderate |
| **Katsoularis 2023** | N | NA | N | Y | Y | Y | Y | Y | Y | NA | Y | High |
| **Kim 2024** | Y | N | Y | U | U | Y | Y | N | Y | NA | U | High |
| **Kompaniyets 2022** | Y | N | Y | Y | U | Y | Y | Y | U | Y | Y | Moderate |
| **Kyung 2024** | Y | U | U | Y | U | Y | Y | U | U | N | U | Moderate |
| **Lam 2023** | Y | N | Y | U | U | Y | Y | Y | U | Y | U | Moderate |
| **Lee 2023** | N | NA | Y | Y | U | Y | Y | N | U | U | U | High |
| **Lee 2024** | Y | N | Y | Y | Y | Y | Y | N | Y | NA | U | High |
| **Lim 2023** | Y | N | Y | Y | Y | Y | Y | N | Y | NA | U | High |
| **Lin 2024** | Y | Y | Y | U | Y | Y | Y | N | U | N | Y | High |
| **Ma 2024** | Y | N | U | U | U | U | Y | Y | U | N | U | High |
| **Meng 2024** | Y | N | Y | U | Y | U | Y | Y | U | N | U | High |
| **Mizrahi 2023** | Y | N | Y | Y | Y | Y | Y | Y | Y | NA | Y | Moderate |
| **Moniti 2024** | Y | N | N | Y | Y | Y | U | Y | N | N | Y | High |
| **Murata 2024** | Y | N | Y | Y | Y | N | Y | N | U | Y | Y | High |
| **Naveed 2023** | Y | Y | Y | Y | U | Y | Y | Y | U | Y | Y | Low |
| **Nersesjan 2023** | Y | Y | Y | U | U | Y | Y | N | Y | NA | U | Moderate |
| **Nielsen 2024** | Y | N | Y | U | U | Y | Y | Y | Y | NA | Y | Moderate |
| **Noorzae 2023** | Y | Y | Y | U | U | Y | Y | U | Y | NA | U | Low |
| **Oh 2024** | Y | N | U | Y | Y | Y | Y | U | U | U | U | Moderate |
| **Pajor 2022** | Y | N | U | Y | Y | Y | Y | N | Y | NA | U | High |
| **Park 2021** | Y | N | U | Y | Y | Y | Y | Y | Y | NA | Y | Moderate |
| **Pietropaolo 2022** | Y | N | Y | Y | Y | Y | U | Y | U | U | Y | Moderate |
| **Prahalad 2023** | Y | N | U | Y | U | Y | U | Y | U | N | U | High |
| **Qureshi 2022** | Y | Y | Y | Y | Y | Y | Y | Y | U | U | U | Low |
| **Rahman 2024** | U | N | U | U | Y | Y | Y | N | U | N | Y | High |
| **Rao 2022** | Y | Y | Y | Y | Y | Y | Y | Y | U | U | Y | Low |
| **Rezel-Potts 2022** | Y | N | U | Y | Y | Y | Y | Y | Y | NA | Y | Moderate |
| **Robertson 2023** | Y | U | U | Y | Y | Y | Y | U | N | N | U | High |
| **Roessler 2022** | Y | N | U | Y | Y | Y | U | Y | U | Y | U | Moderate |
| **Salah 2022** | Y | N | U | U | Y | Y | U | Y | U | N | U | High |
| **Schmitt 2024** | Y | N | Y | U | U | Y | Y | U | U | Y | Y | Moderate |
| **Smith 2024** | Y | N | U | U | U | Y | Y | U | U | N | U | High |
| **Taquet 2022** | Y | N | Y | Y | Y | Y | Y | Y | Y | U | Y | Moderate |
| **Tartof 2022** | Y | Y | U | Y | Y | U | Y | Y | Y | NA | Y | Low |
| **Taylor 2024** | Y | N | Y | Y | Y | Y | Y | Y | U | Y | Y | Moderate |
| **Tesch 2023** | Y | N | Y | Y | Y | Y | Y | U | Y | Y | Y | Moderate |
| **Valdivieso-Martinez 2024** | Y | U | U | Y | U | Y | U | Y | Y | NA | N | Moderate |

| | | | | | | | | | | | | |
|---|---|---|---|---|---|---|---|---|---|---|---|---|
| **Velasquez-Garcia 2024** | Y | Y | Y | Y | Y | Y | Y | N | N | Y | Y | High |
| **Wan 2023** | Y | N | U | Y | Y | Y | Y | Y | U | Y | U | Moderate |
| **Wan 2024** | Y | N | U | Y | Y | Y | Y | N | U | Y | U | High |
| **Wang 2022a** | Y | N | Y | Y | Y | Y | Y | Y | U | U | Y | Moderate |
| **Wang 2022b** | Y | Y | Y | Y | Y | Y | Y | Y | Y | NA | Y | Low |
| **Westman 2022** | Y | N | Y | Y | Y | Y | Y | U | Y | NA | Y | Moderate |
| **Xie 2022a** | Y | N | Y | Y | Y | Y | Y | U | Y | NA | Y | Moderate |
| **Xie 2022b** | Y | N | Y | Y | Y | Y | Y | U | U | U | Y | Moderate |
| **Xu 2022** | Y | N | Y | Y | Y | Y | Y | U | U | U | Y | Moderate |
| **Yang 2024** | Y | N | Y | Y | Y | Y | Y | Y | U | Y | Y | Moderate |
| **Zang 2023** | Y | Y | Y | U | Y | Y | Y | Y | U | Y | Y | Low |
| **Zappacosta 2022** | Y | N | Y | Y | U | Y | Y | Y | U | Y | Y | Moderate |
| **Zarifkar 2022** | Y | Y | Y | Y | U | U | Y | Y | U | U | U | Low |
| **Zhang 2024** | Y | Y | Y | Y | Y | Y | Y | Y | N | Y | U | Moderate |
| **Zhang-James 2024** | U | N | Y | Y | Y | U | Y | U | U | Y | Y | Moderate |

Abbreviations: N = no; NA = not applicable; U = unsure; Y = yes

1. Were the two groups similar and recruited from the same population?
2. Were the exposures measured similarly to assign people to both exposed and unexposed groups?*
3. Was the exposure measured in a valid and reliable way?
4. Were confounding factors identified?
5. Were strategies to deal with confounding factors stated?
6. Were the groups/participants free of the condition/diagnosis of interest at the start of the study (or at the moment of exposure)?
7. Were the outcomes measured in a valid and reliable way?
8. Was the follow up time reported and sufficient to be long enough for outcomes to occur?
9. Was follow up complete, and if not, were the reasons for loss to follow up described and explored?
10. Were strategies to address incomplete follow up utilized?
11. Was appropriate statistical analysis used?

*Most studies got “No” for Q2 question because they relied on the *absence* of a positive SARS-CoV-2 test/diagnosis to identify the control group sample (i.e., the control group was not tested and we considered this differential ascertainment in exposure).

#### Table S3. Detailed summary of findings table

| Outcome | Subgroup<br>No. Studies | Relative findings<br>HR (95% CI) | Conclusion<br>Certainty for relative findings | Excess cases per 1000 people over 6 mos (95% CI) |
|---|---|---|---|---|
| **Autoimmune disorders** | | | | |
| Autoimmune disorders, overall | Outpatients/mixed, <18 y<br>5 studies | 1.13 (0.86 to 1.50) | Little-to-no difference<br>Low; B | NE |
| | Outpatients/mixed, 18-64 y<br>10 studies | 1.51 (1.31 to 1.75) | Small-to-moderate increase<br>Low; b, c[1] | 4.67 (2.84 to 6.87) |
| | Outpatients/mixed, ≥65 y<br>9 studies | 1.44 (1.23 to 1.69) | Small-to-moderate increase<br>Low; b, c[1] | 2.19 (1.14 to 3.43) |
| | Inpatients, 18-64 y<br>1 study | 1.04 (0.70 to 1.56) | Very low; c[2], D | NE |
| | Inpatients, ≥65 y<br>1 study | 1.14 (0.78 to 1.66) | Little-to-no difference<br>Low; c[2], d | NE |
| Allergic diseases | Outpatients/mixed, <18 y<br>1 study | 1.36 (1.21 to 1.53) | Very low; C[1,2], d | NE |
| | Outpatients/mixed, 18-64 y<br>3 studies | 1.61 (1.22 to 2.13) | Small-to-moderate increase<br>Low; c[1], d | 2.9 (1.04 to 5.36) |
| | Outpatients/mixed, ≥65 y<br>3 studies | 1.41 (1.08 to 1.85) | Very low; C[1,2], d | NE |
| | Inpatients, 18-64 y<br>1 study | 1.04 (0.70 to 1.56) | Very low; c[2], D | NE |
| | Inpatients, ≥65 y<br>1 study | 1.14 (0.78 to 1.66) | Very low; c[2], d | NE |
| Alopecia | Outpatients/mixed, <18 y<br>1 study | 0.96 (0.44 to 2.11) | Little-to-no difference<br>Low; D | NE |
| | Outpatients/mixed, 18-64 y<br>2 studies | 1.42 (1.17 to 1.73) | Small-to-moderate increase<br>Low; c[1], d | 0.35 (0.14 to 0.61) |
| | Outpatients/mixed, ≥65 y<br>1 study | 0.95 (0.40 to 2.24) | Little-to-no difference<br>Low; D | NE |
| Celiac disease | Outpatients/mixed, <18y<br>1 study | 0.75 (0.57 to 0.99) | Little-to-no difference<br>Moderate; d | NE |
| | Outpatients/mixed, 18-64 y<br>2 studies | 1.33 (0.96 to 1.85) | Small-to-moderate increase<br>Low; b, c[1] | 0.1 (-0.01 to 0.26) |
| | Outpatients/mixed, ≥65 y<br>1 study | 2.15 (1.41 to 3.28) | Small-to-moderate increase* [μ]<br>Moderate; c[1] | 0.37 (0.13 to 0.74) |

| Outcome | Subgroup<br>No. Studies | Relative findings<br>HR (95% CI) | Conclusion<br>Certainty for relative findings | Excess cases per 1000 people over 6 mos (95% CI) |
|---|---|---|---|---|
| Connective tissue disease | Outpatients/mixed, <18y<br>1 study | 2.59 (1.28 to 5.25) | Small-to-moderate increase* [µ]<br>Low; C[1,3] | NE (No CER available) |
| | Outpatients/mixed, 18-64 y<br>2 studies | 1.95 (1.08 to 3.54) | Small-to-moderate increase<br>Low; c[1], d | NE (No CER available) |
| | Outpatients/mixed, ≥65 y<br>1 study | 2.87 (1.84 to 4.47) | Small-to-moderate increase* [µ]<br>Moderate; c[1] | NE (No CER available) |
| Hyperthyroid | Outpatients/mixed, 18-64 y<br>1 study | 1.74 (1.63 to 1.86) | Small-to-moderate increase<br>Low; a, c[1] | 0.41 (0.35 to 0.47) |
| | Outpatients/mixed, ≥65 y<br>1 study | 1.97 (1.75 to 2.22) | Small-to-moderate increase<br>Low; a, c[1] | 1.09 (0.84 to 1.37) |
| Hypothyroid | Outpatients/mixed, 18-64 y<br>1 study | 2.01 (1.96 to 2.06) | Small-to-moderate increase* [µ]<br>Low; a, c[1] | 3.6 (3.42 to 3.78) |
| | Outpatients/mixed, ≥65 y<br>1 study | 2.21 (2.14 to 2.29) | Large increase<br>Low; a, c[1] | 15.85 (14.94 to 16.9) |
| Inflammatory rheumatic diseases | Outpatients/mixed, 18-64 y<br>3 studies | 1.61 (1.22 to 2.13) | Small-to-moderate increase<br>Low; a, c[1] | 18.95 (6.83 to 35.1) |
| | Outpatients/mixed, ≥65 y<br>2 studies | 1.95 (0.69 to 5.46) | Very low; a, c[1], d | NE |
| **Cardiovascular disorders** | | | | |
| Cardiovascular disorders, overall | Outpatients/mixed, <18 y<br>7 studies | 1.17 (1.13 to 1.21) | Little-to-no difference<br>Moderate; c[4] | NE |
| | Outpatients/mixed, 18-64 y<br>12 studies | 1.32 (1.12 to 1.55) | Small-to-moderate increase<br>Low; b, c[1] | 0.81 (0.3 to 1.38) |
| | Outpatients/mixed, ≥65 y<br>10 studies | 1.33 (1.14 to 1.54) | Small-to-moderate increase<br>Low; b, c[1] | 10.96 (4.65 to 17.93) |
| | Inpatients, 18-64 y<br>5 studies | 2.03 (1.13 to 3.66) | Small-to-moderate increase*[•]<br>Moderate; c[1] | 36.26 (4.58 to 93.65) |
| | Inpatients, ≥65 y<br>3 studies | 0.85 (0.29 to 2.49) | Little-to-no difference<br>Low; a, b | NE |
| Acute coronary disease | Outpatients/mixed, <18 y<br>4 studies | 1.45 (1.17 to 1.79) | Small to moderate increase<br>Low; c[1], d | 0.09 (0.03 to 0.15) |
| | Outpatients/mixed, 18-64 y<br>9 studies | 1.43 (1.14 to 1.81) | Small-to-moderate increase<br>Low; b, c[1] | 0.28 (0.09 to 0.52) |
| | Outpatients/mixed, ≥65 y<br>7 studies | 1.55 (1.27 to 1.89) | Small-to-moderate increase<br>Moderate; c[1] | 3.76 (1.85 to 6.09) |

| Outcome | Subgroup<br>No. Studies | Relative findings<br>HR (95% CI) | Conclusion<br>Certainty for relative findings | Excess cases per 1000 people over 6 mos (95% CI) |
|---|---|---|---|---|
| | Inpatients, 18-64 y<br>4 studies | 2.70 (0.90 to 8.06) | Small-to-moderate increase*•<br>Low; c[1], d | 3.96 (-0.23 to 16.45) |
| | Inpatients, ≥65 y<br>3 studies | 1.02 (0.30 to 3.49) | Very low; a, b, d | NE |
| Arrhythmias/<br>dysrhythmias | Outpatients/mixed, <18 y<br>7 studies | 1.28 (1.17 to 1.41) | Small-to-moderate increase<br>Low; b, c[1] | 0.7 (0.43 to 1.03) |
| | Outpatients/mixed, 18-64 y<br>8 studies | 1.56 (1.38 to 1.77) | Small-to-moderate increase<br>Low; b, c[1] | 1.84 (1.25 to 2.53) |
| | Outpatients/mixed, ≥65 y<br>8 studies | 1.43 (1.18 to 1.73) | Small-to-moderate increase<br>Low; b, c[1] | 7.26 (3.04 to 12.32) |
| | Inpatients, 18-64 y<br>2 studies | 2.22 (0.49 to 10.01) | Very low; c[1], D | NE |
| | Inpatients, ≥65 y<br>2 studies | 1.59 (0.38 to 6.75) | Very low; B, c[1] | NE |
| Cardiomyopathy | Outpatients/mixed, <18 y<br>1 study | 1.50 (1.19 to 1.90) | Small-to-moderate increase<br>Low; c[1], d | 0.14 (0.05 to 0.26) |
| | Outpatients/mixed, 18-64 y<br>2 studies | 2.81 (2.31 to 3.42) | Large increase<br>Moderate; c[1] | 1.26 (0.91 to 1.69) |
| | Outpatients/mixed, ≥65 y<br>3 studies | 1.57 (0.90 to 2.74) | Small-to-moderate increase<br>Low; b, c[1] | 3.35 (-0.59 to 10.22) |
| | Inpatients, 18-64 y<br>1 study | 5.63 (4.04 to 7.85) | Large increase<br>Moderate; c[1] | 13.78 (9.05 to 20.39) |
| | Inpatients, ≥65 y<br>2 studies | 1.40 (0.83 to 2.38) | Small-to-moderate increase<br>Low; b, c[1] | 5.13 (-2.18 to 17.68) |
| Heart failure | Outpatients/mixed, <18 y<br>2 studies | 1.51 (0.70 to 3.25) | Very low; c[1], D | NE |
| | Outpatients/mixed, 18-64 y<br>7 studies | 1.81 (1.40 to 2.34) | Small to moderate increase<br>Low; b, c[1] | 4.8 (2.37 to 7.94) |
| | Outpatients/mixed, ≥65 y<br>6 studies | 1.61 (1.22 to 2.12) | Small to moderate increase<br>Low; c[1], d | 6.71 (2.42 to 12.31) |
| | Inpatients, 18-64 y<br>3 studies | 1.91 (0.91 to 4.02) | Small to moderate increase<br>Low; c[1], d | 2.36 (-0.23 to 7.82) |
| | Inpatients, ≥65 y<br>2 studies | 1.15 (0.23 to 5.69) | Very low; a, D | NE |
| Hypertension | Outpatients/mixed, <18 y<br>2 studies | 1.47 (1.35 to 1.60) | Small to moderate increase<br>Moderate; c[1] | 0.96 (0.71 to 1.22) |

| Outcome | Subgroup<br>No. Studies | Relative findings<br>HR (95% CI) | Conclusion<br>Certainty for relative findings | Excess cases per 1000 people over 6 mos (95% CI) |
|---|---|---|---|---|
| | Outpatients/mixed, 18-64 y<br>2 studies | 1.30 (0.94 to 1.79) | Very low; B, c[1] | NE |
| | Outpatients/mixed, ≥65 y<br>2 studies | 1.47 (1.00 to 2.15) | Small-to-moderate increase<br>Low; b, c[1] | 2.06 (0 to 5.03) |
| | Inpatients, 18-64 y<br>1 study | 3.05 (2.75 to 3.38) | Large increase<br>Moderate; c[1] | 72.17 (61.61 to 83.79) |
| | Inpatients, ≥65 y<br>1 study | 2.77 (2.64 to 2.91) | Large increase<br>Moderate; c[1] | 189 (175.12 to 203.95) |
| **Chronic kidney disease** | | | | |
| Chronic kidney disease | Outpatients/mixed, <18 y<br>1 study | 1.07 (0.94 to 1.21) | Little-to-no difference<br>High, | NE |
| | Outpatients/mixed, 18-64 y<br>3 studies | 1.39 (1.00 to 1.95) | Small-to-moderate increase<br>Low; b, c[1] | 0.67 (0 to 1.62) |
| | Outpatients/mixed, ≥65 y<br>2 studies | 1.35 (1.20 to 1.52) | Small-to-moderate increase<br>Low; c[1], d | 3.77 (2.15 to 5.6) |
| | Inpatients, 18-64 y<br>2 studies | 3.13 (2.66 to 3.69) | Large increase<br>Moderate; c[1] | 10.55 (8.22 to 13.32) |
| | Inpatients, ≥65 y<br>2 studies | 1.66 (1.51 to 1.82) | Small-to-moderate increase<br>Moderate; c[1] | 19.95 (15.41 to 24.78) |
| **Diabetes** | | | | |
| Diabetes, overall | Outpatients/mixed, <18 y<br>6 studies | 1.10 (0.95 to 1.28) | Little-to-no difference<br>Moderate; b | NE |
| | Outpatients/mixed, 18-64 y<br>10 studies | 1.53 (1.29 to 1.81) | Small-to-moderate increase<br>Low; b, c[1] | 6.24 (3.42 to 9.54) |
| | Outpatients/mixed, ≥65 y<br>6 studies | 1.61 (1.36 to 1.92) | Small-to-moderate increase<br>Moderate; c[1] | 14.42 (8.51 to 21.74) |
| | Inpatients, 18-64 y<br>3 studies | 2.46 (1.45 to 4.17) | Small to moderate increase*•<br>Moderate; c[1] | 29.57 (9.11 to 64.2) |
| | Inpatients, ≥65 y<br>2 studies | 1.89 (1.03 to 3.44) | Small to moderate increase<br>Low; c[1], d | 27.48 (0.93 to 75.33) |
| Type 1 | Outpatients/mixed, <18 y<br>3 studies | 1.01 (0.79 to 1.28) | Little-to-no difference<br>Moderate; d | NE |
| | Outpatients/mixed, 18-64 y<br>2 studies | 1.84 (1.25 to 2.71) | Small to moderate increase<br>Low; b, c[1] | 0.04 (0.01 to 0.09) |
| | Outpatients/mixed, ≥65 y<br>1 study | 3.74 (3.26 to 4.29) | Large increase<br>Moderate; c[1] | NE (No CER available) |

| Outcome | Subgroup<br>No. Studies | Relative findings<br>HR (95% CI) | Conclusion<br>Certainty for relative findings | Excess cases per 1000 people over 6 mos (95% CI) |
|---|---|---|---|---|
| Type 2 | Outpatients/mixed, <18 y<br>2 studies | 1.17 (1.11 to 1.23) | Little-to-no difference<br>High | NE |
| | Outpatients/mixed, 18-64 y<br>4 studies | 1.38 (1.22 to 1.55) | Small to moderate increase<br>Moderate; d | 1.02 (0.59 to 1.48) |
| | Outpatients/mixed, ≥65 y<br>2 studies | 1.44 (1.23 to 1.70) | Small to moderate increase<br>Moderate; d | 2.71 (1.41 to 4.31) |
| | Inpatients, 18-64 y<br>1 study | 3.67 (3.06 to 4.39) | Large increase<br>Moderate; $c^1$ | 28.68 (22.13 to 36.42) |
| | Inpatients, ≥65 y<br>1 study | 3.03 (2.72 to 3.38) | Large increase<br>Moderate; $c^1$ | 48.08 (40.74 to 56.37) |
| **Fibromyalgia** | | | | |
| Fibromyalgia | Outpatients/mixed, 18-64 y<br>1 study | 1.17 (1.09 to 1.26) | Little-to-no difference<br>Moderate; d | NE |
| | Outpatients/mixed, ≥65 y<br>1 study | 1.30 (1.18 to 1.42) | Small-to-moderate increase<br>Low; $c^1$, d | 11.28 (6.77 to 15.8) |
| **Inflammatory bowel disease** | | | | |
| Inflammatory bowel disease, overall | Outpatients/mixed, <18 y<br>1 study | 0.96 (0.90 to 1.02) | Little-to-no difference<br>High | NE |
| | Outpatients/mixed, 18-64 y<br>3 studies | 1.71 (1.58 to 1.86) | Small-to-moderate increase<br>Moderate; $c^1$ | NE (No CER available) |
| | Outpatients/mixed, ≥65 y<br>3 studies | 1.83 (1.71 to 1.97) | Small-to-moderate increase<br>Moderate; $c^1$ | 0.67 (0.57 to 0.78) |
| Crohn's disease | Outpatients/mixed, 18-64 y<br>1 study | 2.08 (1.47 to 2.95) | Small-to-moderate increase* $^{\mu}$<br>Low; a, $c^5$ | NE (No CER available) |
| Ulcerative colitis | Outpatients/mixed, 18-64 y<br>1 study | 1.02 (0.72 to 1.45) | Very low; a, $c^5$, d | NE |
| **Mental illness** | | | | |
| Mental illness, overall | Outpatients/mixed, <18 y<br>11 studies | 1.01 (0.88 to 1.16) | Little-to-no difference<br>Moderate; b | NE |
| | Outpatients/mixed, 18-64 y<br>12 studies | 1.24 (1.13 to 1.36) | Little-to-no difference<br>Moderate; b | NE |
| | Outpatients/mixed, ≥65 y<br>13 studies | 1.20 (1.07 to 1.35) | Little-to-no difference<br>Low; B | NE |
| | Inpatients, <18 y<br>1 study | 1.28 (1.20 to 1.36) | Small-to-moderate increase<br>Low; $c^1$, d | 10.15 (7.25 to 13.05) |

| Outcome | Subgroup<br>No. Studies | Relative findings<br>HR (95% CI) | Conclusion<br>Certainty for relative findings | Excess cases per 1000 people over 6 mos (95% CI) |
|---|---|---|---|---|
| | Inpatients, 18-64 y<br>3 studies | 1.66 (1.35 to 2.03) | Small-to-moderate increase<br>Moderate; c[6] | 24.69 (13.09 to 38.53) |
| | Inpatients, ≥65 y<br>4 studies | 1.14 (0.76 to 1.71) | Little-to-no difference<br>Moderate; b | NE |
| Anxiety/anxiety disorders | Outpatients/mixed, <18 y<br>6 studies | 0.87 (0.81 to 0.92) | Little-to-no difference<br>High | NE |
| | Outpatients/mixed, 18-64 y<br>6 studies | 1.08 (0.96 to 1.22) | Little-to-no difference<br>Moderate; b | NE |
| | Outpatients/mixed, ≥65 y<br>6 studies | 0.84 (0.66 to 1.08) | Little-to-no difference<br>Low; B | NE |
| | Inpatients, 18-64 y<br>1 study | 2.12 (1.87 to 2.39) | Small to moderate increase* µ<br>Low; b, c[1] | 31.31 (24.32 to 38.86) |
| | Inpatients, ≥65 y<br>1 study | 0.49 (0.36 to 0.66) | Very low; a, b, d | NE |
| Depression & mood disorders | Outpatients/mixed, <18 y<br>6 studies | 1.37 (0.84 to 2.23) | Very low; B, c[1] | NE |
| | Outpatients/mixed, 18-64 y<br>5 studies | 1.11 (1.04 to 1.19) | Little-to-no difference<br>High | NE |
| | Outpatients/mixed, ≥65 y<br>5 studies | 1.07 (1.05 to 1.09) | Little-to-no difference<br>High | NE |
| | Inpatients, 18-64 y<br>1 study | 2.27 (1.96 to 2.63) | Small-to-moderate increase* µ<br>Low; b, c[1] | 24.16 (18.27 to 31.01) |
| | Inpatients, ≥65 y<br>1 study | 1.17 (0.91 to 1.50) | Little-to-no difference<br>Low; a, d | NE |
| Psychosis/psychotic disorders | Outpatients/mixed, <18 y<br>4 studies | 1.23 (0.70 to 2.16) | Little-to-no difference<br>Low; B | NE |
| | <12 mos follow-up<br>2 studies; 31.0% | 0.42 (0.10 to 1.75) | Very low; B, c | NE |
| | ≥12 mos follow-up<br>2 studies; 69.0% | 2.14 (1.46 to 3.14) | Small-to-moderate increase*•<br>Moderate; c[7] | 0.1 (0.04 to 0.18) |
| | Outpatients/mixed, 18-64 y<br>3 studies | 0.89 (0.86 to 0.93) | Little-to-no difference<br>High | NE |
| | Outpatients/mixed, ≥65 y<br>4 studies | 1.60 (1.13 to 2.28) | Small to moderate increase<br>Low; c[1], d | 0.84 (0.18 to 1.8) |
| | Inpatients, 18-64 y<br>1 study | 1.70 (1.51 to 1.92) | Small to moderate increase<br>Moderate; a | 2.22 (1.61 to 2.91) |

| Outcome | Subgroup<br>No. Studies | Relative findings<br>HR (95% CI) | Conclusion<br>Certainty for relative findings | Excess cases per 1000 people over 6 mos (95% CI) |
|---|---|---|---|---|
| | Inpatients, ≥65 y<br>3 studies | 1.72 (1.16 to 2.54) | Small to moderate increase<br>Low; c[1], d | 1.90 (0.42 to 4.05) |
| Trauma & stress disorders | Outpatients/mixed, <18 y<br>2 studies | 1.63 (1.09 to 2.43) | Very low; C[1,8], d | NE |
| | Outpatients/mixed, 18-64 y<br>2 studies | 1.81 (1.11 to 2.95) | Small-to-moderate increase<br>Low; c[1], d | 1.17 (0.16 to 2.82) |
| | Outpatients/mixed, ≥65 y<br>1 study | 1.72 (1.18 to 2.50) | Very low; a, c[1], d | NE |
| | Inpatients, 18-64 y<br>1 study | 2.88 (1.88 to 4.39) | Small to moderate increase*[μ]<br>Moderate; c[1] | 4.38 (2.05 to 7.90) |
| **Musculoskeletal disorders** | | | | |
| Musculoskeletal disorders, overall | Outpatients/mixed, <18 y<br>2 studies | 0.94 (0.92 to 0.97) | Little-to-no difference<br>High | NE |
| Inflammatory arthritides | Outpatients/mixed, <18 y<br>1 study | 1.12 (0.50 to 2.52) | Little-to-no difference<br>Low; D | NE |
| **Neurological disorders** | | | | |
| Neurological disorders, overall | Outpatients/mixed, <18 y<br>10 studies | 1.26 (1.05 to 1.52) | Little-to-no difference<br>Low; B | NE |
| | Outpatients/mixed, 18-64 y<br>10 studies | 1.16 (1.00 to 1.35) | Little-to-no difference<br>Moderate; b | NE |
| | Outpatients/mixed, ≥65 y<br>14 studies | 1.55 (1.18 to 2.04) | Very low; B, c[1] | NE |
| | Inpatients, 18-64 y<br>2 studies | 2.53 (1.14 to 5.63) | Small-to-moderate increase*[±]<br>Moderate; b | 5.42 (0.5 to 16.4) |
| | Inpatients, ≥65 y<br>4 studies | 1.34 (0.89 to 2.04) | Very low; B, c[1] | NE |
| Chronic fatigue syndrome | Outpatients/mixed, <18 y<br>2 studies | 2.46 (1.89 to 3.19) | Small-to-moderate increase*[μ]<br>Low; a, c[1] | 0.63 (0.38 to 0.95) |
| | Outpatients/mixed, 18-64 y<br>1 study | 2.03 (1.34 to 3.06) | Small to moderate increase*[μ]<br>Low; a, c[1] | 1.94 (0.64 to 3.88) |
| | Outpatients/mixed, ≥65 y<br>1 study | 1.12 (1.00 to 1.24) | Little-to-no difference<br>Low; a, b, | NE |
| Communication & motor disorders | Outpatients/mixed, <18 y<br>4 studies | 1.50 (1.12 to 1.99) | Small to moderate increase<br>Low; c[1], d | 0.07 (0.02 to 0.14) |
| | Outpatients/mixed, 18-64 y<br>6 studies | 1.46 (0.72 to 2.96) | Very low; B, c[1] | NE |

| Outcome | Subgroup<br>No. Studies | Relative findings<br>HR (95% CI) | Conclusion<br>Certainty for relative findings | Excess cases per 1000 people over 6 mos (95% CI) |
|---|---|---|---|---|
| | Outpatients/mixed, ≥65 y<br>7 studies | 1.31 (0.99 to 1.74) | Very low; B, c[1] | NE |
| | Inpatients, 18-64 y<br>1 study | 1.53 (0.41 to 5.67) | Very low; C[1,8], D | NE |
| | Inpatients, ≥65 y<br>1 study | 0.67 (0.23 to 1.98) | Very low; b, C[1,9], D | NE |
| Dementia/mild cognitive impairment | Outpatients/mixed, 18-64 y<br>7 studies | 1.42 (1.05 to 1.92) | Very low; B, c[1] | NE |
| | Outpatients/mixed, ≥65 y<br>11 study | 1.42 (1.23 to 1.63) | Small to moderate increase<br>Moderate; b | 4.75 (2.6 to 7.13) |
| | Inpatients, 18-64 y<br>3 studies | 1.64 (0.57 to 4.68) | Very low; B, c[1] | NE |
| | Inpatients, ≥65 y<br>4 studies | 1.42 (0.85 to 2.38) | Very low; B, c[1] | NE |
| Encephalopathy | Outpatients/mixed, 18-64 y<br>2 studies | 2.96 (0.70 to 12.44) | Very low; c[1], D | NE |
| | Outpatients/mixed, ≥65 y<br>2 studies | 2.20 (0.97 to 5.02) | Small-to-moderate increase* [µ]<br>Low; c[1], d | 22.28 (-0.56 to 74.63) |
| | Inpatients, 18-64 y<br>1 study | 24.14 (13.80 to 42.23) | Large increase<br>Moderate; c[1] | 23.92 (13.25 to 1.27) |
| | Inpatients, ≥65 y<br>1 study | 5.84 (5.13 to 6.66) | Large increase<br>Moderate; c[1] | 73.3 (62.54 to 85.71) |
| Epilepsy | Outpatients/mixed, <18 y<br>4 studies | 1.09 (0.61 to 1.95) | Little-to-no difference<br>Low; B | NE |
| | Outpatients/mixed, 18-64 y<br>5 studies | 1.05 (0.94 to 1.17) | Little-to-no difference<br>Moderate; b | NE |
| | Outpatients/mixed, ≥65 y<br>5 studies | 1.26 (1.04 to 1.53) | Very low; B, c[1] | NE |
| | Inpatients, 18-64 y<br>1 study | 5.86 (3.58 to 9.57) | Large increase<br>Low; b, c[1] | 7.55 (4.01 to 13.31) |
| | Inpatients, ≥65 y<br>1 study | 3.86 (2.84 to 5.23) | Large increase<br>Low; b, c[1] | 8.88 (5.72 to 13.14) |
| Guillain-Barre syndrome | Outpatients/mixed, <18 y<br>1 study | 0.64 (0.34 to 1.22) | Very low; b, c[1], d | NE |
| | Outpatients/mixed, 18-64 y<br>4 studies | 1.18 (1.04 to 1.33) | Little-to-no difference<br>Moderate; d | NE |

| Outcome | Subgroup<br>No. Studies | Relative findings<br>HR (95% CI) | Conclusion<br>Certainty for relative findings | Excess cases per 1000 people over 6 mos (95% CI) |
|---|---|---|---|---|
| | Outpatients/mixed, ≥65 y<br>3 studies | 2.32 (2.00 to 2.70) | Large increase<br>Moderate; c[1] | 0.18 (0.14 to 0.24) |
| | Inpatients, 18-64 y<br>2 studies | 3.48 (0.81 to 14.91) | Small-to-moderate increase*• µ<br>Low; c[1], d | 0.29 (-0.02 to 0.45) |
| | Inpatients, ≥65 y<br>2 studies | 2.63 (0.91 to 7.61) | Small-to-moderate increase*• µ<br>Low; c[1], d | 0.23 (-0.01 to 0.94) |
| Migraine | Outpatients/mixed, <18 y<br>1 study | 0.95 (0.43 to 2.11) | Little-to-no difference<br>Low; D | NE |
| | Outpatients/mixed, 18-64 y<br>1 study | 1.29 (1.12 to 1.48) | Small-to-moderate increase<br>Low; c[1], d | 1.53 (0.63 to 2.54) |
| | Outpatients/mixed, ≥65 y<br>1 study | 1.26 (1.03 to 1.55) | Small-to-moderate increase<br>Low; c[1], d | 1.92 (0.22 to 4.06) |
| | Inpatients, 18-64 y<br>1 study | 1.55 (1.21 to 2.00) | Small to moderate increase<br>Low; c[1], d | 4.13 (1.58 to 7.51) |
| | Inpatients, ≥65 y<br>1 study | 0.64 (0.42 to 0.98) | Very low; b, c[1], d | NE |
| Multiple sclerosis | Outpatients/mixed, <18 y<br>1 study | 0.37 (0.10 to 1.37) | Very low; b, D | NE |
| | Outpatients/mixed, 18-64 y<br>2 studies | 0.88 (0.71 to 1.08) | Little-to-no difference<br>Moderate; d | NE |
| | Outpatients/mixed, ≥65 y<br>2 studies | 1.68 (0.80 to 3.51) | Very low; B, c[1] | NE |
| | Inpatients, 18-64 y<br>1 study | 0.30 (0.05 to 1.79) | Very low; c[1], D | NE |
| | Inpatients, ≥65 y<br>1 study | 0.97 (0.16 to 6.03) | Little-to-no difference<br>Low; D | NE |
| Nerve disorders | Outpatients/mixed, <18 y<br>4 studies | 1.10 (0.76 to 1.61) | Little-to-no difference<br>Low; B | NE |
| | Outpatients/mixed, 18-64 y<br>6 studies | 1.08 (0.97 to 1.20) | Little-to-no difference<br>Moderate; b | NE |
| | Outpatients/mixed, ≥65 y<br>6 studies | 1.06 (0.93 to 1.22) | Little-to-no difference<br>Moderate; b | NE |
| | Inpatients, 18-64 y<br>1 study | 3.52 (2.64 to 4.71) | Small to moderate increase*±<br>Low; b, c[10] | 11.42 (7.43 to 16.81) |
| | Inpatients, ≥65 y<br>1 study | 1.44 (1.20 to 1.74) | Very low; C[1,10], d | NE |

| Outcome | Subgroup<br>No. Studies | Relative findings<br>HR (95% CI) | Conclusion<br>Certainty for relative findings | Excess cases per 1000 people over 6 mos (95% CI) |
|---|---|---|---|---|
| **Respiratory disorders** | | | | |
| Respiratory disorders, overall | Outpatients/mixed, <18 y<br>3 studies | 0.99 (0.94 to 1.05) | Little-to-no difference<br>High | NE |
| | Outpatients/mixed, 18-64 y<br>6 studies | 1.89 (1.42 to 2.51) | Very low; B, c[1] | NE |
| | Outpatients/mixed, ≥65 y<br>5 studies | 1.83 (1.31 to 2.56) | Small to moderate increase<br>Moderate; c[1] | 0.9 (0.34 to 1.7) |
| | Inpatients, 18-64 y<br>3 studies | 9.06 (4.93 to 16.67) | Small to moderate increase*[±]<br>High | 59.22 (28.87 to 41.66) |
| | Inpatients, ≥65 y<br>2 studies | 6.85 (4.38 to10.70) | Small to moderate increase*[±]<br>High | 385.26 (222.6 to -19.76) |
| Asthma | Outpatients/mixed, <18 y<br>1 study | 1.00 (0.99 to 1.01) | Little-to-no difference<br>Moderate; b | NE |
| | Outpatients/mixed, 18-64 y<br>2 studies | 2.29 (1.87 to 2.80) | Small-to-moderate increase*[•]<br>Moderate; c[1] | 1.77 (1.2 to 2.48) |
| | Outpatients/mixed, ≥65 y<br>2studies | 1.76 (1.37 to 2.25) | Small to moderate increase<br>Moderate; c[1] | 0.83 (0.4 to 1.37) |
| COPD & bronchiectasis | Outpatients/mixed, 18-64 y<br>3 studies | 1.22 (1.09 to 1.36) | Little-to-no difference<br>Moderate; b | NE |
| | Outpatients/mixed, ≥65 y<br>3 studies | 1.32 (1.04 to 1.67) | Small to moderate increase<br>Low; b, c[1] | 0.35 (0.04 to 0.73) |
| Interstitial lung disease | Outpatients/mixed, 18-64 y<br>2 studies | 3.61 (0.82 to 15.78) | Small-to-moderate increase*[•]<br>Low; c[1], d | 0.93 (-0.06 to 1.7) |
| | Outpatients/mixed, ≥65 y<br>2 studies | 2.45 (1.56 to 3.83) | Small to moderate increase* [µ]<br>Moderate; c[1] | 5.93 (2.29 to 11.58) |
| | Inpatients, 18-64 y<br>1 study | 12.57 (8.22 to 19.20) | Small to moderate increase*[±]<br>High | 19.35 (12.15 to 13.8) |
| | Inpatients, ≥65 y<br>1 study | 7.63 (6.29 to 9.26) | Small to moderate increase*[±]<br>High | 11.16 (8.90 to 13.90) |
| Respiratory failure | Outpatients/mixed, 18-64 y<br>1 study | 12.85 (6.39 to 25.84) | Small to moderate increase*[±]<br>High | 1.74 (0.79 to 3.66) |
| | Outpatients/mixed, ≥65 y<br>2 studies | 2.65 (0.71 to 9.96) | Small to moderate increase*[±]<br>Moderate; b | 4.08 (-0.72 to 22.13) |
| | Inpatients, 18-64 y<br>1 study | 30.78 (15.85 to 59.76) | Small to moderate increase*[±]<br>High | 19.22 (9.58 to 5.67) |

| Outcome | Subgroup<br>No. Studies | Relative findings<br>HR (95% CI) | Conclusion<br>Certainty for relative findings | Excess cases per 1000 people over 6 mos (95% CI) |
|---|---|---|---|---|
| | Inpatients, ≥65 y<br>1 study | 6.44 (5.26 to 7.87) | Small to moderate increase*±<br>High | 32.39 (25.36 to 40.9) |
| **Sleep disorders** | | | | |
| Sleep disorders, overall | Outpatients/mixed, <18 y<br>6 studies | 0.97 (0.91 to 1.04) | Little-to-no difference<br>High | NE |
| | Outpatients/mixed, 18-64 y<br>6 studies | 1.52 (1.23 to 1.87) | Very low; C[1,11], d | NE |
| | Outpatients/mixed, ≥65 y<br>8 studies | 1.15 (0.98 to 1.35) | Very low; a, B | NE |
| | Inpatients, 18-64 y<br>2 studies | 3.69 (3.18 to 4.29) | Small-to-moderate increase*±<br>Moderate; c[11] | 40.74 (33.01 to 49.82) |
| | Inpatients, ≥65 y<br>3 studies | 1.23 (0.77 to 1.98) | Little-to-no difference<br>Low; a, d | NE |
| Insomnia | Outpatients/mixed, <18 y<br>3 studies | 1.03 (0.98 to 1.08) | Little-to-no difference<br>Moderate; c[1] | NE |
| | Outpatients/mixed, 18-64 y<br>2 studies | 0.95 (0.2 to 4.51) | Little-to-no difference<br>Low; D | NE |
| | Outpatients/mixed, ≥65 y<br>3 studies | 0.77 (0.50 to 1.19) | Little-to-no difference<br>Low; a, d | NE |
| | Inpatients, ≥65 y<br>1 study | 0.92 (0.78 to 1.09) | Little-to-no difference<br>Moderate; a | NE |
| Narcolepsy | Outpatients/mixed, 18-64 y<br>1 study | 0.89 (0.20 to 3.89) | Very low; b, D | NE |
| | Outpatients/mixed, ≥65 y<br>1 study | 3.0 (0.80 to 11.27) | Small-to-moderate increase*±µ<br>Moderate; d | 0.09 (-0.01 to 0.01) |
| | Inpatients, 18-64 y<br>1 study | 3.80 (0.54 to 26.68) | Small-to-moderate increase*±<br>Low; D | 0.12 (-0.02 to 0.25) |
| | Inpatients, ≥65 y<br>1 study | 4.94 (0.48 to 51.12) | Small-to-moderate increase*±<br>Low; D | 0.07 (-0.01 to 0) |
| Sleep apnea | Outpatients/mixed, 18-64 y<br>2 studies | 1.69 (1.65 to 1.74) | Small-to-moderate increase<br>High | 4.12 (3.88 to 4.42) |
| | Outpatients/mixed, ≥65 y<br>2 studies | 1.50 (1.48 to 1.53) | Small-to-moderate increase<br>High | 4.01 (3.85 to 4.25) |
| | Inpatients, 18-64 y<br>1 study | 3.69 (3.18 to 4.29) | Small-to-moderate increase*±<br>High | 40.74 (33.01 to 49.82) |

| Outcome | Subgroup<br>No. Studies | Relative findings<br>HR (95% CI) | Conclusion<br>Certainty for relative findings | Excess cases per 1000 people over 6 mos (95% CI) |
|---|---|---|---|---|
| | Inpatients, ≥65 y<br>1 study | 2.15 (1.85 to 2.49) | Small-to-moderate increase*[µ]<br>Moderate; c[1] | 16.52 (12.21 to 21.41) |
| **Stroke** | | | | |
| Stroke, overall | Outpatients/mixed, <18 y<br>5 studies | 1.14 (0.99 to 1.30) | Little-to-no difference<br>Moderate; d | NE |
| | Outpatients/mixed, 18-64 y<br>14 studies | 1.11 (0.95 to 1.31) | Little-to-no difference<br>Low; B | NE |
| | Outpatients/mixed, ≥65 y<br>12 studies | 1.17 (0.99 to 1.39) | Little-to-no difference<br>Low; B | NE |
| | Inpatients, 18-64 y<br>4 studies | 1.12 (0.42 to 3.03) | Very low; B, c[12] | NE |
| | Inpatients, ≥65 y<br>4 studies | 0.75 (0.30 to 1.92) | Very low; B, c[12] | NE |
| Haemorrhagic stroke | Outpatients/mixed, 18-64 y<br>1 study | 2.59 (1.41 to 4.75) | Small-to-moderate increase*[µ]<br>Moderate; c[1] | 0.38 (0.1 to 0.9) |
| | Outpatients/mixed, ≥65 y<br>2 studies | 1.41 (0.68 to 2.94) | Very low; B, c[1] | NE |
| | Inpatients, 18-64 y<br>1 study | 4.86 (2.61 to 9.03) | Small-to-moderate increase*[±]<br>High | 4.00 (1.67 to 8.31) |
| | Inpatients, ≥65 y<br>1 study | 2.79 (2.39 to 3.26) | Small-to-moderate increase*[±]<br>High | 22.01 (17.09 to 27.79) |
| Ischemic stroke | Outpatients/mixed, <18 y<br>1 study | 1.10 (0.89 to 1.34) | Little-to-no difference<br>Moderate; d | NE |
| | Outpatients/mixed, 18-64 y<br>3 studies | 0.78 (0.53 to 1.17) | Little-to-no difference<br>Low; B | NE |
| | Outpatients/mixed, ≥65 y<br>4 studies | 0.98 (0.76 to 1.28) | Little-to-no difference<br>Low; B | NE |
| | Inpatients, 18-64 y<br>3 studies | 0.94 (0.24 to 3.76) | Little-to-no difference<br>Low; B | NE |
| | Inpatients, ≥65 y<br>3 studies | 0.80 (0.27 to 2.36) | Little-to-no difference<br>Low; B | NE |
| Transient ischemic attack | Outpatients/mixed, 18-64 y<br>2 studies | 1.33 (1.03 to 1.73) | Small to moderate increase<br>Low; b, c[1] | 0.04 (0 to 0.1) |
| | Outpatients/mixed, ≥65 y<br>3 studies | 1.44 (1.11 to 1.87) | Small-to-moderate increase<br>Low; b, c[1] | 1.1 (0.27 to 2.17) |

**Note:** CER = control event rate; CI = confidence interval; COPD = chronic obstructive pulmonary disease; HR = hazard ratio; mos= months; NE = not estimated; y = years

**GRADE legend:** A = ROB, B = inconsistency; C = Indirectness; D = imprecision. Lowercase and capital letters represent downrating that domain for one or two steps, respectively. *Rated as small-to-moderate association (instead of large association) for greater certainty; [µ]Large effect would be rated down for imprecision; [•]Large effect would be rated down for inconsistency; [±]Large effect would be rated down for indirectness.

#### Footnotes:

[1] Concerns about applicability of conclusions to a contemporary population (highly vaccinated and exposed to Omicron variant)
[2] ≥60% of weight from single condition (atopic dermatitis)
[3] 100% of weight from single condition (myositis)
[4] 95% of weight from single condition type (arrhythmias)
[5] Based on data from 18-40y only
[6] >50% of weight from single condition (schizophrenia & related disorders)
[7] Concerns about outcome definition in 100% of weight (inclusion of ICD-10 F23, Acute and transient psychotic disorders)
[8] 100% weight from single condition (adjustment disorder)
[9] 100% of weight from only 2 conditions (Parkinson’s and myasthenia gravis)
[10] 100% of weight from single condition (peripheral neuropathy)
[11] >65% of weight from single condition (sleep apnea)
[12] >50% of weight from ischemic stroke

### Appendix C. Forest Plots for primary analyses (new diagnoses)

#### Autoimmune disorders

##### Overall

| Study or Subgroup | log[Hazard Ratio] | SE | Weight | Hazard Ratio IV, Random, 95% CI |
|---|---|---|---|---|
| **1.1.1 Autoimmune disorders, overall (outpatients/mixed, <18 y)** | | | | |
| Mizrahi 2023, 0-4 y | -0.46960737 | 0.30738814 | 11.1% | 0.63 [0.34, 1.14] |
| Mizrahi 2023, 12-18 y | -0.13926 | 0.213002 | 15.2% | 0.87 [0.57, 1.32] |
| Mizrahi 2023, 5-11 y | -0.3597 | 0.232888 | 14.3% | 0.70 [0.44, 1.10] |
| Rao 2022 | 0.951658 | 0.360659 | 9.3% | 2.59 [1.28, 5.25] |
| Schmitt 2024 | 0.307485 | 0.061521 | 22.0% | 1.36 [1.21, 1.53] |
| Tartof 2022 | -0.04082 | 0.401122 | 8.2% | 0.96 [0.44, 2.11] |
| Tesch 2023 | 0.48858 | 0.118669 | 19.8% | 1.63 [1.29, 2.06] |
| **Subtotal (95% CI)** | | | **100.0%** | **1.13 [0.86, 1.50]** |
| Heterogeneity: Tau² = 0.09; Chi² = 24.73, df = 6 (P = 0.0004); I² = 76% | | | | |
| Test for overall effect: Z = 0.88 (P = 0.38) | | | | |
| **1.1.2 Autoimmune disorders, overall (outpatients/mixed, 18-64 y)** | | | | |
| Chang 2023, 18-40 y | 0.656266 | 0.123921 | 4.7% | 1.93 [1.51, 2.46] |
| Chang 2023, 41-64 y | 0.946803 | 0.098617 | 5.0% | 2.58 [2.12, 3.13] |
| Daugherty 2021 | 0.215111 | 0.10761 | 4.9% | 1.24 [1.00, 1.53] |
| Huang 2024 | 0.678034 | 0.013018 | 5.4% | 1.97 [1.92, 2.02] |
| Kim 2024, 20-39 y | 0.058269 | 0.040807 | 5.3% | 1.06 [0.98, 1.15] |
| Kim 2024, 40-59 y | 0.139762 | 0.019888 | 5.4% | 1.15 [1.11, 1.20] |
| Kyung 2024, Japan cohort, 20-39 y | 0.300105 | 0.078828 | 5.1% | 1.35 [1.16, 1.58] |
| Kyung 2024, Japan cohort, 40-59 y | 0.329304 | 0.060195 | 5.2% | 1.39 [1.24, 1.56] |
| Kyung 2024, South Korea cohort, 20-39 y | 0.405465 | 0.16005 | 4.3% | 1.50 [1.10, 2.05] |
| Kyung 2024, South Korea cohort, 40-59y | 0.756122 | 0.126909 | 4.7% | 2.13 [1.66, 2.73] |
| Lim 2023 | 0.04697 | 0.219831 | 3.7% | 1.05 [0.68, 1.61] |
| Mizrahi 2023, 19-40 y | -0.26379 | 0.240527 | 3.5% | 0.77 [0.48, 1.23] |
| Mizrahi 2023, 41-60 y | 0.128753 | 0.302499 | 2.9% | 1.14 [0.63, 2.06] |
| Mizrahi 2023, 60+ y | -0.82098055 | 1.764631 | 0.2% | 0.44 [0.01, 13.98] |
| Oh 2024, Japan cohort, 20-39 y | 0.95958 | 0.032756 | 5.4% | 2.61 [2.45, 2.78] |
| Oh 2024, Japan cohort, 40-59 y | 0.945678 | 0.025499 | 5.4% | 2.57 [2.45, 2.71] |
| Oh 2024, Japan cohort, 60+ y | 0.747603 | 0.118029 | 4.8% | 2.11 [1.68, 2.66] |
| Oh 2024, South Korea cohort, 20-39 y | 0.211086 | 0.129947 | 4.7% | 1.24 [0.96, 1.59] |
| Oh 2024, South Korea cohort, 40-59 y | 0.18091 | 0.094441 | 5.0% | 1.20 [1.00, 1.44] |
| Oh 2024, South Korea cohort, 60+ y | 0.183624 | 0.217209 | 3.7% | 1.20 [0.78, 1.84] |
| Schmitt 2024 | 0.262364 | 0.031437 | 5.4% | 1.30 [1.22, 1.38] |
| Tesch 2023 | 0.364643 | 0.023126 | 5.4% | 1.44 [1.38, 1.51] |
| **Subtotal (95% CI)** | | | **100.0%** | **1.51 [1.31, 1.75]** |
| Heterogeneity: Tau² = 0.10; Chi² = 1303.50, df = 21 (P < 0.00001); I² = 98% | | | | |
| Test for overall effect: Z = 5.50 (P < 0.00001) | | | | |
| **1.1.3 Autoimmune disorders, overall (outpatients/mixed, ≥ 65 y)** | | | | |
| Chang 2023 | 1.070937 | 0.158296 | 7.6% | 2.92 [2.14, 3.98] |
| Cohen 2022 | -0.03046 | 0.180547 | 7.0% | 0.97 [0.68, 1.38] |
| Huang 2024 | 0.779325 | 0.191731 | 6.7% | 2.18 [1.50, 3.17] |
| Kim 2024 | 0.14842 | 0.044135 | 10.2% | 1.16 [1.06, 1.26] |
| Kyung 2024, Japan cohort | 0.322083 | 0.17357 | 7.2% | 1.38 [0.98, 1.94] |
| Kyung 2024, South Korea cohort | -0.56212 | 0.390963 | 3.1% | 0.57 [0.26, 1.23] |
| Mizrahi 2023, 60+ y | -0.82098 | 0.770148 | 1.0% | 0.44 [0.10, 1.99] |
| Oh 2024, Japan cohort | 0.747603 | 0.057592 | 10.0% | 2.11 [1.89, 2.36] |
| Oh 2024, South Korea cohort | 0.183624 | 0.105987 | 9.0% | 1.20 [0.98, 1.48] |
| Schmitt 2024, 65-79 y | 0.350657 | 0.093141 | 9.3% | 1.42 [1.18, 1.70] |
| Schmitt 2024, 80+ y | 0.371564 | 0.104872 | 9.0% | 1.45 [1.18, 1.78] |
| Tesch 2023, 65-79 y | 0.350657 | 0.052096 | 10.1% | 1.42 [1.28, 1.57] |
| Tesch 2023, 80+ y | 0.300105 | 0.067926 | 9.8% | 1.35 [1.18, 1.54] |
| **Subtotal (95% CI)** | | | **100.0%** | **1.44 [1.23, 1.69]** |
| Heterogeneity: Tau² = 0.06; Chi² = 109.46, df = 12 (P < 0.00001); I² = 89% | | | | |
| Test for overall effect: Z = 4.45 (P < 0.00001) | | | | |
| **1.1.4 Autoimmune disorders, overall (inpatients, 18-64 y)** | | | | |
| Daugherty 2021 | 0.041718 | 0.204182 | 100.0% | 1.04 [0.70, 1.56] |
| **Subtotal (95% CI)** | | | **100.0%** | **1.04 [0.70, 1.56]** |
| Heterogeneity: Not applicable | | | | |
| Test for overall effect: Z = 0.20 (P = 0.84) | | | | |
| **1.1.5 Autoimmune disorders, overall (inpatients, ≥ 65 y)** | | | | |
| Cohen 2022 | 0.128657 | 0.191952 | 100.0% | 1.14 [0.78, 1.66] |
| **Subtotal (95% CI)** | | | **100.0%** | **1.14 [0.78, 1.66]** |
| Heterogeneity: Not applicable | | | | |
| Test for overall effect: Z = 0.67 (P = 0.50) | | | | |

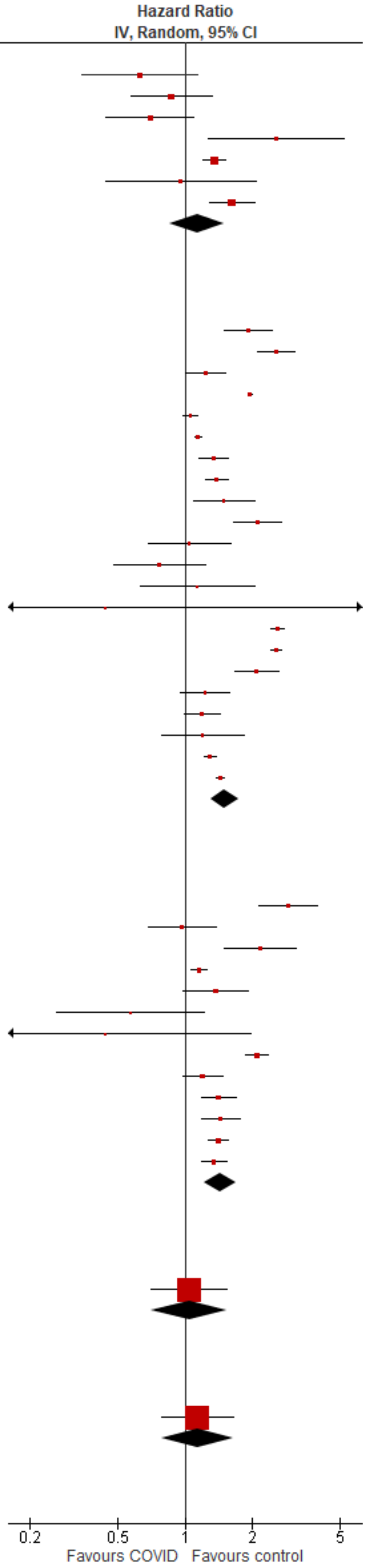

Test for subgroup differences: Chi² = 6.57, df = 4 (P = 0.16), I² = 39.1%

#### Allergic diseases

| Study or Subgroup | log[Hazard Ratio] | SE | Weight | Hazard Ratio IV, Random, 95% CI |
|---|---|---|---|---|
| **1.2.1 Allergic diseases (outpatients/mixed, <18 y)** | | | | |
| Schmitt 2024 | 0.307485 | 0.061521 | 100.0% | 1.36 [1.21, 1.53] |
| **Subtotal (95% CI)** | | | **100.0%** | **1.36 [1.21, 1.53]** |
| Heterogeneity: Not applicable | | | | |
| Test for overall effect: Z = 5.00 (P < 0.00001) | | | | |
| **1.2.2 Allergic diseases (outpatients/mixed, 18-64 y)** | | | | |
| Daugherty 2021 | 0.215111 | 0.10761 | 12.5% | 1.24 [1.00, 1.53] |
| Oh 2024, Japan cohort, 20-39 y | 0.95958 | 0.032756 | 13.4% | 2.61 [2.45, 2.78] |
| Oh 2024, Japan cohort, 40-59 y | 0.945678 | 0.025499 | 13.4% | 2.57 [2.45, 2.71] |
| Oh 2024, Japan cohort, 60+ y | 0.747603 | 0.118029 | 12.3% | 2.11 [1.68, 2.66] |
| Oh 2024, South Korea cohort, 20-39 y | 0.211086 | 0.129947 | 12.1% | 1.24 [0.96, 1.59] |
| Oh 2024, South Korea cohort, 40-59 y | 0.18091 | 0.094441 | 12.7% | 1.20 [1.00, 1.44] |
| Oh 2024, South Korea cohort, 60+ y | 0.183624 | 0.217209 | 10.3% | 1.20 [0.78, 1.84] |
| Schmitt 2024 | 0.262364 | 0.031437 | 13.4% | 1.30 [1.22, 1.38] |
| **Subtotal (95% CI)** | | | **100.0%** | **1.61 [1.22, 2.13]** |
| Heterogeneity: $Tau^2 = 0.15$; $Chi^2 = 419.48$, df = 7 (P < 0.00001); $I^2 = 98\%$ | | | | |
| Test for overall effect: Z = 3.33 (P = 0.0009) | | | | |
| **1.2.3 Allergic diseases (outpatients/mixed, ≥ 65 y)** | | | | |
| Cohen 2022 | -0.03046 | 0.180547 | 16.5% | 0.97 [0.68, 1.38] |
| Oh 2024, Japan cohort | 0.747603 | 0.057592 | 22.1% | 2.11 [1.89, 2.36] |
| Oh 2024, South Korea cohort | 0.183624 | 0.105987 | 20.3% | 1.20 [0.98, 1.48] |
| Schmitt 2024, 65-79 y | 0.350657 | 0.093141 | 20.8% | 1.42 [1.18, 1.70] |
| Schmitt 2024, 80+ y | 0.371564 | 0.104872 | 20.3% | 1.45 [1.18, 1.78] |
| **Subtotal (95% CI)** | | | **100.0%** | **1.41 [1.08, 1.85]** |
| Heterogeneity: $Tau^2 = 0.08$; $Chi^2 = 40.09$, df = 4 (P < 0.00001); $I^2 = 90\%$ | | | | |
| Test for overall effect: Z = 2.50 (P = 0.01) | | | | |
| **1.2.4 Allergic diseases (inpatients, 18-64 y)** | | | | |
| Daugherty 2021 | 0.041718 | 0.204182 | 100.0% | 1.04 [0.70, 1.56] |
| **Subtotal (95% CI)** | | | **100.0%** | **1.04 [0.70, 1.56]** |
| Heterogeneity: Not applicable | | | | |
| Test for overall effect: Z = 0.20 (P = 0.84) | | | | |
| **1.2.5 Allergic diseases (inpatients, ≥ 65 y)** | | | | |
| Cohen 2022 | 0.128657 | 0.191952 | 100.0% | 1.14 [0.78, 1.66] |
| **Subtotal (95% CI)** | | | **100.0%** | **1.14 [0.78, 1.66]** |
| Heterogeneity: Not applicable | | | | |
| Test for overall effect: Z = 0.67 (P = 0.50) | | | | |

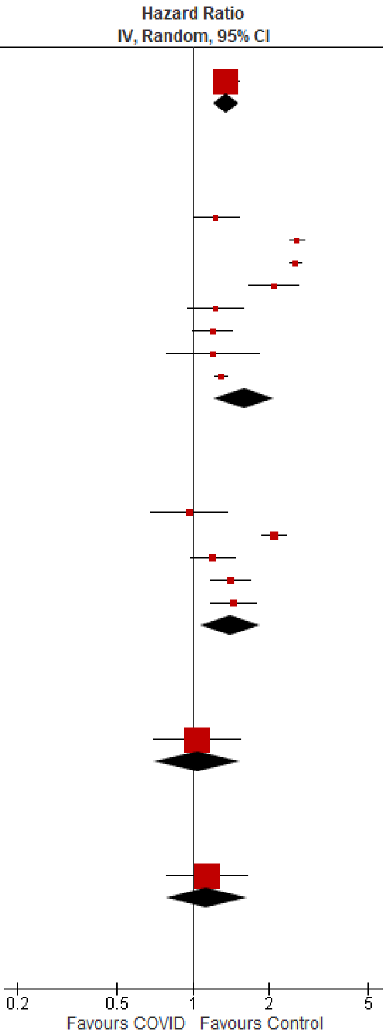

Test for subgroup differences: $Chi^2 = 4.03$, df = 4 (P = 0.40), $I^2 = 0.7\%$

#### Alopecia

| Study or Subgroup | log[Hazard Ratio] | SE | Weight | Hazard Ratio IV, Random, 95% CI |
|---|---|---|---|---|
| **1.3.1 Alopecia (outpatients/mixed, <18 y)** | | | | |
| Tartof 2022 | -0.04082 | 0.401122 | 100.0% | 0.96 [0.44, 2.11] |
| **Subtotal (95% CI)** | | | **100.0%** | **0.96 [0.44, 2.11]** |
| Heterogeneity: Not applicable | | | | |
| Test for overall effect: Z = 0.10 (P = 0.92) | | | | |
| **1.3.2 Alopecia (outpatients/mixed, 18-64 y)** | | | | |
| Kyung 2024, Japan cohort, 20-39 y | 0.300105 | 0.078828 | 23.5% | 1.35 [1.16, 1.58] |
| Kyung 2024, Japan cohort, 40-59 y | 0.329304 | 0.060195 | 25.1% | 1.39 [1.24, 1.56] |
| Kyung 2024, South Korea cohort, 20-39 y | 0.405465 | 0.16005 | 16.0% | 1.50 [1.10, 2.05] |
| Kyung 2024, South Korea cohort, 40-59y | 0.756122 | 0.126909 | 18.9% | 2.13 [1.66, 2.73] |
| Lim 2023 | -0.05104 | 0.153892 | 16.5% | 0.95 [0.70, 1.28] |
| **Subtotal (95% CI)** | | | **100.0%** | **1.42 [1.17, 1.73]** |
| Heterogeneity: $Tau^2 = 0.03$; $Chi^2 = 17.65$, df = 4 (P = 0.001); $I^2 = 77\%$ | | | | |
| Test for overall effect: Z = 3.59 (P = 0.0003) | | | | |
| **1.3.3 Alopecia (outpatients/mixed, ≥ 65 y)** | | | | |
| Kyung 2024, Japan cohort | 0.322083 | 0.17357 | 57.8% | 1.38 [0.98, 1.94] |
| Kyung 2024, South Korea cohort | -0.56212 | 0.390963 | 42.2% | 0.57 [0.26, 1.23] |
| **Subtotal (95% CI)** | | | **100.0%** | **0.95 [0.40, 2.24]** |
| Heterogeneity: $Tau^2 = 0.30$; $Chi^2 = 4.27$, df = 1 (P = 0.04); $I^2 = 77\%$ | | | | |
| Test for overall effect: Z = 0.12 (P = 0.91) | | | | |

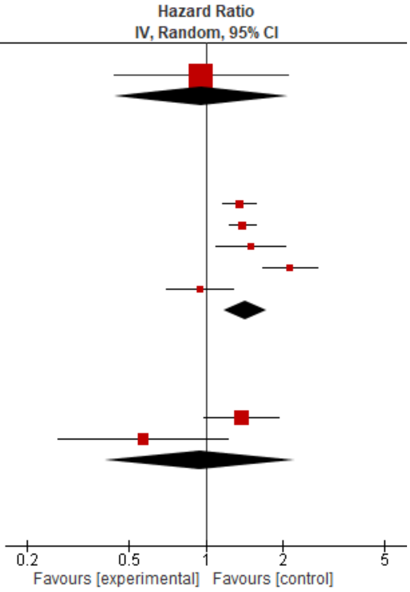

Test for subgroup differences: $Chi^2 = 1.64$, df = 2 (P = 0.44), $I^2 = 0\%$

#### Celiac disease

| Study or Subgroup | log[Hazard Ratio] | SE | Weight | Hazard Ratio IV, Random, 95% CI |
|---|---|---|---|---|
| **1.4.1 Celiac disease (outpatients/mixed, <18 y)** | | | | |
| Mizrahi 2023, 0-4 y | -0.46960737 | 0.30738814 | 20.7% | 0.63 [0.34, 1.14] |
| Mizrahi 2023, 12-18 y | -0.13926 | 0.213002 | 43.2% | 0.87 [0.57, 1.32] |
| Mizrahi 2023, 5-11 y | -0.3597 | 0.232888 | 36.1% | 0.70 [0.44, 1.10] |
| **Subtotal (95% CI)** | | | **100.0%** | **0.75 [0.57, 0.99]** |
| Heterogeneity: Tau² = 0.00; Chi² = 0.93, df = 2 (P = 0.63); I² = 0% | | | | |
| Test for overall effect: Z = 2.05 (P = 0.04) | | | | |
| **1.4.2 Celiac disease (outpatients/mixed, 18-64 y)** | | | | |
| Chang 2023, 18-40 y | 0.431782 | 0.111534 | 31.3% | 1.54 [1.24, 1.92] |
| Chang 2023, 41-64 y | 0.636577 | 0.122666 | 30.4% | 1.89 [1.49, 2.40] |
| Mizrahi 2023, 19-40 y | -0.26379 | 0.240527 | 20.8% | 0.77 [0.48, 1.23] |
| Mizrahi 2023, 41-60 y | 0.128753 | 0.302499 | 16.7% | 1.14 [0.63, 2.06] |
| Mizrahi 2023, 60+ y | -0.82098055 | 1.764631 | 0.9% | 0.44 [0.01, 13.98] |
| **Subtotal (95% CI)** | | | **100.0%** | **1.33 [0.96, 1.85]** |
| Heterogeneity: Tau² = 0.08; Chi² = 12.63, df = 4 (P = 0.01); I² = 68% | | | | |
| Test for overall effect: Z = 1.71 (P = 0.09) | | | | |
| **1.4.3 Celiac disease (outpatients/mixed, ≥ 65 y)** | | | | |
| Chang 2023 | 0.765468 | 0.215371 | 100.0% | 2.15 [1.41, 3.28] |
| **Subtotal (95% CI)** | | | **100.0%** | **2.15 [1.41, 3.28]** |
| Heterogeneity: Not applicable | | | | |
| Test for overall effect: Z = 3.55 (P = 0.0004) | | | | |

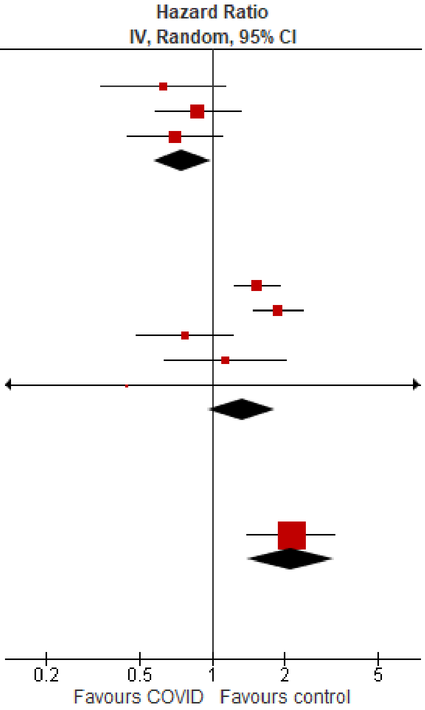

Test for subgroup differences: Chi² = 18.44, df = 2 (P < 0.0001), I² = 89.2%

#### Connective tissue diseases

| Study or Subgroup | log[Hazard Ratio] | SE | Weight | Hazard Ratio IV, Random, 95% CI |
|---|---|---|---|---|
| **1.5.1 Connective tissue diseases (outpatients/mixed, <18 y)** | | | | |
| Rao 2022 | 0.951658 | 0.360659 | 100.0% | 2.59 [1.28, 5.25] |
| **Subtotal (95% CI)** | | | **100.0%** | **2.59 [1.28, 5.25]** |
| Heterogeneity: Not applicable | | | | |
| Test for overall effect: Z = 2.64 (P = 0.008) | | | | |
| **1.5.2 Connective tissue diseases (outpatients/mixed, 18-64 y)** | | | | |
| Chang 2023, 18-40 y | 0.839476 | 0.198527 | 33.2% | 2.32 [1.57, 3.42] |
| Chang 2023, 41-64 y | 1.092313 | 0.132331 | 36.1% | 2.98 [2.30, 3.86] |
| Lim 2023 | -0.00868 | 0.247873 | 30.7% | 0.99 [0.61, 1.61] |
| **Subtotal (95% CI)** | | | **100.0%** | **1.95 [1.08, 3.54]** |
| Heterogeneity: Tau² = 0.24; Chi² = 15.35, df = 2 (P = 0.0005); I² = 87% | | | | |
| Test for overall effect: Z = 2.21 (P = 0.03) | | | | |
| **1.5.3 Connective tissue diseases (outpatients/mixed, ≥ 65 y)** | | | | |
| Chang 2023 | 1.055285 | 0.225984 | 100.0% | 2.87 [1.84, 4.47] |
| **Subtotal (95% CI)** | | | **100.0%** | **2.87 [1.84, 4.47]** |
| Heterogeneity: Not applicable | | | | |
| Test for overall effect: Z = 4.67 (P < 0.00001) | | | | |

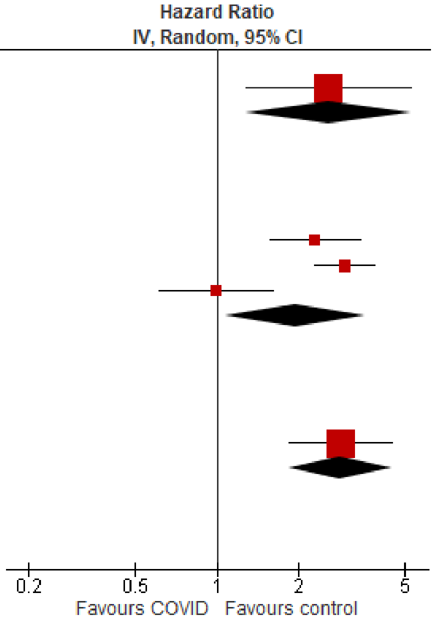

Test for subgroup differences: Chi² = 1.05, df = 2 (P = 0.59), I² = 0%

#### Hyperthyroid

| Study or Subgroup | log[Hazard Ratio] | SE | Weight | Hazard Ratio IV, Random, 95% CI |
|---|---|---|---|---|
| **1.6.1 Hyperthyroid (outpatients/mixed, 18-64 y)** | | | | |
| Huang 2024 | 0.553885 | 0.033867 | 100.0% | 1.74 [1.63, 1.86] |
| **Subtotal (95% CI)** | | | **100.0%** | **1.74 [1.63, 1.86]** |
| Heterogeneity: Not applicable | | | | |
| Test for overall effect: Z = 16.35 (P < 0.00001) | | | | |
| **1.6.2 Hyperthyroid (outpatients/mixed, ≥ 65 y)** | | | | |
| Huang 2024 | 0.678034 | 0.060687 | 100.0% | 1.97 [1.75, 2.22] |
| **Subtotal (95% CI)** | | | **100.0%** | **1.97 [1.75, 2.22]** |
| Heterogeneity: Not applicable | | | | |
| Test for overall effect: Z = 11.17 (P < 0.00001) | | | | |

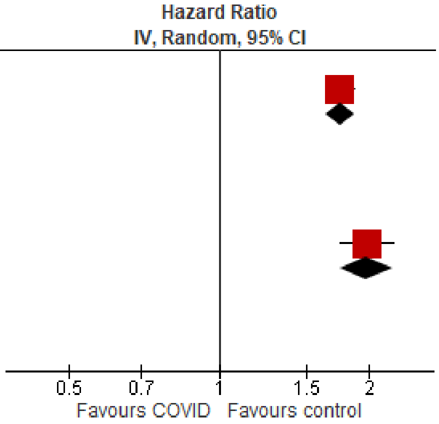

Test for subgroup differences: Chi² = 3.19, df = 1 (P = 0.07), I² = 68.7%

#### Hypothyroid

| Study or Subgroup | log[Hazard Ratio] | SE | Weight | Hazard Ratio IV, Random, 95% CI |
|---|---|---|---|---|
| **1.7.1 Hypothyroid (outpatients/mixed, 18-64 y)** | | | | |
| Huang 2024 | 0.698135 | 0.012694 | 100.0% | 2.01 [1.96, 2.06] |
| **Subtotal (95% CI)** | | | **100.0%** | **2.01 [1.96, 2.06]** |
| Heterogeneity: Not applicable | | | | |
| Test for overall effect: Z = 55.00 (P < 0.00001) | | | | |
| **1.7.2 Hypothyroid (outpatients/mixed, ≥ 65 y)** | | | | |
| Huang 2024 | 0.792993 | 0.017361 | 100.0% | 2.21 [2.14, 2.29] |
| **Subtotal (95% CI)** | | | **100.0%** | **2.21 [2.14, 2.29]** |
| Heterogeneity: Not applicable | | | | |
| Test for overall effect: Z = 45.68 (P < 0.00001) | | | | |

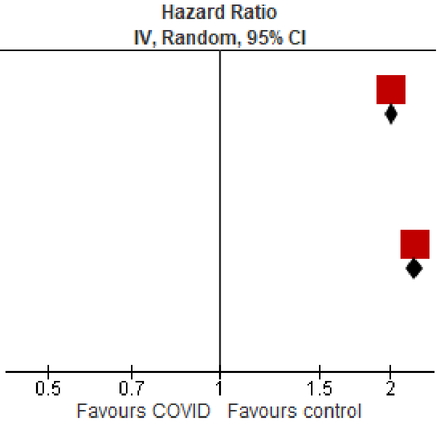

Test for subgroup differences: Chi² = 19.45, df = 1 (P < 0.0001), I² = 94.9%

#### Inflammatory rheumatic diseases

| Study or Subgroup | log[Hazard Ratio] | SE | Weight | Hazard Ratio IV, Random, 95% CI |
|---|---|---|---|---|
| **1.8.1 Inflammatory rheumatic diseases (outpatients/mixed, 18-64 y)** | | | | |
| Chang 2023, 18-40 y | 0.926525 | 0.142032 | 18.6% | 2.53 [1.91, 3.34] |
| Chang 2023, 41-64 y | 1.087735 | 0.107007 | 20.3% | 2.97 [2.41, 3.66] |
| Kim 2024, 20-39 y | 0.058269 | 0.040807 | 22.5% | 1.06 [0.98, 1.15] |
| Kim 2024, 40-59 y | 0.139762 | 0.019888 | 22.9% | 1.15 [1.11, 1.20] |
| Lim 2023 | 0.25546 | 0.200947 | 15.7% | 1.29 [0.87, 1.91] |
| **Subtotal (95% CI)** | | | **100.0%** | **1.61 [1.22, 2.13]** |
| Heterogeneity: Tau² = 0.09; Chi² = 111.74, df = 4 (P < 0.00001); I² = 96% | | | | |
| Test for overall effect: Z = 3.38 (P = 0.0007) | | | | |
| **1.8.2 Inflammatory rheumatic diseases (outpatients/mixed, ≥ 65 y)** | | | | |
| Chang 2023 | 1.200721 | 0.137226 | 49.2% | 3.32 [2.54, 4.35] |
| Kim 2024 | 0.14842 | 0.044135 | 50.8% | 1.16 [1.06, 1.26] |
| **Subtotal (95% CI)** | | | **100.0%** | **1.95 [0.69, 5.46]** |
| Heterogeneity: Tau² = 0.54; Chi² = 53.29, df = 1 (P < 0.00001); I² = 98% | | | | |
| Test for overall effect: Z = 1.27 (P = 0.21) | | | | |

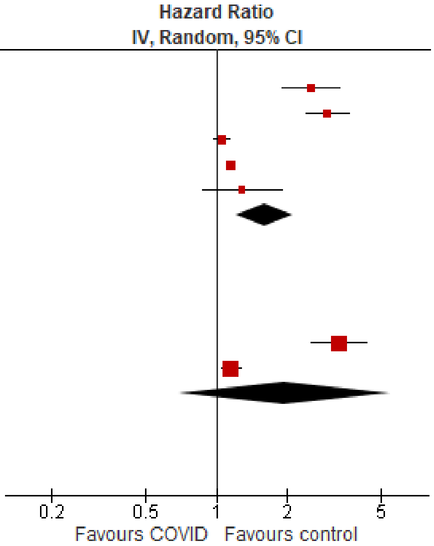

Test for subgroup differences: Chi² = 0.12, df = 1 (P = 0.73), I² = 0%

### Cardiovascular disorders

#### Overall

| Study or Subgroup | log[Hazard Ratio] | SE | Weight | Hazard Ratio IV, Random, 95% CI |
|---|---|---|---|---|
| **2.1.1 Cardiovascular disorders, overall (outpatients/mixed, <18 y)** | | | | |
| Gordon 2024, 0-1 y | 0.071479 | 0.21927 | 0.6% | 1.07 [0.70, 1.65] |
| Gordon 2024, 12-17 y | 0.237643 | 2.813738 | 0.0% | 1.27 [0.01, 314.97] |
| Gordon 2024, 2-4 y | 0.36921 | 2.813829 | 0.0% | 1.45 [0.01, 359.32] |
| Gordon 2024, 5-11 y | 0.321619 | 0.173146 | 1.0% | 1.38 [0.98, 1.94] |
| Kompaniyets 2022 | 0.14842 | 0.0176 | 95.2% | 1.16 [1.12, 1.20] |
| Mizrahi 2023, 12-18 y | -0.05374 | 0.386854 | 0.2% | 0.95 [0.44, 2.02] |
| Roessler 2022 | 0.355874 | 0.624046 | 0.1% | 1.43 [0.42, 4.85] |
| Tartof 2022 | 0.576613 | 0.624046 | 0.1% | 1.78 [0.52, 6.05] |
| Valdivieso-Martinez 2023, unvaccinated cohort | 1.098612 | 1.154739 | 0.0% | 3.00 [0.31, 28.84] |
| Valdivieso-Martinez 2023, vaccinated cohort | 1.94591 | 3.75576 | 0.0% | 7.00 [0.00, 11015.56] |
| Zhang 2024 | 0.398109 | 0.102964 | 2.8% | 1.49 [1.22, 1.82] |
| **Subtotal (95% CI)** | | | **100.0%** | **1.17 [1.13, 1.21]** |
| Heterogeneity: Tau² = 0.00; Chi² = 8.53, df = 10 (P = 0.58); I² = 0% | | | | |
| Test for overall effect: Z = 9.14 (P < 0.00001) | | | | |
| **2.1.2 Cardiovascular disorders, overall (outpatients/mixed, 18-64 y)** | | | | |
| Batistoni 2024 | 0.836308 | 0.216484 | 4.5% | 2.31 [1.51, 3.53] |
| Daugherty 2021 | 0.715483 | 0.094995 | 5.9% | 2.05 [1.70, 2.46] |
| Katsoularis 2023, 60+ y | 0.227813 | 0.25172 | 4.0% | 1.26 [0.77, 2.06] |
| Katsoularis 2023, 60 y & under | 0.348127 | 0.183357 | 4.9% | 1.42 [0.99, 2.03] |
| Lam 2023 | 0.385263 | 0.163468 | 5.1% | 1.47 [1.07, 2.03] |
| Lim 2023 | 0.690556 | 0.220029 | 4.4% | 1.99 [1.30, 3.07] |
| Mizrahi 2023, 19-40 y | 0.074487 | 0.201878 | 4.6% | 1.08 [0.73, 1.60] |
| Mizrahi 2023, 41-60 y | 0.044255 | 0.158501 | 5.2% | 1.05 [0.77, 1.43] |
| Mizrahi 2023, 60+ y | -0.13266 | 0.359911 | 2.9% | 0.88 [0.43, 1.77] |
| Rezel-Potts 2022 | -0.05033 | 0.054413 | 6.2% | 0.95 [0.85, 1.06] |
| Velasquez Garcia 2024 | 0.329304 | 0.1379 | 5.4% | 1.39 [1.06, 1.82] |
| Wang 2022b, 20-44y | 0.527832 | 0.083008 | 6.0% | 1.70 [1.44, 1.99] |
| Wang 2022b, 45-64y | 0.721112 | 0.045725 | 6.3% | 2.06 [1.88, 2.25] |
| Xie 2022b | 0.51721819 | 0.043625 | 6.3% | 1.68 [1.54, 1.83] |
| Zang 2023, INSIGHT cohort | 0.110557 | 0.063558 | 6.2% | 1.12 [0.99, 1.27] |
| Zappacosta 2022, 18-30 y | 0.350657 | 0.37779 | 2.8% | 1.42 [0.68, 2.98] |
| Zappacosta 2022, 31-40 y | 0.29267 | 0.254071 | 4.0% | 1.34 [0.81, 2.20] |
| Zappacosta 2022, 41-50 y | -0.57982 | 0.168459 | 5.1% | 0.56 [0.40, 0.78] |
| Zappacosta 2022, 51-60 y | -0.08338 | 0.131688 | 5.5% | 0.92 [0.71, 1.19] |
| Zappacosta 2022, 61-70 y | -0.0202 | 0.186986 | 4.8% | 0.98 [0.68, 1.41] |
| **Subtotal (95% CI)** | | | **100.0%** | **1.32 [1.12, 1.55]** |
| Heterogeneity: Tau² = 0.11; Chi² = 227.01, df = 19 (P < 0.00001); I² = 92% | | | | |
| Test for overall effect: Z = 3.30 (P = 0.0010) | | | | |
| **2.1.3 Cardiovascular disorders, overall (outpatients/mixed, ≥ 65 y)** | | | | |
| Batistoni 2024 | 0.43628 | 0.1502 | 7.8% | 1.55 [1.15, 2.08] |
| Cohen 2022 | 0.61252 | 0.0677 | 10.1% | 1.85 [1.62, 2.11] |
| Katsoularis 2023, 60+ y | 0.22781 | 0.25172 | 5.1% | 1.26 [0.77, 2.06] |
| Mizrahi 2023, 60+ y | -0.1327 | 0.15708 | 7.6% | 0.88 [0.64, 1.19] |
| Wan 2023a, UK cohort | 0.34905 | 0.09979 | 9.3% | 1.42 [1.17, 1.72] |
| Wang 2022b | 0.715354 | 0.043015 | 10.6% | 2.04 [1.88, 2.22] |
| Xie 2022b | 0.494319 | 0.027525 | 10.8% | 1.64 [1.55, 1.73] |
| Yang 2024 | 0.21486 | 0.01727 | 10.9% | 1.24 [1.20, 1.28] |
| Zang 2023, INSIGHT cohort | 0.2389 | 0.0423 | 10.6% | 1.27 [1.17, 1.38] |
| Zappacosta 2022, 61-70 y | -0.0202 | 0.15267 | 7.7% | 0.98 [0.73, 1.32] |
| Zappacosta 2022, 70+ y | -0.2485 | 0.08638 | 9.6% | 0.78 [0.66, 0.92] |
| **Subtotal (95% CI)** | | | **100.0%** | **1.33 [1.14, 1.54]** |
| Heterogeneity: Tau² = 0.05; Chi² = 241.39, df = 10 (P < 0.00001); I² = 96% | | | | |
| Test for overall effect: Z = 3.64 (P = 0.0003) | | | | |

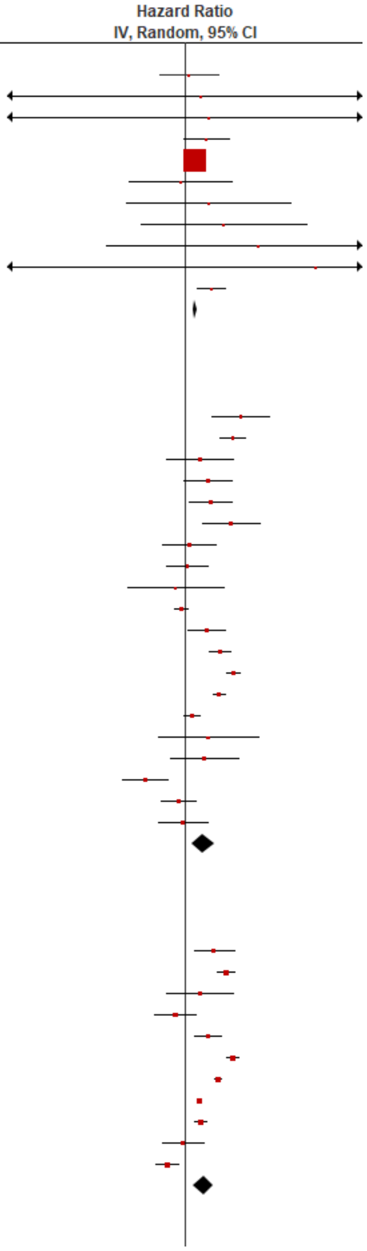

| | | | | |
|---|---|---|---|---|
| **2.1.4 Cardiovascular disorders, overall (inpatients, 18-64 y)** | | | | |
| Daugherty 2021 | 1.42149 | 0.10551 | 18.9% | 4.14 [3.37, 5.10] |
| Ho 2021 | 0.182322 | 0.725333 | 9.1% | 1.20 [0.29, 4.97] |
| Lee 2023, 18-39 y | 0.833253 | 0.228239 | 17.4% | 2.30 [1.47, 3.60] |
| Lee 2023, 40-64 y | -0.45089 | 0.132605 | 18.6% | 0.64 [0.49, 0.83] |
| Salah 2022 | 0.425268 | 0.033177 | 19.3% | 1.53 [1.43, 1.63] |
| Velasquez Garcia 2024 | 1.684545 | 0.267412 | 16.8% | 5.39 [3.19, 9.10] |
| **Subtotal (95% CI)** | | | **100.0%** | **2.03 [1.13, 3.66]** |
| Heterogeneity: Tau² = 0.46; Chi² = 154.46, df = 5 (P < 0.00001); I² = 97% | | | | |
| Test for overall effect: Z = 2.37 (P = 0.02) | | | | |
| **2.1.5 Cardiovascular disorders, overall (inpatients, ≥ 65 y)** | | | | |
| Cohen 2022 | 0.936224 | 0.048991 | 27.4% | 2.55 [2.32, 2.81] |
| Ho 2021, 66-80 y | -0.6746 | 0.16321 | 26.8% | 0.51 [0.37, 0.70] |
| Ho 2021, 80+ y | -0.4308 | 0.71803 | 18.7% | 0.65 [0.16, 2.66] |
| Lee 2023 | -0.5865 | 0.09818 | 27.2% | 0.56 [0.46, 0.67] |
| **Subtotal (95% CI)** | | | **100.0%** | **0.85 [0.29, 2.49]** |
| Heterogeneity: Tau² = 1.10; Chi² = 254.27, df = 3 (P < 0.00001); I² = 99% | | | | |
| Test for overall effect: Z = 0.30 (P = 0.76) | | | | |

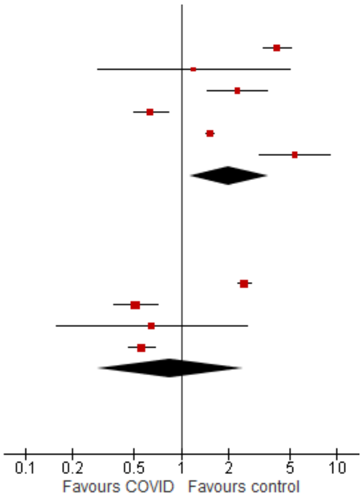

Test for subgroup differences: Chi² = 7.88, df = 4 (P = 0.10), I² = 49.2%

#### Acute coronary disease

| Study or Subgroup | log[Hazard Ratio] | SE | Weight | Hazard Ratio IV, Random, 95% CI |
|---|---|---|---|---|
| **2.2.1 Acute coronary disease (outpatients/mixed, <18 y)** | | | | |
| Gordon 2024, 0-1 y | 0.287682 | 0.381718 | 8.2% | 1.33 [0.63, 2.82] |
| Gordon 2024, 12-17 y | 0.267315 | 0.168705 | 42.2% | 1.31 [0.94, 1.82] |
| Gordon 2024, 2-4 y | 0.223143 | 0.387205 | 8.0% | 1.25 [0.59, 2.67] |
| Gordon 2024, 5-11 y | 0.476924 | 0.300027 | 13.4% | 1.61 [0.89, 2.90] |
| Mizrahi 2023, 12-18 y | 0.336472 | 0.614454 | 3.2% | 1.40 [0.42, 4.67] |
| Roessler 2022 | 1.199965 | 1.05458 | 1.1% | 3.32 [0.42, 26.23] |
| Zhang 2024 | 0.530794 | 0.224256 | 23.9% | 1.70 [1.10, 2.64] |
| **Subtotal (95% CI)** | | | **100.0%** | **1.45 [1.17, 1.79]** |
| Heterogeneity: Tau² = 0.00; Chi² = 1.82, df = 6 (P = 0.94); I² = 0% | | | | |
| Test for overall effect: Z = 3.36 (P = 0.0008) | | | | |
| **2.2.2 Acute coronary disease (outpatients/mixed, 18-64 y)** | | | | |
| Batistoni 2024 | 0.530628 | 0.281988 | 6.8% | 1.70 [0.98, 2.95] |
| Daugherty 2021 | 0.802002 | 0.156599 | 9.2% | 2.23 [1.64, 3.03] |
| Lam 2023 | 0.274072 | 0.188926 | 8.6% | 1.32 [0.91, 1.90] |
| Lim 2023 | 1.652497 | 0.377588 | 5.2% | 5.22 [2.49, 10.94] |
| Mizrahi 2023, 19-40 y | 0.250003 | 0.326955 | 6.0% | 1.28 [0.68, 2.44] |
| Mizrahi 2023, 41-60 y | -0.33604 | 0.147626 | 9.4% | 0.71 [0.54, 0.95] |
| Mizrahi 2023, 60+ y | -0.28462 | 0.384812 | 5.1% | 0.75 [0.35, 1.60] |
| Rezel-Potts 2022 | -0.17732 | 0.123467 | 9.8% | 0.84 [0.66, 1.07] |
| Velasquez Garcia 2024 | 0.329304 | 0.1379 | 9.6% | 1.39 [1.06, 1.82] |
| Wang 2022b, 20-44y | 0.372339 | 0.138893 | 9.5% | 1.45 [1.11, 1.91] |
| Wang 2022b, 45-64y | 0.662593 | 0.086399 | 10.4% | 1.94 [1.64, 2.30] |
| Xie 2022b | 0.506818 | 0.070291 | 10.6% | 1.66 [1.45, 1.91] |
| **Subtotal (95% CI)** | | | **100.0%** | **1.43 [1.14, 1.81]** |
| Heterogeneity: Tau² = 0.13; Chi² = 80.19, df = 11 (P < 0.00001); I² = 86% | | | | |
| Test for overall effect: Z = 3.04 (P = 0.002) | | | | |
| **2.2.3 Acute coronary disease (outpatients/mixed, ≥ 65 y)** | | | | |
| Batistoni 2024 | 1.1474 | 0.29663 | 7.0% | 3.15 [1.76, 5.63] |
| Cohen 2022 | 0.732368 | 0.088939 | 15.8% | 2.08 [1.75, 2.48] |
| Mizrahi 2023, 60+ y | -0.2846 | 0.16795 | 12.0% | 0.75 [0.54, 1.05] |
| Wan 2023a, UK cohort | 0.48734 | 0.12869 | 13.9% | 1.63 [1.27, 2.10] |
| Wang 2022b | 0.589343 | 0.084561 | 16.0% | 1.80 [1.53, 2.13] |
| Xie 2022b | 0.470004 | 0.031731 | 17.7% | 1.60 [1.50, 1.70] |
| Yang 2024 | 0.18462 | 0.02869 | 17.7% | 1.20 [1.14, 1.27] |
| **Subtotal (95% CI)** | | | **100.0%** | **1.55 [1.27, 1.89]** |
| Heterogeneity: Tau² = 0.06; Chi² = 96.66, df = 6 (P < 0.00001); I² = 94% | | | | |
| Test for overall effect: Z = 4.37 (P < 0.0001) | | | | |
| **2.2.4 Acute coronary disease (inpatients, 18-64 y)** | | | | |
| Daugherty 2021 | 1.781288 | 0.191085 | 21.5% | 5.94 [4.08, 8.63] |
| Ho 2021 | 0.182322 | 0.725333 | 16.1% | 1.20 [0.29, 4.97] |
| Lee 2023, 18-39 y | 1.721862 | 0.348452 | 20.3% | 5.59 [2.83, 11.08] |
| Lee 2023, 40-64 y | -0.57478 | 0.22663 | 21.2% | 0.56 [0.36, 0.88] |
| Velasquez Garcia 2024 | 1.684545 | 0.267412 | 21.0% | 5.39 [3.19, 9.10] |
| **Subtotal (95% CI)** | | | **100.0%** | **2.70 [0.90, 8.06]** |
| Heterogeneity: Tau² = 1.42; Chi² = 76.81, df = 4 (P < 0.00001); I² = 95% | | | | |
| Test for overall effect: Z = 1.77 (P = 0.08) | | | | |
| **2.2.5 Acute coronary disease (inpatients, ≥ 65 y)** | | | | |
| Cohen 2022 | 1.090681 | 0.072668 | 28.1% | 2.98 [2.58, 3.43] |
| Ho 2021, 66-80 y | -0.6746 | 0.16321 | 27.7% | 0.51 [0.37, 0.70] |
| Ho 2021, 80+ y | -0.4308 | 0.71803 | 20.6% | 0.65 [0.16, 2.66] |
| Lee 2023 | -0.0408 | 0.52574 | 23.6% | 0.96 [0.34, 2.69] |
| **Subtotal (95% CI)** | | | **100.0%** | **1.02 [0.30, 3.49]** |
| Heterogeneity: Tau² = 1.39; Chi² = 102.99, df = 3 (P < 0.00001); I² = 97% | | | | |
| Test for overall effect: Z = 0.03 (P = 0.97) | | | | |

Hazard Ratio
IV, Random, 95% CI

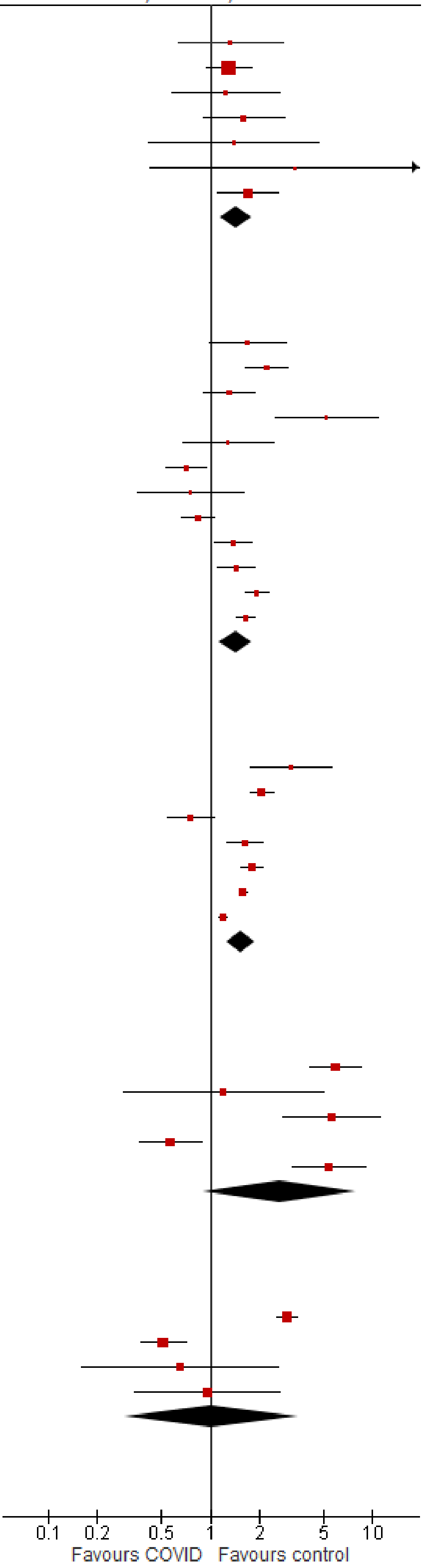

Test for subgroup differences: Chi² = 1.84, df = 4 (P = 0.77), I² = 0%

#### Arrhythmia/dysrhythmias

| Study or Subgroup | log[Hazard Ratio] | SE | Weight | Hazard Ratio IV, Random, 95% CI |
|---|---|---|---|---|
| **2.3.1 Arrhythmia/dysrhythmia (outpatients/mixed, <18 y)** | | | | |
| Gordon 2024, 0-1 y | 0.057432 | 0.097299 | 11.8% | 1.06 [0.88, 1.28] |
| Gordon 2024, 12-17 y | 0.207361 | 0.039932 | 19.3% | 1.23 [1.14, 1.33] |
| Gordon 2024, 2-4 y | 0.518563 | 0.124121 | 9.1% | 1.68 [1.32, 2.14] |
| Gordon 2024, 5-11 y | 0.310877 | 0.078909 | 14.0% | 1.36 [1.17, 1.59] |
| Kompaniyets 2022 | 0.14842 | 0.0176 | 21.5% | 1.16 [1.12, 1.20] |
| Mizrahi 2023, 12-18 y | -0.58949 | 0.340836 | 1.9% | 0.55 [0.28, 1.08] |
| Roessler 2022 | 0.360835 | 0.161986 | 6.4% | 1.43 [1.04, 1.97] |
| Tartof 2022 | 0.576613 | 0.263687 | 3.0% | 1.78 [1.06, 2.98] |
| Valdivieso-Martinez 2023, unvaccinated cohort | 1.098612 | 1.154739 | 0.2% | 3.00 [0.31, 28.84] |
| Valdivieso-Martinez 2023, vaccinated cohort | 1.94591 | 3.75576 | 0.0% | 7.00 [0.00, 11015.56] |
| Zhang 2024 | 0.385262 | 0.087884 | 12.9% | 1.47 [1.24, 1.75] |
| **Subtotal (95% CI)** | | | **100.0%** | **1.28 [1.17, 1.41]** |
| Heterogeneity: Tau² = 0.01; Chi² = 30.28, df = 10 (P = 0.0008); I² = 67% | | | | |
| Test for overall effect: Z = 5.14 (P < 0.00001) | | | | |
| **2.3.2 Arrhythmia/dysrhythmia (outpatients/mixed, 18-64 y)** | | | | |
| Batistoni 2024 | 0.916291 | 0.166762 | 7.2% | 2.50 [1.80, 3.47] |
| Daugherty 2021 | 0.916291 | 0.061931 | 12.5% | 2.50 [2.21, 2.82] |
| Katsoularis 2023, 60+ y | 0.227813 | 0.25172 | 4.4% | 1.26 [0.77, 2.06] |
| Katsoularis 2023, 60 y & under | 0.348127 | 0.183357 | 6.5% | 1.42 [0.99, 2.03] |
| Lam 2023 | 0.285179 | 0.217401 | 5.4% | 1.33 [0.87, 2.04] |
| Mizrahi 2023, 19-40 y | -0.07429 | 0.223295 | 5.2% | 0.93 [0.60, 1.44] |
| Mizrahi 2023, 41-60 y | 0.140799 | 0.174525 | 6.9% | 1.15 [0.82, 1.62] |
| Mizrahi 2023, 60+ y | 0.007484 | 0.410256 | 2.1% | 1.01 [0.45, 2.25] |
| Rezel-Potts 2022 | 0.001457 | 0.129349 | 9.0% | 1.00 [0.78, 1.29] |
| Wang 2022b, 20-44y | 0.445361 | 0.038156 | 13.4% | 1.56 [1.45, 1.68] |
| Wang 2022b, 45-64y | 0.577963 | 0.030978 | 13.7% | 1.78 [1.68, 1.89] |
| Xie 2022b | 0.518794 | 0.027359 | 13.8% | 1.68 [1.59, 1.77] |
| **Subtotal (95% CI)** | | | **100.0%** | **1.56 [1.38, 1.77]** |
| Heterogeneity: Tau² = 0.03; Chi² = 85.99, df = 11 (P < 0.00001); I² = 87% | | | | |
| Test for overall effect: Z = 6.95 (P < 0.00001) | | | | |
| **2.3.3 Arrhythmia/dysrhythmia (outpatients/mixed, ≥ 65 y)** | | | | |
| Batistoni 2024 | 0.3001 | 0.08647 | 13.3% | 1.35 [1.14, 1.60] |
| Cohen 2022 | 0.652325 | 0.056924 | 14.1% | 1.92 [1.72, 2.15] |
| Katsoularis 2023, 60+ y | 0.22781 | 0.25172 | 7.4% | 1.26 [0.77, 2.06] |
| Mizrahi 2023, 60+ y | 0.00748 | 0.17905 | 9.9% | 1.01 [0.71, 1.43] |
| Wan 2023a, UK cohort | 0.04038 | 0.14434 | 11.2% | 1.04 [0.78, 1.38] |
| Wang 2022b | 0.665519 | 0.03084304 | 14.6% | 1.95 [1.83, 2.07] |
| Xie 2022b | 0.518794 | 0.031834 | 14.6% | 1.68 [1.58, 1.79] |
| Yang 2024 | 0.21486 | 0.01365 | 14.8% | 1.24 [1.21, 1.27] |
| **Subtotal (95% CI)** | | | **100.0%** | **1.43 [1.18, 1.73]** |
| Heterogeneity: Tau² = 0.06; Chi² = 264.27, df = 7 (P < 0.00001); I² = 97% | | | | |
| Test for overall effect: Z = 3.69 (P = 0.0002) | | | | |
| **2.3.4 Arrhythmia/dysrhythmia (inpatients, 18-64 y)** | | | | |
| Daugherty 2021 | 1.909543 | 0.081702 | 34.0% | 6.75 [5.75, 7.92] |
| Lee 2023, 18-39 y | 0.631272 | 0.303679 | 32.4% | 1.88 [1.04, 3.41] |
| Lee 2023, 40-64 y | -0.17435 | 0.172536 | 33.6% | 0.84 [0.60, 1.18] |
| **Subtotal (95% CI)** | | | **100.0%** | **2.22 [0.49, 10.01]** |
| Heterogeneity: Tau² = 1.73; Chi² = 127.39, df = 2 (P < 0.00001); I² = 98% | | | | |
| Test for overall effect: Z = 1.03 (P = 0.30) | | | | |
| **2.3.5 Arrhythmia/dysrhythmia (inpatients, ≥ 65 y)** | | | | |
| Cohen 2022 | 1.198216 | 0.04372 | 50.3% | 3.31 [3.04, 3.61] |
| Lee 2023 | -0.2744 | 0.12254 | 49.7% | 0.76 [0.60, 0.97] |
| **Subtotal (95% CI)** | | | **100.0%** | **1.59 [0.38, 6.75]** |
| Heterogeneity: Tau² = 1.08; Chi² = 128.11, df = 1 (P < 0.00001); I² = 99% | | | | |
| Test for overall effect: Z = 0.63 (P = 0.53) | | | | |

Test for subgroup differences: Chi² = 6.37, df = 4 (P = 0.17), I² = 37.2%

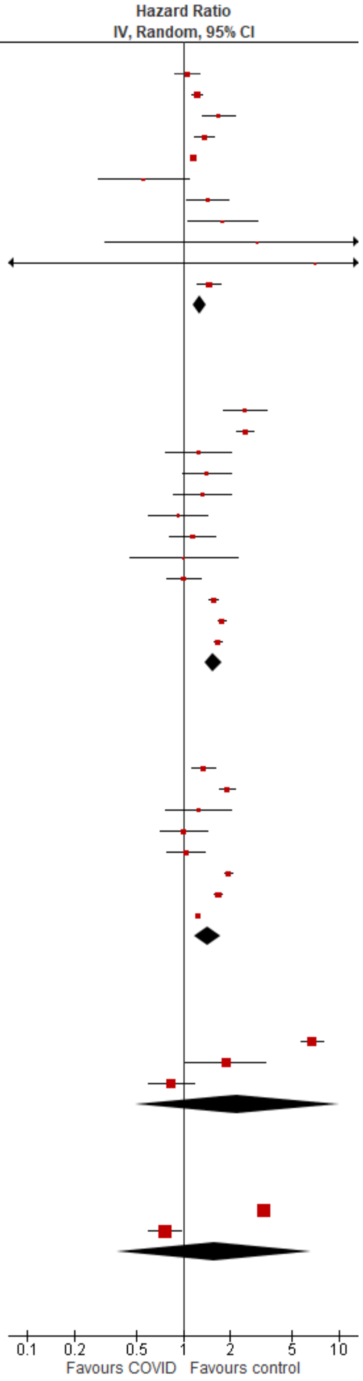

#### Cardiomyopathy

| Study or Subgroup | log[Hazard Ratio] | SE | Weight | Hazard Ratio IV, Random, 95% CI |
|---|---|---|---|---|
| **2.4.1 Cardiomyopathy (outpatients/mixed, <18 y)** | | | | |
| Zhang 2024 | 0.405465 | 0.119362 | 100.0% | 1.50 [1.19, 1.90] |
| **Subtotal (95% CI)** | | | **100.0%** | **1.50 [1.19, 1.90]** |
| Heterogeneity: Not applicable | | | | |
| Test for overall effect: Z = 3.40 (P = 0.0007) | | | | |
| **2.4.2 Cardiomyopathy (outpatients/mixed, 18-64 y)** | | | | |
| Daugherty 2021 | 1.18479 | 0.175721 | 22.6% | 3.27 [2.32, 4.61] |
| Wang 2022b, 20-44y | 0.777391 | 0.168159 | 24.1% | 2.18 [1.56, 3.03] |
| Wang 2022b, 45-64y | 1.083469 | 0.07449 | 53.3% | 2.95 [2.55, 3.42] |
| **Subtotal (95% CI)** | | | **100.0%** | **2.81 [2.31, 3.42]** |
| Heterogeneity: Tau² = 0.01; Chi² = 3.42, df = 2 (P = 0.18); I² = 41% | | | | |
| Test for overall effect: Z = 10.35 (P < 0.00001) | | | | |
| **2.4.3 Cardiomyopathy (outpatients/mixed, ≥ 65 y)** | | | | |
| Cohen 2022 | 0.385262 | 0.102014 | 32.6% | 1.47 [1.20, 1.80] |
| Wang 2022b | 0.89155598 | 0.067941 | 33.4% | 2.44 [2.13, 2.79] |
| Yang 2024 | 0.076603 | 0.022796 | 34.0% | 1.08 [1.03, 1.13] |
| **Subtotal (95% CI)** | | | **100.0%** | **1.57 [0.90, 2.74]** |
| Heterogeneity: Tau² = 0.24; Chi² = 134.03, df = 2 (P < 0.00001); I² = 99% | | | | |
| Test for overall effect: Z = 1.58 (P = 0.11) | | | | |
| **2.4.4 Cardiomyopathy (inpatients, 18-64 y)** | | | | |
| Daugherty 2021 | 1.728837 | 0.169466 | 100.0% | 5.63 [4.04, 7.85] |
| **Subtotal (95% CI)** | | | **100.0%** | **5.63 [4.04, 7.85]** |
| Heterogeneity: Not applicable | | | | |
| Test for overall effect: Z = 10.20 (P < 0.00001) | | | | |
| **2.4.5 Cardiomyopathy (inpatients, ≥ 65 y)** | | | | |
| Cohen 2022 | 0.614699 | 0.081488 | 48.9% | 1.85 [1.58, 2.17] |
| Yang 2024 | 0.0766 | 0.0228 | 51.1% | 1.08 [1.03, 1.13] |
| **Subtotal (95% CI)** | | | **100.0%** | **1.40 [0.83, 2.38]** |
| Heterogeneity: Tau² = 0.14; Chi² = 40.44, df = 1 (P < 0.00001); I² = 98% | | | | |
| Test for overall effect: Z = 1.26 (P = 0.21) | | | | |

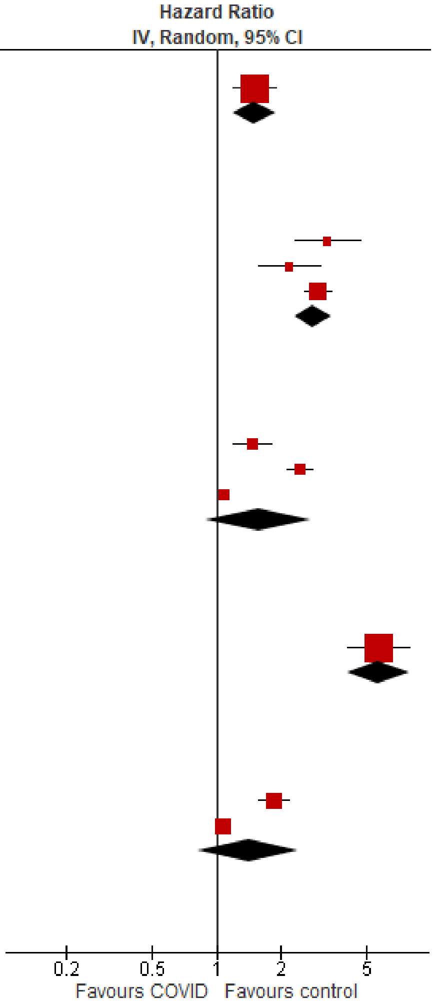

Test for subgroup differences: Chi² = 49.56, df = 4 (P < 0.00001), I² = 91.9%

#### Heart failure

| Study or Subgroup | log[Hazard Ratio] | SE | Weight | Hazard Ratio IV, Random, 95% CI |
|---|---|---|---|---|
| **2.5.1 Heart failure (outpatients/mixed, <18 y)** | | | | |
| Roessler 2022 | -0.57982 | 0.993461 | 13.6% | 0.56 [0.08, 3.92] |
| Zhang 2024 | 0.565314 | 0.176823 | 86.4% | 1.76 [1.24, 2.49] |
| **Subtotal (95% CI)** | | | **100.0%** | **1.51 [0.70, 3.25]** |
| Heterogeneity: Tau² = 0.15; Chi² = 1.29, df = 1 (P = 0.26); I² = 22% | | | | |
| Test for overall effect: Z = 1.05 (P = 0.30) | | | | |
| **2.5.2 Heart failure (outpatients/mixed, 18-64 y)** | | | | |
| Batistoni 2024 | 0.947789 | 0.339347 | 7.6% | 2.58 [1.33, 5.02] |
| Daugherty 2021 | 0.963174 | 0.160982 | 12.4% | 2.62 [1.91, 3.59] |
| Lam 2023 | 0.582216 | 0.194005 | 11.4% | 1.79 [1.22, 2.62] |
| Lim 2023 | 0.615186 | 0.105693 | 13.9% | 1.85 [1.50, 2.28] |
| Rezel-Potts 2022 | -0.04229 | 0.237164 | 10.2% | 0.96 [0.60, 1.53] |
| Wang 2022b, 20-44y | 0.756122 | 0.063915 | 14.7% | 2.13 [1.88, 2.41] |
| Wang 2022b, 45-64y | 0.836381 | 0.035453 | 15.1% | 2.31 [2.15, 2.47] |
| Zang 2023, INSIGHT cohort | 0.110557 | 0.063558 | 14.7% | 1.12 [0.99, 1.27] |
| **Subtotal (95% CI)** | | | **100.0%** | **1.81 [1.40, 2.34]** |
| Heterogeneity: Tau² = 0.11; Chi² = 114.74, df = 7 (P < 0.00001); I² = 94% | | | | |
| Test for overall effect: Z = 4.50 (P < 0.00001) | | | | |
| **2.5.3 Heart failure (outpatients/mixed, ≥ 65 y)** | | | | |
| Batistoni 2024 | 0.67294 | 0.15286 | 14.7% | 1.96 [1.45, 2.64] |
| Cohen 2022 | 0.615186 | 0.0714 | 17.0% | 1.85 [1.61, 2.13] |
| Wan 2023a, UK cohort | 0.47 | 0.13678 | 15.3% | 1.60 [1.22, 2.09] |
| Wang 2022b | 0.744315 | 0.030161 | 17.7% | 2.10 [1.98, 2.23] |
| Yang 2024 | 0.139 | 0.02173 | 17.8% | 1.15 [1.10, 1.20] |
| Zang 2023, INSIGHT cohort | 0.2389 | 0.0423 | 17.5% | 1.27 [1.17, 1.38] |
| **Subtotal (95% CI)** | | | **100.0%** | **1.61 [1.22, 2.12]** |
| Heterogeneity: Tau² = 0.11; Chi² = 291.13, df = 5 (P < 0.00001); I² = 98% | | | | |
| Test for overall effect: Z = 3.37 (P = 0.0007) | | | | |
| **2.5.4 Heart failure (inpatients, 18-64 y)** | | | | |
| Daugherty 2021 | 2.047693 | 0.177093 | 24.4% | 7.75 [5.48, 10.97] |
| Lee 2023, 18-39 y | 0.667829 | 0.179601 | 24.4% | 1.95 [1.37, 2.77] |
| Lee 2023, 40-64 y | -0.4943 | 0.079448 | 25.5% | 0.61 [0.52, 0.71] |
| Salah 2022 | 0.425268 | 0.033177 | 25.7% | 1.53 [1.43, 1.63] |
| **Subtotal (95% CI)** | | | **100.0%** | **1.91 [0.91, 4.02]** |
| Heterogeneity: Tau² = 0.56; Chi² = 213.11, df = 3 (P < 0.00001); I² = 99% | | | | |
| Test for overall effect: Z = 1.70 (P = 0.09) | | | | |
| **2.5.5 Heart failure (inpatients, ≥ 65 y)** | | | | |
| Cohen 2022 | 0.956473 | 0.053257 | 50.0% | 2.60 [2.34, 2.89] |
| Lee 2023 | -0.6733 | 0.04047 | 50.0% | 0.51 [0.47, 0.55] |
| **Subtotal (95% CI)** | | | **100.0%** | **1.15 [0.23, 5.69]** |
| Heterogeneity: Tau² = 1.33; Chi² = 593.67, df = 1 (P < 0.00001); I² = 100% | | | | |
| Test for overall effect: Z = 0.17 (P = 0.86) | | | | |

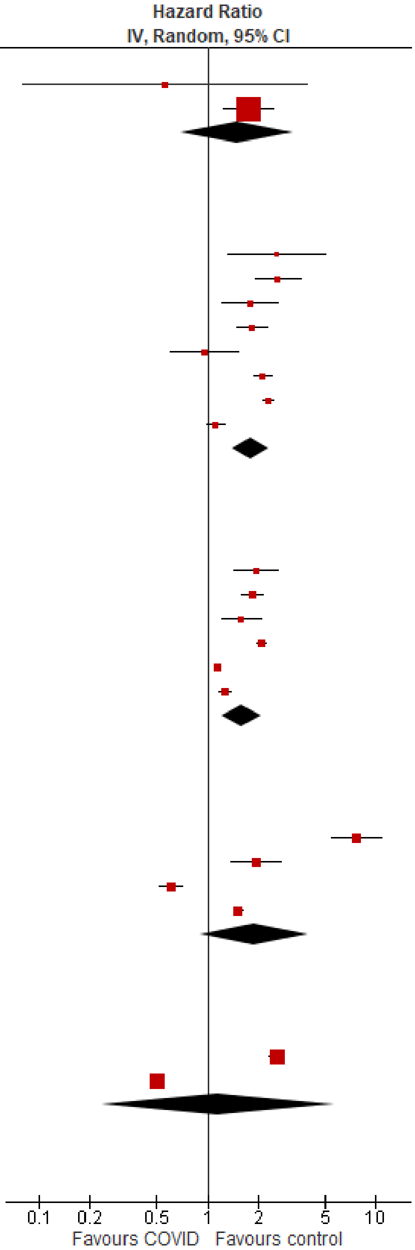

Test for subgroup differences: Chi² = 0.80, df = 4 (P = 0.94), I² = 0%

#### Hypertension

| Study or Subgroup | log[Hazard Ratio] | SE | Weight | Hazard Ratio IV, Random, 95% CI |
|---|---|---|---|---|
| **2.6.1 Hypertension (outpatients/mixed, <18 y)** | | | | |
| Mizrahi 2023, 12-18 y | 0.690675 | 0.456109 | 0.9% | 2.00 [0.82, 4.88] |
| Zhang 2024 | 0.385262 | 0.043342 | 99.1% | 1.47 [1.35, 1.60] |
| **Subtotal (95% CI)** | | | **100.0%** | **1.47 [1.35, 1.60]** |
| Heterogeneity: Tau² = 0.00; Chi² = 0.44, df = 1 (P = 0.51); I² = 0% | | | | |
| Test for overall effect: Z = 8.99 (P < 0.00001) | | | | |
| **2.6.2 Hypertension (outpatients/mixed, 18-64 y)** | | | | |
| Daugherty 2021 | 0.530628 | 0.047878 | 34.6% | 1.70 [1.55, 1.87] |
| Mizrahi 2023, 19-40 y | 0.120958 | 0.186089 | 24.4% | 1.13 [0.78, 1.63] |
| Mizrahi 2023, 41-60 y | 0.110494 | 0.057276 | 34.2% | 1.12 [1.00, 1.25] |
| Mizrahi 2023, 60+ y | 0.124964 | 0.561782 | 6.8% | 1.13 [0.38, 3.41] |
| **Subtotal (95% CI)** | | | **100.0%** | **1.30 [0.94, 1.79]** |
| Heterogeneity: Tau² = 0.07; Chi² = 33.39, df = 3 (P < 0.00001); I² = 91% | | | | |
| Test for overall effect: Z = 1.59 (P = 0.11) | | | | |
| **2.6.3 Hypertension (outpatients/mixed, ≥ 65 y)** | | | | |
| Cohen 2022 | 0.530628 | 0.115643 | 64.2% | 1.70 [1.36, 2.13] |
| Mizrahi 2023, 60+ y | 0.12496 | 0.24518 | 35.8% | 1.13 [0.70, 1.83] |
| **Subtotal (95% CI)** | | | **100.0%** | **1.47 [1.00, 2.15]** |
| Heterogeneity: Tau² = 0.05; Chi² = 2.24, df = 1 (P = 0.13); I² = 55% | | | | |
| Test for overall effect: Z = 1.98 (P = 0.05) | | | | |
| **2.6.4 Hypertension (inpatients, 18-64 y)** | | | | |
| Daugherty 2021 | 1.114748 | 0.051955 | 100.0% | 3.05 [2.75, 3.38] |
| **Subtotal (95% CI)** | | | **100.0%** | **3.05 [2.75, 3.38]** |
| Heterogeneity: Not applicable | | | | |
| Test for overall effect: Z = 21.46 (P < 0.00001) | | | | |
| **2.6.5 Hypertension (inpatients, ≥ 65 y)** | | | | |
| Cohen 2022 | 1.02061 | 0.02485 | 100.0% | 2.77 [2.64, 2.91] |
| **Subtotal (95% CI)** | | | **100.0%** | **2.77 [2.64, 2.91]** |
| Heterogeneity: Not applicable | | | | |
| Test for overall effect: Z = 41.07 (P < 0.00001) | | | | |

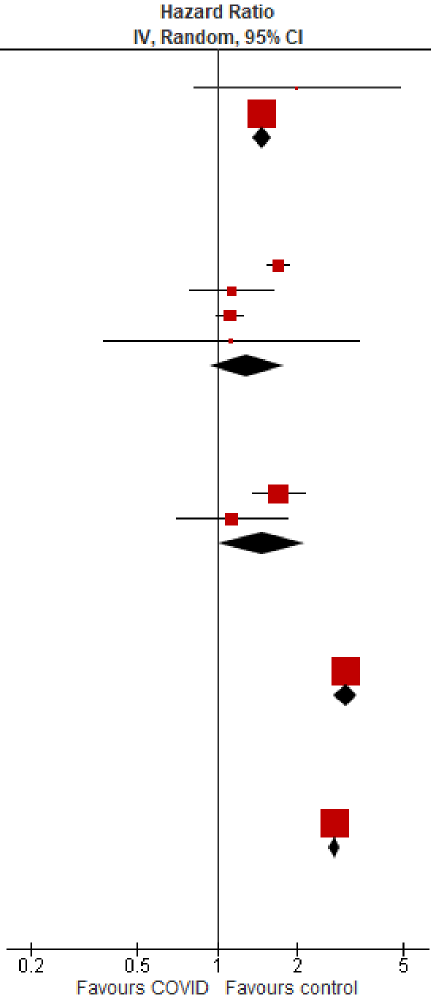

Test for subgroup differences: Chi² = 203.26, df = 4 (P < 0.00001), I² = 98.0%

### Chronic kidney disease

#### Overall

| Study or Subgroup | log[Hazard Ratio] | SE | Weight | Hazard Ratio IV, Random, 95% CI |
|---|---|---|---|---|
| **3.1.1 Chronic kidney disease (outpatients/mixed, <18 y)** | | | | |
| Kompaniyets 2022 | 0.067659 | 0.063812 | 100.0% | 1.07 [0.94, 1.21] |
| **Subtotal (95% CI)** | | | **100.0%** | **1.07 [0.94, 1.21]** |
| Heterogeneity: Not applicable | | | | |
| Test for overall effect: Z = 1.06 (P = 0.29) | | | | |
| **3.1.2 Chronic kidney disease (outpatients/mixed, 18-64 y)** | | | | |
| Daugherty 2021 | 0.470004 | 0.108008 | 65.9% | 1.60 [1.29, 1.98] |
| Lam 2023 | 0.530628 | 0.437449 | 13.1% | 1.70 [0.72, 4.01] |
| Mizrahi 2023, 41-60 y | -0.3899 | 0.526782 | 9.5% | 0.68 [0.24, 1.90] |
| Mizrahi 2023, 60+ y | -0.08726 | 0.471534 | 11.5% | 0.92 [0.36, 2.31] |
| **Subtotal (95% CI)** | | | **100.0%** | **1.39 [1.00, 1.95]** |
| Heterogeneity: Tau² = 0.03; Chi² = 3.80, df = 3 (P = 0.28); I² = 21% | | | | |
| Test for overall effect: Z = 1.94 (P = 0.05) | | | | |
| **3.1.3 Chronic kidney disease (outpatients/mixed, ≥ 65 y)** | | | | |
| Cohen 2022 | 0.307485 | 0.061521 | 98.6% | 1.36 [1.21, 1.53] |
| Mizrahi 2023, 60+ y | -0.08726 | 0.514495 | 1.4% | 0.92 [0.33, 2.51] |
| **Subtotal (95% CI)** | | | **100.0%** | **1.35 [1.20, 1.52]** |
| Heterogeneity: Tau² = 0.00; Chi² = 0.58, df = 1 (P = 0.45); I² = 0% | | | | |
| Test for overall effect: Z = 4.94 (P < 0.00001) | | | | |
| **3.1.4 Chronic kidney disease (inpatients, 18-64 y)** | | | | |
| Ayoubkhani 2021, 18-69 y | 1.252763 | 0.142831 | 34.2% | 3.50 [2.65, 4.63] |
| Daugherty 2021 | 1.085291 | 0.103034 | 65.8% | 2.96 [2.42, 3.62] |
| **Subtotal (95% CI)** | | | **100.0%** | **3.13 [2.66, 3.69]** |
| Heterogeneity: Tau² = 0.00; Chi² = 0.90, df = 1 (P = 0.34); I² = 0% | | | | |
| Test for overall effect: Z = 13.67 (P < 0.00001) | | | | |
| **3.1.5 Chronic kidney disease (inpatients, ≥ 65 y)** | | | | |
| Ayoubkhani 2021, 18-69 y | 1.252763 | 0.490074 | 0.9% | 3.50 [1.34, 9.15] |
| Ayoubkhani 2021, 70+ y | 0.470004 | 0.064111 | 37.3% | 1.60 [1.41, 1.81] |
| Cohen 2022 | 0.51408 | 0.041514 | 61.7% | 1.67 [1.54, 1.81] |
| **Subtotal (95% CI)** | | | **100.0%** | **1.66 [1.51, 1.82]** |
| Heterogeneity: Tau² = 0.00; Chi² = 2.67, df = 2 (P = 0.26); I² = 25% | | | | |
| Test for overall effect: Z = 10.62 (P < 0.00001) | | | | |

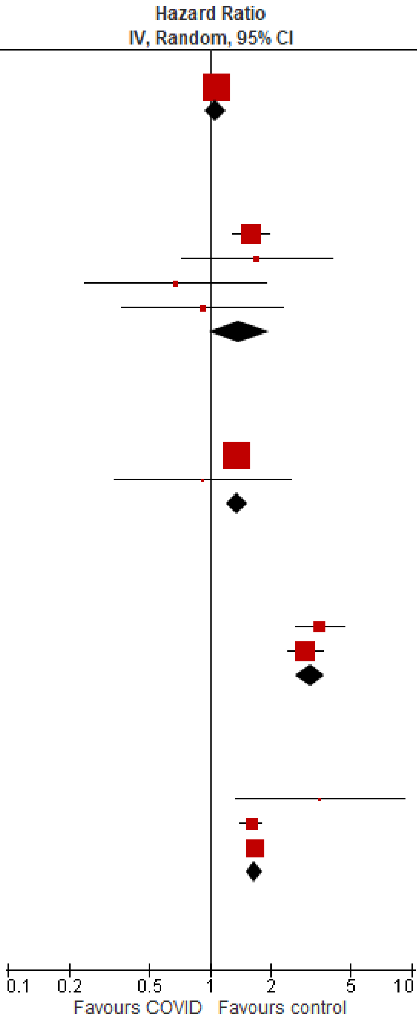

Test for subgroup differences: Chi² = 112.09, df = 4 (P < 0.00001), I² = 96.4%

### Diabetes

#### Overall

| Study or Subgroup | log[Hazard Ratio] | SE | Weight | Hazard Ratio IV, Random, 95% CI |
|---|---|---|---|---|
| **4.1.1 Diabetes overall (outpatients/mixed, <18 y)** | | | | |
| Gordon 2024, 0-1 y | 0.920402 | 0.730235 | 1.0% | 2.51 [0.60, 10.50] |
| Gordon 2024, 12-17 y | 0.177811 | 0.101203 | 15.7% | 1.19 [0.98, 1.46] |
| Gordon 2024, 2-4 y | 0.828814 | 0.385061 | 3.3% | 2.29 [1.08, 4.87] |
| Gordon 2024, 5-11 y | 0.362418 | 0.159062 | 11.2% | 1.44 [1.05, 1.96] |
| Kompaniyets 2022 | 0.171211 | 0.029592 | 20.9% | 1.19 [1.12, 1.26] |
| Mizrahi 2023, 12-18 y | 0.431782 | 0.485066 | 2.2% | 1.54 [0.60, 3.98] |
| Noorzae 2023, 0-4 y | 0.086178 | 0.28434 | 5.4% | 1.09 [0.62, 1.90] |
| Noorzae 2023, 11-17 y | -0.09431 | 0.159087 | 11.2% | 0.91 [0.67, 1.24] |
| Noorzae 2023, 5-10 y | -0.27444 | 0.138961 | 12.6% | 0.76 [0.58, 1.00] |
| Pietropaolo 2022 | 0.188465 | 0.202801 | 8.6% | 1.21 [0.81, 1.80] |
| Tartof 2022 | -0.35667 | 0.215354 | 8.0% | 0.70 [0.46, 1.07] |
| **Subtotal (95% CI)** | | | **100.0%** | **1.10 [0.95, 1.28]** |
| Heterogeneity: Tau² = 0.03; Chi² = 24.24, df = 10 (P = 0.007); I² = 59% | | | | |
| Test for overall effect: Z = 1.28 (P = 0.20) | | | | |
| **4.1.2 Diabetes overall (outpatients/mixed, 18-64 y)** | | | | |
| Chang 2023, 18-40 y | 1.022451 | 0.056719 | 6.0% | 2.78 [2.49, 3.11] |
| Chang 2023, 41-64 y | 0.951658 | 0.050104 | 6.0% | 2.59 [2.35, 2.86] |
| Choi 2023 | 0.431782 | 0.021478 | 6.1% | 1.54 [1.48, 1.61] |
| Daugherty 2021 | 0.604316 | 0.127551 | 5.4% | 1.83 [1.43, 2.35] |
| Mizrahi 2023, 19-40 y | 0.301483 | 0.296006 | 3.5% | 1.35 [0.76, 2.41] |
| Mizrahi 2023, 41-60 y | 0.115372 | 0.125237 | 5.4% | 1.12 [0.88, 1.43] |
| Mizrahi 2023, 60+ y | 0.05005 | 0.372991 | 2.8% | 1.05 [0.51, 2.18] |
| Naveed 2023 | 0.157004 | 0.04811 | 6.0% | 1.17 [1.06, 1.29] |
| Pietropaolo 2022 | -0.07035 | 0.152822 | 5.1% | 0.93 [0.69, 1.26] |
| Rezel-Potts 2022 | 0.110593 | 0.048347 | 6.0% | 1.12 [1.02, 1.23] |
| Taylor 2024, pre-vaccination cohort, 18-39 y | 0.395044 | 0.075306 | 5.8% | 1.48 [1.28, 1.72] |
| Taylor 2024, pre-vaccination cohort, 40-59 y | 0.212603 | 0.035455 | 6.0% | 1.24 [1.15, 1.33] |
| Taylor 2024, pre-vaccination cohort, 60-79 y | 0.252617 | 0.087333 | 5.7% | 1.29 [1.08, 1.53] |
| Taylor 2024, unvaccinated cohort, 40-59 y | 0.604316 | 0.153076 | 5.1% | 1.83 [1.36, 2.47] |
| Taylor 2024, unvaccinated cohort, 60-79 y | 0.883768 | 0.54832 | 1.7% | 2.42 [0.83, 7.09] |
| Taylor 2024, vaccinated cohort, 40-59 y | 0.173953 | 0.059796 | 5.9% | 1.19 [1.06, 1.34] |
| Taylor 2024, vaccinated cohort, 60-79 y | 0.223144 | 0.146779 | 5.2% | 1.25 [0.94, 1.67] |
| Xie 2022a | 0.307485 | 0.018766 | 6.1% | 1.36 [1.31, 1.41] |
| Zang 2023, INSIGHT cohort | 1.308333 | 0.041054 | 6.0% | 3.70 [3.41, 4.01] |
| **Subtotal (95% CI)** | | | **100.0%** | **1.53 [1.29, 1.81]** |
| Heterogeneity: Tau² = 0.12; Chi² = 878.07, df = 18 (P < 0.00001); I² = 98% | | | | |
| Test for overall effect: Z = 4.95 (P < 0.00001) | | | | |
| **4.1.3 Diabetes overall (outpatients/mixed, ≥ 65 y)** | | | | |
| Chang 2023 | 1.181727 | 0.070225 | 12.2% | 3.26 [2.84, 3.74] |
| Cohen 2022 | 0.672944 | 0.103435 | 11.2% | 1.96 [1.60, 2.40] |
| Mizrahi 2023, 60+ y | 0.05005 | 0.837214 | 1.0% | 1.05 [0.20, 5.42] |
| Taylor 2024, pre-vaccination cohort, 60-79 y | 0.252617 | 0.050422 | 12.7% | 1.29 [1.17, 1.42] |
| Taylor 2024, pre-vaccination cohort, 80-110 y | 0.240626 | 0.093384 | 11.6% | 1.27 [1.06, 1.53] |
| Taylor 2024, unvaccinated cohort, 60-79 y | 0.883768 | 0.316573 | 4.9% | 2.42 [1.30, 4.50] |
| Taylor 2024, vaccinated cohort, 60-79 y | 0.223144 | 0.084743 | 11.8% | 1.25 [1.06, 1.48] |
| Taylor 2024, vaccination cohort, 80-110 y | 0.350657 | 0.184439 | 8.4% | 1.42 [0.99, 2.04] |
| Xie 2022a | 0.357674 | 0.02142 | 13.2% | 1.43 [1.37, 1.49] |
| Zang 2023, INSIGHT cohort | 0.401926 | 0.037497 | 13.0% | 1.49 [1.39, 1.61] |
| **Subtotal (95% CI)** | | | **100.0%** | **1.61 [1.36, 1.92]** |
| Heterogeneity: Tau² = 0.06; Chi² = 153.17, df = 9 (P < 0.00001); I² = 94% | | | | |
| Test for overall effect: Z = 5.43 (P < 0.00001) | | | | |
| **4.1.4 Diabetes overall (inpatients, 18-64 y)** | | | | |
| Ayoubkhani 2021, 18-69 y | 0.530628 | 0.031574 | 34.6% | 1.70 [1.60, 1.81] |
| Daugherty 2021 | 1.299292 | 0.092114 | 33.4% | 3.67 [3.06, 4.39] |
| Naveed 2023 | 0.883768 | 0.133027 | 32.0% | 2.42 [1.86, 3.14] |
| **Subtotal (95% CI)** | | | **100.0%** | **2.46 [1.45, 4.17]** |
| Heterogeneity: Tau² = 0.21; Chi² = 66.30, df = 2 (P < 0.00001); I² = 97% | | | | |
| Test for overall effect: Z = 3.35 (P = 0.0008) | | | | |
| **4.1.5 Diabetes overall (inpatients, ≥ 65 y)** | | | | |
| Ayoubkhani 2021, 18-69 y | 0.530628 | 0.108337 | 32.6% | 1.70 [1.37, 2.10] |
| Ayoubkhani 2021, 70+ y | 0.262364 | 0.039324 | 33.8% | 1.30 [1.20, 1.40] |
| Cohen 2022 | 1.10942 | 0.055677 | 33.6% | 3.03 [2.72, 3.38] |
| **Subtotal (95% CI)** | | | **100.0%** | **1.89 [1.03, 3.44]** |
| Heterogeneity: Tau² = 0.28; Chi² = 154.44, df = 2 (P < 0.00001); I² = 99% | | | | |
| Test for overall effect: Z = 2.07 (P = 0.04) | | | | |

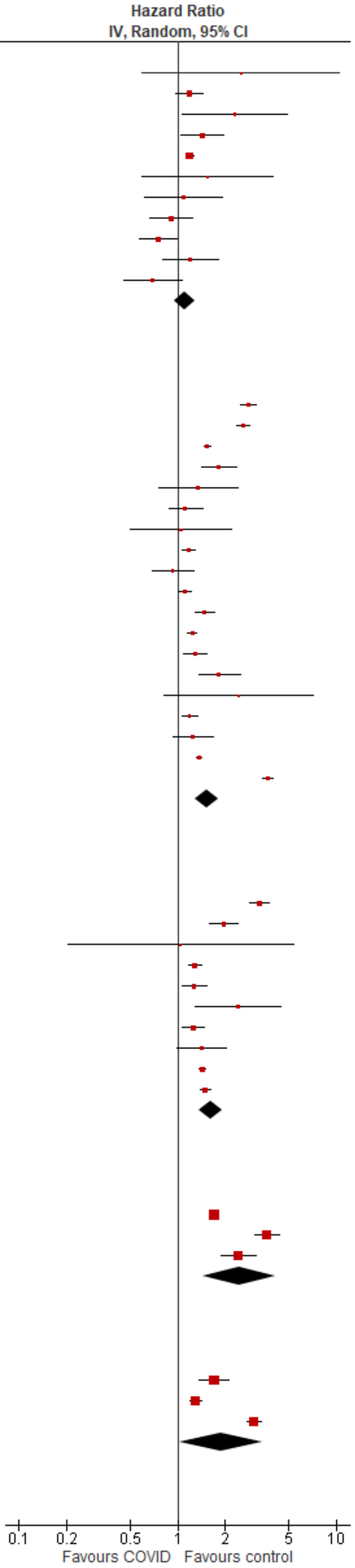

Test for subgroup differences: Chi² = 18.69, df = 4 (P = 0.0009), I² = 78.6%

Type 1

| Study or Subgroup | log[Hazard Ratio] | SE | Weight | Hazard Ratio IV, Random, 95% CI |
|---|---|---|---|---|
| **4.2.1 Diabetes, type 1 (outpatients/mixed, <18 y)** | | | | |
| Kompaniyets 2022 | 0.20701417 | 0.041572 | 31.4% | 1.23 [1.13, 1.33] |
| Noorzae 2023, 0-4 y | 0.086178 | 0.28434 | 11.8% | 1.09 [0.62, 1.90] |
| Noorzae 2023, 11-17 y | -0.09431 | 0.159087 | 21.0% | 0.91 [0.67, 1.24] |
| Noorzae 2023, 5-10 y | -0.27444 | 0.138961 | 23.0% | 0.76 [0.58, 1.00] |
| Pietropaolo 2022 | 0.142367 | 0.268617 | 12.7% | 1.15 [0.68, 1.95] |
| **Subtotal (95% CI)** | | | **100.0%** | **1.01 [0.79, 1.28]** |
| Heterogeneity: $Tau^2 = 0.05$; $Chi^2 = 13.62$, df = 4 (P = 0.009); $I^2 = 71\%$ | | | | |
| Test for overall effect: Z = 0.09 (P = 0.93) | | | | |
| **4.2.2 Diabetes, type 1 (outpatients/mixed, 18-64 y)** | | | | |
| Chang 2023, 18-40 y | 1.022451 | 0.056719 | 38.2% | 2.78 [2.49, 3.11] |
| Chang 2023, 41-64 y | 0.951658 | 0.050104 | 38.5% | 2.59 [2.35, 2.86] |
| Pietropaolo 2022 | -0.62548853 | 0.260083 | 23.3% | 0.54 [0.32, 0.89] |
| **Subtotal (95% CI)** | | | **100.0%** | **1.84 [1.25, 2.71]** |
| Heterogeneity: $Tau^2 = 0.10$; $Chi^2 = 38.33$, df = 2 (P < 0.00001); $I^2 = 95\%$ | | | | |
| Test for overall effect: Z = 3.11 (P = 0.002) | | | | |
| **4.2.3 Diabetes, type 1 (outpatients/mixed, ≥ 65 y)** | | | | |
| Chang 2023 | 1.319086 | 0.070225 | 100.0% | 3.74 [3.26, 4.29] |
| **Subtotal (95% CI)** | | | **100.0%** | **3.74 [3.26, 4.29]** |
| Heterogeneity: Not applicable | | | | |
| Test for overall effect: Z = 18.78 (P < 0.00001) | | | | |

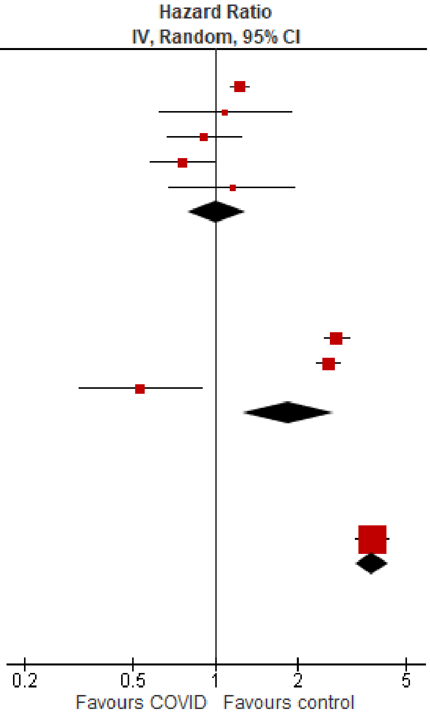

Test for subgroup differences: $Chi^2 = 89.29$, df = 2 (P < 0.00001), $I^2 = 97.8\%$

#### Type 2

| Study or Subgroup | log[Hazard Ratio] | SE | Weight | Hazard Ratio IV, Random, 95% CI |
|---|---|---|---|---|
| **4.3.1 Diabetes, type 2 (outpatients/mixed, <18 y)** | | | | |
| Kompaniyets 2022 | 0.157004 | 0.026187 | 98.3% | 1.17 [1.11, 1.23] |
| Pietropaolo 2022 | 0.213497 | 0.197942 | 1.7% | 1.24 [0.84, 1.82] |
| **Subtotal (95% CI)** | | | **100.0%** | **1.17 [1.11, 1.23]** |
| Heterogeneity: Tau² = 0.00; Chi² = 0.08, df = 1 (P = 0.78); I² = 0% | | | | |
| Test for overall effect: Z = 6.09 (P < 0.00001) | | | | |
| **4.3.2 Diabetes, type 2 (outpatients/mixed, 18-64 y)** | | | | |
| Choi 2023 | 0.431782 | 0.021478 | 13.5% | 1.54 [1.48, 1.61] |
| Daugherty 2021 | 0.60431597 | 0.069371 | 11.7% | 1.83 [1.60, 2.10] |
| Pietropaolo 2022 | -0.02224561 | 0.076557 | 11.3% | 0.98 [0.84, 1.14] |
| Taylor 2024, pre-vaccination cohort, 18-39 y | 0.395044 | 0.075306 | 11.4% | 1.48 [1.28, 1.72] |
| Taylor 2024, pre-vaccination cohort, 40-59 y | 0.212603 | 0.035455 | 13.1% | 1.24 [1.15, 1.33] |
| Taylor 2024, pre-vaccination cohort, 60-79 y | 0.252617 | 0.087333 | 10.7% | 1.29 [1.08, 1.53] |
| Taylor 2024, unvaccinated cohort, 40-59 y | 0.604316 | 0.153076 | 7.4% | 1.83 [1.36, 2.47] |
| Taylor 2024, unvaccinated cohort, 60-79 y | 0.883768 | 0.54832 | 1.1% | 2.42 [0.83, 7.09] |
| Taylor 2024, vaccinated cohort, 40-59 y | 0.173953 | 0.059796 | 12.1% | 1.19 [1.06, 1.34] |
| Taylor 2024, vaccinated cohort, 60-79 y | 0.223144 | 0.146779 | 7.7% | 1.25 [0.94, 1.67] |
| **Subtotal (95% CI)** | | | **100.0%** | **1.38 [1.22, 1.55]** |
| Heterogeneity: Tau² = 0.03; Chi² = 81.30, df = 9 (P < 0.00001); I² = 89% | | | | |
| Test for overall effect: Z = 5.21 (P < 0.00001) | | | | |
| **4.3.3 Diabetes, type 2 (outpatients/mixed, ≥ 65 y)** | | | | |
| Cohen 2022 | 0.672944 | 0.103435 | 18.7% | 1.96 [1.60, 2.40] |
| Taylor 2024, pre-vaccination cohort, 60-79 y | 0.252617 | 0.050422 | 24.1% | 1.29 [1.17, 1.42] |
| Taylor 2024, pre-vaccination cohort, 80-110 y | 0.240626 | 0.093384 | 19.8% | 1.27 [1.06, 1.53] |
| Taylor 2024, unvaccinated cohort, 60-79 y | 0.883768 | 0.316573 | 5.4% | 2.42 [1.30, 4.50] |
| Taylor 2024, vaccinated cohort, 60-79 y | 0.223144 | 0.084743 | 20.7% | 1.25 [1.06, 1.48] |
| Taylor 2024, vaccination cohort, 80-110 y | 0.350657 | 0.184439 | 11.4% | 1.42 [0.99, 2.04] |
| **Subtotal (95% CI)** | | | **100.0%** | **1.44 [1.23, 1.70]** |
| Heterogeneity: Tau² = 0.03; Chi² = 18.54, df = 5 (P = 0.002); I² = 73% | | | | |
| Test for overall effect: Z = 4.46 (P < 0.00001) | | | | |
| **4.3.4 Diabetes, type 2 (inpatients, 18-64 y)** | | | | |
| Daugherty 2021 | 1.299292 | 0.092114 | 100.0% | 3.67 [3.06, 4.39] |
| **Subtotal (95% CI)** | | | **100.0%** | **3.67 [3.06, 4.39]** |
| Heterogeneity: Not applicable | | | | |
| Test for overall effect: Z = 14.11 (P < 0.00001) | | | | |
| **4.3.5 Diabetes, type 2 (inpatients, ≥ 65 y)** | | | | |
| Cohen 2022 | 1.10942 | 0.055677 | 100.0% | 3.03 [2.72, 3.38] |
| **Subtotal (95% CI)** | | | **100.0%** | **3.03 [2.72, 3.38]** |
| Heterogeneity: Not applicable | | | | |
| Test for overall effect: Z = 19.93 (P < 0.00001) | | | | |

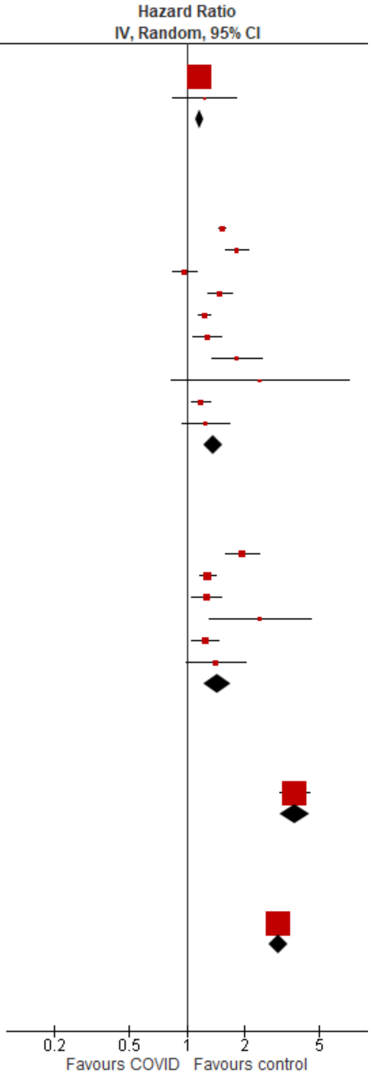

Test for subgroup differences: Chi² = 345.38, df = 4 (P < 0.00001), I² = 98.8%

#### Fibromyalgia

| Study or Subgroup | log[Hazard Ratio] | SE | Weight | Hazard Ratio IV, Random, 95% CI |
|---|---|---|---|---|
| **5.1.1 Fibromyalgia (outpatients/mixed, 18-64 y)** | | | | |
| Zang 2023, INSIGHT cohort | 0.156405 | 0.036665 | 100.0% | 1.17 [1.09, 1.26] |
| **Subtotal (95% CI)** | | | **100.0%** | **1.17 [1.09, 1.26]** |
| Heterogeneity: Not applicable | | | | |
| Test for overall effect: Z = 4.27 (P < 0.0001) | | | | |
| **5.1.2 Fibromyalgia (outpatients/mixed, ≥ 65 y)** | | | | |
| Zang 2023, INSIGHT cohort | 0.260131 | 0.046507 | 100.0% | 1.30 [1.18, 1.42] |
| **Subtotal (95% CI)** | | | **100.0%** | **1.30 [1.18, 1.42]** |
| Heterogeneity: Not applicable | | | | |
| Test for overall effect: Z = 5.59 (P < 0.00001) | | | | |

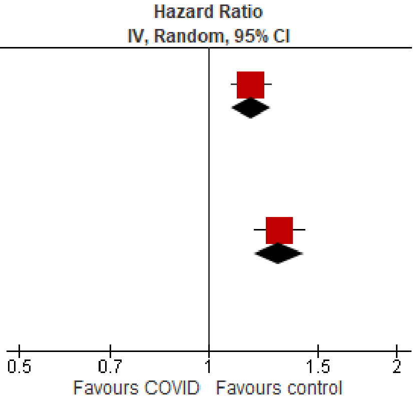

Test for subgroup differences: Chi² = 3.07, df = 1 (P = 0.08), I² = 67.4%

### Inflammatory bowel disease

#### Overall

| Study or Subgroup | log[Hazard Ratio] | SE | Weight | Hazard Ratio IV, Random, 95% CI |
|---|---|---|---|---|
| **6.1.1 Inflammatory bowel disease, overall (outpatients/mixed, <18 y)** | | | | |
| Gordon 2024, 0-1 y | -0.05873 | 0.061691 | 29.0% | 0.94 [0.84, 1.06] |
| Gordon 2024, 12-17 y | -0.06364 | 0.055569 | 35.8% | 0.94 [0.84, 1.05] |
| Gordon 2024, 2-4 y | -0.05109 | 0.088181 | 14.2% | 0.95 [0.80, 1.13] |
| Gordon 2024, 5-11 y | 0.015831 | 0.072446 | 21.0% | 1.02 [0.88, 1.17] |
| **Subtotal (95% CI)** | | | **100.0%** | **0.96 [0.90, 1.02]** |
| Heterogeneity: Tau² = 0.00; Chi² = 0.87, df = 3 (P = 0.83); I² = 0% | | | | |
| Test for overall effect: Z = 1.32 (P = 0.19) | | | | |
| **6.1.2 Inflammatory bowel disease, overall (outpatients/mixed, 18-64 y)** | | | | |
| Chang 2023, 18-40 y | 0.576613 | 0.027333 | 45.5% | 1.78 [1.69, 1.88] |
| Chang 2023, 41-64 y | 0.559616 | 0.029186 | 44.2% | 1.75 [1.65, 1.85] |
| Lim 2023 | 0.381461 | 0.155055 | 6.1% | 1.46 [1.08, 1.98] |
| Ma 2023, 60+ y | 0.667829 | 0.531157 | 0.6% | 1.95 [0.69, 5.52] |
| Ma 2023, under 60 y | 0.039221 | 0.209729 | 3.5% | 1.04 [0.69, 1.57] |
| **Subtotal (95% CI)** | | | **100.0%** | **1.71 [1.58, 1.86]** |
| Heterogeneity: Tau² = 0.00; Chi² = 7.90, df = 4 (P = 0.10); I² = 49% | | | | |
| Test for overall effect: Z = 13.25 (P < 0.00001) | | | | |
| **6.1.3 Inflammatory bowel disease, overall (outpatients/mixed, ≥ 65 y)** | | | | |
| Chang 2023 | 0.615186 | 0.037197 | 94.7% | 1.85 [1.72, 1.99] |
| Ma 2023, 60+ y | 0.667829 | 0.259178 | 1.9% | 1.95 [1.17, 3.24] |
| Wan 2023a, UK cohort | 0.336472 | 0.196754 | 3.4% | 1.40 [0.95, 2.06] |
| **Subtotal (95% CI)** | | | **100.0%** | **1.83 [1.71, 1.97]** |
| Heterogeneity: Tau² = 0.00; Chi² = 1.99, df = 2 (P = 0.37); I² = 0% | | | | |
| Test for overall effect: Z = 16.77 (P < 0.00001) | | | | |

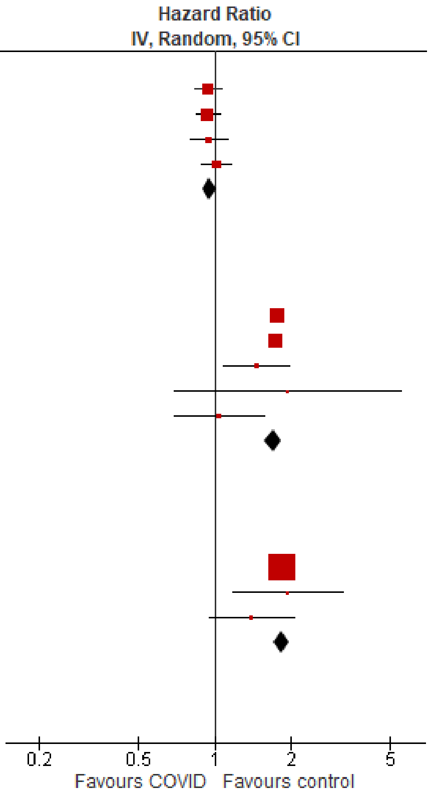

Test for subgroup differences: Chi² = 211.32, df = 2 (P < 0.00001), I² = 99.1%

#### Crohn's disease

| Study or Subgroup | log[Hazard Ratio] | SE | Weight | Hazard Ratio IV, Random, 95% CI |
|---|---|---|---|---|
| **6.2.1 Crohn's disease (outpatients/mixed, 18-64 y)** | | | | |
| Lim 2023 | 0.732368 | 0.177689 | 100.0% | 2.08 [1.47, 2.95] |
| **Subtotal (95% CI)** | | | **100.0%** | **2.08 [1.47, 2.95]** |
| Heterogeneity: Not applicable | | | | |
| Test for overall effect: Z = 4.12 (P < 0.0001) | | | | |

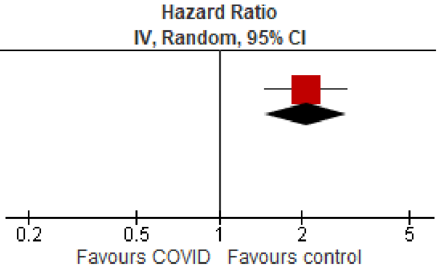

Test for subgroup differences: Not applicable

#### Ulcerative colitis

| Study or Subgroup | log[Hazard Ratio] | SE | Weight | Hazard Ratio IV, Random, 95% CI |
|---|---|---|---|---|
| **6.3.1 Ulcerative colitis (outpatients/mixed, 18-64 y)** | | | | |
| Lim 2023 | 0.019803 | 0.180391 | 100.0% | 1.02 [0.72, 1.45] |
| **Subtotal (95% CI)** | | | **100.0%** | **1.02 [0.72, 1.45]** |
| Heterogeneity: Not applicable | | | | |
| Test for overall effect: Z = 0.11 (P = 0.91) | | | | |

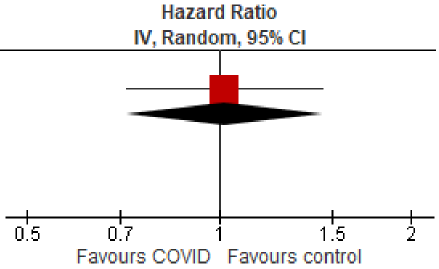

Test for subgroup differences: Not applicable

### Mental illness

#### Overall

| Study or Subgroup | log[Hazard Ratio] | SE | Weight | Hazard Ratio IV, Random, 95% CI |
|---|---|---|---|---|
| **7.1.1 Mental illness, overall (outpatients/mixed, <18 y)** | | | | |
| Donnachie 2022, 0-11 y | 0.144966 | 0.037248 | 7.2% | 1.16 [1.07, 1.24] |
| Donnachie 2022, 12-17 y | -0.23067 | 0.023763 | 7.2% | 0.79 [0.76, 0.83] |
| Gordon 2024, 0-1 y | -0.19237 | 0.234921 | 4.1% | 0.83 [0.52, 1.31] |
| Gordon 2024, 12-17 y | -0.11649 | 0.021231 | 7.3% | 0.89 [0.85, 0.93] |
| Gordon 2024, 2-4 y | 0.016261 | 0.080112 | 6.7% | 1.02 [0.87, 1.19] |
| Gordon 2024, 5-11 y | -0.03881 | 0.03265 | 7.2% | 0.96 [0.90, 1.03] |
| Kompaniyets 2022 | -0.1726 | 0.010074 | 7.3% | 0.84 [0.83, 0.86] |
| Mizrahi 2023, 12-18 y | -0.10932 | 0.200457 | 4.6% | 0.90 [0.61, 1.33] |
| Nielsen 2024 | -0.31471 | 0.027984 | 7.2% | 0.73 [0.69, 0.77] |
| Rao 2022 | 0.482426 | 0.059359 | 6.9% | 1.62 [1.44, 1.82] |
| Roessler 2022 | 0.360624 | 0.096104 | 6.4% | 1.43 [1.19, 1.73] |
| Taquet 2022 | -0.16626 | 0.039868 | 7.1% | 0.85 [0.78, 0.92] |
| Tartof 2022 | -0.1568 | 0.313754 | 3.0% | 0.85 [0.46, 1.58] |
| Valdivieso-Martinez 2023, unvaccinated cohort | -0.24484 | 0.597572 | 1.2% | 0.78 [0.24, 2.53] |
| Valdivieso-Martinez 2023, vaccinated cohort | -0.01384 | 0.427767 | 2.0% | 0.99 [0.43, 2.28] |
| Zhang-James 2024, 13-18 y | 0.04879 | 0.009719 | 7.3% | 1.05 [1.03, 1.07] |
| Zhang-James 2024, under 13 y | 0.451076 | 0.013002 | 7.3% | 1.57 [1.53, 1.61] |
| **Subtotal (95% CI)** | | | **100.0%** | **1.01 [0.88, 1.16]** |
| Heterogeneity: Tau² = 0.07; Chi² = 1879.94, df = 16 (P < 0.00001); I² = 99% | | | | |
| Test for overall effect: Z = 0.12 (P = 0.90) | | | | |
| **7.1.2 Mental illness, overall (outpatients/mixed, 18-64 y)** | | | | |
| Abel 2021, 16-24 y | 0.24686 | 0.109916 | 3.3% | 1.28 [1.03, 1.59] |
| Abel 2021, 25-34 y | 0.463734 | 0.120405 | 3.2% | 1.59 [1.26, 2.01] |
| Abel 2021, 35-49 y | 0.693147 | 0.102396 | 3.4% | 2.00 [1.64, 2.44] |
| Abel 2021, 50-59 y | 0.806476 | 0.127085 | 3.1% | 2.24 [1.75, 2.87] |
| Abel 2021, 60-69 y | 1.128171 | 0.236563 | 2.0% | 3.09 [1.94, 4.91] |
| Daugherty 2021 | 0.329304 | 0.036505 | 4.0% | 1.39 [1.29, 1.49] |
| Donnachie 2022, 18-39y | 0.325556 | 0.008473 | 4.1% | 1.38 [1.36, 1.41] |
| Donnachie 2022, 40-59y | -0.00321 | 0.009212 | 4.1% | 1.00 [0.98, 1.01] |
| Donnachie 2022, 60+ y | 0.094219 | 0.037287 | 4.0% | 1.10 [1.02, 1.18] |
| Jacob 2022, 18-30 y | -0.01855 | 0.095097 | 3.5% | 0.98 [0.81, 1.18] |
| Jacob 2022, 31-40 y | -0.05372 | 0.107087 | 3.4% | 0.95 [0.77, 1.17] |
| Jacob 2022, 41-50 y | -0.00605 | 0.103521 | 3.4% | 0.99 [0.81, 1.22] |
| Jacob 2022, 51-60 y | -0.05027 | 0.086349 | 3.6% | 0.95 [0.80, 1.13] |
| Jacob 2022, 61-70 y | 0.122487 | 0.191274 | 2.4% | 1.13 [0.78, 1.64] |
| Lam 2023 | 0.523995 | 0.307968 | 1.5% | 1.69 [0.92, 3.09] |
| Mizrahi 2023, 19-40 y | 0.028888 | 0.134651 | 3.0% | 1.03 [0.79, 1.34] |
| Mizrahi 2023, 41-60 y | 0.091665 | 0.170862 | 2.6% | 1.10 [0.78, 1.53] |
| Mizrahi 2023, 60+ y | 0.062342 | 0.503706 | 0.7% | 1.06 [0.40, 2.86] |
| Nielsen 2024, 18-39 y | -0.27444 | 0.02015 | 4.1% | 0.76 [0.73, 0.79] |
| Nielsen 2024, 40-64 y | -0.34249 | 0.028774 | 4.0% | 0.71 [0.67, 0.75] |
| Park 2021, 20-39 y | 0.470004 | 0.083952 | 3.6% | 1.60 [1.36, 1.89] |
| Park 2021, 40-59 y | 0.81978 | 0.066238 | 3.8% | 2.27 [1.99, 2.58] |
| Smith 2024, 16-30 y | 0.157004 | 0.076811 | 3.7% | 1.17 [1.01, 1.36] |
| Smith 2024, 31-40 y | 0.254642 | 0.078027 | 3.7% | 1.29 [1.11, 1.50] |
| Smith 2024, 41-50 y | 0.231112 | 0.078648 | 3.7% | 1.26 [1.08, 1.47] |
| Smith 2024, 51-60 y | 0.182322 | 0.074001 | 3.7% | 1.20 [1.04, 1.39] |
| Smith 2024, 60+ y | 0.223144 | 0.196996 | 2.4% | 1.25 [0.85, 1.84] |
| Taquet 2022 | -0.01706 | 0.008532 | 4.1% | 0.98 [0.97, 1.00] |
| Xu 2022 | 0.425268 | 0.024948 | 4.0% | 1.53 [1.46, 1.61] |
| Zang 2023, INSIGHT cohort | 0.121421 | 0.033472 | 4.0% | 1.13 [1.06, 1.21] |
| **Subtotal (95% CI)** | | | **100.0%** | **1.24 [1.13, 1.36]** |
| Heterogeneity: Tau² = 0.05; Chi² = 2064.61, df = 29 (P < 0.00001); I² = 99% | | | | |
| Test for overall effect: Z = 4.59 (P < 0.00001) | | | | |
| **7.1.3 Mental illness, overall (outpatients/mixed, ≥ 65 y)** | | | | |
| Abel 2021, 60-69 y | 1.128171 | 0.259142 | 3.1% | 3.09 [1.86, 5.14] |
| Abel 2021, 70-79 y | 1.297463 | 0.307646 | 2.5% | 3.66 [2.00, 6.69] |
| Abel 2021, 80+ y | 1.427916 | 0.228145 | 3.6% | 4.17 [2.67, 6.52] |
| Cohen 2022 | 0.565314 | 0.056278 | 7.1% | 1.76 [1.58, 1.97] |
| Donnachie 2022, 60+ y | 0.094219 | 0.014441 | 7.6% | 1.10 [1.07, 1.13] |
| Jacob 2022, 61-70 y | 0.122487 | 0.174608 | 4.6% | 1.13 [0.80, 1.59] |
| Jacob 2022, 70+ y | 0.084079 | 0.145272 | 5.2% | 1.09 [0.82, 1.45] |
| Mizrahi 2023, 60+ y | -0.31714 | 0.219835 | 3.7% | 0.73 [0.47, 1.12] |
| Murata 2024, alpha variant | -0.71335 | 0.689518 | 0.7% | 0.49 [0.13, 1.89] |
| Murata 2024, delta variant | 0.086178 | 0.339601 | 2.2% | 1.09 [0.56, 2.12] |
| Murata 2024, omicron BA.1/BA.2 variant | -0.52763 | 0.131419 | 5.5% | 0.59 [0.46, 0.76] |
| Murata 2024, omicron BA.5 variant | -0.79851 | 0.101126 | 6.2% | 0.45 [0.37, 0.55] |
| Nielsen 2024 | -0.30111 | 0.055374 | 7.2% | 0.74 [0.66, 0.82] |
| Park 2021, 60+ y | 0.620576 | 0.070584 | 6.9% | 1.86 [1.62, 2.14] |
| Smith 2024, 60+ y | 0.223144 | 0.085976 | 6.6% | 1.25 [1.06, 1.48] |
| Taquet 2022 | 0.045458 | 0.015112 | 7.6% | 1.05 [1.02, 1.08] |
| Wan 2023a, UK cohort | 0.53487 | 0.14634 | 5.2% | 1.71 [1.28, 2.27] |
| Xu 2022 | 0.270027 | 0.02524 | 7.5% | 1.31 [1.25, 1.38] |
| Zang 2023, INSIGHT cohort | 0.18938 | 0.052341 | 7.2% | 1.21 [1.09, 1.34] |
| **Subtotal (95% CI)** | | | **100.0%** | **1.20 [1.07, 1.35]** |
| Heterogeneity: Tau² = 0.05; Chi² = 418.18, df = 18 (P < 0.00001); I² = 96% | | | | |
| Test for overall effect: Z = 3.10 (P = 0.002) | | | | |

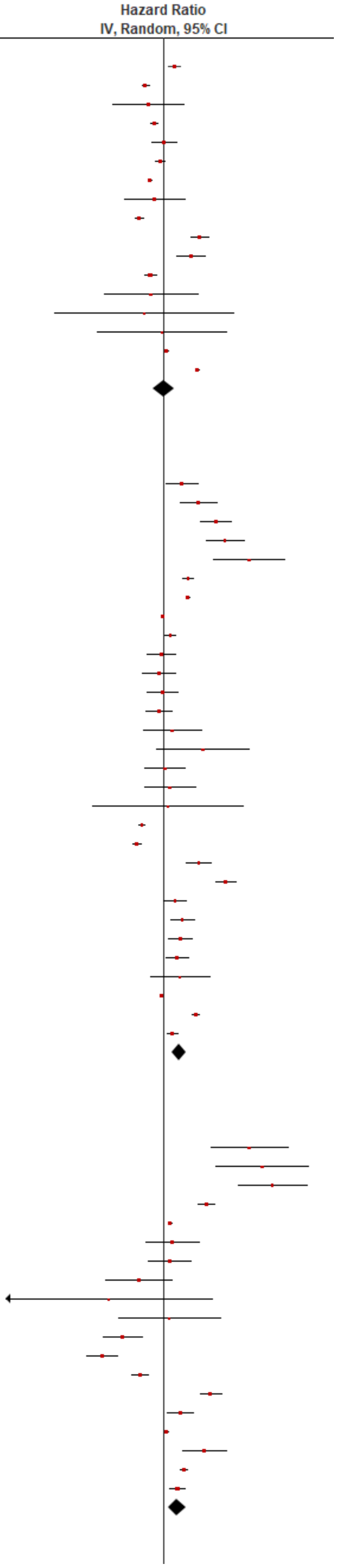

| | | | | |
|---|---|---|---|---|
| **7.1.4 Mental illness, overall (inpatients, <18 y)** | | | | |
| Zhang-James 2024, 13-18 y | 0.277632 | 0.056924 | 32.3% | 1.32 [1.18, 1.48] |
| Zhang-James 2024, under 13 y | 0.231112 | 0.039324 | 67.7% | 1.26 [1.17, 1.36] |
| **Subtotal (95% CI)** | | | **100.0%** | **1.28 [1.20, 1.36]** |
| Heterogeneity: Tau² = 0.00; Chi² = 0.45, df = 1 (P = 0.50); I² = 0% | | | | |
| Test for overall effect: Z = 7.61 (P < 0.00001) | | | | |
| **7.1.5 Mental illness, overall (inpatients, 18-64 y)** | | | | |
| Daugherty 2021 | 0.717693 | 0.054073 | 18.3% | 2.05 [1.84, 2.28] |
| Nersejan 2023, 18-29 y | 0.34359 | 0.295914 | 7.5% | 1.41 [0.79, 2.52] |
| Nersejan 2023, 30-49 y | 1.033184 | 0.333664 | 6.5% | 2.81 [1.46, 5.40] |
| Nersejan 2023, 50-69 y | -0.94161 | 0.340953 | 6.3% | 0.39 [0.20, 0.76] |
| Rahman 2024, 22-29 y | 0.596746 | 0.163051 | 13.1% | 1.82 [1.32, 2.50] |
| Rahman 2024, 30-39 y | 0.680366 | 0.131314 | 14.8% | 1.97 [1.53, 2.55] |
| Rahman 2024, 40-49 y | 0.450056 | 0.125181 | 15.1% | 1.57 [1.23, 2.00] |
| Rahman 2024, 50-59 y | 0.462223 | 0.102046 | 16.3% | 1.59 [1.30, 1.94] |
| Rahman 2024, 60+ y | 0.844623 | 0.655761 | 2.2% | 2.33 [0.64, 8.41] |
| **Subtotal (95% CI)** | | | **100.0%** | **1.66 [1.35, 2.03]** |
| Heterogeneity: Tau² = 0.06; Chi² = 31.22, df = 8 (P = 0.0001); I² = 74% | | | | |
| Test for overall effect: Z = 4.86 (P < 0.00001) | | | | |
| **7.1.6 Mental illness, overall (inpatients, ≥ 65 y)** | | | | |
| Cohen 2022 | 0.804107 | 0.041652 | 14.1% | 2.23 [2.06, 2.42] |
| Murata 2024, alpha variant | -0.08338 | 0.211931 | 12.3% | 0.92 [0.61, 1.39] |
| Murata 2024, delta variant | 0.173953 | 0.160164 | 13.1% | 1.19 [0.87, 1.63] |
| Murata 2024, omicron BA.1/BA.2 variant | -0.18633 | 0.092095 | 13.8% | 0.83 [0.69, 0.99] |
| Murata 2024, omicron BA.5 variant | -0.19845 | 0.104701 | 13.7% | 0.82 [0.67, 1.01] |
| Nersejan 2023, 50-69 y | -0.94161 | 0.590548 | 6.5% | 0.39 [0.12, 1.24] |
| Nersejan 2023, 70+ y | 0.039221 | 0.178565 | 12.8% | 1.04 [0.73, 1.48] |
| Rahman 2024, 60+ y | 0.844623 | 0.099187 | 13.7% | 2.33 [1.92, 2.83] |
| **Subtotal (95% CI)** | | | **100.0%** | **1.14 [0.76, 1.71]** |
| Heterogeneity: Tau² = 0.30; Chi² = 189.29, df = 7 (P < 0.00001); I² = 96% | | | | |
| Test for overall effect: Z = 0.64 (P = 0.52) | | | | |

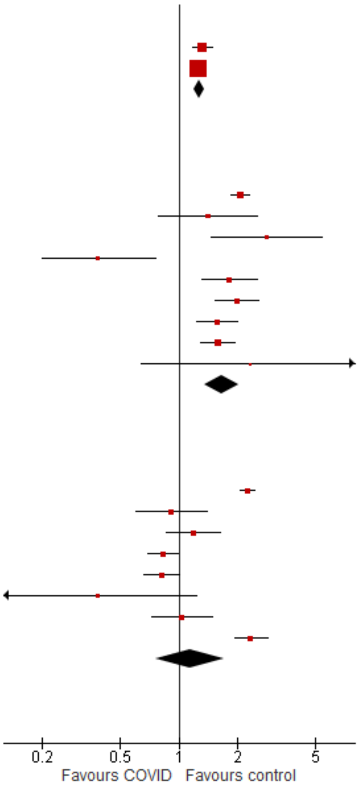

Test for subgroup differences: Chi² = 17.64, df = 5 (P = 0.003), I² = 71.7%

#### Anxiety/anxiety disorders

| Study or Subgroup | log[Hazard Ratio] | SE | Weight | Hazard Ratio IV, Random, 95% CI |
|---|---|---|---|---|
| **7.2.1 Anxiety/anxiety disorders (outpatients/mixed, <18 y)** | | | | |
| Kompaniyets 2022 | -0.16252 | 0.006003 | 36.3% | 0.85 [0.84, 0.86] |
| Mizrahi 2023, 0-4 y | -0.13926 | 0.451107 | 0.5% | 0.87 [0.36, 2.11] |
| Mizrahi 2023, 12-18 y | 0.020456 | 0.127184 | 5.8% | 1.02 [0.80, 1.31] |
| Mizrahi 2023, 5-11 y | 0.036664 | 0.151149 | 4.3% | 1.04 [0.77, 1.40] |
| Roessler 2022 | 0.409575 | 0.188676 | 2.9% | 1.51 [1.04, 2.18] |
| Taquet 2022 | -0.23383 | 0.009443 | 35.6% | 0.79 [0.78, 0.81] |
| Tartof 2022 | -0.12783 | 0.080929 | 11.7% | 0.88 [0.75, 1.03] |
| Valdivieso-Martinez 2023, unvaccinated cohort | -0.69315 | 0.499375 | 0.4% | 0.50 [0.19, 1.33] |
| Valdivieso-Martinez 2023, vaccinated cohort | 0.115113 | 0.21385 | 2.3% | 1.12 [0.74, 1.71] |
| **Subtotal (95% CI)** | | | **100.0%** | **0.87 [0.81, 0.92]** |
| Heterogeneity: Tau² = 0.00; Chi² = 58.55, df = 8 (P < 0.00001); I² = 86% | | | | |
| Test for overall effect: Z = 4.28 (P < 0.0001) | | | | |
| **7.2.2 Anxiety/anxiety disorders (outpatients/mixed, 18-64 y)** | | | | |
| Daugherty 2021 | 0.350657 | 0.037662 | 12.3% | 1.42 [1.32, 1.53] |
| Jacob 2022, 18-30 y | -0.10536 | 0.13104 | 8.1% | 0.90 [0.70, 1.16] |
| Jacob 2022, 31-40 y | -0.26136 | 0.153403 | 7.1% | 0.77 [0.57, 1.04] |
| Jacob 2022, 41-50 y | 0.067659 | 0.149946 | 7.3% | 1.07 [0.80, 1.44] |
| Jacob 2022, 51-60 y | 0.067659 | 0.121884 | 8.5% | 1.07 [0.84, 1.36] |
| Jacob 2022, 61-70 y | 0.04879 | 0.274293 | 3.6% | 1.05 [0.61, 1.80] |
| Lam 2023 | 0.29267 | 0.140606 | 7.7% | 1.34 [1.02, 1.77] |
| Mizrahi 2023, 19-40 y | 0.041275 | 0.085815 | 10.3% | 1.04 [0.88, 1.23] |
| Mizrahi 2023, 41-60 y | 0.118999 | 0.113247 | 8.9% | 1.13 [0.90, 1.41] |
| Mizrahi 2023, 60+ y | 0.22156 | 0.57783 | 1.0% | 1.25 [0.40, 3.87] |
| Taquet 2022 | -0.02922 | 0.003546 | 12.8% | 0.97 [0.96, 0.98] |
| Zang 2023, INSIGHT cohort | 0.121421 | 0.033472 | 12.4% | 1.13 [1.06, 1.21] |
| **Subtotal (95% CI)** | | | **100.0%** | **1.08 [0.96, 1.22]** |
| Heterogeneity: Tau² = 0.03; Chi² = 131.22, df = 11 (P < 0.00001); I² = 92% | | | | |
| Test for overall effect: Z = 1.29 (P = 0.20) | | | | |
| **7.2.3 Anxiety/anxiety disorders (outpatients/mixed, ≥ 65 y)** | | | | |
| Jacob 2022, 61-70 y | -0.06188 | 0.019888 | 13.4% | 0.94 [0.90, 0.98] |
| Jacob 2022, 70+ y | 0.04879 | 0.211401 | 9.8% | 1.05 [0.69, 1.59] |
| Mizrahi 2023, 60+ y | 0.22156 | 0.252186 | 8.8% | 1.25 [0.76, 2.05] |
| Murata 2024, alpha variant | 0.131028 | 0.574672 | 3.6% | 1.14 [0.37, 3.52] |
| Murata 2024, delta variant | -0.4943 | 0.572963 | 3.6% | 0.61 [0.20, 1.88] |
| Murata 2024, omicron BA.1/BA.2 variant | -1.13943 | 0.210887 | 9.8% | 0.32 [0.21, 0.48] |
| Murata 2024, omicron BA.5 variant | -1.10866 | 0.04262 | 13.3% | 0.33 [0.30, 0.36] |
| Taquet 2022 | 0.013706 | 0.008429 | 13.5% | 1.01 [1.00, 1.03] |
| Wan 2023a, UK cohort | 0.530628 | 0.16273 | 11.0% | 1.70 [1.24, 2.34] |
| Zang 2023, INSIGHT cohort | 0.18938 | 0.052341 | 13.2% | 1.21 [1.09, 1.34] |
| **Subtotal (95% CI)** | | | **100.0%** | **0.84 [0.66, 1.08]** |
| Heterogeneity: Tau² = 0.12; Chi² = 728.94, df = 9 (P < 0.00001); I² = 99% | | | | |
| Test for overall effect: Z = 1.34 (P = 0.18) | | | | |
| **7.2.4 Anxiety/anxiety disorders (inpatients, 18-64 y)** | | | | |
| Daugherty 2021 | 0.749102 | 0.062339 | 100.0% | 2.12 [1.87, 2.39] |
| **Subtotal (95% CI)** | | | **100.0%** | **2.12 [1.87, 2.39]** |
| Heterogeneity: Not applicable | | | | |
| Test for overall effect: Z = 12.02 (P < 0.00001) | | | | |
| **7.2.5 Anxiety/anxiety disorders (inpatients, ≥ 65 y)** | | | | |
| Murata 2024, alpha variant | -0.22314 | 0.440284 | 12.4% | 0.80 [0.34, 1.90] |
| Murata 2024, delta variant | -0.84397 | 0.578117 | 7.2% | 0.43 [0.14, 1.34] |
| Murata 2024, omicron BA.1/BA.2 variant | -0.71335 | 0.243753 | 40.5% | 0.49 [0.30, 0.79] |
| Murata 2024, omicron BA.5 variant | -0.8675 | 0.245258 | 40.0% | 0.42 [0.26, 0.68] |
| **Subtotal (95% CI)** | | | **100.0%** | **0.49 [0.36, 0.66]** |
| Heterogeneity: Tau² = 0.00; Chi² = 1.68, df = 3 (P = 0.64); I² = 0% | | | | |
| Test for overall effect: Z = 4.67 (P < 0.00001) | | | | |

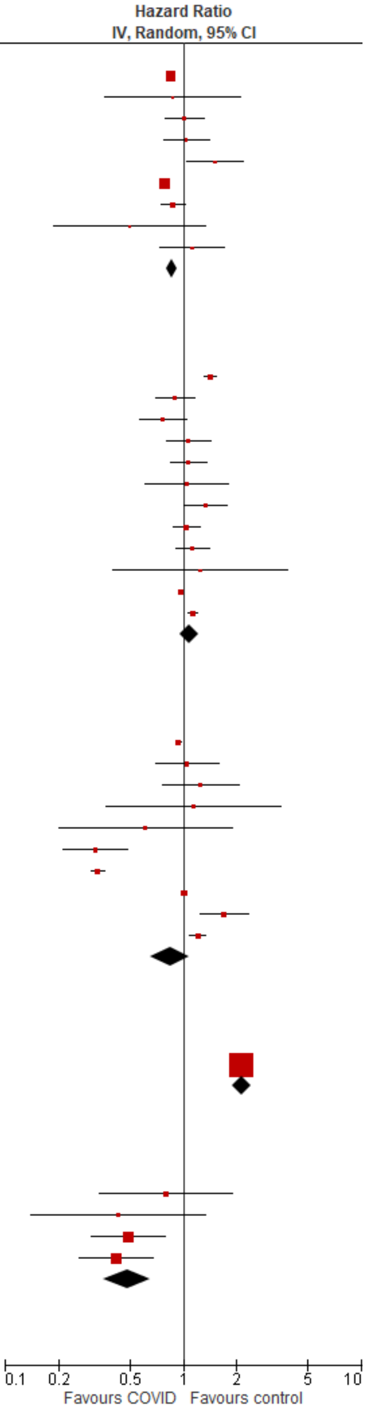

Test for subgroup differences: Chi² = 186.64, df = 4 (P < 0.00001), I² = 97.9%

#### Depression & mood disorders

| Study or Subgroup | log[Hazard Ratio] | SE | Weight | Hazard Ratio IV, Random, 95% CI |
|---|---|---|---|---|
| **7.3.1 Depression & mood disorders (outpatients/mixed, <18 y)** | | | | |
| Kompaniyets 2022 | -0.24846136 | 0.01646391 | 14.5% | 0.78 [0.76, 0.81] |
| Mizrahi 2023, 12-18 y | -0.44514 | 0.21327 | 13.1% | 0.64 [0.42, 0.97] |
| Roessler 2022 | 0.371564 | 0.130768 | 13.9% | 1.45 [1.12, 1.87] |
| Taquet 2022 | -0.05933 | 0.012802 | 14.5% | 0.94 [0.92, 0.97] |
| Valdivieso-Martinez 2023, unvaccinated cohort | 1.098612 | 0.864466 | 5.3% | 3.00 [0.55, 16.33] |
| Valdivieso-Martinez 2023, vaccinated cohort | -0.48548 | 0.448891 | 9.8% | 0.62 [0.26, 1.48] |
| Zhang-James 2024, 13-18 y | 1.435085 | 0.030047 | 14.5% | 4.20 [3.96, 4.45] |
| Zhang-James 2024, under 13 y | 1.029619 | 0.054875 | 14.4% | 2.80 [2.51, 3.12] |
| **Subtotal (95% CI)** | | | **100.0%** | **1.37 [0.84, 2.23]** |
| Heterogeneity: $Tau^2 = 0.43$; $Chi^2 = 2858.70$, df = 7 ($P < 0.00001$); $I^2 = 100\%$ | | | | |
| Test for overall effect: Z = 1.27 (P = 0.20) | | | | |
| **7.3.2 Depression & mood disorders (outpatients/mixed, 18-64 y)** | | | | |
| Daugherty 2021 | 0.239017 | 0.050181 | 10.1% | 1.27 [1.15, 1.40] |
| Jacob 2022, 18-30 y | 0.019803 | 0.087103 | 7.3% | 1.02 [0.86, 1.21] |
| Jacob 2022, 31-40 y | 0.019803 | 0.091285 | 7.0% | 1.02 [0.85, 1.22] |
| Jacob 2022, 41-50 y | -0.03046 | 0.086279 | 7.3% | 0.97 [0.82, 1.15] |
| Jacob 2022, 51-60 y | -0.06188 | 0.075739 | 8.1% | 0.94 [0.81, 1.09] |
| Jacob 2022, 61-70 y | 0.14842 | 0.162712 | 3.6% | 1.16 [0.84, 1.60] |
| Mizrahi 2023, 19-40 y | 0.062594 | 0.120899 | 5.2% | 1.06 [0.84, 1.35] |
| Mizrahi 2023, 41-60 y | 0.111062 | 0.1459 | 4.1% | 1.12 [0.84, 1.49] |
| Mizrahi 2023, 60+ y | 0.183801 | 0.592558 | 0.4% | 1.20 [0.38, 3.84] |
| Smith 2024, 16-30 y | 0.157004 | 0.076811 | 8.0% | 1.17 [1.01, 1.36] |
| Smith 2024, 31-40 y | 0.254642 | 0.078027 | 7.9% | 1.29 [1.11, 1.50] |
| Smith 2024, 41-50 y | 0.231112 | 0.078648 | 7.9% | 1.26 [1.08, 1.47] |
| Smith 2024, 51-60 y | 0.182322 | 0.074001 | 8.2% | 1.20 [1.04, 1.39] |
| Smith 2024, 60+ y | 0.223144 | 0.196996 | 2.7% | 1.25 [0.85, 1.84] |
| Taquet 2022 | 0.006976 | 0.004509 | 12.4% | 1.01 [1.00, 1.02] |
| **Subtotal (95% CI)** | | | **100.0%** | **1.11 [1.04, 1.19]** |
| Heterogeneity: $Tau^2 = 0.01$; $Chi^2 = 51.77$, df = 14 ($P < 0.00001$); $I^2 = 73\%$ | | | | |
| Test for overall effect: Z = 2.92 (P = 0.003) | | | | |
| **7.3.3 Depression & mood disorders (outpatients/mixed, ≥ 65 y)** | | | | |
| Jacob 2022, 61-70 y | 0.14842 | 0.148535 | 0.4% | 1.16 [0.87, 1.55] |
| Jacob 2022, 70+ y | 0.131028 | 0.119899 | 0.6% | 1.14 [0.90, 1.44] |
| Mizrahi 2023, 60+ y | 0.183801 | 0.258614 | 0.1% | 1.20 [0.72, 2.00] |
| Murata 2024, alpha variant | 0.81093 | 0.727984 | 0.0% | 2.25 [0.54, 9.37] |
| Murata 2024, delta variant | -0.19845 | 1.016738 | 0.0% | 0.82 [0.11, 6.02] |
| Murata 2024, omicron BA.1/BA.2 variant | 0.139762 | 0.263712 | 0.1% | 1.15 [0.69, 1.93] |
| Murata 2024, omicron BA.5 variant | -0.15082 | 0.22717 | 0.2% | 0.86 [0.55, 1.34] |
| Smith 2024, 60+ y | 0.223144 | 0.085976 | 1.2% | 1.25 [1.06, 1.48] |
| Taquet 2022 | 0.062881 | 0.009486 | 97.4% | 1.06 [1.05, 1.08] |
| **Subtotal (95% CI)** | | | **100.0%** | **1.07 [1.05, 1.09]** |
| Heterogeneity: $Tau^2 = 0.00$; $Chi^2 = 6.36$, df = 8 ($P = 0.61$); $I^2 = 0\%$ | | | | |
| Test for overall effect: Z = 7.00 ($P < 0.00001$) | | | | |
| **7.3.4 Depression & mood disorders (inpatients, 18-64 y)** | | | | |
| Daugherty 2021 | 0.818413 | 0.075001 | 100.0% | 2.27 [1.96, 2.63] |
| **Subtotal (95% CI)** | | | **100.0%** | **2.27 [1.96, 2.63]** |
| Heterogeneity: Not applicable | | | | |
| Test for overall effect: Z = 10.91 ($P < 0.00001$) | | | | |
| **7.3.5 Depression & mood disorders (inpatients, ≥ 65 y)** | | | | |
| Murata 2024, alpha variant | 0.122218 | 0.540884 | 5.7% | 1.13 [0.39, 3.26] |
| Murata 2024, delta variant | 0.285179 | 0.248211 | 27.0% | 1.33 [0.82, 2.16] |
| Murata 2024, omicron BA.1/BA.2 variant | 0.086178 | 0.21 | 37.7% | 1.09 [0.72, 1.65] |
| Murata 2024, omicron BA.5 variant | 0.131028 | 0.236566 | 29.7% | 1.14 [0.72, 1.81] |
| **Subtotal (95% CI)** | | | **100.0%** | **1.17 [0.91, 1.50]** |
| Heterogeneity: $Tau^2 = 0.00$; $Chi^2 = 0.40$, df = 3 ($P = 0.94$); $I^2 = 0\%$ | | | | |
| Test for overall effect: Z = 1.20 (P = 0.23) | | | | |

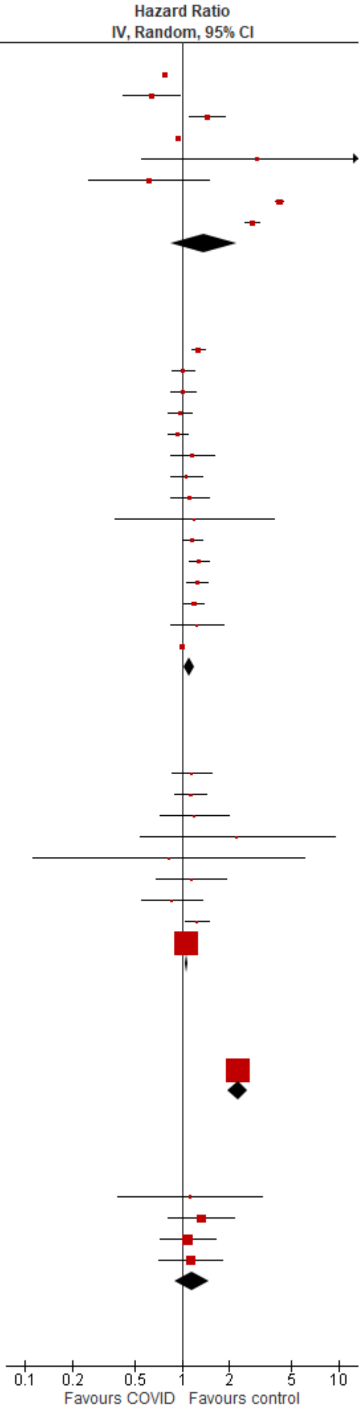

Test for subgroup differences: $Chi^2 = 101.07$, df = 4 ($P < 0.00001$), $I^2 = 96.0\%$

#### Psychosis/psychotic disorders

| Study or Subgroup | log[Hazard Ratio] | SE | Weight | Hazard Ratio IV, Random, 95% CI |
|---|---|---|---|---|
| **7.4.1 Psychosis/psychotic disorders (outpatients/mixed, <18 y)** | | | | |
| Mizrahi 2023, 12-18 y | -0.2013 | 0.379172 | 18.3% | 0.82 [0.39, 1.72] |
| Taquet 2022 | 1.048967 | 0.10743 | 25.9% | 2.85 [2.31, 3.52] |
| Tartof 2022 | -1.66073 | 0.583107 | 12.7% | 0.19 [0.06, 0.60] |
| Zhang-James 2024, 13-18 y | 0.788457 | 0.118016 | 25.7% | 2.20 [1.75, 2.77] |
| Zhang-James 2024, under 13 y | -0.10536 | 0.410571 | 17.3% | 0.90 [0.40, 2.01] |
| **Subtotal (95% CI)** | | | **100.0%** | **1.23 [0.70, 2.16]** |
| Heterogeneity: Tau² = 0.31; Chi² = 35.02, df = 4 (P < 0.00001); I² = 89% | | | | |
| Test for overall effect: Z = 0.72 (P = 0.47) | | | | |
| **7.4.2 Psychosis/psychotic disorders (outpatients/mixed, 18-64 y)** | | | | |
| Lam 2023 | 0.198851 | 0.74678 | 0.1% | 1.22 [0.28, 5.27] |
| Mizrahi 2023, 19-40 y | -0.26794 | 0.272813 | 0.6% | 0.76 [0.45, 1.31] |
| Mizrahi 2023, 41-60 y | -0.27864 | 0.348862 | 0.4% | 0.76 [0.38, 1.50] |
| Mizrahi 2023, 60+ y | -0.31714 | 0.679131 | 0.1% | 0.73 [0.19, 2.76] |
| Taquet 2022 | -0.11127 | 0.020921 | 98.9% | 0.89 [0.86, 0.93] |
| **Subtotal (95% CI)** | | | **100.0%** | **0.89 [0.86, 0.93]** |
| Heterogeneity: Tau² = 0.00; Chi² = 0.82, df = 4 (P = 0.94); I² = 0% | | | | |
| Test for overall effect: Z = 5.42 (P < 0.00001) | | | | |
| **7.4.3 Psychosis/psychotic disorders (outpatients/mixed, ≥ 65 y)** | | | | |
| Cohen 2022 | 1.007958 | 0.22021 | 18.1% | 2.74 [1.78, 4.22] |
| Mizrahi 2023, 60+ y | -0.31714 | 0.296397 | 14.9% | 0.73 [0.41, 1.30] |
| Murata 2024, alpha variant | 1.211941 | 0.820379 | 4.0% | 3.36 [0.67, 16.77] |
| Murata 2024, delta variant | 1.826161 | 0.588692 | 6.8% | 6.21 [1.96, 19.69] |
| Murata 2024, omicron BA.1/BA.2 variant | 0.34359 | 0.257242 | 16.5% | 1.41 [0.85, 2.33] |
| Murata 2024, omicron BA.5 variant | 0.139762 | 0.290515 | 15.1% | 1.15 [0.65, 2.03] |
| Taquet 2022 | 0.348118 | 0.03457 | 24.6% | 1.42 [1.32, 1.52] |
| **Subtotal (95% CI)** | | | **100.0%** | **1.60 [1.13, 2.28]** |
| Heterogeneity: Tau² = 0.13; Chi² = 21.85, df = 6 (P = 0.001); I² = 73% | | | | |
| Test for overall effect: Z = 2.62 (P = 0.009) | | | | |
| **7.4.4 Psychosis/psychotic disorders (inpatients, 18-64 y)** | | | | |
| Rahman 2024, 22-29 y | 0.596746 | 0.163051 | 14.6% | 1.82 [1.32, 2.50] |
| Rahman 2024, 30-39 y | 0.680366 | 0.131314 | 22.5% | 1.97 [1.53, 2.55] |
| Rahman 2024, 40-49 y | 0.450056 | 0.125181 | 24.8% | 1.57 [1.23, 2.00] |
| Rahman 2024, 50-59 y | 0.462223 | 0.102046 | 37.3% | 1.59 [1.30, 1.94] |
| Rahman 2024, 60+ y | 0.844623 | 0.655761 | 0.9% | 2.33 [0.64, 8.41] |
| **Subtotal (95% CI)** | | | **100.0%** | **1.70 [1.51, 1.92]** |
| Heterogeneity: Tau² = 0.00; Chi² = 2.56, df = 4 (P = 0.63); I² = 0% | | | | |
| Test for overall effect: Z = 8.53 (P < 0.00001) | | | | |
| **7.4.5 Psychosis/psychotic disorders (inpatients, ≥ 65 y)** | | | | |
| Cohen 2022 | 1.12441 | 0.167906 | 17.8% | 3.08 [2.22, 4.28] |
| Murata 2024, alpha variant | 0.65752 | 0.344369 | 12.7% | 1.93 [0.98, 3.79] |
| Murata 2024, delta variant | 0.518794 | 0.280258 | 14.6% | 1.68 [0.97, 2.91] |
| Murata 2024, omicron BA.1/BA.2 variant | 0.19062 | 0.163433 | 18.0% | 1.21 [0.88, 1.67] |
| Murata 2024, omicron BA.5 variant | -0.09431 | 0.178809 | 17.5% | 0.91 [0.64, 1.29] |
| Rahman 2024, 60+ y | 0.844623 | 0.099187 | 19.4% | 2.33 [1.92, 2.83] |
| **Subtotal (95% CI)** | | | **100.0%** | **1.72 [1.16, 2.54]** |
| Heterogeneity: Tau² = 0.20; Chi² = 37.15, df = 5 (P < 0.00001); I² = 87% | | | | |
| Test for overall effect: Z = 2.71 (P = 0.007) | | | | |

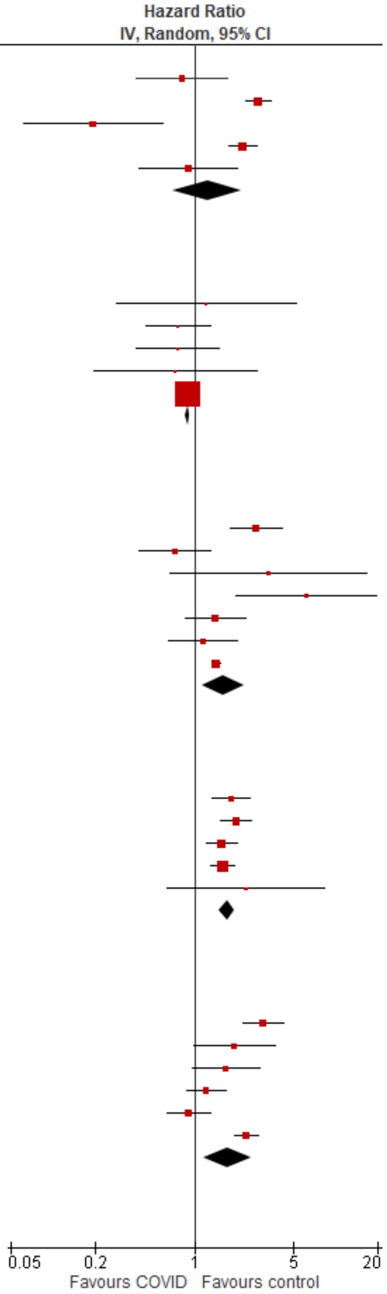

Test for subgroup differences: Chi² = 113.60, df = 4 (P < 0.00001), I² = 96.5%

#### Trauma & stress disorders

| Study or Subgroup | log[Hazard Ratio] | SE | Weight | Hazard Ratio IV, Random, 95% CI |
|---|---|---|---|---|
| **7.5.1 Trauma & stress disorders (outpatients/mixed, <18 y)** | | | | |
| Roessler 2022 | 0.536493 | 0.09491 | 94.6% | 1.71 [1.42, 2.06] |
| Valdivieso-Martinez 2023, vaccinated cohort | -0.36624 | 0.857509 | 5.4% | 0.69 [0.13, 3.72] |
| **Subtotal (95% CI)** | | | **100.0%** | **1.63 [1.09, 2.43]** |
| Heterogeneity: Tau² = 0.04; Chi² = 1.09, df = 1 (P = 0.30); I² = 9% | | | | |
| Test for overall effect: Z = 2.38 (P = 0.02) | | | | |
| **7.5.2 Trauma & stress disorders (outpatients/mixed, 18-64 y)** | | | | |
| Daugherty 2021 | 0.350657 | 0.136326 | 51.6% | 1.42 [1.09, 1.85] |
| Lam 2023 | 0.850151 | 0.162947 | 48.4% | 2.34 [1.70, 3.22] |
| **Subtotal (95% CI)** | | | **100.0%** | **1.81 [1.11, 2.95]** |
| Heterogeneity: Tau² = 0.10; Chi² = 5.53, df = 1 (P = 0.02); I² = 82% | | | | |
| Test for overall effect: Z = 2.37 (P = 0.02) | | | | |
| **7.5.3 Trauma & stress disorders (outpatients/mixed, ≥ 65 y)** | | | | |
| Wan 2023a, UK cohort | 0.542217 | 0.191374 | 100.0% | 1.72 [1.18, 2.50] |
| **Subtotal (95% CI)** | | | **100.0%** | **1.72 [1.18, 2.50]** |
| Heterogeneity: Not applicable | | | | |
| Test for overall effect: Z = 2.83 (P = 0.005) | | | | |
| **7.5.4 Trauma & stress disorders (inpatients, 18-64 y)** | | | | |
| Daugherty 2021 | 1.056053 | 0.216218 | 100.0% | 2.88 [1.88, 4.39] |
| **Subtotal (95% CI)** | | | **100.0%** | **2.88 [1.88, 4.39]** |
| Heterogeneity: Not applicable | | | | |
| Test for overall effect: Z = 4.88 (P < 0.00001) | | | | |

Test for subgroup differences: Chi² = 4.52, df = 3 (P = 0.21), I² = 33.6%

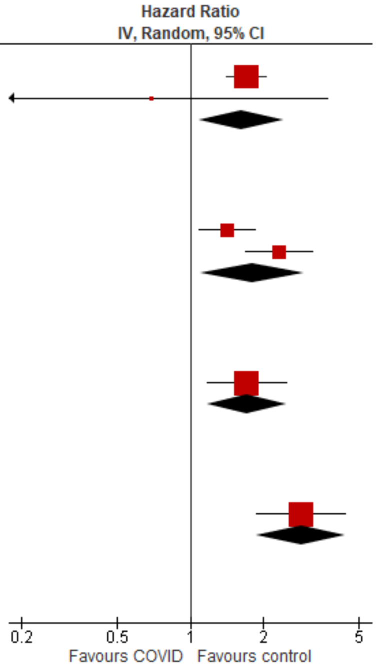

#### Musculoskeletal disorders

##### Overall

| Study or Subgroup | log[Hazard Ratio] | SE | Weight | Hazard Ratio IV, Random, 95% CI |
|---|---|---|---|---|
| **8.1.1 Musculoskeletal disorders, overall (outpatients/mixed, <18 y)** | | | | |
| Kompaniyets 2022 | -0.06188 | 0.013645 | 99.9% | 0.94 [0.92, 0.97] |
| Roessler 2022 | 0.113329 | 0.413614 | 0.1% | 1.12 [0.50, 2.52] |
| **Subtotal (95% CI)** | | | **100.0%** | **0.94 [0.92, 0.97]** |
| Heterogeneity: Tau² = 0.00; Chi² = 0.18, df = 1 (P = 0.67); I² = 0% | | | | |
| Test for overall effect: Z = 4.52 (P < 0.00001) | | | | |

Test for subgroup differences: Not applicable

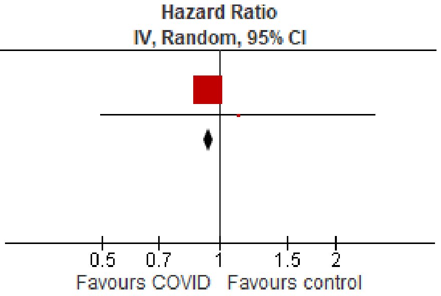

##### Inflammatory arthritides

| Study or Subgroup | log[Hazard Ratio] | SE | Weight | Hazard Ratio IV, Random, 95% CI |
|---|---|---|---|---|
| **8.2.1 Inflammatory arthritides (outpatients/mixed, >18 y)** | | | | |
| Roessler 2022 | 0.113329 | 0.413614 | 100.0% | 1.12 [0.50, 2.52] |
| **Subtotal (95% CI)** | | | **100.0%** | **1.12 [0.50, 2.52]** |
| Heterogeneity: Not applicable | | | | |
| Test for overall effect: Z = 0.27 (P = 0.78) | | | | |
| **Total (95% CI)** | | | **100.0%** | **1.12 [0.50, 2.52]** |
| Heterogeneity: Not applicable | | | | |
| Test for overall effect: Z = 0.27 (P = 0.78) | | | | |
| Test for subgroup differences: Not applicable | | | | |

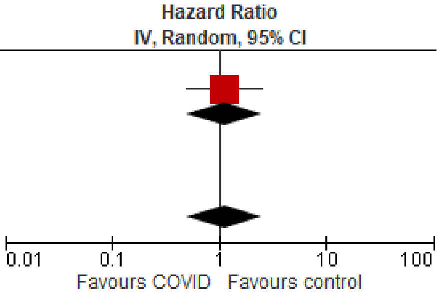

### Neurological disorders

#### Overall

| Study or Subgroup | log[Hazard Ratio] | SE | Weight | Hazard Ratio IV, Random, 95% CI |
|---|---|---|---|---|
| **9.1.1 Neurological disorders, overall (outpatients/mixed, <18 y)** | | | | |
| Donnachie 2022, 0-11 y | 1.029619 | 0.368361 | 4.4% | 2.80 [1.36, 5.76] |
| Donnachie 2022, 12-17 y | 0.897923 | 0.145685 | 10.1% | 2.45 [1.84, 3.27] |
| Gronkjaer 2023 | -0.11653 | 0.069693 | 12.4% | 0.89 [0.78, 1.02] |
| Kompaniyets 2022 | -0.06188 | 0.008186 | 13.2% | 0.94 [0.93, 0.96] |
| Mizrahi 2023, 0-4 y | 0.866777 | 0.464457 | 3.2% | 2.38 [0.96, 5.91] |
| Mizrahi 2023, 12-18 y | 0.312513 | 0.366435 | 4.5% | 1.37 [0.67, 2.80] |
| Mizrahi 2023, 5-11 y | -0.13955 | 0.325134 | 5.2% | 0.87 [0.46, 1.64] |
| Nielsen 2024 | 0.20678 | 0.32787 | 5.1% | 1.23 [0.65, 2.34] |
| Rao 2022 | 0.165514 | 0.05038 | 12.8% | 1.18 [1.07, 1.30] |
| Roessler 2022 | 0.195264 | 0.535629 | 2.5% | 1.22 [0.43, 3.47] |
| Taquet 2022 | 0.679011 | 0.11325 | 11.2% | 1.97 [1.58, 2.46] |
| Valdivieso-Martinez 2023, unvaccinated cohort | -0.28768 | 0.76335 | 1.4% | 0.75 [0.17, 3.35] |
| Valdivieso-Martinez 2023, vaccinated cohort | 0 | 0.40773 | 3.8% | 1.00 [0.45, 2.22] |
| Westman 2022 | -0.22314 | 0.143126 | 10.2% | 0.80 [0.60, 1.06] |
| **Subtotal (95% CI)** | | | **100.0%** | **1.26 [1.05, 1.52]** |
| Heterogeneity: Tau² = 0.07; Chi² = 121.03, df = 13 (P < 0.00001); I² = 89% | | | | |
| Test for overall effect: Z = 2.46 (P = 0.01) | | | | |
| **9.1.2 Neurological disorders, overall (outpatients/mixed, 18-64 y)** | | | | |
| Daugherty 2021 | 0.488405 | 0.235579 | 3.6% | 1.63 [1.03, 2.59] |
| Donnachie 2022, 18-39y | 0.761549 | 0.077881 | 5.1% | 2.14 [1.84, 2.49] |
| Donnachie 2022, 40-59y | 1.134362 | 0.051255 | 5.3% | 3.11 [2.81, 3.44] |
| Donnachie 2022, 60+ y | 0.135997 | 0.126196 | 4.7% | 1.15 [0.89, 1.47] |
| Gronkjaer 2023, 20-29 y | 0.076961 | 0.059041 | 5.3% | 1.08 [0.96, 1.21] |
| Gronkjaer 2023, 30-39 y | 0.122218 | 0.058435 | 5.3% | 1.13 [1.01, 1.27] |
| Gronkjaer 2023, 40-49 y | 0.254642 | 0.049401 | 5.3% | 1.29 [1.17, 1.42] |
| Gronkjaer 2023, 50-59 y | 0.076961 | 0.04953 | 5.3% | 1.08 [0.98, 1.19] |
| Gronkjaer 2023, 60-69 y | 0.113329 | 0.084131 | 5.1% | 1.12 [0.95, 1.32] |
| Mizrahi 2023, 19-40 y | 0.017494 | 0.227602 | 3.7% | 1.02 [0.65, 1.59] |
| Mizrahi 2023, 41-60 y | 0.174803 | 0.313754 | 2.8% | 1.19 [0.64, 2.20] |
| Mizrahi 2023, 60+ y | -0.08154 | 0.76822 | 0.8% | 0.92 [0.20, 4.15] |
| Nielsen 2024, 18-39 y | 0.100848 | 0.170984 | 4.3% | 1.11 [0.79, 1.55] |
| Nielsen 2024, 40-64 y | 0.109452 | 0.080718 | 5.1% | 1.12 [0.95, 1.31] |
| Taquet 2022 | 0.029667 | 0.027151 | 5.4% | 1.03 [0.98, 1.09] |
| Westman 2022, 21-40 y | -0.52763 | 0.124706 | 4.7% | 0.59 [0.46, 0.75] |
| Westman 2022, 41-60 y | -0.43078 | 0.113063 | 4.8% | 0.65 [0.52, 0.81] |
| Westman 2022, 61-80y | 0.24686 | 0.187368 | 4.1% | 1.28 [0.89, 1.85] |
| Xu 2022 | 0.34359 | 0.023453 | 5.4% | 1.41 [1.35, 1.48] |
| Zang 2023, INSIGHT cohort | 0.320223 | 0.081779 | 5.1% | 1.38 [1.17, 1.62] |
| Zang 2023, OneFlorida+ cohort | -0.35667 | 0.1281 | 4.7% | 0.70 [0.54, 0.90] |
| Zarifkar 2022, 18-39 y | -0.3191 | 0.324026 | 2.7% | 0.73 [0.39, 1.37] |
| Zarifkar 2022, 40-59 y | -0.26227 | 0.914212 | 0.6% | 0.77 [0.13, 4.62] |
| Zarifkar 2022, 60-79 y | 0.760617 | 0.831084 | 0.7% | 2.14 [0.42, 10.91] |
| **Subtotal (95% CI)** | | | **100.0%** | **1.16 [1.00, 1.35]** |
| Heterogeneity: Tau² = 0.11; Chi² = 559.38, df = 23 (P < 0.00001); I² = 96% | | | | |
| Test for overall effect: Z = 1.97 (P = 0.05) | | | | |
| **9.1.3 Neurological disorders, overall (outpatients/mixed, ≥ 65 y)** | | | | |
| Cohen 2022 | 0.793707 | 0.232087 | 4.6% | 2.21 [1.40, 3.49] |
| Donnachie 2022, 60+ y | 0.135997 | 0.049883 | 5.3% | 1.15 [1.04, 1.26] |
| Gollop 2023 | 0.029559 | 0.106419 | 5.1% | 1.03 [0.84, 1.27] |
| Gronkjaer 2023, 60-69 y | 0.113329 | 0.094061 | 5.2% | 1.12 [0.93, 1.35] |
| Gronkjaer 2023, 70-79 y | 0.131028 | 0.060436 | 5.3% | 1.14 [1.01, 1.28] |
| Gronkjaer 2023, 80+ y | 0.182322 | 0.070304 | 5.2% | 1.20 [1.05, 1.38] |
| Mizrahi 2023, 60+ y | 0.023683 | 0.405044 | 3.7% | 1.02 [0.46, 2.26] |
| Murata 2024, alpha variant | -0.31471 | 1.389474 | 0.9% | 0.73 [0.05, 11.12] |
| Murata 2024, delta variant | 1.811562 | 0.422699 | 3.6% | 6.12 [2.67, 14.01] |
| Murata 2024, omicron BA.1/BA.2 variant | 0.71295 | 0.200632 | 4.8% | 2.04 [1.38, 3.02] |
| Murata 2024, omicron BA.5 variant | 0.029559 | 0.183813 | 4.9% | 1.03 [0.72, 1.48] |
| Nielsen 2024 | 0.052315 | 0.148244 | 5.0% | 1.05 [0.79, 1.41] |
| Taquet 2022 | 0.091728 | 0.045347 | 5.3% | 1.10 [1.00, 1.20] |
| Wan 2023a, UK cohort | 0.09531 | 0.240453 | 4.6% | 1.10 [0.69, 1.76] |
| Wang 2022a | 0.524729 | 0.029861 | 5.3% | 1.69 [1.59, 1.79] |
| Westman 2022, 100+ y | 0.004689 | 1.997988 | 0.5% | 1.00 [0.02, 50.43] |
| Westman 2022, 61-80y | 0.24686 | 0.118502 | 5.1% | 1.28 [1.01, 1.61] |
| Westman 2022, 81-100 y | 0.29267 | 0.163323 | 4.9% | 1.34 [0.97, 1.85] |
| Xu 2022 | 0.34359 | 0.023621 | 5.3% | 1.41 [1.35, 1.48] |
| Zang 2023, INSIGHT cohort | 0.320368 | 0.078479 | 5.2% | 1.38 [1.18, 1.61] |
| Zang 2023, OneFlorida+ cohort | 2.734368 | 0.062372 | 5.2% | 15.40 [13.63, 17.40] |
| Zarifkar 2022, 60-79 y | 0.779905 | 0.582952 | 2.8% | 2.18 [0.70, 6.84] |
| Zarifkar 2022, 80+ y | 0.267583 | 0.688153 | 2.3% | 1.31 [0.34, 5.03] |
| **Subtotal (95% CI)** | | | **100.0%** | **1.55 [1.18, 2.04]** |
| Heterogeneity: Tau² = 0.37; Chi² = 1576.28, df = 22 (P < 0.00001); I² = 99% | | | | |
| Test for overall effect: Z = 3.13 (P = 0.002) | | | | |

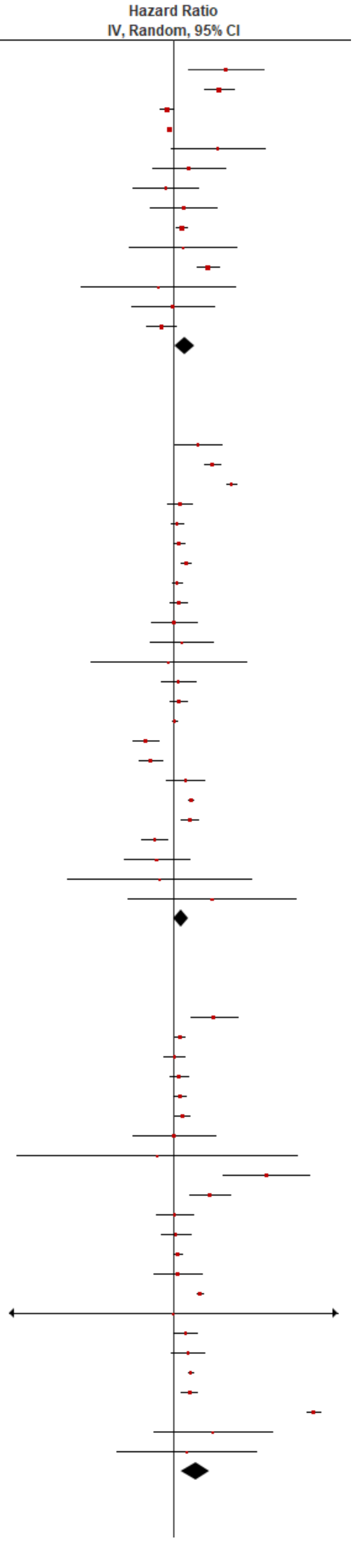

| Study or Subgroup | log[Hazard Ratio] | SE | Weight | Hazard Ratio IV, Random, 95% CI |
|---|---|---|---|---|
| **9.1.4 Neurological disorders, overall (inpatients, 18-64 y)** | | | | |
| Daugherty 2021 | 1.369809 | 0.220199 | 66.1% | 3.93 [2.56, 6.06] |
| Zarifkar 2022, 18-39 y | 0.458818 | 1.186798 | 10.3% | 1.58 [0.15, 16.20] |
| Zarifkar 2022, 40-59 y | -0.30593 | 1.011888 | 13.5% | 0.74 [0.10, 5.35] |
| Zarifkar 2022, 60-79 y | 0.180954 | 1.198991 | 10.1% | 1.20 [0.11, 12.57] |
| **Subtotal (95% CI)** | | | **100.0%** | **2.53 [1.14, 5.63]** |
| Heterogeneity: Tau² = 0.20; Chi² = 3.89, df = 3 (P = 0.27); I² = 23% | | | | |
| Test for overall effect: Z = 2.29 (P = 0.02) | | | | |
| **9.1.5 Neurological disorders, overall (inpatients, ≥ 65 y)** | | | | |
| Cohen 2022 | 1.157562 | 0.170471 | 14.8% | 3.18 [2.28, 4.44] |
| Murata 2024, alpha variant | 0.270027 | 0.244237 | 13.4% | 1.31 [0.81, 2.11] |
| Murata 2024, delta variant | -0.04082 | 0.231226 | 13.7% | 0.96 [0.61, 1.51] |
| Murata 2024, omicron BA.1/BA.2 variant | -0.10536 | 0.087269 | 15.9% | 0.90 [0.76, 1.07] |
| Murata 2024, omicron BA.5 variant | -0.21072 | 0.117434 | 15.5% | 0.81 [0.64, 1.02] |
| Qureshi 2022, 55-70 y | -0.00934 | 0.24769 | 13.3% | 0.99 [0.61, 1.61] |
| Qureshi 2022, 70+ y | 3.89182 | 1.006886 | 3.5% | 49.00 [6.81, 352.58] |
| Zarifkar 2022, 60-79 y | 0.000803 | 0.799008 | 4.9% | 1.00 [0.21, 4.79] |
| Zarifkar 2022, 80+ y | 0.190591 | 0.79266 | 5.0% | 1.21 [0.26, 5.72] |
| **Subtotal (95% CI)** | | | **100.0%** | **1.34 [0.89, 2.04]** |
| Heterogeneity: Tau² = 0.27; Chi² = 65.88, df = 8 (P < 0.00001); I² = 88% | | | | |
| Test for overall effect: Z = 1.40 (P = 0.16) | | | | |

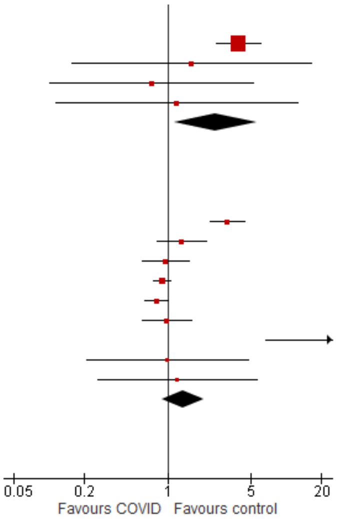

Test for subgroup differences: Chi² = 6.32, df = 4 (P = 0.18), I² = 36.7%

#### Chronic fatigue syndrome

| Study or Subgroup | log[Hazard Ratio] | SE | Weight | Hazard Ratio IV, Random, 95% CI |
|---|---|---|---|---|
| **9.2.1 Chronic fatigue syndrome (outpatients/mixed, <18 y)** | | | | |
| Donnachie 2022, 0-11 y | 1.029619 | 0.368361 | | Not estimable |
| Donnachie 2022, 12-17 y | 0.897923 | 0.145685 | | Not estimable |
| Roessler 2022 | 0.223144 | 0.845842 | 100.0% | 1.25 [0.24, 6.56] |
| **Subtotal (95% CI)** | | | **100.0%** | **1.25 [0.24, 6.56]** |
| Heterogeneity: Not applicable | | | | |
| Test for overall effect: Z = 0.26 (P = 0.79) | | | | |
| **9.2.2 Chronic fatigue syndrome (outpatients/mixed, 18-64 y)** | | | | |
| Donnachie 2022, 18-39y | 0.760479 | 0.039957 | 34.7% | 2.14 [1.98, 2.31] |
| Donnachie 2022, 40-59y | 1.17545 | 0.037273 | 34.8% | 3.24 [3.01, 3.49] |
| Donnachie 2022, 60+ y | 0.110915 | 0.139018 | 30.5% | 1.12 [0.85, 1.47] |
| **Subtotal (95% CI)** | | | **100.0%** | **2.03 [1.34, 3.06]** |
| Heterogeneity: Tau² = 0.13; Chi² = 95.51, df = 2 (P < 0.00001); I² = 98% | | | | |
| Test for overall effect: Z = 3.36 (P = 0.0008) | | | | |
| **9.2.3 Chronic fatigue syndrome (outpatients/mixed, ≥ 65 y)** | | | | |
| Donnachie 2022, 60+ y | 0.110915 | 0.054952 | 100.0% | 1.12 [1.00, 1.24] |
| **Subtotal (95% CI)** | | | **100.0%** | **1.12 [1.00, 1.24]** |
| Heterogeneity: Not applicable | | | | |
| Test for overall effect: Z = 2.02 (P = 0.04) | | | | |

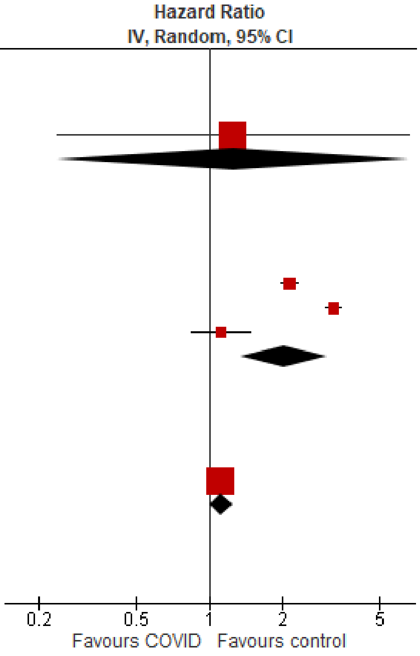

Test for subgroup differences: Chi² = 7.53, df = 2 (P = 0.02), I² = 73.5%

#### Communication & motor disorders

| Study or Subgroup | log[Hazard Ratio] | SE | Weight | Hazard Ratio IV, Random, 95% CI |
|---|---|---|---|---|
| **9.3.1 Communication & motor disorders (outpatients/mixed, <18 y)** | | | | |
| Nielsen 2024 | 0.912283 | 0.334132 | 12.2% | 2.49 [1.29, 4.79] |
| Rao 2022 | 0.165514 | 0.05038 | 32.7% | 1.18 [1.07, 1.30] |
| Roessler 2022 | 0.211468 | 0.120209 | 27.7% | 1.24 [0.98, 1.56] |
| Taquet 2022 | 0.654666 | 0.123437 | 27.4% | 1.92 [1.51, 2.45] |
| **Subtotal (95% CI)** | | | **100.0%** | **1.50 [1.12, 1.99]** |
| Heterogeneity: Tau² = 0.06; Chi² = 17.57, df = 3 (P = 0.0005); I² = 83% | | | | |
| Test for overall effect: Z = 2.76 (P = 0.006) | | | | |
| **9.3.2 Communication & motor disorders (outpatients/mixed, 18-64 y)** | | | | |
| Gronkjaer 2023, 20-39 y | 1.191618 | 0.236124 | 10.8% | 3.29 [2.07, 5.23] |
| Gronkjaer 2023, 40-59 y | 2.123265 | 0.136175 | 11.1% | 8.36 [6.40, 10.92] |
| Gronkjaer 2023, 60-79 y | 1.290414 | 0.216985 | 10.9% | 3.63 [2.38, 5.56] |
| Nielsen 2024, 18-39 y | -0.36817 | 0.323975 | 10.4% | 0.69 [0.37, 1.31] |
| Nielsen 2024, 40-64 y | -0.06253 | 0.154557 | 11.1% | 0.94 [0.69, 1.27] |
| Taquet 2022 | -1.17054 | 0.051588 | 11.3% | 0.31 [0.28, 0.34] |
| Xu 2022 | 0.364643 | 0.037139 | 11.3% | 1.44 [1.34, 1.55] |
| Zang 2023, INSIGHT cohort | -0.01228 | 0.130172 | 11.1% | 0.99 [0.77, 1.27] |
| Zarifkar 2022, 18-39 y | -0.31732 | 1.496508 | 3.8% | 0.73 [0.04, 13.68] |
| Zarifkar 2022, 40-59 y | 0.375821 | 0.911733 | 6.6% | 1.46 [0.24, 8.70] |
| Zarifkar 2022, 60-79 y | -0.03695 | 2.615193 | 1.6% | 0.96 [0.01, 162.18] |
| **Subtotal (95% CI)** | | | **100.0%** | **1.46 [0.72, 2.96]** |
| Heterogeneity: Tau² = 1.15; Chi² = 916.60, df = 10 (P < 0.00001); I² = 99% | | | | |
| Test for overall effect: Z = 1.05 (P = 0.29) | | | | |
| **9.3.3 Communication & motor disorders (outpatients/mixed, ≥ 65 y)** | | | | |
| Gronkjaer 2023, 60-79 y | 1.673351 | 0.324743 | 10.4% | 5.33 [2.82, 10.07] |
| Mizrahi 2023, 60+ y | -0.08154 | 0.441947 | 7.2% | 0.92 [0.39, 2.19] |
| Nielsen 2024 | 0.053452 | 0.149542 | 17.7% | 1.05 [0.79, 1.41] |
| Taquet 2022 | -0.08921 | 0.028377 | 21.8% | 0.91 [0.87, 0.97] |
| Xu 2022 | 0.314811 | 0.050255 | 21.4% | 1.37 [1.24, 1.51] |
| Zang 2023, INSIGHT cohort | 0.267505 | 0.200016 | 15.4% | 1.31 [0.88, 1.93] |
| Zarifkar 2022, 60-79 y | -0.03697 | 1.120579 | 1.5% | 0.96 [0.11, 8.67] |
| Zarifkar 2022, 80+ y | 0.173882 | 0.592904 | 4.6% | 1.19 [0.37, 3.80] |
| **Subtotal (95% CI)** | | | **100.0%** | **1.31 [0.99, 1.74]** |
| Heterogeneity: Tau² = 0.09; Chi² = 76.84, df = 7 (P < 0.00001); I² = 91% | | | | |
| Test for overall effect: Z = 1.90 (P = 0.06) | | | | |
| **9.3.4 Communication & motor disorders (inpatients, 18-64 y)** | | | | |
| Zarifkar 2022, 18-39 y | 1.958572 | 1.490987 | 20.0% | 7.09 [0.38, 131.74] |
| Zarifkar 2022, 40-59 y | 0.169804 | 0.825965 | 65.3% | 1.19 [0.23, 5.98] |
| Zarifkar 2022, 60-79 y | -0.52164 | 1.744669 | 14.6% | 0.59 [0.02, 18.14] |
| **Subtotal (95% CI)** | | | **100.0%** | **1.53 [0.41, 5.67]** |
| Heterogeneity: Tau² = 0.00; Chi² = 1.45, df = 2 (P = 0.48); I² = 0% | | | | |
| Test for overall effect: Z = 0.64 (P = 0.52) | | | | |
| **9.3.5 Communication & motor disorders (inpatients, ≥ 65 y)** | | | | |
| Zarifkar 2022, 60-79 y | -0.52164 | 1.042639 | 28.1% | 0.59 [0.08, 4.58] |
| Zarifkar 2022, 80+ y | -0.3509 | 0.651637 | 71.9% | 0.70 [0.20, 2.53] |
| **Subtotal (95% CI)** | | | **100.0%** | **0.67 [0.23, 1.98]** |
| Heterogeneity: Tau² = 0.00; Chi² = 0.02, df = 1 (P = 0.89); I² = 0% | | | | |
| Test for overall effect: Z = 0.72 (P = 0.47) | | | | |

Test for subgroup differences: Chi² = 2.18, df = 4 (P = 0.70), I² = 0%

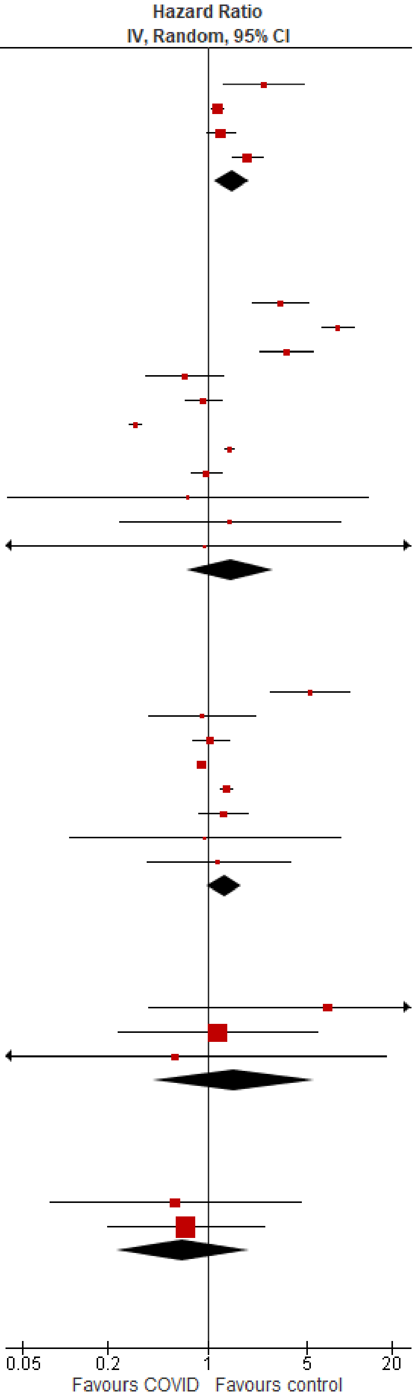

#### Dementia/mild cognitive impairment

| Study or Subgroup | log[Hazard Ratio] | SE | Weight | Hazard Ratio IV, Random, 95% CI |
|---|---|---|---|---|
| **9.4.1 Dementia/mild cognitive impairment (outpatients/mixed, 18-64 y)** | | | | |
| Daugherty 2021 | 1.019158 | 0.540074 | 4.7% | 2.77 [0.96, 7.99] |
| Donnachie 2022, 18-39y | 0.773205 | 0.131887 | 10.2% | 2.17 [1.67, 2.81] |
| Donnachie 2022, 40-59y | 0.951388 | 0.078656 | 10.7% | 2.59 [2.22, 3.02] |
| Donnachie 2022, 60+ y | 0.166107 | 0.152318 | 10.0% | 1.18 [0.88, 1.59] |
| Nielsen 2024, 18-39 y | 0.24686 | 0.486549 | 5.3% | 1.28 [0.49, 3.32] |
| Nielsen 2024, 40-64 y | -0.41552 | 0.130313 | 10.2% | 0.66 [0.51, 0.85] |
| Taquet 2022 | -0.09519 | 0.033674 | 11.0% | 0.91 [0.85, 0.97] |
| Xu 2022 | 0.565314 | 0.030388 | 11.0% | 1.76 [1.66, 1.87] |
| Zang 2023, INSIGHT cohort | 0.435024 | 0.122723 | 10.3% | 1.55 [1.21, 1.97] |
| Zang 2023, OneFlorida+ cohort | -0.35667 | 0.1281 | 10.3% | 0.70 [0.54, 0.90] |
| Zarifkar 2022, 18-39 y | 3.049951 | 1.999941 | 0.6% | 21.11 [0.42, 1063.97] |
| Zarifkar 2022, 40-59 y | 1.844352 | 1.632594 | 0.8% | 6.32 [0.26, 155.12] |
| Zarifkar 2022, 60-79 y | 0.832909 | 0.511689 | 5.0% | 2.30 [0.84, 6.27] |
| **Subtotal (95% CI)** | | | **100.0%** | **1.42 [1.05, 1.92]** |
| Heterogeneity: Tau² = 0.22; Chi² = 359.48, df = 12 (P < 0.00001); I² = 97% | | | | |
| Test for overall effect: Z = 2.26 (P = 0.02) | | | | |
| **9.4.2 Dementia/mild cognitive impairment (outpatients/mixed, ≥ 65 y)** | | | | |
| Cohen 2022 | 0.81978 | 0.081838 | 7.8% | 2.27 [1.93, 2.66] |
| Donnachie 2022, 60+ y | 0.166107 | 0.060209 | 8.2% | 1.18 [1.05, 1.33] |
| Gollop 2023 | 0.029559 | 0.106419 | 7.3% | 1.03 [0.84, 1.27] |
| Murata 2024, alpha variant | -0.31471 | 1.389474 | 0.3% | 0.73 [0.05, 11.12] |
| Murata 2024, delta variant | 1.811562 | 0.422699 | 2.2% | 6.12 [2.67, 14.01] |
| Murata 2024, omicron BA.1/BA.2 variant | 0.71295 | 0.200632 | 5.3% | 2.04 [1.38, 3.02] |
| Murata 2024, omicron BA.5 variant | 0.029559 | 0.183813 | 5.6% | 1.03 [0.72, 1.48] |
| Nielsen 2024 | -0.31471 | 0.049074 | 8.4% | 0.73 [0.66, 0.80] |
| Taquet 2022 | 0.289231 | 0.014765 | 8.7% | 1.34 [1.30, 1.37] |
| Wan 2023a, UK cohort | 0.09531 | 0.240453 | 4.5% | 1.10 [0.69, 1.76] |
| Wang 2022a | 0.524729 | 0.029861 | 8.6% | 1.69 [1.59, 1.79] |
| Xu 2022 | 0.512824 | 0.048739 | 8.4% | 1.67 [1.52, 1.84] |
| Zang 2023, INSIGHT cohort | 0.360747 | 0.053668 | 8.3% | 1.43 [1.29, 1.59] |
| Zang 2023, OneFlorida+ cohort | 0.352838 | 0.065038 | 8.1% | 1.42 [1.25, 1.62] |
| Zarifkar 2022, 60-79 y | 0.832909 | 0.334979 | 3.1% | 2.30 [1.19, 4.43] |
| Zarifkar 2022, 80+ y | 0.262364 | 0.203701 | 5.2% | 1.30 [0.87, 1.94] |
| **Subtotal (95% CI)** | | | **100.0%** | **1.42 [1.23, 1.63]** |
| Heterogeneity: Tau² = 0.06; Chi² = 308.26, df = 15 (P < 0.00001); I² = 95% | | | | |
| Test for overall effect: Z = 4.76 (P < 0.00001) | | | | |
| **9.4.3 Dementia/mild cognitive impairment (inpatients, 18-64 y)** | | | | |
| Daugherty 2021 | 2.261763 | 0.169586 | 23.6% | 9.60 [6.89, 13.39] |
| Qureshi 2022, 18-35y | -1.09691 | 1.6251 | 7.5% | 0.33 [0.01, 8.07] |
| Qureshi 2022, 35-54 y | -0.0005 | 0.391276 | 21.4% | 1.00 [0.46, 2.15] |
| Qureshi 2022, 55-70 y | -0.00934 | 0.201401 | 23.4% | 0.99 [0.67, 1.47] |
| Zarifkar 2022, 60-79 y | 0.182322 | 0.062958 | 24.1% | 1.20 [1.06, 1.36] |
| **Subtotal (95% CI)** | | | **100.0%** | **1.64 [0.57, 4.68]** |
| Heterogeneity: Tau² = 1.19; Chi² = 138.47, df = 4 (P < 0.00001); I² = 97% | | | | |
| Test for overall effect: Z = 0.92 (P = 0.36) | | | | |
| **9.4.4 Dementia/mild cognitive impairment (inpatients, ≥ 65 y)** | | | | |
| Cohen 2022 | 1.07432 | 0.059607 | 13.2% | 2.93 [2.61, 3.29] |
| Murata 2024, alpha variant | 0.270027 | 0.244237 | 12.0% | 1.31 [0.81, 2.11] |
| Murata 2024, delta variant | -0.04082 | 0.231226 | 12.1% | 0.96 [0.61, 1.51] |
| Murata 2024, omicron BA.1/BA.2 variant | -0.10536 | 0.087269 | 13.1% | 0.90 [0.76, 1.07] |
| Murata 2024, omicron BA.5 variant | -0.21072 | 0.117434 | 13.0% | 0.81 [0.64, 1.02] |
| Qureshi 2022, 55-70 y | -0.00934 | 0.24769 | 11.9% | 0.99 [0.61, 1.61] |
| Qureshi 2022, 70+ y | 3.89182 | 1.006886 | 4.5% | 49.00 [6.81, 352.58] |
| Zarifkar 2022, 60-79 y | 0.182322 | 0.53599 | 8.6% | 1.20 [0.42, 3.43] |
| Zarifkar 2022, 80+ y | 0.262364 | 0.280258 | 11.6% | 1.30 [0.75, 2.25] |
| **Subtotal (95% CI)** | | | **100.0%** | **1.42 [0.85, 2.38]** |
| Heterogeneity: Tau² = 0.52; Chi² = 200.48, df = 8 (P < 0.00001); I² = 96% | | | | |
| Test for overall effect: Z = 1.33 (P = 0.19) | | | | |

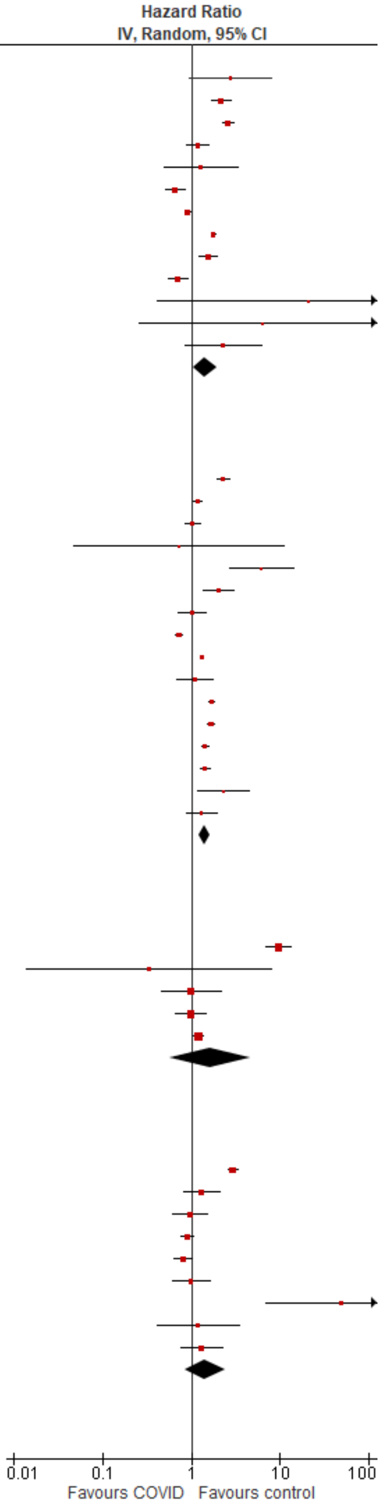

Test for subgroup differences: Chi² = 0.07, df = 3 (P = 0.99), I² = 0%

#### Encephalopathy

| Study or Subgroup | log[Hazard Ratio] | SE | Weight | Hazard Ratio IV, Random, 95% CI |
|---|---|---|---|---|
| **9.5.1 Encephalopathy (outpatients/mixed, 18-64 y)** | | | | |
| Daugherty 2021 | 1.83418 | 0.226278 | 48.9% | 6.26 [4.02, 9.75] |
| Zang 2023, INSIGHT cohort | 0.368386 | 0.052429 | 51.1% | 1.45 [1.30, 1.60] |
| **Subtotal (95% CI)** | | | **100.0%** | **2.96 [0.70, 12.44]** |
| Heterogeneity: Tau² = 1.05; Chi² = 39.82, df = 1 (P < 0.00001); I² = 97% | | | | |
| Test for overall effect: Z = 1.48 (P = 0.14) | | | | |
| **9.5.2 Encephalopathy (outpatients/mixed, ≥ 65 y)** | | | | |
| Cohen 2022 | 1.211941 | 0.080186 | 49.7% | 3.36 [2.87, 3.93] |
| Zang 2023, INSIGHT cohort | 0.371839 | 0.053515 | 50.3% | 1.45 [1.31, 1.61] |
| **Subtotal (95% CI)** | | | **100.0%** | **2.20 [0.97, 5.02]** |
| Heterogeneity: Tau² = 0.35; Chi² = 75.94, df = 1 (P < 0.00001); I² = 99% | | | | |
| Test for overall effect: Z = 1.88 (P = 0.06) | | | | |
| **9.5.3 Encephalopathy (inpatients, 18-64 y)** | | | | |
| Daugherty 2021 | 3.183994 | 0.28532 | 100.0% | 24.14 [13.80, 42.23] |
| **Subtotal (95% CI)** | | | **100.0%** | **24.14 [13.80, 42.23]** |
| Heterogeneity: Not applicable | | | | |
| Test for overall effect: Z = 11.16 (P < 0.00001) | | | | |
| **9.5.4 Encephalopathy (inpatients, ≥ 65 y)** | | | | |
| Cohen 2022 | 1.765261 | 0.066469 | 100.0% | 5.84 [5.13, 6.66] |
| **Subtotal (95% CI)** | | | **100.0%** | **5.84 [5.13, 6.66]** |
| Heterogeneity: Not applicable | | | | |
| Test for overall effect: Z = 26.56 (P < 0.00001) | | | | |

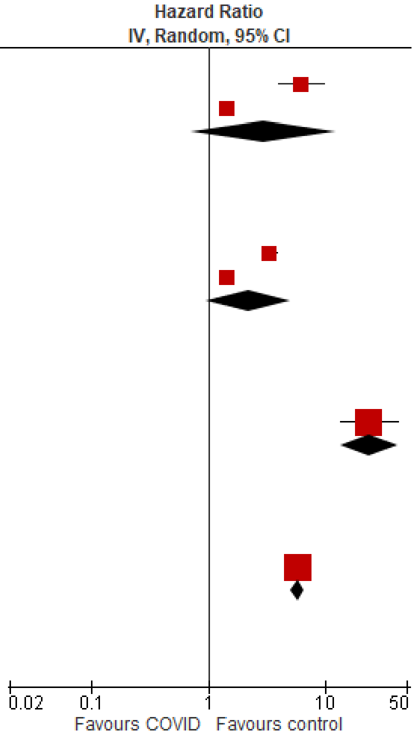

Test for subgroup differences: Chi² = 30.52, df = 3 (P < 0.00001), I² = 90.2%

#### Epilepsy

| Study or Subgroup | log[Hazard Ratio] | SE | Weight | Hazard Ratio IV, Random, 95% CI |
|---|---|---|---|---|
| **9.6.1 Epilepsy (outpatients/mixed, <18 y)** | | | | |
| Gronkjaer 2023 | -0.28768 | 0.147913 | 18.9% | 0.75 [0.56, 1.00] |
| Mizrahi 2023, 0-4 y | 0.866777 | 0.464457 | 13.4% | 2.38 [0.96, 5.91] |
| Mizrahi 2023, 12-18 y | -0.22314 | 0.50477 | 12.6% | 0.80 [0.30, 2.15] |
| Mizrahi 2023, 5-11 y | -0.33165 | 0.313322 | 16.3% | 0.72 [0.39, 1.33] |
| Taquet 2022 | 0.736015 | 0.025306 | 19.8% | 2.09 [1.99, 2.19] |
| Westman 2022 | -0.22314 | 0.143126 | 19.0% | 0.80 [0.60, 1.06] |
| **Subtotal (95% CI)** | | | **100.0%** | **1.09 [0.61, 1.95]** |
| Heterogeneity: Tau² = 0.45; Chi² = 101.19, df = 5 (P < 0.00001); I² = 95% | | | | |
| Test for overall effect: Z = 0.28 (P = 0.78) | | | | |
| **9.6.2 Epilepsy (outpatients/mixed, 18-64 y)** | | | | |
| Daugherty 2021 | 0.5988365 | 0.178809 | 7.0% | 1.82 [1.28, 2.58] |
| Gronkjaer 2023, 20-39 y | -0.19845 | 0.207918 | 5.6% | 0.82 [0.55, 1.23] |
| Gronkjaer 2023, 40-59 y | -0.16252 | 0.191771 | 6.3% | 0.85 [0.58, 1.24] |
| Gronkjaer 2023, 60-79 y | 0.058269 | 0.312814 | 2.8% | 1.06 [0.57, 1.96] |
| Mizrahi 2023, 19-40 y | 0.417722 | 0.389851 | 1.9% | 1.52 [0.71, 3.26] |
| Mizrahi 2023, 41-60 y | -0.3285 | 0.540475 | 1.0% | 0.72 [0.25, 2.08] |
| Mizrahi 2023, 60+ y | -0.60666 | 1.260595 | 0.2% | 0.55 [0.05, 6.45] |
| Taquet 2022 | 0.263517 | 0.012367 | 25.7% | 1.30 [1.27, 1.33] |
| Westman 2022, 21-40 y | -0.52763274 | 0.124706 | 11.2% | 0.59 [0.46, 0.75] |
| Westman 2022, 41-60 y | -0.43078292 | 0.113063 | 12.5% | 0.65 [0.52, 0.81] |
| Westman 2022, 61-80y | 0.263517 | 0.01223 | 25.7% | 1.30 [1.27, 1.33] |
| **Subtotal (95% CI)** | | | **100.0%** | **1.05 [0.94, 1.17]** |
| Heterogeneity: Tau² = 0.01; Chi² = 92.53, df = 10 (P < 0.00001); I² = 89% | | | | |
| Test for overall effect: Z = 0.86 (P = 0.39) | | | | |
| **9.6.3 Epilepsy (outpatients/mixed, ≥ 65 y)** | | | | |
| Cohen 2022 | 0.774727 | 0.159395 | 15.3% | 2.17 [1.59, 2.97] |
| Gronkjaer 2023, 60-79 y | 0.058269 | 0.186942 | 13.3% | 1.06 [0.73, 1.53] |
| Gronkjaer 2023, 80+ y | 0.09531 | 0.245479 | 9.9% | 1.10 [0.68, 1.78] |
| Mizrahi 2023, 60+ y | -0.60666 | 0.550169 | 2.9% | 0.55 [0.19, 1.60] |
| Taquet 2022 | 0.10084 | 0.020774 | 24.8% | 1.11 [1.06, 1.15] |
| Westman 2022, 100+ y | 0.004689 | 1.997988 | 0.2% | 1.00 [0.02, 50.43] |
| Westman 2022, 61-80y | 0.24686 | 0.118502 | 18.5% | 1.28 [1.01, 1.61] |
| Westman 2022, 81-100 y | 0.29267 | 0.163323 | 15.0% | 1.34 [0.97, 1.85] |
| **Subtotal (95% CI)** | | | **100.0%** | **1.26 [1.04, 1.53]** |
| Heterogeneity: Tau² = 0.04; Chi² = 21.83, df = 7 (P = 0.003); I² = 68% | | | | |
| Test for overall effect: Z = 2.33 (P = 0.02) | | | | |
| **9.6.4 Epilepsy (inpatients, 18-64 y)** | | | | |
| Daugherty 2021 | 1.767655 | 0.250715 | 100.0% | 5.86 [3.58, 9.57] |
| **Subtotal (95% CI)** | | | **100.0%** | **5.86 [3.58, 9.57]** |
| Heterogeneity: Not applicable | | | | |
| Test for overall effect: Z = 7.05 (P < 0.00001) | | | | |
| **9.6.5 Epilepsy (inpatients, ≥ 65 y)** | | | | |
| Cohen 2022 | 1.349916 | 0.155764 | 100.0% | 3.86 [2.84, 5.23] |
| **Subtotal (95% CI)** | | | **100.0%** | **3.86 [2.84, 5.23]** |
| Heterogeneity: Not applicable | | | | |
| Test for overall effect: Z = 8.67 (P < 0.00001) | | | | |

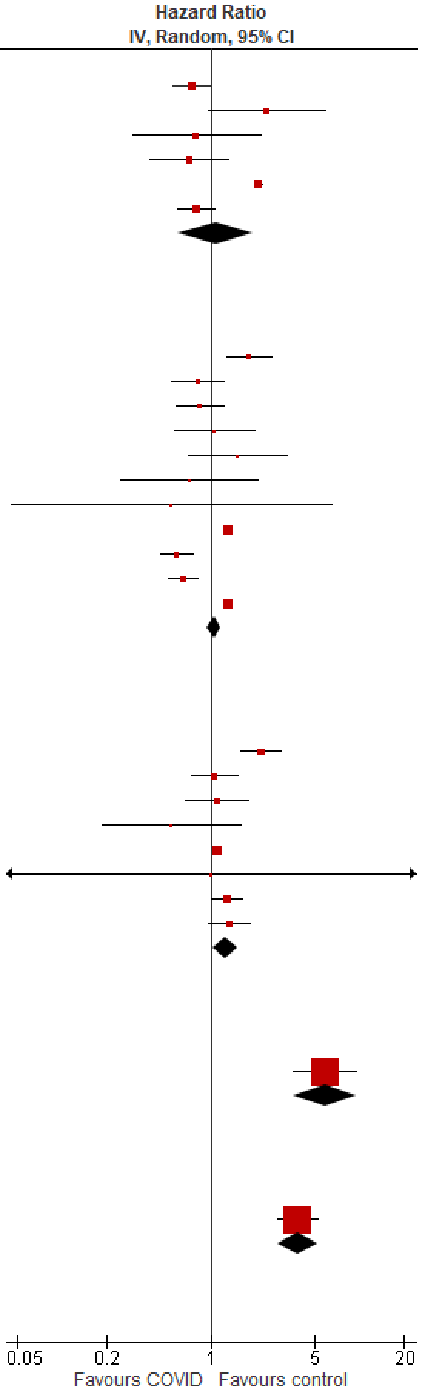

Test for subgroup differences: Chi² = 100.12, df = 4 (P < 0.00001), I² = 96.0%

#### Guillain-Barre syndrome

| Study or Subgroup | log[Hazard Ratio] | SE | Weight | Hazard Ratio IV, Random, 95% CI |
|---|---|---|---|---|
| **9.7.1 Guillian-Barre syndrome (outpatients/mixed, <18 y)** | | | | |
| Taquet 2022 | -0.44177 | 0.3264 | 100.0% | 0.64 [0.34, 1.22] |
| **Subtotal (95% CI)** | | | **100.0%** | **0.64 [0.34, 1.22]** |
| Heterogeneity: Not applicable | | | | |
| Test for overall effect: Z = 1.35 (P = 0.18) | | | | |
| **9.7.2 Guillian-Barre syndrome (outpatients/mixed, 18-64 y)** | | | | |
| Daugherty 2021 | 1.211941 | 0.78529 | 0.7% | 3.36 [0.72, 15.66] |
| Mizrahi 2023, 41-60 y | 0.076961 | 0.313754 | 4.1% | 1.08 [0.58, 2.00] |
| Taquet 2022 | 0.161523 | 0.065576 | 94.4% | 1.18 [1.03, 1.34] |
| Zarifkar 2022, 18-39 y | -0.31732 | 1.00442 | 0.4% | 0.73 [0.10, 5.21] |
| Zarifkar 2022, 40-59 y | -0.72052 | 1.435453 | 0.2% | 0.49 [0.03, 8.11] |
| Zarifkar 2022, 60-79 y | 0.503741 | 1.336515 | 0.2% | 1.65 [0.12, 22.72] |
| **Subtotal (95% CI)** | | | **100.0%** | **1.18 [1.04, 1.33]** |
| Heterogeneity: Tau² = 0.00; Chi² = 2.53, df = 5 (P = 0.77); I² = 0% | | | | |
| Test for overall effect: Z = 2.54 (P = 0.01) | | | | |
| **9.7.3 Guillian-Barre syndrome (outpatients/mixed, ≥ 65 y)** | | | | |
| Cohen 2022 | 1.669592 | 1.057887 | 0.5% | 5.31 [0.67, 42.22] |
| Taquet 2022 | 0.841912 | 0.077164 | 98.5% | 2.32 [2.00, 2.70] |
| Zarifkar 2022, 60-79 y | 0.503741 | 0.874722 | 0.8% | 1.65 [0.30, 9.19] |
| Zarifkar 2022, 80+ y | 0.65269 | 1.536664 | 0.2% | 1.92 [0.09, 39.04] |
| **Subtotal (95% CI)** | | | **100.0%** | **2.32 [2.00, 2.70]** |
| Heterogeneity: Tau² = 0.00; Chi² = 0.78, df = 3 (P = 0.86); I² = 0% | | | | |
| Test for overall effect: Z = 11.01 (P < 0.00001) | | | | |
| **9.7.4 Guillian-Barre syndrome (inpatients, 18-64 y)** | | | | |
| Daugherty 2021 | 2.397895 | 0.685224 | 48.5% | 11.00 [2.87, 42.14] |
| Zarifkar 2022, 18-39 y | 0.859974 | 1.440847 | 20.1% | 2.36 [0.14, 39.80] |
| Zarifkar 2022, 40-59 y | -0.40437 | 1.442761 | 20.0% | 0.67 [0.04, 11.28] |
| Zarifkar 2022, 60-79 y | -0.05646 | 2.038969 | 11.4% | 0.95 [0.02, 51.41] |
| **Subtotal (95% CI)** | | | **100.0%** | **3.48 [0.81, 14.91]** |
| Heterogeneity: Tau² = 0.67; Chi² = 4.21, df = 3 (P = 0.24); I² = 29% | | | | |
| Test for overall effect: Z = 1.68 (P = 0.09) | | | | |
| **9.7.5 Guillian-Barre syndrome (inpatients, ≥ 65 y)** | | | | |
| Cohen 2022 | 1.252763 | 0.659858 | 67.4% | 3.50 [0.96, 12.76] |
| Zarifkar 2022, 60-79 y | -0.05646 | 1.217804 | 19.8% | 0.95 [0.09, 10.28] |
| Zarifkar 2022, 80+ y | 1.048371 | 1.511702 | 12.8% | 2.85 [0.15, 55.22] |
| **Subtotal (95% CI)** | | | **100.0%** | **2.63 [0.91, 7.61]** |
| Heterogeneity: Tau² = 0.00; Chi² = 0.90, df = 2 (P = 0.64); I² = 0% | | | | |
| Test for overall effect: Z = 1.79 (P = 0.07) | | | | |

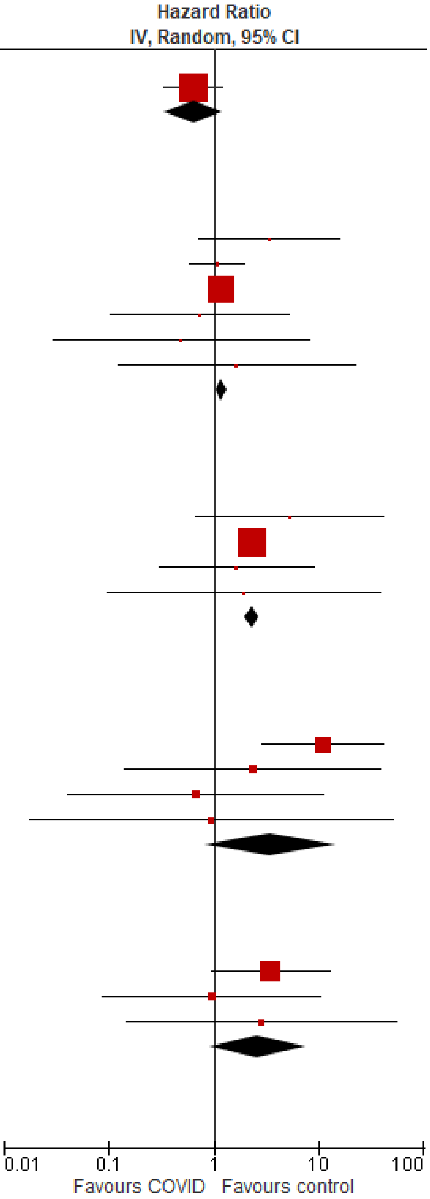

Test for subgroup differences: Chi² = 56.14, df = 4 (P < 0.00001), I² = 92.9%

#### Migraine & headache disorders

| Study or Subgroup | log[Hazard Ratio] | SE | Weight | Hazard Ratio IV, Random, 95% CI |
|---|---|---|---|---|
| **9.8.1 Migraine/headache disorders (outpatients/mixed, <18 y)** | | | | |
| Valdivieso-Martinez 2023, unvaccinated cohort | -0.28768 | 0.943196 | 18.8% | 0.75 [0.12, 4.76] |
| Valdivieso-Martinez 2023, vaccinated cohort | 0 | 0.453779 | 81.2% | 1.00 [0.41, 2.43] |
| **Subtotal (95% CI)** | | | **100.0%** | **0.95 [0.43, 2.11]** |
| Heterogeneity: Tau² = 0.00; Chi² = 0.08, df = 1 (P = 0.78); I² = 0% | | | | |
| Test for overall effect: Z = 0.13 (P = 0.89) | | | | |
| **9.8.2 Migraine/headache disorders (outpatients/mixed, 18-64 y)** | | | | |
| Daugherty 2021 | 0.254642 | 0.0711 | 100.0% | 1.29 [1.12, 1.48] |
| **Subtotal (95% CI)** | | | **100.0%** | **1.29 [1.12, 1.48]** |
| Heterogeneity: Not applicable | | | | |
| Test for overall effect: Z = 3.58 (P = 0.0003) | | | | |
| **9.8.3 Migraine/headache disorders (outpatients/mixed, ≥ 65 y)** | | | | |
| Cohen 2022 | 0.23111172 | 0.104259 | 100.0% | 1.26 [1.03, 1.55] |
| **Subtotal (95% CI)** | | | **100.0%** | **1.26 [1.03, 1.55]** |
| Heterogeneity: Not applicable | | | | |
| Test for overall effect: Z = 2.22 (P = 0.03) | | | | |
| **9.8.4 Migraine/headache disorders (inpatients, 18-64 y)** | | | | |
| Daugherty 2021 | 0.439802 | 0.129186 | 100.0% | 1.55 [1.21, 2.00] |
| **Subtotal (95% CI)** | | | **100.0%** | **1.55 [1.21, 2.00]** |
| Heterogeneity: Not applicable | | | | |
| Test for overall effect: Z = 3.40 (P = 0.0007) | | | | |
| **9.8.5 Migraine/headache disorders (inpatients, ≥ 65 y)** | | | | |
| Cohen 2022 | -0.44738 | 0.215981 | 100.0% | 0.64 [0.42, 0.98] |
| **Subtotal (95% CI)** | | | **100.0%** | **0.64 [0.42, 0.98]** |
| Heterogeneity: Not applicable | | | | |
| Test for overall effect: Z = 2.07 (P = 0.04) | | | | |
| Test for subgroup differences: Chi² = 13.07, df = 4 (P = 0.01), I² = 69.4% | | | | |

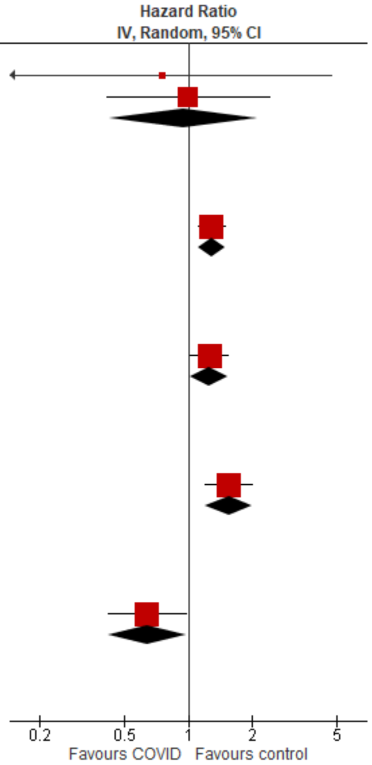

#### Multiple sclerosis

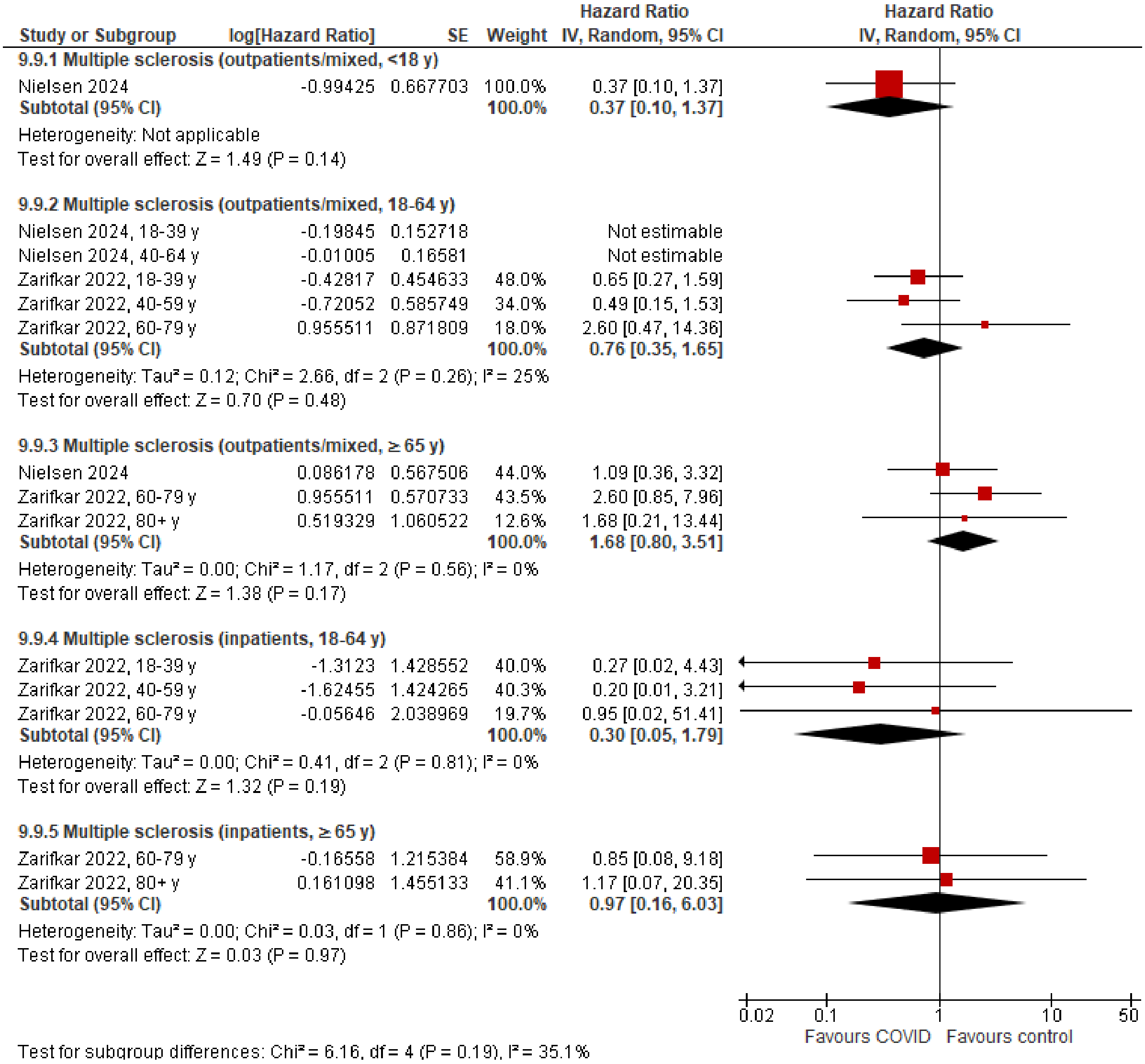

| Study or Subgroup | log[Hazard Ratio] | SE | Weight | Hazard Ratio IV, Random, 95% CI |
|---|---|---|---|---|
| **9.9.1 Multiple sclerosis (outpatients/mixed, <18 y)** | | | | |
| Nielsen 2024 | -0.99425 | 0.667703 | 100.0% | 0.37 [0.10, 1.37] |
| **Subtotal (95% CI)** | | | **100.0%** | **0.37 [0.10, 1.37]** |
| Heterogeneity: Not applicable | | | | |
| Test for overall effect: Z = 1.49 (P = 0.14) | | | | |
| **9.9.2 Multiple sclerosis (outpatients/mixed, 18-64 y)** | | | | |
| Nielsen 2024, 18-39 y | -0.19845 | 0.152718 | | Not estimable |
| Nielsen 2024, 40-64 y | -0.01005 | 0.16581 | | Not estimable |
| Zarifkar 2022, 18-39 y | -0.42817 | 0.454633 | 48.0% | 0.65 [0.27, 1.59] |
| Zarifkar 2022, 40-59 y | -0.72052 | 0.585749 | 34.0% | 0.49 [0.15, 1.53] |
| Zarifkar 2022, 60-79 y | 0.955511 | 0.871809 | 18.0% | 2.60 [0.47, 14.36] |
| **Subtotal (95% CI)** | | | **100.0%** | **0.76 [0.35, 1.65]** |
| Heterogeneity: Tau² = 0.12; Chi² = 2.66, df = 2 (P = 0.26); I² = 25% | | | | |
| Test for overall effect: Z = 0.70 (P = 0.48) | | | | |
| **9.9.3 Multiple sclerosis (outpatients/mixed, ≥ 65 y)** | | | | |
| Nielsen 2024 | 0.086178 | 0.567506 | 44.0% | 1.09 [0.36, 3.32] |
| Zarifkar 2022, 60-79 y | 0.955511 | 0.570733 | 43.5% | 2.60 [0.85, 7.96] |
| Zarifkar 2022, 80+ y | 0.519329 | 1.060522 | 12.6% | 1.68 [0.21, 13.44] |
| **Subtotal (95% CI)** | | | **100.0%** | **1.68 [0.80, 3.51]** |
| Heterogeneity: Tau² = 0.00; Chi² = 1.17, df = 2 (P = 0.56); I² = 0% | | | | |
| Test for overall effect: Z = 1.38 (P = 0.17) | | | | |
| **9.9.4 Multiple sclerosis (inpatients, 18-64 y)** | | | | |
| Zarifkar 2022, 18-39 y | -1.3123 | 1.428552 | 40.0% | 0.27 [0.02, 4.43] |
| Zarifkar 2022, 40-59 y | -1.62455 | 1.424265 | 40.3% | 0.20 [0.01, 3.21] |
| Zarifkar 2022, 60-79 y | -0.05646 | 2.038969 | 19.7% | 0.95 [0.02, 51.41] |
| **Subtotal (95% CI)** | | | **100.0%** | **0.30 [0.05, 1.79]** |
| Heterogeneity: Tau² = 0.00; Chi² = 0.41, df = 2 (P = 0.81); I² = 0% | | | | |
| Test for overall effect: Z = 1.32 (P = 0.19) | | | | |
| **9.9.5 Multiple sclerosis (inpatients, ≥ 65 y)** | | | | |
| Zarifkar 2022, 60-79 y | -0.16558 | 1.215384 | 58.9% | 0.85 [0.08, 9.18] |
| Zarifkar 2022, 80+ y | 0.161098 | 1.455133 | 41.1% | 1.17 [0.07, 20.35] |
| **Subtotal (95% CI)** | | | **100.0%** | **0.97 [0.16, 6.03]** |
| Heterogeneity: Tau² = 0.00; Chi² = 0.03, df = 1 (P = 0.86); I² = 0% | | | | |
| Test for overall effect: Z = 0.03 (P = 0.97) | | | | |
| Test for subgroup differences: Chi² = 6.16, df = 4 (P = 0.19), I² = 35.1% | | | | |

#### Nerve disorders

| Study or Subgroup | log[Hazard Ratio] | SE | Weight | Hazard Ratio IV, Random, 95% CI |
|---|---|---|---|---|
| **9.10.1 Nerve disorders (outpatients/mixed, <18 y)** | | | | |
| Gronkjaer 2023 | -0.2627 | 0.22258 | 26.1% | 0.77 [0.50, 1.19] |
| Nielsen 2024 | 0.113329 | 0.160774 | 31.3% | 1.12 [0.82, 1.53] |
| Roessler 2022 | -0.79851 | 0.925426 | 3.9% | 0.45 [0.07, 2.76] |
| Taquet 2022 | 0.417328 | 0.058337 | 38.7% | 1.52 [1.35, 1.70] |
| **Subtotal (95% CI)** | | | **100.0%** | **1.10 [0.76, 1.61]** |
| Heterogeneity: Tau² = 0.09; Chi² = 12.59, df = 3 (P = 0.006); I² = 76% | | | | |
| Test for overall effect: Z = 0.51 (P = 0.61) | | | | |
| **9.10.2 Nerve disorders (outpatients/mixed, 18-64 y)** | | | | |
| Daugherty 2021 | 0.71784 | 0.130313 | 7.5% | 2.05 [1.59, 2.65] |
| Gronkjaer 2023, 20-39 y | -0.20239 | 0.108552 | 8.7% | 0.82 [0.66, 1.01] |
| Gronkjaer 2023, 40-59 y | -0.01541 | 0.067573 | 11.2% | 0.98 [0.86, 1.12] |
| Gronkjaer 2023, 60-79 y | -0.1201 | 0.083145 | 10.3% | 0.89 [0.75, 1.04] |
| Nielsen 2024, 18-39 y | 0.086178 | 0.046511 | 12.4% | 1.09 [1.00, 1.19] |
| Nielsen 2024, 40-64 y | 0.019803 | 0.030047 | 13.2% | 1.02 [0.96, 1.08] |
| Taquet 2022 | -0.00441 | 0.005842 | 13.7% | 1.00 [0.98, 1.01] |
| Xu 2022 | 0.277632 | 0.027082 | 13.3% | 1.32 [1.25, 1.39] |
| Zang 2023, INSIGHT cohort | 0.057231 | 0.092442 | 9.7% | 1.06 [0.88, 1.27] |
| **Subtotal (95% CI)** | | | **100.0%** | **1.08 [0.97, 1.20]** |
| Heterogeneity: Tau² = 0.02; Chi² = 142.72, df = 8 (P < 0.00001); I² = 94% | | | | |
| Test for overall effect: Z = 1.46 (P = 0.14) | | | | |
| **9.10.3 Nerve disorders (outpatients/mixed, ≥ 65 y)** | | | | |
| Cohen 2022 | 0.19062 | 0.175336 | 8.6% | 1.21 [0.86, 1.71] |
| Gronkjaer 2023, 60-79 y | -0.1201 | 0.099377 | 13.9% | 0.89 [0.73, 1.08] |
| Gronkjaer 2023, 80+ y | -0.45442 | 0.218488 | 6.6% | 0.63 [0.41, 0.97] |
| Nielsen 2024 | 0.165514 | 0.043342 | 18.1% | 1.18 [1.08, 1.28] |
| Taquet 2022 | 0.008762 | 0.01024 | 19.4% | 1.01 [0.99, 1.03] |
| Xu 2022 | 0.270027 | 0.027289 | 18.9% | 1.31 [1.24, 1.38] |
| Zang 2023, INSIGHT cohort | 0.057231 | 0.089785 | 14.6% | 1.06 [0.89, 1.26] |
| **Subtotal (95% CI)** | | | **100.0%** | **1.06 [0.93, 1.22]** |
| Heterogeneity: Tau² = 0.02; Chi² = 96.95, df = 6 (P < 0.00001); I² = 94% | | | | |
| Test for overall effect: Z = 0.88 (P = 0.38) | | | | |
| **9.10.4 Nerve disorders (inpatients, 18-64 y)** | | | | |
| Daugherty 2021 | 1.25954 | 0.148013 | 100.0% | 3.52 [2.64, 4.71] |
| **Subtotal (95% CI)** | | | **100.0%** | **3.52 [2.64, 4.71]** |
| Heterogeneity: Not applicable | | | | |
| Test for overall effect: Z = 8.51 (P < 0.00001) | | | | |
| **9.10.5 Nerve disorders (inpatients, ≥ 65 y)** | | | | |
| Cohen 2022 | 0.365754 | 0.094932 | 100.0% | 1.44 [1.20, 1.74] |
| **Subtotal (95% CI)** | | | **100.0%** | **1.44 [1.20, 1.74]** |
| Heterogeneity: Not applicable | | | | |
| Test for overall effect: Z = 3.85 (P = 0.0001) | | | | |

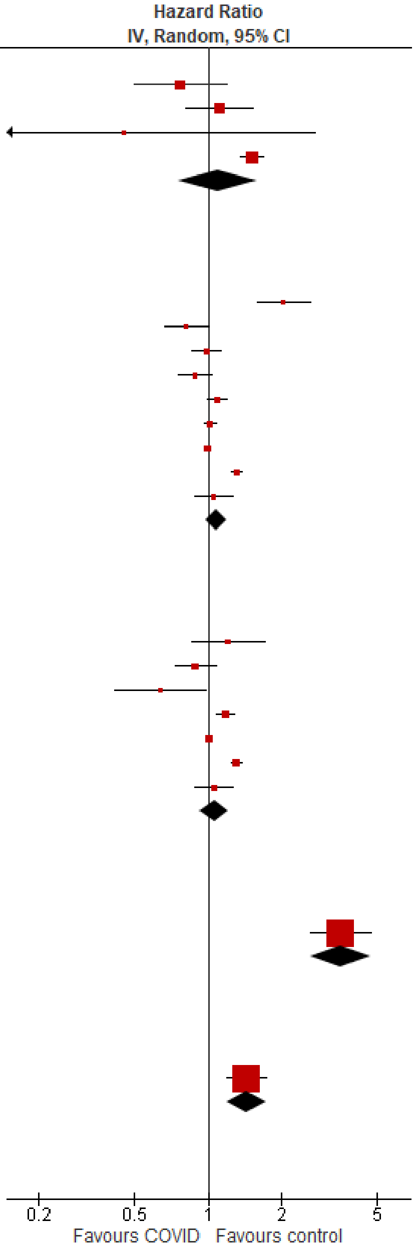

Test for subgroup differences: Chi² = 63.93, df = 4 (P < 0.00001), I² = 93.7%

### Respiratory disorders

#### Overall

| Study or Subgroup | log[Hazard Ratio] | SE | Weight | Hazard Ratio IV, Random, 95% CI |
|---|---|---|---|---|
| **10.1.1 Respiratory disorders, overall (outpatients/mixed, <18 y)** | | | | |
| Gordon 2024, 0-1 y | -0.08234 | 0.060864 | 13.0% | 0.92 [0.82, 1.04] |
| Gordon 2024, 12-17 y | 0.045781 | 0.04267 | 20.0% | 1.05 [0.96, 1.14] |
| Gordon 2024, 2-4 y | -0.12433 | 0.072268 | 10.1% | 0.88 [0.77, 1.02] |
| Gordon 2024, 5-11 y | 0.037271 | 0.054244 | 15.1% | 1.04 [0.93, 1.15] |
| Kompaniyets 2022 | 0 | 0.005102 | 41.2% | 1.00 [0.99, 1.01] |
| Roessler 2022 | 0.65752 | 0.378978 | 0.5% | 1.93 [0.92, 4.06] |
| **Subtotal (95% CI)** | | | **100.0%** | **0.99 [0.94, 1.05]** |
| Heterogeneity: Tau² = 0.00; Chi² = 9.42, df = 5 (P = 0.09); I² = 47% | | | | |
| Test for overall effect: Z = 0.20 (P = 0.84) | | | | |
| **10.1.2 Respiratory disorders, overall (outpatients/mixed, 18-64 y)** | | | | |
| Daugherty 2021 | 2.190555 | 0.254895 | 9.6% | 8.94 [5.42, 14.73] |
| Meng 2024 | 0.410333 | 0.110206 | 12.5% | 1.51 [1.21, 1.87] |
| Mizrahi 2023, 19-40 y | -0.4943 | 0.543782 | 4.7% | 0.61 [0.21, 1.77] |
| Mizrahi 2023, 41-60 y | 0.193874 | 0.247303 | 9.7% | 1.21 [0.75, 1.97] |
| Mizrahi 2023, 60+ y | -0.01137 | 1.620559 | 0.8% | 0.99 [0.04, 23.69] |
| Oh 2024, Japan cohort, 20-39 y | 1.068153 | 0.039324 | 13.4% | 2.91 [2.69, 3.14] |
| Oh 2024, Japan cohort, 40-59 y | 0.947789 | 0.031681 | 13.4% | 2.58 [2.42, 2.75] |
| Oh 2024, Japan cohort, 60+ y | 0.727549 | 0.178137 | 11.2% | 2.07 [1.46, 2.93] |
| Robertson 2024, BMI 20 to 25 | 0.038949 | 0.162655 | 11.6% | 1.04 [0.76, 1.43] |
| Zang 2023, INSIGHT cohort | 0.173953 | 0.069947 | 13.1% | 1.19 [1.04, 1.36] |
| **Subtotal (95% CI)** | | | **100.0%** | **1.89 [1.42, 2.51]** |
| Heterogeneity: Tau² = 0.16; Chi² = 215.47, df = 9 (P < 0.00001); I² = 96% | | | | |
| Test for overall effect: Z = 4.37 (P < 0.0001) | | | | |
| **10.1.3 Respiratory disorders, overall (outpatients/mixed, ≥ 65 y)** | | | | |
| Cohen 2022 | 1.2947 | 0.108294 | 18.2% | 3.65 [2.95, 4.51] |
| Meng 2024 | 0.521952 | 0.097869 | 18.5% | 1.69 [1.39, 2.04] |
| Mizrahi 2023, 60+ y | 0.00878 | 0.387936 | 9.8% | 1.01 [0.47, 2.16] |
| Oh 2024, Japan cohort | 0.530628 | 0.206854 | 15.3% | 1.70 [1.13, 2.55] |
| Oh 2024, South Korea cohort | 0.727549 | 0.077745 | 18.9% | 2.07 [1.78, 2.41] |
| Zang 2023, INSIGHT cohort | 0.280733 | 0.050088 | 19.3% | 1.32 [1.20, 1.46] |
| **Subtotal (95% CI)** | | | **100.0%** | **1.83 [1.31, 2.56]** |
| Heterogeneity: Tau² = 0.15; Chi² = 82.86, df = 5 (P < 0.00001); I² = 94% | | | | |
| Test for overall effect: Z = 3.54 (P = 0.0004) | | | | |
| **10.1.4 Respiratory disorders, overall (inpatients, 18-64 y)** | | | | |
| Ayoubkhani 2021, 18-69 y | 2.351375 | 0.043289 | 36.5% | 10.50 [9.65, 11.43] |
| Daugherty 2021 | 2.790793 | 0.242229 | 30.1% | 16.29 [10.14, 26.19] |
| Robertson 2024, BMI 20 to 25 | 1.515537 | 0.162655 | 33.4% | 4.55 [3.31, 6.26] |
| **Subtotal (95% CI)** | | | **100.0%** | **9.06 [4.93, 16.67]** |
| Heterogeneity: Tau² = 0.26; Chi² = 28.71, df = 2 (P < 0.00001); I² = 93% | | | | |
| Test for overall effect: Z = 7.09 (P < 0.00001) | | | | |
| **10.1.5 Respiratory disorders, overall (inpatients, ≥ 65 y)** | | | | |
| Ayoubkhani 2021, 18-69 y | 2.351375 | 0.149958 | 30.8% | 10.50 [7.83, 14.09] |
| Ayoubkhani 2021, 70+ y | 1.526056 | 0.028061 | 35.4% | 4.60 [4.35, 4.86] |
| Cohen 2022 | 1.950872 | 0.08737 | 33.8% | 7.03 [5.93, 8.35] |
| **Subtotal (95% CI)** | | | **100.0%** | **6.85 [4.38, 10.70]** |
| Heterogeneity: Tau² = 0.15; Chi² = 48.03, df = 2 (P < 0.00001); I² = 96% | | | | |
| Test for overall effect: Z = 8.46 (P < 0.00001) | | | | |

Hazard Ratio
IV, Random, 95% CI

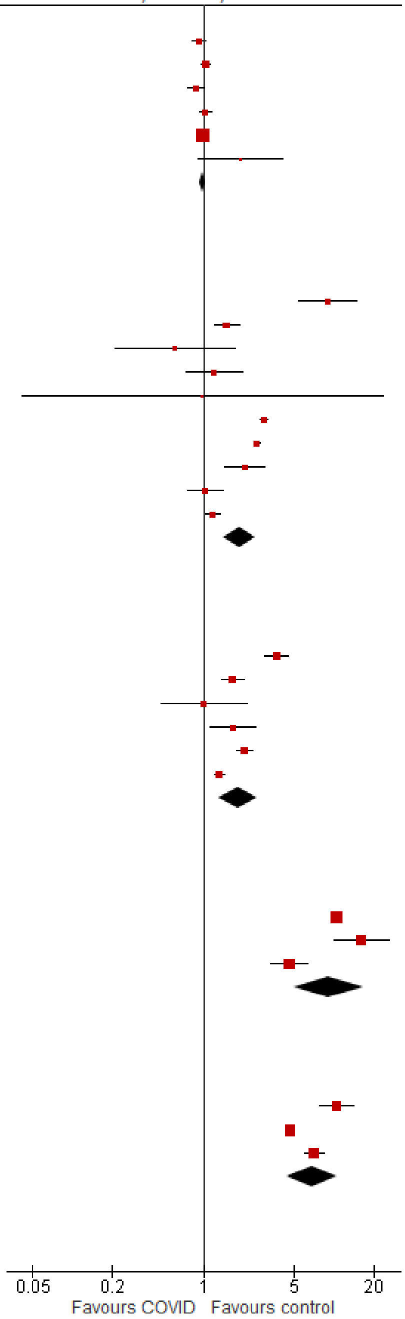

Test for subgroup differences: Chi² = 146.55, df = 4 (P < 0.00001), I² = 97.3%

#### Asthma

| Study or Subgroup | log[Hazard Ratio] | SE | Weight | Hazard Ratio IV, Random, 95% CI |
|---|---|---|---|---|
| **10.2.1 Asthma (outpatients/mixed, <18 y)** | | | | |
| Kompaniyets 2022 | 0 | 0.005102 | 100.0% | 1.00 [0.99, 1.01] |
| **Subtotal (95% CI)** | | | **100.0%** | **1.00 [0.99, 1.01]** |
| Heterogeneity: Not applicable | | | | |
| Test for overall effect: Z = 0.00 (P = 1.00) | | | | |
| **10.2.2 Asthma (outpatients/mixed, 18-64 y)** | | | | |
| Meng 2024 | 0.438255 | 0.101418 | 23.9% | 1.55 [1.27, 1.89] |
| Oh 2024, Japan cohort, 20-39 y | 1.068153 | 0.039324 | 29.8% | 2.91 [2.69, 3.14] |
| Oh 2024, Japan cohort, 40-59 y | 0.947789 | 0.031681 | 30.2% | 2.58 [2.42, 2.75] |
| Oh 2024, Japan cohort, 60+ y | 0.727549 | 0.178137 | 16.1% | 2.07 [1.46, 2.93] |
| **Subtotal (95% CI)** | | | **100.0%** | **2.29 [1.87, 2.80]** |
| Heterogeneity: Tau² = 0.03; Chi² = 35.88, df = 3 (P < 0.00001); I² = 92% | | | | |
| Test for overall effect: Z = 8.04 (P < 0.00001) | | | | |
| **10.2.3 Asthma (outpatients/mixed, ≥ 65 y)** | | | | |
| Meng 2024 | 0.371564 | 0.130823 | 33.9% | 1.45 [1.12, 1.87] |
| Oh 2024, Japan cohort | 0.530628 | 0.206854 | 22.0% | 1.70 [1.13, 2.55] |
| Oh 2024, South Korea cohort | 0.727549 | 0.077745 | 44.1% | 2.07 [1.78, 2.41] |
| **Subtotal (95% CI)** | | | **100.0%** | **1.76 [1.37, 2.25]** |
| Heterogeneity: Tau² = 0.03; Chi² = 5.70, df = 2 (P = 0.06); I² = 65% | | | | |
| Test for overall effect: Z = 4.43 (P < 0.00001) | | | | |

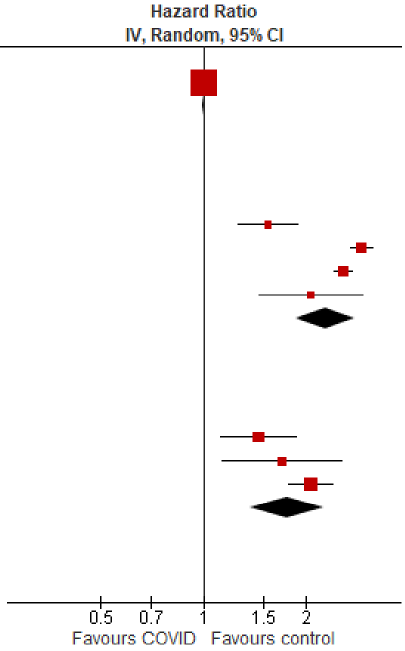

Test for subgroup differences: Chi² = 83.95, df = 2 (P < 0.00001), I² = 97.6%

#### COPD & bronchiectasis

| Study or Subgroup | log[Hazard Ratio] | SE | Weight | Hazard Ratio IV, Random, 95% CI |
|---|---|---|---|---|
| **10.3.1 COPD & bronchiectasis (outpatients/mixed, 18-64 y)** | | | | |
| Meng 2024 | 0.309376 | 0.122734 | 22.7% | 1.36 [1.07, 1.73] |
| Mizrahi 2023, 19-40 y | -0.4943 | 0.543782 | 1.2% | 0.61 [0.21, 1.77] |
| Mizrahi 2023, 41-60 y | 0.193874 | 0.247303 | 5.6% | 1.21 [0.75, 1.97] |
| Mizrahi 2023, 60+ y | -0.01137 | 0.642501 | 0.8% | 0.99 [0.28, 3.48] |
| Zang 2023, INSIGHT cohort | 0.173953 | 0.069947 | 69.8% | 1.19 [1.04, 1.36] |
| **Subtotal (95% CI)** | | | **100.0%** | **1.22 [1.09, 1.36]** |
| Heterogeneity: Tau² = 0.00; Chi² = 2.67, df = 4 (P = 0.61); I² = 0% | | | | |
| Test for overall effect: Z = 3.36 (P = 0.0008) | | | | |
| **10.3.2 COPD & bronchiectasis (outpatients/mixed, ≥ 65 y)** | | | | |
| Meng 2024 | 0.519845 | 0.103179 | 32.3% | 1.68 [1.37, 2.06] |
| Mizrahi 2023, 60+ y | -0.01137 | 0.131345 | 28.2% | 0.99 [0.76, 1.28] |
| Zang 2023, INSIGHT cohort | 0.280733 | 0.050088 | 39.5% | 1.32 [1.20, 1.46] |
| **Subtotal (95% CI)** | | | **100.0%** | **1.32 [1.04, 1.67]** |
| Heterogeneity: Tau² = 0.03; Chi² = 10.26, df = 2 (P = 0.006); I² = 81% | | | | |
| Test for overall effect: Z = 2.28 (P = 0.02) | | | | |

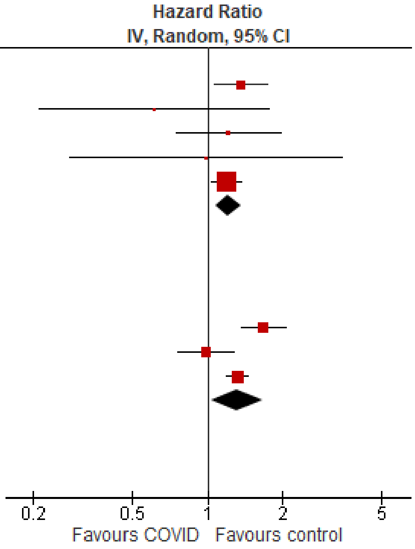

Test for subgroup differences: Chi² = 0.35, df = 1 (P = 0.56), I² = 0%

#### Interstitial lung disease

| Study or Subgroup | log[Hazard Ratio] | SE | Weight | Hazard Ratio IV, Random, 95% CI |
|---|---|---|---|---|
| **10.4.1 Interstitial lung disease (outpatients/mixed, 18-64 y)** | | | | |
| Daugherty 2021 | 2.042518 | 0.227619 | 49.6% | 7.71 [4.94, 12.04] |
| Meng 2024 | 0.536493 | 0.177875 | 50.4% | 1.71 [1.21, 2.42] |
| **Subtotal (95% CI)** | | | **100.0%** | **3.61 [0.82, 15.78]** |
| Heterogeneity: Tau² = 1.09; Chi² = 27.18, df = 1 (P < 0.00001); I² = 96% | | | | |
| Test for overall effect: Z = 1.70 (P = 0.09) | | | | |
| **10.4.2 Interstitial lung disease (outpatients/mixed, ≥ 65 y)** | | | | |
| Cohen 2022 | 1.12167756 | 0.118055 | 50.4% | 3.07 [2.44, 3.87] |
| Meng 2024 | 0.662688 | 0.125154 | 49.6% | 1.94 [1.52, 2.48] |
| **Subtotal (95% CI)** | | | **100.0%** | **2.45 [1.56, 3.83]** |
| Heterogeneity: Tau² = 0.09; Chi² = 7.12, df = 1 (P = 0.008); I² = 86% | | | | |
| Test for overall effect: Z = 3.90 (P < 0.0001) | | | | |
| **10.4.3 Interstitial lung disease (inpatients, 18-64 y)** | | | | |
| Daugherty 2021 | 2.530931 | 0.216399 | 100.0% | 12.57 [8.22, 19.20] |
| **Subtotal (95% CI)** | | | **100.0%** | **12.57 [8.22, 19.20]** |
| Heterogeneity: Not applicable | | | | |
| Test for overall effect: Z = 11.70 (P < 0.00001) | | | | |
| **10.4.4 Interstitial lung disease (inpatients, ≥ 65 y)** | | | | |
| Cohen 2022 | 2.032717 | 0.09874 | 100.0% | 7.63 [6.29, 9.26] |
| **Subtotal (95% CI)** | | | **100.0%** | **7.63 [6.29, 9.26]** |
| Heterogeneity: Not applicable | | | | |
| Test for overall effect: Z = 20.59 (P < 0.00001) | | | | |

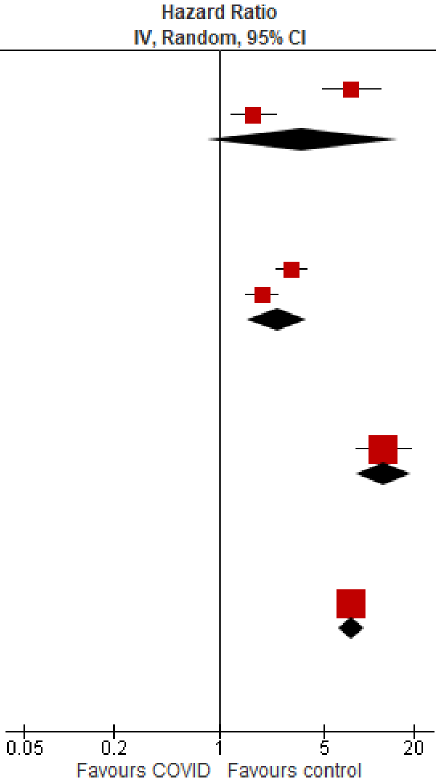

Test for subgroup differences: Chi² = 29.86, df = 3 (P < 0.00001), I² = 90.0%

#### Respiratory failure

| Study or Subgroup | log[Hazard Ratio] | SE | Weight | Hazard Ratio IV, Random, 95% CI |
|---|---|---|---|---|
| **10.5.1 Respiratory failure (outpatients/mixed, 18-64 y)** | | | | |
| Daugherty 2021 | 2.55334381 | 0.356327 | 100.0% | 12.85 [6.39, 25.84] |
| **Subtotal (95% CI)** | | | **100.0%** | **12.85 [6.39, 25.84]** |
| Heterogeneity: Not applicable | | | | |
| Test for overall effect: Z = 7.17 (P < 0.00001) | | | | |
| **10.5.2 Respiratory failure (outpatients/mixed, ≥ 65 y)** | | | | |
| Cohen 2022 | 1.51072194 | 0.131911 | 61.3% | 4.53 [3.50, 5.87] |
| Mizrahi 2023, 60+ y | 0.124079 | 0.673456 | 38.7% | 1.13 [0.30, 4.24] |
| **Subtotal (95% CI)** | | | **100.0%** | **2.65 [0.71, 9.96]** |
| Heterogeneity: Tau² = 0.73; Chi² = 4.08, df = 1 (P = 0.04); I² = 76% | | | | |
| Test for overall effect: Z = 1.44 (P = 0.15) | | | | |
| **10.5.3 Respiratory failure (inpatients, 18-64 y)** | | | | |
| Daugherty 2021 | 3.426794 | 0.338542 | 100.0% | 30.78 [15.85, 59.76] |
| **Subtotal (95% CI)** | | | **100.0%** | **30.78 [15.85, 59.76]** |
| Heterogeneity: Not applicable | | | | |
| Test for overall effect: Z = 10.12 (P < 0.00001) | | | | |
| **10.5.4 Respiratory failure (inpatients, ≥ 65 y)** | | | | |
| Cohen 2022 | 1.861783 | 0.103017 | 100.0% | 6.44 [5.26, 7.87] |
| **Subtotal (95% CI)** | | | **100.0%** | **6.44 [5.26, 7.87]** |
| Heterogeneity: Not applicable | | | | |
| Test for overall effect: Z = 18.07 (P < 0.00001) | | | | |

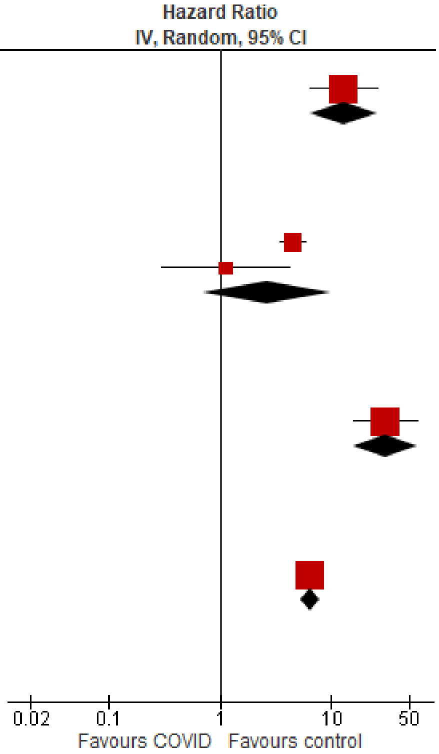

Test for subgroup differences: Chi² = 24.26, df = 3 (P < 0.0001), I² = 87.6%

#### Sleep disorders

##### Overall

| Study or Subgroup | log[Hazard Ratio] | SE | Weight | Hazard Ratio IV, Random, 95% CI |
|---|---|---|---|---|
| **11.1.1 Sleep disorders, overall (outpatients/mixed, <18 y)** | | | | |
| Kompaniyets 2022, 12-17 y | -0.09431 | 0.013943 | 24.3% | 0.91 [0.89, 0.94] |
| Kompaniyets 2022, 2-4 y | -0.01005 | 0.033377 | 20.3% | 0.99 [0.93, 1.06] |
| Kompaniyets 2022, 5-11 y | -0.11653 | 0.017204 | 23.7% | 0.89 [0.86, 0.92] |
| Mizrahi 2023, 0-4 y | 0.350657 | 0.446788 | 0.6% | 1.42 [0.59, 3.41] |
| Roessler 2022 | 0.398776 | 0.144739 | 4.5% | 1.49 [1.12, 1.98] |
| Taquet 2022 | 0.02557 | 0.026073 | 22.0% | 1.03 [0.97, 1.08] |
| Tartof 2022 | -0.09431 | 0.154972 | 4.0% | 0.91 [0.67, 1.23] |
| Valdivieso-Martinez 2023, unvaccinated cohort | 0.875469 | 0.53192 | 0.4% | 2.40 [0.85, 6.81] |
| Valdivieso-Martinez 2023, vaccinated cohort | 0 | 0.999776 | 0.1% | 1.00 [0.14, 7.10] |
| **Subtotal (95% CI)** | | | **100.0%** | **0.97 [0.91, 1.04]** |
| Heterogeneity: Tau² = 0.00; Chi² = 41.24, df = 8 (P < 0.00001); I² = 81% | | | | |
| Test for overall effect: Z = 0.80 (P = 0.42) | | | | |
| **11.1.2 Sleep disorders, overall (outpaient/mixed, 18-64 y)** | | | | |
| Daugherty 2021 | 0.576613 | 0.062672 | 30.2% | 1.78 [1.57, 2.01] |
| Lin 2024 | 0.524729 | 0.013629 | 33.5% | 1.69 [1.65, 1.74] |
| Mizrahi 2023, 19-40 y | 0.487125 | 1.617151 | 0.4% | 1.63 [0.07, 38.73] |
| Mizrahi 2023, 41-60 y | 0.444686 | 1.581163 | 0.5% | 1.56 [0.07, 34.59] |
| Mizrahi 2023, 60+ y | -1.20397 | 1.620559 | 0.4% | 0.30 [0.01, 7.19] |
| Taquet 2022 | 0.030044 | 1.54256 | 0.5% | 1.03 [0.05, 21.19] |
| Zang 2023, INSIGHT cohort | 0.218493 | 0.033683 | 32.6% | 1.24 [1.16, 1.33] |
| Zarifkar 2022, 18-39 y | -0.84188 | 1.42859 | 0.6% | 0.43 [0.03, 7.09] |
| Zarifkar 2022, 40-59 y | -0.27589 | 1.055883 | 1.0% | 0.76 [0.10, 6.01] |
| Zarifkar 2022, 60-79 y | 1.196767 | 1.617151 | 0.4% | 3.31 [0.14, 78.75] |
| **Subtotal (95% CI)** | | | **100.0%** | **1.52 [1.23, 1.87]** |
| Heterogeneity: Tau² = 0.03; Chi² = 75.99, df = 9 (P < 0.00001); I² = 88% | | | | |
| Test for overall effect: Z = 3.89 (P = 0.0001) | | | | |
| **11.1.3 Sleep disorders, overall (outpatients/mixed, ≥ 65 y)** | | | | |
| Cohen 2022 | 0.405465 | 0.094435 | 13.4% | 1.50 [1.25, 1.80] |
| Lin 2024 | 0.405465 | 0.008533 | 16.4% | 1.50 [1.48, 1.53] |
| Mizrahi 2023, 60+ y | -1.20397 | 0.724736 | 1.2% | 0.30 [0.07, 1.24] |
| Murata 2024, alpha variant | -0.82098 | 1.002182 | 0.6% | 0.44 [0.06, 3.14] |
| Murata 2024, delta variant | -0.37106 | 0.166409 | 9.7% | 0.69 [0.50, 0.96] |
| Murata 2024, omicron BA.1/BA.2 variant | -0.54473 | 0.126888 | 11.7% | 0.58 [0.45, 0.74] |
| Murata 2024, omicron BA.5 variant | 0.322083 | 0.451043 | 2.7% | 1.38 [0.57, 3.34] |
| Taquet 2022 | 0.14842 | 0.026413 | 16.2% | 1.16 [1.10, 1.22] |
| Wan 2023a, UK cohort | 0.336472 | 0.139424 | 11.1% | 1.40 [1.07, 1.84] |
| Zang 2023, INSIGHT cohort | 0.359142 | 0.046804 | 15.6% | 1.43 [1.31, 1.57] |
| Zarifkar 2022, 60-79 y | 1.196767 | 0.933663 | 0.7% | 3.31 [0.53, 20.63] |
| Zarifkar 2022, 80+ y | 0.989169 | 0.979509 | 0.7% | 2.69 [0.39, 18.34] |
| **Subtotal (95% CI)** | | | **100.0%** | **1.15 [0.98, 1.35]** |
| Heterogeneity: Tau² = 0.04; Chi² = 166.62, df = 11 (P < 0.00001); I² = 93% | | | | |
| Test for overall effect: Z = 1.76 (P = 0.08) | | | | |
| **11.1.4 Sleep disorders, overall (inpatients, 18-64 y)** | | | | |
| Daugherty 2021 | 1.306141 | 0.076897 | 99.4% | 3.69 [3.18, 4.29] |
| Zarifkar 2022, 18-39 y | 1.590846 | 1.467219 | 0.3% | 4.91 [0.28, 87.06] |
| Zarifkar 2022, 40-59 y | 1.083702 | 1.511694 | 0.3% | 2.96 [0.15, 57.20] |
| Zarifkar 2022, 60-79 y | 1.255445 | 3.02356 | 0.1% | 3.51 [0.01, 1314.90] |
| **Subtotal (95% CI)** | | | **100.0%** | **3.69 [3.18, 4.29]** |
| Heterogeneity: Tau² = 0.00; Chi² = 0.06, df = 3 (P = 1.00); I² = 0% | | | | |
| Test for overall effect: Z = 17.04 (P < 0.00001) | | | | |
| **11.1.5 Sleep disorders, overall (inpatients, ≥ 65 y)** | | | | |
| Cohen 2022 | 0.763699 | 0.076538 | 21.3% | 2.15 [1.85, 2.49] |
| Murata 2024, alpha variant | 0.122218 | 0.306312 | 16.1% | 1.13 [0.62, 2.06] |
| Murata 2024, delta variant | 0.113329 | 0.233028 | 18.1% | 1.12 [0.71, 1.77] |
| Murata 2024, omicron BA.1/BA.2 variant | -0.18633 | 0.127153 | 20.5% | 0.83 [0.65, 1.06] |
| Murata 2024, omicron BA.5 variant | -0.07257 | 0.145864 | 20.2% | 0.93 [0.70, 1.24] |
| Zarifkar 2022, 60-79 y | 1.255445 | 1.745653 | 1.8% | 3.51 [0.11, 107.43] |
| Zarifkar 2022, 80+ y | 1.895654 | 1.632792 | 2.0% | 6.66 [0.27, 163.35] |
| **Subtotal (95% CI)** | | | **100.0%** | **1.23 [0.77, 1.98]** |
| Heterogeneity: Tau² = 0.27; Chi² = 57.71, df = 6 (P < 0.00001); I² = 90% | | | | |
| Test for overall effect: Z = 0.87 (P = 0.38) | | | | |

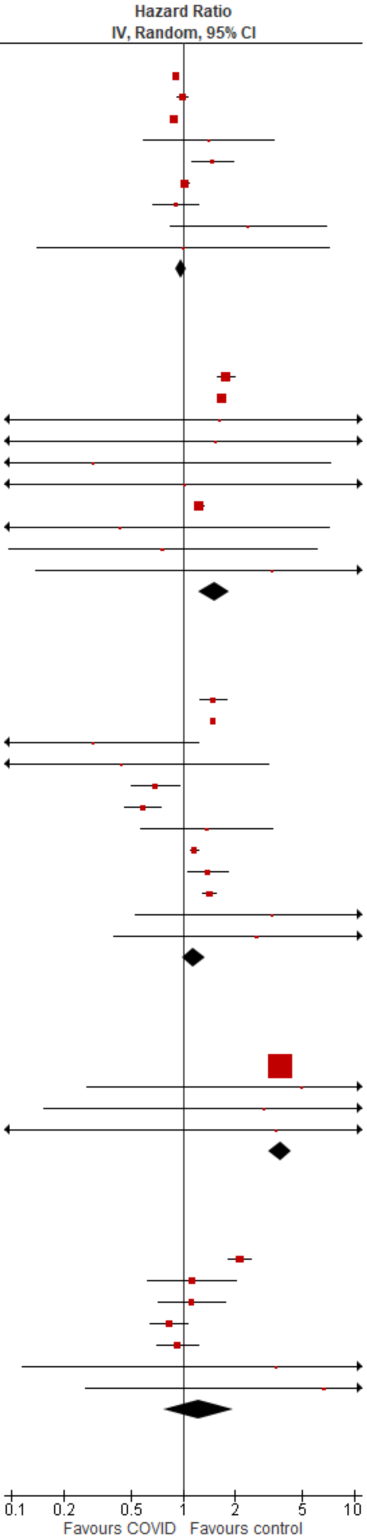

Test for subgroup differences: Chi² = 257.48, df = 4 (P < 0.00001), I² = 98.4%

#### Insomnia

| Study or Subgroup | log[Hazard Ratio] | SE | Weight | Hazard Ratio IV, Random, 95% CI |
|---|---|---|---|---|
| **11.2.1 Insomnia (outpatients/mixed, <18 y)** | | | | |
| Mizrahi 2023, 0-4 y | 0.350657 | 0.446788 | 0.3% | 1.42 [0.59, 3.41] |
| Taquet 2022 | 0.02557 | 0.026471 | 99.4% | 1.03 [0.97, 1.08] |
| Valdivieso-Martinez 2023, unvaccinated cohort | 0 | 1.339193 | 0.0% | 1.00 [0.07, 13.80] |
| Valdivieso-Martinez 2023, vaccinated cohort | 0.875469 | 0.612855 | 0.2% | 2.40 [0.72, 7.98] |
| **Subtotal (95% CI)** | | | **100.0%** | **1.03 [0.98, 1.08]** |
| Heterogeneity: Tau² = 0.00; Chi² = 2.44, df = 3 (P = 0.49); I² = 0% | | | | |
| Test for overall effect: Z = 1.07 (P = 0.28) | | | | |
| **11.2.2 Insomnia (outpatients/mixed, 18-64 y)** | | | | |
| Mizrahi 2023, 19-40 y | 0.487125 | 1.617151 | 24.1% | 1.63 [0.07, 38.73] |
| Mizrahi 2023, 41-60 y | 0.444686 | 1.581163 | 25.3% | 1.56 [0.07, 34.59] |
| Mizrahi 2023, 60+ y | -1.20397 | 1.620559 | 24.0% | 0.30 [0.01, 7.19] |
| Taquet 2022 | 0.030044 | 1.54256 | 26.5% | 1.03 [0.05, 21.19] |
| **Subtotal (95% CI)** | | | **100.0%** | **0.95 [0.20, 4.51]** |
| Heterogeneity: Tau² = 0.00; Chi² = 0.72, df = 3 (P = 0.87); I² = 0% | | | | |
| Test for overall effect: Z = 0.06 (P = 0.95) | | | | |
| **11.2.3 Insomnia (outpatients/mixed, ≥ 65 y)** | | | | |
| Mizrahi 2023, 60+ y | -1.20397 | 0.724736 | 7.0% | 0.30 [0.07, 1.24] |
| Murata 2024, alpha variant | -0.82098 | 1.002182 | 4.2% | 0.44 [0.06, 3.14] |
| Murata 2024, delta variant | -0.37106 | 0.166409 | 23.7% | 0.69 [0.50, 0.96] |
| Murata 2024, omicron BA.1/BA.2 variant | -0.54473 | 0.126888 | 25.1% | 0.58 [0.45, 0.74] |
| Murata 2024, omicron BA.5 variant | 0.322083 | 0.451043 | 12.9% | 1.38 [0.57, 3.34] |
| Taquet 2022 | 0.14842 | 0.026413 | 27.2% | 1.16 [1.10, 1.22] |
| **Subtotal (95% CI)** | | | **100.0%** | **0.77 [0.50, 1.19]** |
| Heterogeneity: Tau² = 0.18; Chi² = 41.45, df = 5 (P < 0.00001); I² = 88% | | | | |
| Test for overall effect: Z = 1.17 (P = 0.24) | | | | |
| **11.2.4 Insomnia (inpatients, ≥ 65 y)** | | | | |
| Murata 2024, alpha variant | 0.122218 | 0.306312 | 7.7% | 1.13 [0.62, 2.06] |
| Murata 2024, delta variant | 0.113329 | 0.233028 | 13.4% | 1.12 [0.71, 1.77] |
| Murata 2024, omicron BA.1/BA.2 variant | -0.18633 | 0.127153 | 44.8% | 0.83 [0.65, 1.06] |
| Murata 2024, omicron BA.5 variant | -0.07257 | 0.145864 | 34.1% | 0.93 [0.70, 1.24] |
| **Subtotal (95% CI)** | | | **100.0%** | **0.92 [0.78, 1.09]** |
| Heterogeneity: Tau² = 0.00; Chi² = 1.82, df = 3 (P = 0.61); I² = 0% | | | | |
| Test for overall effect: Z = 0.98 (P = 0.33) | | | | |

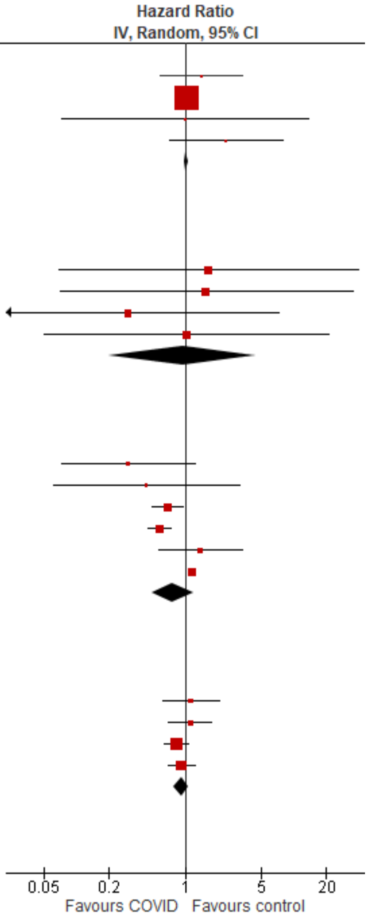

Test for subgroup differences: Chi² = 3.15, df = 3 (P = 0.37), I² = 4.7%

Narcolepsy

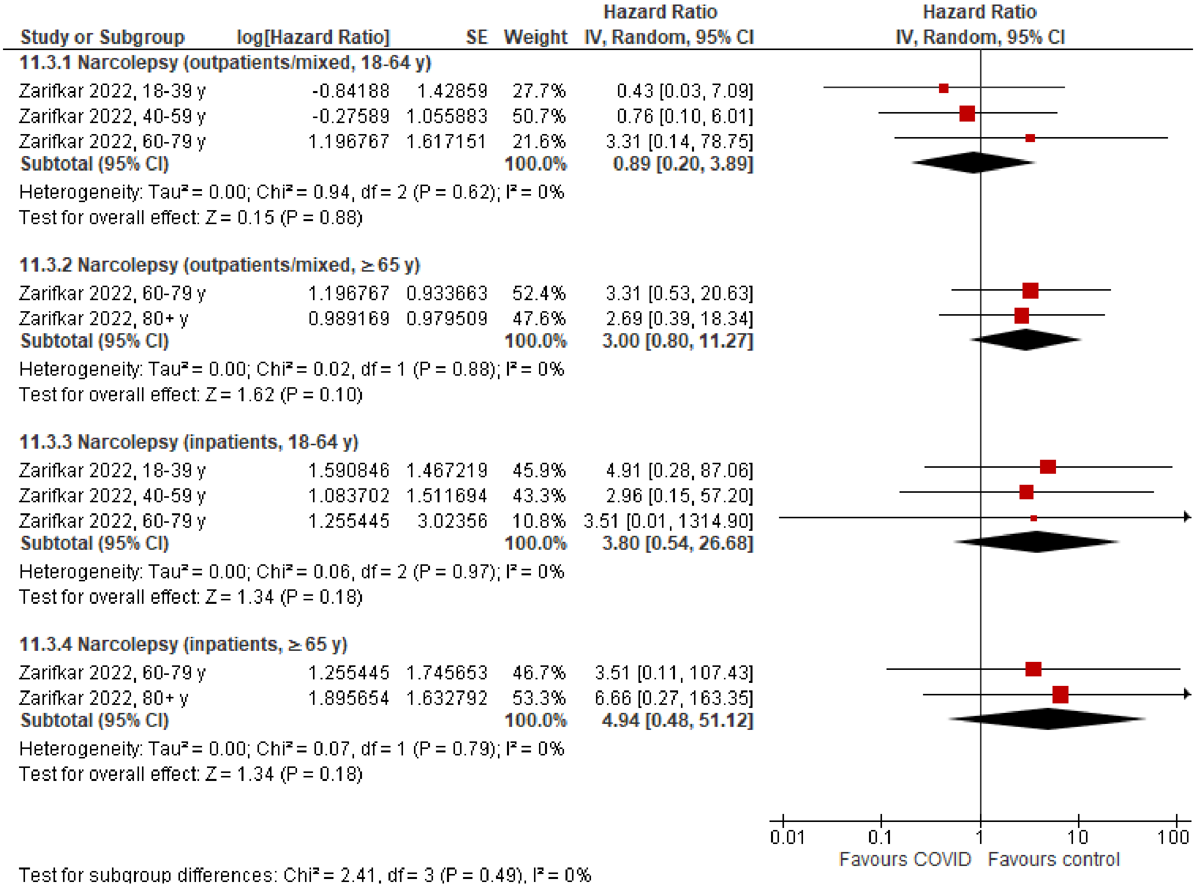

Sleep apnea

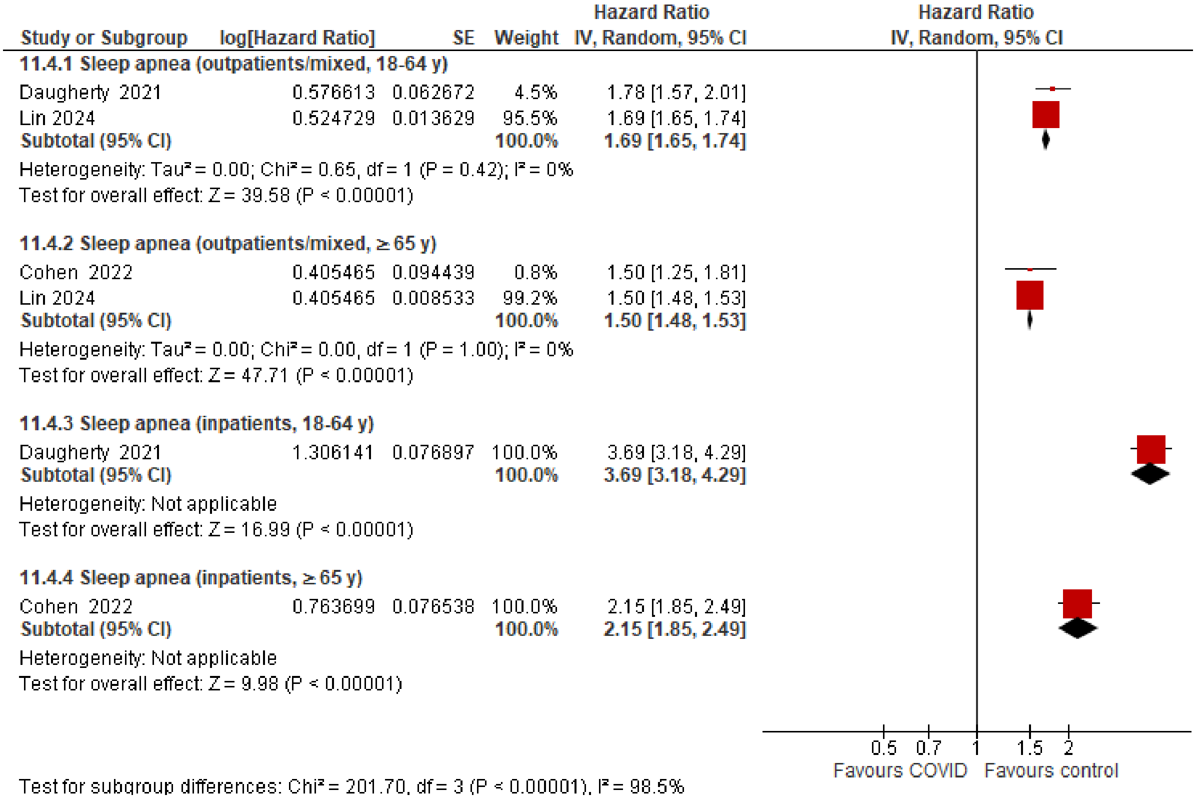

### Stroke

#### Overall

| Study or Subgroup | log[Hazard Ratio] | SE | Weight | Hazard Ratio IV, Random, 95% CI |
|---|---|---|---|---|
| **12.1.1 Stroke, overall (outpatients/mixed, <18 y)** | | | | |
| Kompaniyets 2022 | 0.182322 | 0.094787 | 51.9% | 1.20 [1.00, 1.44] |
| Nielsen 2024 | -0.10536 | 0.401708 | 2.9% | 0.90 [0.41, 1.98] |
| Roessler 2022 | 0.223144 | 0.603648 | 1.3% | 1.25 [0.38, 4.08] |
| Taquet 2022 | 0.092397 | 0.104033 | 43.1% | 1.10 [0.89, 1.34] |
| Tartof 2022 | -0.65393 | 0.740858 | 0.8% | 0.52 [0.12, 2.22] |
| **Subtotal (95% CI)** | | | **100.0%** | **1.14 [0.99, 1.30]** |
| Heterogeneity: Tau² = 0.00; Chi² = 1.92, df = 4 (P = 0.75); I² = 0% | | | | |
| Test for overall effect: Z = 1.88 (P = 0.06) | | | | |
| **12.1.2 Stroke, overall (outpatients/mixed, 18-64 y)** | | | | |
| Batistoni 2024 | 0.494696 | 0.382586 | 2.8% | 1.64 [0.77, 3.47] |
| Daugherty 2021 | 0.900161 | 0.157049 | 5.5% | 2.46 [1.81, 3.35] |
| Lam 2023 | -0.24846 | 0.391448 | 2.7% | 0.78 [0.36, 1.68] |
| Lim 2023 | 0.48858 | 0.135964 | 5.8% | 1.63 [1.25, 2.13] |
| Mizrahi 2023, 19-40 y | -1.34707 | 0.772582 | 1.0% | 0.26 [0.06, 1.18] |
| Mizrahi 2023, 41-60 y | -0.04602 | 0.413093 | 2.5% | 0.96 [0.42, 2.15] |
| Mizrahi 2023, 60+ y | 0.198851 | 0.918382 | 0.7% | 1.22 [0.20, 7.38] |
| Nielsen 2024, 18-39 y | 0.19062 | 0.123234 | 6.0% | 1.21 [0.95, 1.54] |
| Nielsen 2024, 40-64 y | -0.21072 | 0.037399 | 6.8% | 0.81 [0.75, 0.87] |
| Rezel-Potts 2022 | -0.26487 | 0.118513 | 6.0% | 0.77 [0.61, 0.97] |
| Taquet 2022 | -0.10536 | 0.014369 | 6.8% | 0.90 [0.88, 0.93] |
| Velasquez Garcia 2024 | 0.323025 | 0.121986 | 6.0% | 1.38 [1.09, 1.75] |
| Wang 2022b, 20-44y | 0.267734 | 0.064357 | 6.6% | 1.31 [1.15, 1.48] |
| Wang 2022b, 45-64y | 0.402126 | 0.039324 | 6.7% | 1.49 [1.38, 1.61] |
| Xie 2022b | 0.364643 | 0.054939 | 6.7% | 1.44 [1.29, 1.60] |
| Xu 2022 | 0.470004 | 0.035132 | 6.8% | 1.60 [1.49, 1.71] |
| Zappacosta 2022, 18-30 y | 1.530395 | 1.116326 | 0.5% | 4.62 [0.52, 41.20] |
| Zappacosta 2022, 31-40 y | 0.00995 | 0.537656 | 1.8% | 1.01 [0.35, 2.90] |
| Zappacosta 2022, 41-50 y | -0.30111 | 0.396695 | 2.7% | 0.74 [0.34, 1.61] |
| Zappacosta 2022, 51-60 y | 0.139762 | 0.252027 | 4.2% | 1.15 [0.70, 1.88] |
| Zappacosta 2022, 61-70 y | -0.10536 | 0.71835 | 1.1% | 0.90 [0.22, 3.68] |
| Zarifkar 2022, 18-39 y | -1.87015 | 1.003109 | 0.6% | 0.15 [0.02, 1.10] |
| Zarifkar 2022, 40-59 y | -0.69315 | 0.216147 | 4.7% | 0.50 [0.33, 0.76] |
| Zarifkar 2022, 60-79 y | -0.35667 | 0.188849 | 5.1% | 0.70 [0.48, 1.01] |
| **Subtotal (95% CI)** | | | **100.0%** | **1.11 [0.95, 1.31]** |
| Heterogeneity: Tau² = 0.10; Chi² = 509.93, df = 23 (P < 0.00001); I² = 95% | | | | |
| Test for overall effect: Z = 1.30 (P = 0.19) | | | | |
| **12.1.3 Stroke, overall (outpatients/mixed, ≥ 65 y)** | | | | |
| Batistoni 2024 | 0.463734 | 0.201786 | 6.1% | 1.59 [1.07, 2.36] |
| Cohen 2022 | 0.652325 | 0.076443 | 8.6% | 1.92 [1.65, 2.23] |
| Mizrahi 2023, 60+ y | 0.410713 | 0.918382 | 0.8% | 1.51 [0.25, 9.12] |
| Nielsen 2024 | -0.17435 | 0.033256 | 9.1% | 0.84 [0.79, 0.90] |
| Taquet 2022 | 0.070552 | 0.014475 | 9.2% | 1.07 [1.04, 1.10] |
| Wan 2023a, UK cohort | 0.09531 | 0.197698 | 6.1% | 1.10 [0.75, 1.62] |
| Wang 2022b | 0.535908 | 0.033565 | 9.1% | 1.71 [1.60, 1.83] |
| Xie 2022b | 0.425268 | 0.031617 | 9.1% | 1.53 [1.44, 1.63] |
| Xu 2022 | 0.425268 | 0.058421 | 8.8% | 1.53 [1.36, 1.72] |
| Yang 2024 | 0.002934 | 0.051066 | 8.9% | 1.00 [0.91, 1.11] |
| Zappacosta 2022, 61-70 y | -0.10536 | 0.378604 | 3.3% | 0.90 [0.43, 1.89] |
| Zappacosta 2022, 70+ y | 0.398776 | 0.223997 | 5.6% | 1.49 [0.96, 2.31] |
| Zarifkar 2022, 60-79 y | -0.35667 | 0.119439 | 7.8% | 0.70 [0.55, 0.88] |
| Zarifkar 2022, 80+ y | -0.51083 | 0.119899 | 7.8% | 0.60 [0.47, 0.76] |
| **Subtotal (95% CI)** | | | **100.0%** | **1.17 [0.99, 1.39]** |
| Heterogeneity: Tau² = 0.08; Chi² = 459.52, df = 13 (P < 0.00001); I² = 97% | | | | |
| Test for overall effect: Z = 1.88 (P = 0.06) | | | | |

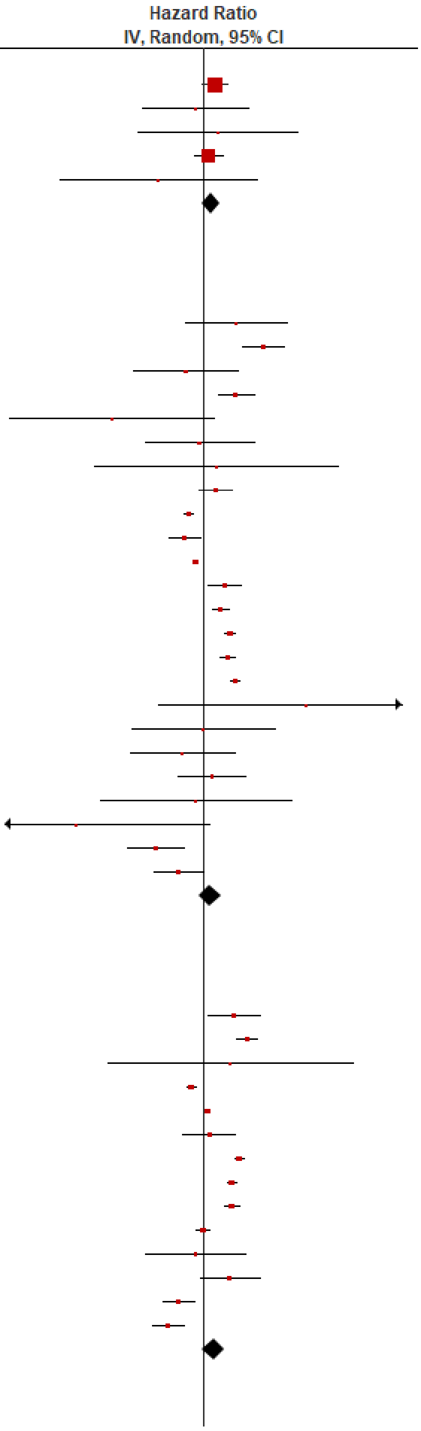

| Study or Subgroup | log[Hazard Ratio] | SE | Weight | Hazard Ratio IV, Random, 95% CI |
|---|---|---|---|---|
| **12.1.4 Stroke, overall (inpatients, 18-64 y)** | | | | |
| Daugherty 2021 | 1.35702 | 0.161755 | 16.4% | 3.88 [2.83, 5.33] |
| Ho 2021 | 0.482426 | 0.61076 | 13.4% | 1.62 [0.49, 5.36] |
| Lee 2023, 18-39 y | 1.366092 | 0.94001 | 10.6% | 3.92 [0.62, 24.74] |
| Lee 2023, 40-64 y | 0.19062 | 0.322976 | 15.6% | 1.21 [0.64, 2.28] |
| Zarifkar 2022, 18-39 y | 0.10499 | 0.712564 | 12.5% | 1.11 [0.27, 4.49] |
| Zarifkar 2022, 40-59 y | -1.60944 | 0.353647 | 15.4% | 0.20 [0.10, 0.40] |
| Zarifkar 2022, 60-79 y | -0.69315 | 0.258592 | 16.0% | 0.50 [0.30, 0.83] |
| **Subtotal (95% CI)** | | | **100.0%** | **1.12 [0.42, 3.03]** |
| Heterogeneity: Tau² = 1.54; Chi² = 86.72, df = 6 (P < 0.00001); I² = 93% | | | | |
| Test for overall effect: Z = 0.23 (P = 0.82) | | | | |
| **12.1.5 Stroke, overall (inpatients, ≥ 65 y)** | | | | |
| Cohen 2022 | 1.089908 | 0.060979 | 17.7% | 2.97 [2.64, 3.35] |
| Ho 2021, 66-80 y | -0.41552 | 0.518223 | 14.7% | 0.66 [0.24, 1.82] |
| Ho 2021, 80+ y | -0.51083 | 0.462342 | 15.3% | 0.60 [0.24, 1.48] |
| Lee 2023 | -0.54473 | 0.217105 | 17.2% | 0.58 [0.38, 0.89] |
| Zarifkar 2022, 60-79 y | -0.69315 | 0.119439 | 17.6% | 0.50 [0.40, 0.63] |
| Zarifkar 2022, 80+ y | -0.69315 | 0.142759 | 17.5% | 0.50 [0.38, 0.66] |
| **Subtotal (95% CI)** | | | **100.0%** | **0.75 [0.30, 1.92]** |
| Heterogeneity: Tau² = 1.28; Chi² = 293.83, df = 5 (P < 0.00001); I² = 98% | | | | |
| Test for overall effect: Z = 0.59 (P = 0.55) | | | | |

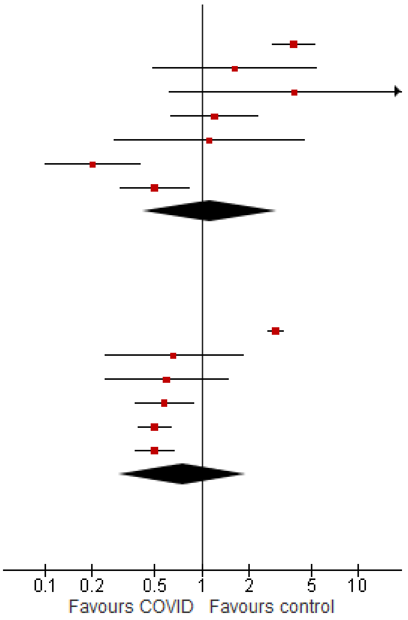

Test for subgroup differences: Chi² = 0.94, df = 4 (P = 0.92), I² = 0%

#### Hemorrhagic stroke

| Study or Subgroup | log[Hazard Ratio] | SE | Weight | Hazard Ratio IV, Random, 95% CI |
|---|---|---|---|---|
| **12.2.1 Hemorrhagic stroke (outpatients/mixed, 18-64 y)** | | | | |
| Daugherty 2021 | 0.951658 | 0.309298 | 100.0% | 2.59 [1.41, 4.75] |
| **Subtotal (95% CI)** | | | **100.0%** | **2.59 [1.41, 4.75]** |
| Heterogeneity: Not applicable | | | | |
| Test for overall effect: Z = 3.08 (P = 0.002) | | | | |
| **12.2.2 Hemorrhagic stroke (outpatients/mixed, ≥ 65 y)** | | | | |
| Cohen 2022 | 0.71295 | 0.098323 | 50.9% | 2.04 [1.68, 2.47] |
| Yang 2024 | -0.03222 | 0.138595 | 49.1% | 0.97 [0.74, 1.27] |
| **Subtotal (95% CI)** | | | **100.0%** | **1.41 [0.68, 2.94]** |
| Heterogeneity: Tau² = 0.26; Chi² = 19.23, df = 1 (P < 0.0001); I² = 95% | | | | |
| Test for overall effect: Z = 0.93 (P = 0.35) | | | | |
| **12.2.3 Hemorrhagic stroke (inpatients, 18-64 y)** | | | | |
| Daugherty 2021 | 1.580442 | 0.31634 | 100.0% | 4.86 [2.61, 9.03] |
| **Subtotal (95% CI)** | | | **100.0%** | **4.86 [2.61, 9.03]** |
| Heterogeneity: Not applicable | | | | |
| Test for overall effect: Z = 5.00 (P < 0.00001) | | | | |
| **12.2.4 Hemorrhagic stroke (inpatients, ≥ 65 y)** | | | | |
| Cohen 2022 | 1.026436 | 0.078765 | 100.0% | 2.79 [2.39, 3.26] |
| **Subtotal (95% CI)** | | | **100.0%** | **2.79 [2.39, 3.26]** |
| Heterogeneity: Not applicable | | | | |
| Test for overall effect: Z = 13.03 (P < 0.00001) | | | | |

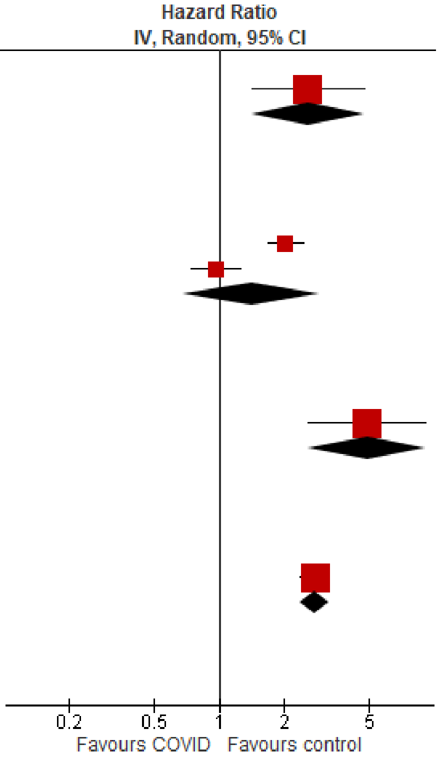

Test for subgroup differences: Chi² = 6.45, df = 3 (P = 0.09), I² = 53.5%

#### Ischemic stroke

| Study or Subgroup | log[Hazard Ratio] | SE | Weight | Hazard Ratio IV, Random, 95% CI |
|---|---|---|---|---|
| **12.3.1 Ischemic stroke (outpatients/mixed, <18 y)** | | | | |
| Taquet 2022 | 0.092397 | 0.104033 | 100.0% | 1.10 [0.89, 1.34] |
| **Subtotal (95% CI)** | | | **100.0%** | **1.10 [0.89, 1.34]** |
| Heterogeneity: Not applicable | | | | |
| Test for overall effect: Z = 0.89 (P = 0.37) | | | | |
| **12.3.2 Ischemic stroke (outpatients/mixed, 18-64 y)** | | | | |
| Daugherty 2021 | 0.947789 | 0.44898 | 12.8% | 2.58 [1.07, 6.22] |
| Taquet 2022 | -0.10536 | 0.014369 | 33.3% | 0.90 [0.88, 0.93] |
| Zarifkar 2022, 18-39 y | -1.87015 | 1.003109 | 3.7% | 0.15 [0.02, 1.10] |
| Zarifkar 2022, 40-59 y | -0.69315 | 0.216147 | 24.3% | 0.50 [0.33, 0.76] |
| Zarifkar 2022, 60-79 y | -0.35667 | 0.188849 | 26.0% | 0.70 [0.48, 1.01] |
| **Subtotal (95% CI)** | | | **100.0%** | **0.78 [0.53, 1.17]** |
| Heterogeneity: Tau² = 0.12; Chi² = 17.71, df = 4 (P = 0.001); I² = 77% | | | | |
| Test for overall effect: Z = 1.20 (P = 0.23) | | | | |
| **12.3.3 Ischemic stroke (outpatients/mixed, ≥ 65 y)** | | | | |
| Cohen 2022 | 0.652325 | 0.080672 | 20.2% | 1.92 [1.64, 2.25] |
| Taquet 2022 | 0.070552 | 0.014475 | 21.8% | 1.07 [1.04, 1.10] |
| Yang 2024 | -0.01525 | 0.059333 | 20.9% | 0.98 [0.88, 1.11] |
| Zarifkar 2022, 60-79 y | -0.51083 | 0.119439 | 18.6% | 0.60 [0.47, 0.76] |
| Zarifkar 2022, 80+ y | -0.35667 | 0.119899 | 18.5% | 0.70 [0.55, 0.89] |
| **Subtotal (95% CI)** | | | **100.0%** | **0.98 [0.76, 1.28]** |
| Heterogeneity: Tau² = 0.08; Chi² = 90.48, df = 4 (P < 0.00001); I² = 96% | | | | |
| Test for overall effect: Z = 0.13 (P = 0.90) | | | | |
| **12.3.4 Ischemic stroke (inpatients, 18-64 y)** | | | | |
| Daugherty 2021 | 1.472243 | 0.185724 | 21.9% | 4.36 [3.03, 6.27] |
| Ho 2021 | 0.482426 | 0.61076 | 19.0% | 1.62 [0.49, 5.36] |
| Zarifkar 2022, 18-39 y | 0.10499 | 0.875049 | 16.5% | 1.11 [0.20, 6.17] |
| Zarifkar 2022, 40-59 y | -1.60944 | 0.353647 | 21.0% | 0.20 [0.10, 0.40] |
| Zarifkar 2022, 60-79 y | -0.69315 | 0.258592 | 21.6% | 0.50 [0.30, 0.83] |
| **Subtotal (95% CI)** | | | **100.0%** | **0.94 [0.24, 3.76]** |
| Heterogeneity: Tau² = 2.24; Chi² = 83.58, df = 4 (P < 0.00001); I² = 95% | | | | |
| Test for overall effect: Z = 0.08 (P = 0.94) | | | | |
| **12.3.5 Ischemic stroke (inpatients, ≥ 65 y)** | | | | |
| Cohen 2022 | 1.135683 | 0.064568 | 21.3% | 3.11 [2.74, 3.53] |
| Ho 2021, 66-80 y | -0.41552 | 0.518223 | 18.0% | 0.66 [0.24, 1.82] |
| Ho 2021, 80+ y | -0.51083 | 0.462342 | 18.6% | 0.60 [0.24, 1.48] |
| Zarifkar 2022, 60-79 y | -0.69315 | 0.119439 | 21.1% | 0.50 [0.40, 0.63] |
| Zarifkar 2022, 80+ y | -0.69315 | 0.142759 | 21.1% | 0.50 [0.38, 0.66] |
| **Subtotal (95% CI)** | | | **100.0%** | **0.80 [0.27, 2.36]** |
| Heterogeneity: Tau² = 1.42; Chi² = 274.46, df = 4 (P < 0.00001); I² = 99% | | | | |
| Test for overall effect: Z = 0.40 (P = 0.69) | | | | |

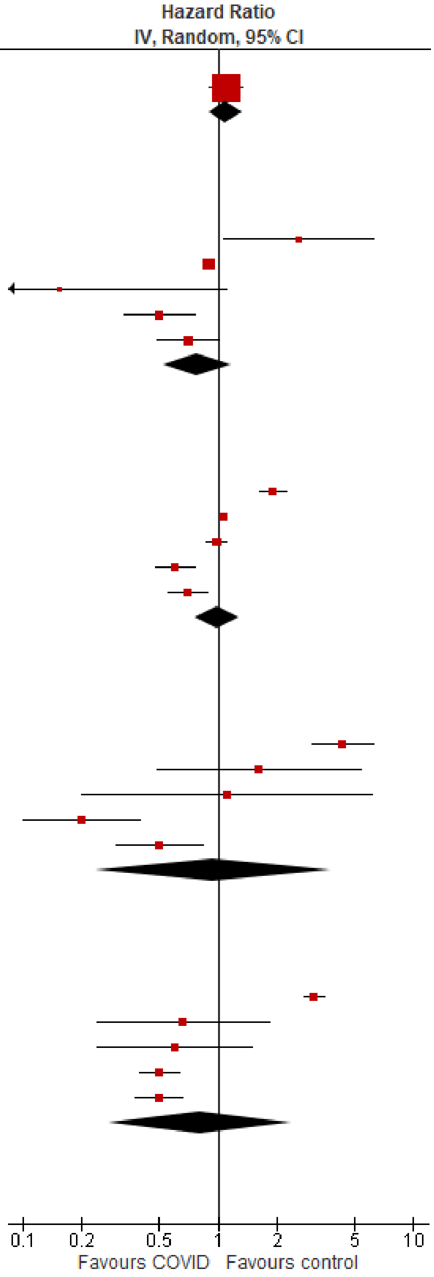

Test for subgroup differences: Chi² = 2.40, df = 4 (P = 0.66), I² = 0%

#### Transient ischemic attack

| Study or Subgroup | log[Hazard Ratio] | SE | Weight | Hazard Ratio IV, Random, 95% CI |
|---|---|---|---|---|
| **12.4.1 Transient ischemic attack (outpatients/mixed, 18-64 y)** | | | | |
| Velasquez Garcia 2024 | -0.12783 | 0.250042 | 18.9% | 0.88 [0.54, 1.44] |
| Wang 2022b, 20-44y | 0.499562 | 0.135602 | 35.8% | 1.65 [1.26, 2.15] |
| Wang 2022b, 45-64y | 0.291923 | 0.089229 | 45.4% | 1.34 [1.12, 1.59] |
| **Subtotal (95% CI)** | | | **100.0%** | **1.33 [1.03, 1.73]** |
| Heterogeneity: Tau² = 0.03; Chi² = 5.06, df = 2 (P = 0.08); I² = 60% | | | | |
| Test for overall effect: Z = 2.16 (P = 0.03) | | | | |
| **12.4.2 Transient ischemic attack (outpatients/mixed, ≥ 65 y)** | | | | |
| Wan 2023a, UK cohort | 0.530628 | 0.256109 | 16.9% | 1.70 [1.03, 2.81] |
| Wang 2022b | 0.489193 | 0.080804 | 38.9% | 1.63 [1.39, 1.91] |
| Yang 2024 | 0.188241 | 0.034017 | 44.2% | 1.21 [1.13, 1.29] |
| **Subtotal (95% CI)** | | | **100.0%** | **1.44 [1.11, 1.87]** |
| Heterogeneity: Tau² = 0.04; Chi² = 13.11, df = 2 (P = 0.001); I² = 85% | | | | |
| Test for overall effect: Z = 2.73 (P = 0.006) | | | | |

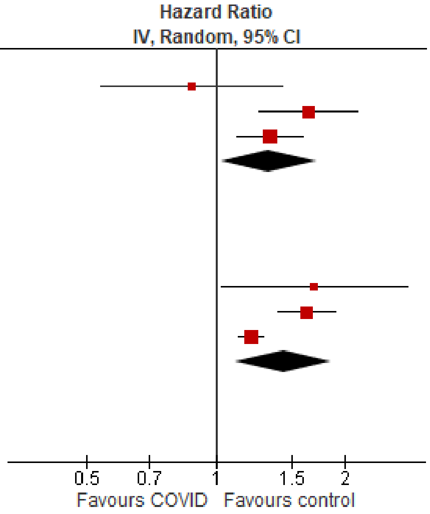

Test for subgroup differences: Chi² = 0.16, df = 1 (P = 0.68), I² = 0%

### Stratified analyses results for a systematic review update examining new diagnoses and exacerbations of chronic conditions after SARS-CoV-2 infection

#### Appendix D. Stratified analysis of new diagnoses of chronic conditions after SARS-CoV-2 infection: studies requiring case confirmation (i.e., documented positive test) vs. others.

Summary

For most outcomes, heterogeneity based on case confirmation methods was not observed. Only outcomes with ≥2 studies contributing to each subgroup are presented.

For the following outcomes, studies of confirmed SARS-CoV-2 cases reported statistically significant smaller associations leading to a difference in conclusions (yellow shading) from studies that did not require case confirmation (i.e., one group indicates little to no association while the other group indicates a small to moderate or large association).

- Overall cardiovascular disorders in outpatients/mixed patients ≥65 y
- Overall mental illness in outpatients/mixed patients ≥65y
- Overall mental illness in inpatients ≥65 y
- Overall sleep disorders in outpatients/mixed patients ≥65y

For two outcomes, a statistically significant difference in association was observed, but did not lead to a difference in conclusions (blue shading) between studies of confirmed SARS-CoV-2 cases compared with studies not based on confirmed cases.

- Acute coronary disease in outpatients/mixed patients ≥65 y
- Depression/mood disorders in outpatients/mixed patients 18-64 y

| Population | Subgroup<br>No. Studies; weight | HR (95% CI) | $I^2$ | Between-groups P-value |
|---|---|---|---|---|
| **Cardiovascular disorders, overall** | | | | |
| Outpatients/mixed, 18-64 y | Overall, 12 studies | 1.32 (1.12 to 1.55) | 92% | P=0.46 |
| | Tested positive<br>9 studies; 83.4% | 1.27 (1.08 to 1.50) | 89% | |
| | Others<br>3 studies;16.6% | 1.62 (0.87 to 3.02) | 97% | |
| Outpatients/mixed, ≥65 y | Overall, 10 studies | 1.33 (1.14 to 1.54) | 96% | P=0.04<br>Difference in conclusions |
| | Tested positive<br>7 studies; 72.9% | 1.23 (1.03 to 1.48) | 97% | |
| | Others<br>3 studies; 27.1% | 1.62 (1.35 to 1.95) | 61% | |
| Inpatients, 18-64 y | Overall, 6 studies | 2.23 (1.29 to 3.87) | 96% | P=0.49 |
| | Tested positive<br>3 studies; 54.1% | 1.78 (0.58 to 5.48) | 95% | |
| | Others<br>3 studies; 45.9% | 2.90 (1.27 to 6.64) | 98% | |
| **Cardiovascular disorders, acute coronary disease** | | | | |
| Outpatients/mixed, 18-64 y | Overall, 9 studies | 1.43 (1.14 to 1.81) | 86% | P=0.97 |
| | Tested positive<br>6 studies; 74.2% | 1.43 (1.12 to 1.84) | 84% | |
| | Others<br>3 studies; 25.8% | 1.45 (0.72 to 2.93) | 92% | |
| Outpatients/mixed, ≥65 y | Overall, 7 studies | 1.55 (1.27 to 1.89) | 94% | P=0.02 |

| Population | Subgroup<br>No. Studies; weight | HR (95% CI) | $I^2$ | Between-groups P-value |
|---|---|---|---|---|
| | Tested positive<br>4 studies; 63.3% | 1.33 (1.05 to 1.67) | 96% | No difference in conclusions |
| | Others<br>3 studies; 36.7% | 2.02 (1.55 to 2.63) | 61% | |
| **Cardiovascular disorders, arrhythmias/dysrhythmias** | | | | |
| Outpatients/mixed, 18-64 y | Overall, 8 studies | 1.56 (1.38 to 1.77) | 87% | P=0.57 |
| | Tested positive<br>5 studies; 71.4% | 1.55 (1.42 to 1.70) | 66% | |
| | Others<br>3 studies; 28.6% | 1.84 (1.02 to 3.34) | 95% | |
| Outpatients/mixed, ≥65 y | Overall, 8 studies | 1.43 (1.18 to 1.73) | 97% | P=0.96 |
| | Tested positive<br>5 studies; 61.4% | 1.44 (1.12 to 1.84) | 98% | |
| | Others<br>3 studies; 38.6% | 1.42 (1.01 to 2.00) | 91% | |
| **Cardiovascular disorders, heart failure** | | | | |
| Outpatients/mixed, 18-64 y | Overall, 7 studies | 1.81 (1.40 to 2.34) | 94% | P=0.92 |
| | Tested positive<br>4 studies; 69.8% | 1.79 (1.31 to 2.43) | 96% | |
| | Others<br>3 studies; 30.2% | 1.86 (0.94 to 3.68) | 85% | |
| Outpatients/mixed, ≥65 y | Overall, 6 studies | 1.61 (1.22 to 2.12) | 98% | P=0.29 |
| | Tested positive<br>3 studies; 53.0% | 1.45 (0.98 to 2.17) | 99% | |
| | Others<br>3 studies; 47.0% | 1.82 (1.62 to 2.04) | 0% | |
| **Diabetes, overall** | | | | |
| Outpatients/mixed, <18y | Overall, 6 studies | 1.10 (0.95 to 1.28) | 59% | P=0.47 |
| | Tested positive<br>4 studies; 70.5% | 1.09 (0.87 to 1.37) | 63% | |
| | Others<br>2 studies; 29.5% | 1.19 (1.12 to 1.26) | 0% | |
| Outpatients/mixed, 18-64 y | Overall, 10 studies | 1.53 (1.29 to 1.81) | 98% | P=0.18 |
| | Tested positive<br>5 studies; 41.8% | 1.75 (1.21 to 2.53) | 99% | |
| | Others<br>5 studies; 58.2% | 1.34 (1.20 to 1.50) | 87% | |
| Outpatients/mixed, ≥65 y | Overall, 6 studies | 1.43 (1.32 to 1.54) | 64% | P=1.00 |
| | Tested positive<br>4 studies; 44.4% | 1.45 (1.39 to 1.50) | 0% | |
| | Others<br>2 studies; 55.6% | 1.44 (1.23 to 1.70) | 73% | |
| **Mental illness, overall** | | | | |
| Outpatients/mixed, <18y | Overall, 11 studies | 1.01 (0.88 to 1.16) | 99% | P=0.69 |
| | Tested positive<br>8 studies; 63.7% | 0.99 (0.86 to 1.14) | 94% | |
| | Others<br>3 studies; 36.3% | 1.05 (0.82 to 1.34) | 100% | |
| Outpatients/mixed, 18-64 y | Overall, 12 studies | 1.24 (1.13 to 1.36) | 99% | P=0.98 |
| | Tested positive<br>8 studies; 55.3% | 1.24 (1.10 to 1.40) | 99% | |
| | Others<br>4 studies; 44.7% | 1.24 (1.07 to 1.42) | 90% | |
| Outpatients/mixed, ≥65 y | Overall, 14 studies | 1.25 (1.11 to 1.41) | 96% | P=0.0007<br>Difference in conclusions |
| | Tested positive<br>8 studies; 60.6% | 1.06 (0.93 to 1.21) | 96% | |
| | Others<br>6 studies; 39.4% | 1.57 (1.30 to 1.89) | 85% | |
| Inpatients, ≥65 y | Overall, 4 studies | 1.16 (0.74 to 1.79) | 97% | P<0.00001 |

| Population | Subgroup<br>No. Studies; weight | HR (95% CI) | I2 | Between-groups P-value |
|---|---|---|---|---|
| | Tested positive<br>2 studies; 68.2% | 0.88 (0.75 to 1.05) | 37% | Difference in conclusions |
| | Others<br>2 studies; 31.8% | 2.25 (2.09 to 2.42) | 0% | |
| **Mental illness, anxiety/anxiety disorders** | | | | |
| Outpatients/mixed, 18-64 y | Overall, 6 studies | 1.08 (0.96 to 1.22) | 92% | P=0.78 |
| | Tested positive<br>4 studies; 53.1% | 1.08 (0.97 to 1.21) | 82% | |
| | Others<br>2 studies; 46.9% | 1.04 (0.83 to 1.32) | 83% | |
| Outpatients/mixed, ≥65 y | Overall, 6 studies | 0.84 (0.66 to 1.08) | 99% | P=0.13 |
| | Tested positive<br>4 studies; 65.7% | 0.72 (0.43 to 1.19) | 99% | |
| | Others<br>2 studies; 34.3% | 1.17 (0.80 to 1.72) | 85% | |
| **Mental illness, depression & mood disorders** | | | | |
| Outpatients/mixed, <18y | Overall, 6 studies | 1.37 (0.84 to 2.23) | 100% | P=0.24 |
| | Tested positive<br>4 studies; 56.6% | 0.98 (0.71 to 1.35) | 76% | |
| | Others<br>2 studies; 43.4% | 2.09 (0.61 to 7.23) | 100% | |
| Outpatients/mixed, 18-64 y | Overall, 5 studies | 1.11 (1.04 to 1.19) | 73% | P=0.002<br>No difference in conclusions |
| | Tested positive<br>2 studies; 22.2% | 1.01 (1.00 to 1.02) | 0% | |
| | Others<br>3 studies; 77.8% | 1.13 (1.05 to 1.22) | 56% | |
| Outpatients/mixed, ≥65 y | Overall, 5 studies | 1.07 (1.05 to 1.09) | 0% | P=0.06 |
| | Tested positive<br>3 studies; 97.8% | 1.06 (1.05 to 1.08) | 0% | |
| | Others<br>2 studies; 2.2% | 1.20 (1.06 to 1.36) | 0% | |
| **Neurological disorders, overall** | | | | |
| Outpatients/mixed, ≥65 y | Overall, 13 studies | 1.49 (1.10 to 2.01) | 99% | P=0.64 |
| | Tested positive<br>10 studies;83.3% | 1.52 (1.09 to 2.11) | 99% | |
| | Others<br>3 studies; 16.7% | 1.32 (0.84 to 2.09) | 78% | |
| **Neurological disorders, dementia/mild cognitive impairment** | | | | |
| Outpatients/mixed, ≥65 y | Overall, 11 studies | 1.42 (1.23 to 1.63) | 95% | P=0.97 |
| | Tested positive<br>8 studies; 80.3% | 1.41 (1.21 to 1.64) | 95% | |
| | Others<br>3 studies; 19.7% | 1.39 (0.76 to 2.55) | 95% | |
| **Respiratory disorders, overall** | | | | |
| Outpatients/mixed, 18-64 y | Overall, 6 studies | 1.89 (1.42 to 2.51) | 96% | P=0.38 |
| | Tested positive<br>4 studies; 77.9% | 1.63 (1.20 to 2.21) | 96% | |
| | Others<br>2 studies; 22.1% | 3.62 (0.63 to 20.70) | 98% | |
| Outpatients/mixed, ≥65 y | Overall, 5 studies | 1.83 (1.31 to 2.56) | 94% | P=0.27 |
| | Tested positive<br>3 studies; 63.3% | 1.57 (1.13 to 2.16) | 88% | |
| | Others<br>2 studies; 36.7% | 2.48 (1.16 to 5.28) | 96% | |
| **Sleep disorders, overall** | | | | |
| Outpatients/mixed, ≥65 y | Overall, 8 studies | 1.15 (0.98 to 1.35) | 93% | P=0.01<br>Difference in conclusions |
| | Tested positive<br>6 studies; 75.5% | 1.07 (0.88 to 1.29) | 95% | |

| Population | Subgroup<br>No. Studies; weight | HR (95% CI) | $I^2$ | Between-groups P-value |
|---|---|---|---|---|
| | Others<br>2 studies; 24.5% | 1.47 (1.26 to 1.71) | 0% | |
| **Stroke, overall** | | | | |
| Outpatients/mixed, 18-64 y | Overall, 14 studies | 1.11 (0.95 to 1.31) | 95% | P=0.52 |
| | Tested positive<br>11 studies; 85.7% | 1.08 (0.90 to 1.28) | 96% | |
| | Others<br>3 studies; 14.3% | 1.44 (0.60 to 3.47) | 94% | |
| Outpatients/mixed, ≥65 y | Overall, 12 studies | 1.17 (0.99 to 1.39) | 97% | P=0.07 |
| | Tested positive<br>9 studies; 79.2% | 1.09 (0.91 to 1.31) | 98% | |
| | Others<br>3 studies; 20.8% | 1.55 (1.11 to 2.17) | 72% | |

### Forest Plots for stratified analyses: SARS-CoV-2 infection confirmation (i.e., documented positive test) vs. others.

#### *Cardiovascular disorders, overall*

##### Outpatient/mixed, 18-64 y

| Study or Subgroup | log[Hazard Ratio] | SE | Weight | Hazard Ratio IV, Random, 95% CI |
|---|---|---|---|---|
| **1.1.1 Tested +ve** | | | | |
| Katsoularis 2023, 60+ y | 0.227813 | 0.25172 | 4.0% | 1.26 [0.77, 2.06] |
| Katsoularis 2023, 60 y & under | 0.348127 | 0.183357 | 4.9% | 1.42 [0.99, 2.03] |
| Lam 2023 | 0.385263 | 0.163468 | 5.1% | 1.47 [1.07, 2.03] |
| Lim 2023 | 0.690556 | 0.220029 | 4.4% | 1.99 [1.30, 3.07] |
| Mizrahi 2023, 19-40 y | 0.074487 | 0.201878 | 4.6% | 1.08 [0.73, 1.60] |
| Mizrahi 2023, 41-60 y | 0.044255 | 0.158501 | 5.2% | 1.05 [0.77, 1.43] |
| Mizrahi 2023, 60+ y | -0.13266 | 0.359911 | 2.9% | 0.88 [0.43, 1.77] |
| Valasquez Garcia 2024 | 0.329304 | 0.1379 | 5.4% | 1.39 [1.06, 1.82] |
| Wang 2022b, 20-44y | 0.527832 | 0.083008 | 6.0% | 1.70 [1.44, 1.99] |
| Wang 2022b, 45-64y | 0.721112 | 0.045725 | 6.3% | 2.06 [1.88, 2.25] |
| Xie 2022b | 0.51721819 | 0.043625 | 6.3% | 1.68 [1.54, 1.83] |
| Zang 2023, INSIGHT cohort | 0.110557 | 0.063558 | 6.2% | 1.12 [0.99, 1.27] |
| Zappacosta 2022, 18-30 y | 0.350657 | 0.37779 | 2.8% | 1.42 [0.68, 2.98] |
| Zappacosta 2022, 31-40 y | 0.29267 | 0.254071 | 4.0% | 1.34 [0.81, 2.20] |
| Zappacosta 2022, 41-50 y | -0.57982 | 0.168459 | 5.1% | 0.56 [0.40, 0.78] |
| Zappacosta 2022, 51-60 y | -0.08338 | 0.131688 | 5.5% | 0.92 [0.71, 1.19] |
| Zappacosta 2022, 61-70 y | -0.0202 | 0.186986 | 4.8% | 0.98 [0.68, 1.41] |
| **Subtotal (95% CI)** | | | **83.4%** | **1.27 [1.08, 1.50]** |
| Heterogeneity: Tau² = 0.09; Chi² = 142.76, df = 16 (P < 0.00001); I² = 89% | | | | |
| Test for overall effect: Z = 2.86 (P = 0.004) | | | | |
| **1.1.2 Other** | | | | |
| Batistoni 2024 | 0.836308 | 0.216484 | 4.5% | 2.31 [1.51, 3.53] |
| Daugherty 2021 | 0.715483 | 0.094995 | 5.9% | 2.05 [1.70, 2.46] |
| Rezel-Potts 2022 | -0.05033 | 0.054413 | 6.2% | 0.95 [0.85, 1.06] |
| **Subtotal (95% CI)** | | | **16.6%** | **1.62 [0.87, 3.02]** |
| Heterogeneity: Tau² = 0.28; Chi² = 58.84, df = 2 (P < 0.00001); I² = 97% | | | | |
| Test for overall effect: Z = 1.52 (P = 0.13) | | | | |
| **Total (95% CI)** | | | **100.0%** | **1.32 [1.12, 1.55]** |
| Heterogeneity: Tau² = 0.11; Chi² = 227.01, df = 19 (P < 0.00001); I² = 92% | | | | |
| Test for overall effect: Z = 3.30 (P = 0.0010) | | | | |
| Test for subgroup differences: Chi² = 0.54, df = 1 (P = 0.46), I² = 0% | | | | |

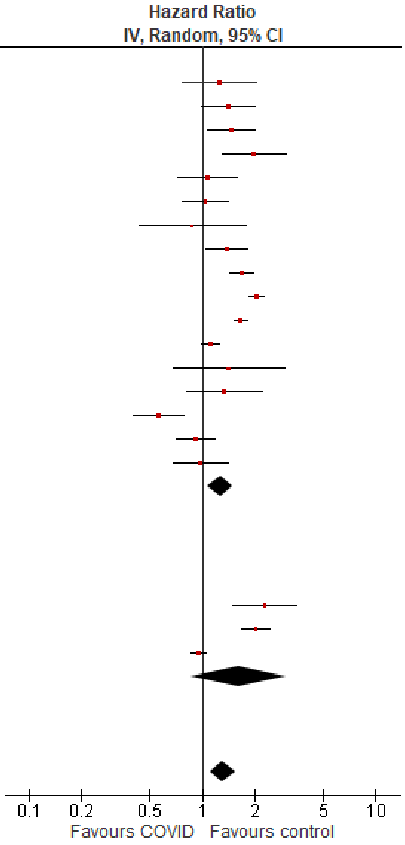

##### Outpatient/mixed ≥65 y

| Study or Subgroup | log[Hazard Ratio] | SE | Weight | Hazard Ratio IV, Random, 95% CI |
|---|---|---|---|---|
| **1.2.1 Tested +ve** | | | | |
| Katsoularis 2023, 60+ y | 0.22781 | 0.25172 | 5.1% | 1.26 [0.77, 2.06] |
| Mizrahi 2023, 60+ y | -0.1327 | 0.15708 | 7.6% | 0.88 [0.64, 1.19] |
| Wang 2022b | 0.715354 | 0.043015 | 10.6% | 2.04 [1.88, 2.22] |
| Xie 2022b | 0.494319 | 0.027525 | 10.8% | 1.64 [1.55, 1.73] |
| Yang 2024 | 0.21486 | 0.01727 | 10.9% | 1.24 [1.20, 1.28] |
| Zang 2023, INSIGHT cohort | 0.2389 | 0.0423 | 10.6% | 1.27 [1.17, 1.38] |
| Zappacosta 2022, 61-70 y | -0.0202 | 0.15267 | 7.7% | 0.98 [0.73, 1.32] |
| Zappacosta 2022, 70+ y | -0.2485 | 0.08638 | 9.6% | 0.78 [0.66, 0.92] |
| **Subtotal (95% CI)** | | | **72.9%** | **1.23 [1.03, 1.48]** |
| Heterogeneity: Tau² = 0.06; Chi² = 221.35, df = 7 (P < 0.00001); I² = 97% | | | | |
| Test for overall effect: Z = 2.27 (P = 0.02) | | | | |
| **1.2.2 Other** | | | | |
| Batistoni 2024 | 0.43628 | 0.1502 | 7.8% | 1.55 [1.15, 2.08] |
| Cohen 2022 | 0.61252 | 0.0677 | 10.1% | 1.85 [1.62, 2.11] |
| Wan 2023a, UK cohort | 0.34905 | 0.09979 | 9.3% | 1.42 [1.17, 1.72] |
| **Subtotal (95% CI)** | | | **27.1%** | **1.62 [1.35, 1.95]** |
| Heterogeneity: Tau² = 0.02; Chi² = 5.11, df = 2 (P = 0.08); I² = 61% | | | | |
| Test for overall effect: Z = 5.16 (P < 0.00001) | | | | |
| **Total (95% CI)** | | | **100.0%** | **1.33 [1.14, 1.54]** |
| Heterogeneity: Tau² = 0.05; Chi² = 241.39, df = 10 (P < 0.00001); I² = 96% | | | | |
| Test for overall effect: Z = 3.64 (P = 0.0003) | | | | |
| Test for subgroup differences: Chi² = 4.28, df = 1 (P = 0.04), I² = 76.6% | | | | |

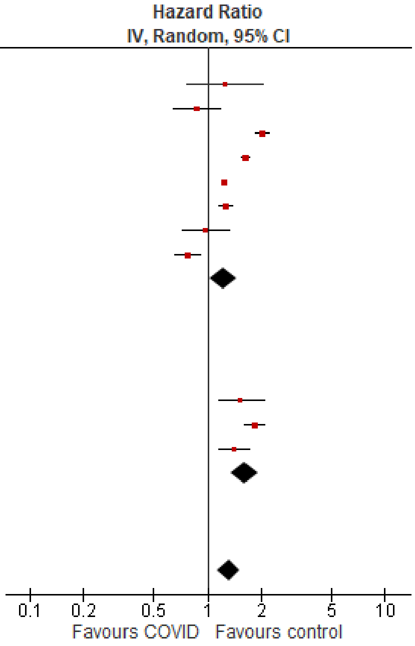

Inpatient, 18-64 y

| Study or Subgroup | log[Hazard Ratio] | SE | Weight | Hazard Ratio IV, Random, 95% CI |
|---|---|---|---|---|
| **1.3.1 Tested +ve** | | | | |
| Ho 2021 | 0.182322 | 0.725333 | 7.9% | 1.20 [0.29, 4.97] |
| Lee 2023, 18-39 y | 0.833253 | 0.228239 | 15.2% | 2.30 [1.47, 3.60] |
| Lee 2023, 40-64 y | -0.45089 | 0.132605 | 16.3% | 0.64 [0.49, 0.83] |
| Valasquez Garcia 2024 | 1.684545 | 0.267412 | 14.7% | 5.39 [3.19, 9.10] |
| **Subtotal (95% CI)** | | | **54.1%** | **1.78 [0.58, 5.48]** |
| Heterogeneity: Tau² = 1.17; Chi² = 62.43, df = 3 (P < 0.00001); I² = 95% | | | | |
| Test for overall effect: Z = 1.01 (P = 0.31) | | | | |
| **1.3.2 Other** | | | | |
| Ayoubkhani 2021, 18-69 y | 1.458615 | 0.408163 | 12.5% | 4.30 [1.93, 9.57] |
| Daugherty 2021 | 1.42149 | 0.10551 | 16.5% | 4.14 [3.37, 5.10] |
| Salah 2022 | 0.425268 | 0.033177 | 16.9% | 1.53 [1.43, 1.63] |
| **Subtotal (95% CI)** | | | **45.9%** | **2.90 [1.27, 6.64]** |
| Heterogeneity: Tau² = 0.48; Chi² = 86.44, df = 2 (P < 0.00001); I² = 98% | | | | |
| Test for overall effect: Z = 2.53 (P = 0.01) | | | | |
| **Total (95% CI)** | | | **100.0%** | **2.23 [1.29, 3.87]** |
| Heterogeneity: Tau² = 0.46; Chi² = 160.12, df = 6 (P < 0.00001); I² = 96% | | | | |
| Test for overall effect: Z = 2.87 (P = 0.004) | | | | |
| Test for subgroup differences: Chi² = 0.47, df = 1 (P = 0.49), I² = 0% | | | | |

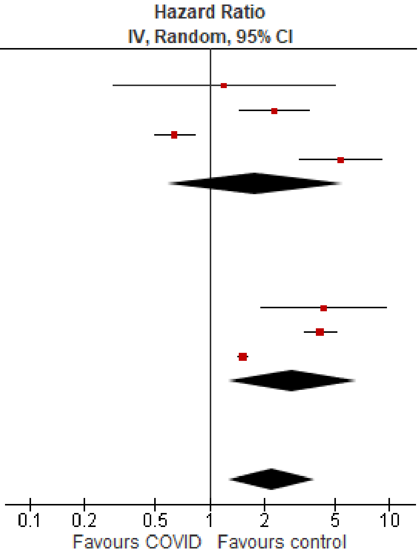

*Cardiovascular disorders, acute coronary disease*

Outpatient/mixed, 18-64 y

| Study or Subgroup | log[Hazard Ratio] | SE | Weight | Hazard Ratio IV, Random, 95% CI |
|---|---|---|---|---|
| **2.1.1 Tested +ve** | | | | |
| Lam 2023 | 0.274072 | 0.188926 | 8.6% | 1.32 [0.91, 1.90] |
| Lim 2023 | 1.652497 | 0.377588 | 5.2% | 5.22 [2.49, 10.94] |
| Mizrahi 2023, 19-40 y | 0.250003 | 0.326955 | 6.0% | 1.28 [0.68, 2.44] |
| Mizrahi 2023, 41-60 y | -0.33604 | 0.147626 | 9.4% | 0.71 [0.54, 0.95] |
| Mizrahi 2023, 60+ y | -0.28462 | 0.384812 | 5.1% | 0.75 [0.35, 1.60] |
| Valasquez Garcia 2024 | 0.329304 | 0.1379 | 9.6% | 1.39 [1.06, 1.82] |
| Wang 2022b, 20-44y | 0.372339 | 0.138893 | 9.5% | 1.45 [1.11, 1.91] |
| Wang 2022b, 45-64y | 0.662593 | 0.086399 | 10.4% | 1.94 [1.64, 2.30] |
| Xie 2022b | 0.506818 | 0.070291 | 10.6% | 1.66 [1.45, 1.91] |
| **Subtotal (95% CI)** | | | **74.2%** | **1.43 [1.12, 1.84]** |
| Heterogeneity: Tau² = 0.10; Chi² = 50.99, df = 8 (P < 0.00001); I² = 84% | | | | |
| Test for overall effect: Z = 2.83 (P = 0.005) | | | | |
| **2.1.2 Other** | | | | |
| Batistoni 2024 | 0.530628 | 0.281988 | 6.8% | 1.70 [0.98, 2.95] |
| Daugherty 2021 | 0.802002 | 0.156599 | 9.2% | 2.23 [1.64, 3.03] |
| Rezel-Potts 2022 | -0.17732 | 0.123467 | 9.8% | 0.84 [0.66, 1.07] |
| **Subtotal (95% CI)** | | | **25.8%** | **1.45 [0.72, 2.93]** |
| Heterogeneity: Tau² = 0.35; Chi² = 25.36, df = 2 (P < 0.00001); I² = 92% | | | | |
| Test for overall effect: Z = 1.04 (P = 0.30) | | | | |
| **Total (95% CI)** | | | **100.0%** | **1.43 [1.14, 1.81]** |
| Heterogeneity: Tau² = 0.13; Chi² = 80.19, df = 11 (P < 0.00001); I² = 86% | | | | |
| Test for overall effect: Z = 3.04 (P = 0.002) | | | | |
| Test for subgroup differences: Chi² = 0.00, df = 1 (P = 0.97), I² = 0% | | | | |

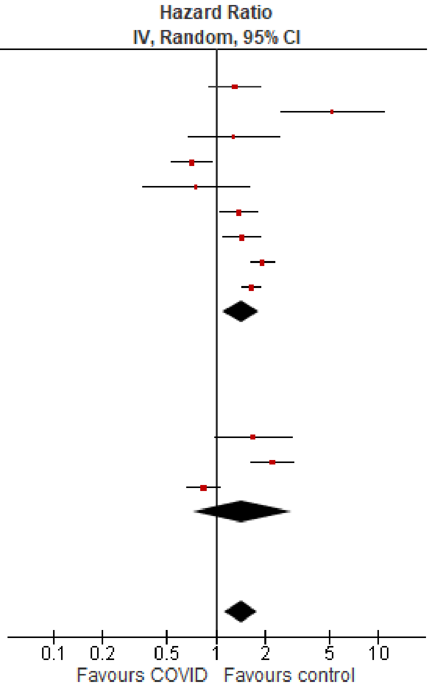

Outpatient/mixed ≥65 y

| Study or Subgroup | log[Hazard Ratio] | SE | Weight | Hazard Ratio IV, Random, 95% CI |
|---|---|---|---|---|
| **2.2.1 Tested +ve** | | | | |
| Mizrahi 2023, 60+ y | -0.2846 | 0.16795 | 12.0% | 0.75 [0.54, 1.05] |
| Wang 2022b | 0.589343 | 0.084561 | 16.0% | 1.80 [1.53, 2.13] |
| Xie 2022b | 0.470004 | 0.031731 | 17.7% | 1.60 [1.50, 1.70] |
| Yang 2024 | 0.18462 | 0.02869 | 17.7% | 1.20 [1.14, 1.27] |
| **Subtotal (95% CI)** | | | **63.3%** | **1.33 [1.05, 1.67]** |
| Heterogeneity: Tau² = 0.05; Chi² = 67.72, df = 3 (P < 0.00001); I² = 96% | | | | |
| Test for overall effect: Z = 2.39 (P = 0.02) | | | | |
| **2.2.2 Other** | | | | |
| Batistoni 2024 | 1.1474 | 0.29663 | 7.0% | 3.15 [1.76, 5.63] |
| Cohen 2022 | 0.732368 | 0.088939 | 15.8% | 2.08 [1.75, 2.48] |
| Wan 2023a, UK cohort | 0.48734 | 0.12869 | 13.9% | 1.63 [1.27, 2.10] |
| **Subtotal (95% CI)** | | | **36.7%** | **2.02 [1.55, 2.63]** |
| Heterogeneity: Tau² = 0.03; Chi² = 5.07, df = 2 (P = 0.08); I² = 61% | | | | |
| Test for overall effect: Z = 5.20 (P < 0.00001) | | | | |
| **Total (95% CI)** | | | **100.0%** | **1.55 [1.27, 1.89]** |
| Heterogeneity: Tau² = 0.06; Chi² = 96.66, df = 6 (P < 0.00001); I² = 94% | | | | |
| Test for overall effect: Z = 4.37 (P < 0.0001) | | | | |
| Test for subgroup differences: Chi² = 5.51, df = 1 (P = 0.02), I² = 81.8% | | | | |

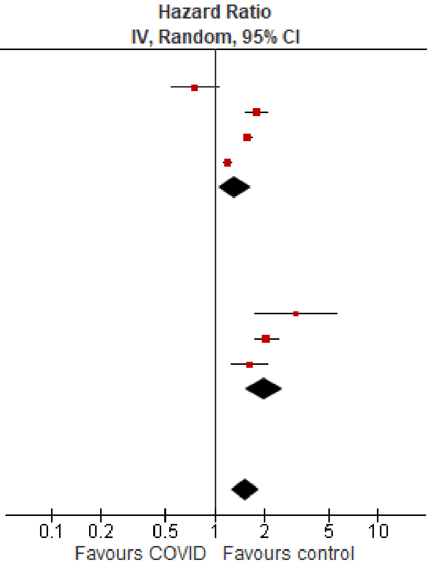

*Cardiovascular disorders, arrhythmias/dysrhythmia*

Outpatient/mixed, 18-64 y

| Study or Subgroup | log[Hazard Ratio] | SE | Weight | Hazard Ratio IV, Random, 95% CI |
|---|---|---|---|---|
| **3.1.1 Tested +ve** | | | | |
| Katsoularis 2023, 60+ y | 0.227813 | 0.25172 | 4.4% | 1.26 [0.77, 2.06] |
| Katsoularis 2023, 60 y & under | 0.348127 | 0.183357 | 6.5% | 1.42 [0.99, 2.03] |
| Lam 2023 | 0.285179 | 0.217401 | 5.4% | 1.33 [0.87, 2.04] |
| Mizrahi 2023, 19-40 y | -0.07429 | 0.223295 | 5.2% | 0.93 [0.60, 1.44] |
| Mizrahi 2023, 41-60 y | 0.140799 | 0.174525 | 6.9% | 1.15 [0.82, 1.62] |
| Mizrahi 2023, 60+ y | 0.007484 | 0.410256 | 2.1% | 1.01 [0.45, 2.25] |
| Wang 2022b, 20-44y | 0.445361 | 0.038156 | 13.4% | 1.56 [1.45, 1.68] |
| Wang 2022b, 45-64y | 0.577963 | 0.030978 | 13.7% | 1.78 [1.68, 1.89] |
| Xie 2022b | 0.518794 | 0.027359 | 13.8% | 1.68 [1.59, 1.77] |
| **Subtotal (95% CI)** | | | **71.4%** | **1.55 [1.42, 1.70]** |
| Heterogeneity: Tau² = 0.01; Chi² = 23.71, df = 8 (P = 0.003); I² = 66% | | | | |
| Test for overall effect: Z = 9.46 (P < 0.00001) | | | | |
| **3.1.2 Other** | | | | |
| Batistoni 2024 | 0.916291 | 0.166762 | 7.2% | 2.50 [1.80, 3.47] |
| Daugherty 2021 | 0.916291 | 0.061931 | 12.5% | 2.50 [2.21, 2.82] |
| Rezel-Potts 2022 | 0.001457 | 0.129349 | 9.0% | 1.00 [0.78, 1.29] |
| **Subtotal (95% CI)** | | | **28.6%** | **1.84 [1.02, 3.34]** |
| Heterogeneity: Tau² = 0.26; Chi² = 41.63, df = 2 (P < 0.00001); I² = 95% | | | | |
| Test for overall effect: Z = 2.02 (P = 0.04) | | | | |
| **Total (95% CI)** | | | **100.0%** | **1.56 [1.38, 1.77]** |
| Heterogeneity: Tau² = 0.03; Chi² = 85.99, df = 11 (P < 0.00001); I² = 87% | | | | |
| Test for overall effect: Z = 6.95 (P < 0.00001) | | | | |
| Test for subgroup differences: Chi² = 0.32, df = 1 (P = 0.57), I² = 0% | | | | |

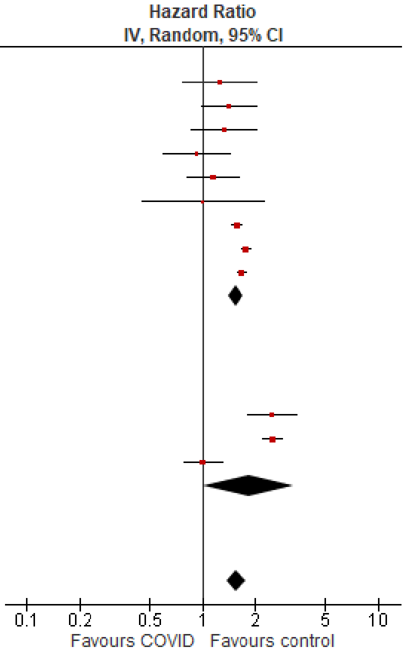

Outpatient/mixed ≥65 y

| Study or Subgroup | log[Hazard Ratio] | SE | Weight | Hazard Ratio IV, Random, 95% CI |
|---|---|---|---|---|
| **3.2.1 Tested +ve** | | | | |
| Katsoularis 2023, 60+ y | 0.22781 | 0.25172 | 7.4% | 1.26 [0.77, 2.06] |
| Mizrahi 2023, 60+ y | 0.00748 | 0.17905 | 9.9% | 1.01 [0.71, 1.43] |
| Wang 2022b | 0.665519 | 0.03084304 | 14.6% | 1.95 [1.83, 2.07] |
| Xie 2022b | 0.518794 | 0.031834 | 14.6% | 1.68 [1.58, 1.79] |
| Yang 2024 | 0.21486 | 0.01365 | 14.8% | 1.24 [1.21, 1.27] |
| **Subtotal (95% CI)** | | | **61.4%** | **1.44 [1.12, 1.84]** |
| Heterogeneity: Tau² = 0.07; Chi² = 226.97, df = 4 (P < 0.00001); I² = 98% | | | | |
| Test for overall effect: Z = 2.84 (P = 0.005) | | | | |
| **3.2.2 Other** | | | | |
| Batistoni 2024 | 0.3001 | 0.08647 | 13.3% | 1.35 [1.14, 1.60] |
| Cohen 2022 | 0.652325 | 0.056924 | 14.1% | 1.92 [1.72, 2.15] |
| Wan 2023a, UK cohort | 0.04038 | 0.14434 | 11.2% | 1.04 [0.78, 1.38] |
| **Subtotal (95% CI)** | | | **38.6%** | **1.42 [1.01, 2.00]** |
| Heterogeneity: Tau² = 0.08; Chi² = 22.64, df = 2 (P < 0.0001); I² = 91% | | | | |
| Test for overall effect: Z = 2.01 (P = 0.04) | | | | |
| **Total (95% CI)** | | | **100.0%** | **1.43 [1.18, 1.73]** |
| Heterogeneity: Tau² = 0.06; Chi² = 264.27, df = 7 (P < 0.00001); I² = 97% | | | | |
| Test for overall effect: Z = 3.69 (P = 0.0002) | | | | |
| Test for subgroup differences: Chi² = 0.00, df = 1 (P = 0.96), I² = 0% | | | | |

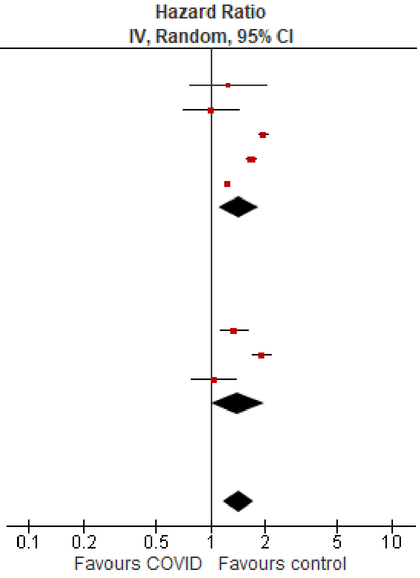

*Cardiovascular disorders, heart failure*

Outpatient/mixed, 18-64 y

| Study or Subgroup | log[Hazard Ratio] | SE | Weight | Hazard Ratio IV, Random, 95% CI |
|---|---|---|---|---|
| **4.1.1 Tested +ve** | | | | |
| Lam 2023 | 0.582216 | 0.194005 | 11.4% | 1.79 [1.22, 2.62] |
| Lim 2023 | 0.615186 | 0.105693 | 13.9% | 1.85 [1.50, 2.28] |
| Wang 2022b, 20-44y | 0.756122 | 0.063915 | 14.7% | 2.13 [1.88, 2.41] |
| Wang 2022b, 45-64y | 0.836381 | 0.035453 | 15.1% | 2.31 [2.15, 2.47] |
| Zang 2023, INSIGHT cohort | 0.110557 | 0.063558 | 14.7% | 1.12 [0.99, 1.27] |
| **Subtotal (95% CI)** | | | **69.8%** | **1.79 [1.31, 2.43]** |
| Heterogeneity: Tau² = 0.11; Chi² = 101.75, df = 4 (P < 0.00001); I² = 96% | | | | |
| Test for overall effect: Z = 3.69 (P = 0.0002) | | | | |
| **4.1.2 Other** | | | | |
| Batistoni 2024 | 0.947789 | 0.339347 | 7.6% | 2.58 [1.33, 5.02] |
| Daugherty 2021 | 0.963174 | 0.160982 | 12.4% | 2.62 [1.91, 3.59] |
| Rezel-Potts 2022 | -0.04229 | 0.237164 | 10.2% | 0.96 [0.60, 1.53] |
| **Subtotal (95% CI)** | | | **30.2%** | **1.86 [0.94, 3.68]** |
| Heterogeneity: Tau² = 0.30; Chi² = 12.99, df = 2 (P = 0.002); I² = 85% | | | | |
| Test for overall effect: Z = 1.77 (P = 0.08) | | | | |
| **Total (95% CI)** | | | **100.0%** | **1.81 [1.40, 2.34]** |
| Heterogeneity: Tau² = 0.11; Chi² = 114.74, df = 7 (P < 0.00001); I² = 94% | | | | |
| Test for overall effect: Z = 4.50 (P < 0.00001) | | | | |
| Test for subgroup differences: Chi² = 0.01, df = 1 (P = 0.92), I² = 0% | | | | |

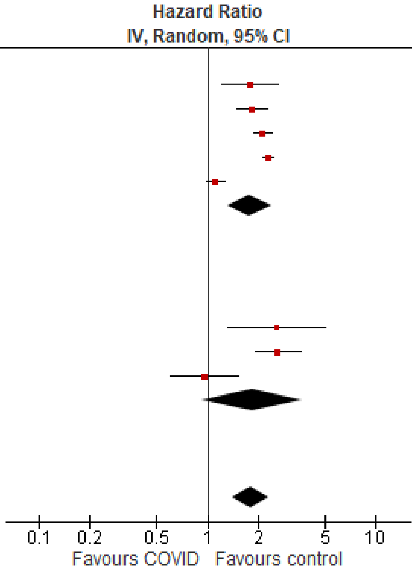

#### Outpatient/mixed ≥65 y

| Study or Subgroup | log[Hazard Ratio] | SE | Weight | Hazard Ratio IV, Random, 95% CI |
|---|---|---|---|---|
| **4.2.1 Tested +ve** | | | | |
| Wang 2022b | 0.744315 | 0.030161 | 17.7% | 2.10 [1.98, 2.23] |
| Yang 2024 | 0.139 | 0.02173 | 17.8% | 1.15 [1.10, 1.20] |
| Zang 2023, INSIGHT cohort | 0.2389 | 0.0423 | 17.5% | 1.27 [1.17, 1.38] |
| **Subtotal (95% CI)** | | | **53.0%** | **1.45 [0.98, 2.17]** |
| Heterogeneity: Tau² = 0.12; Chi² = 270.60, df = 2 (P < 0.00001); I² = 99% | | | | |
| Test for overall effect: Z = 1.84 (P = 0.07) | | | | |
| **4.2.2 Other** | | | | |
| Batistoni 2024 | 0.67294 | 0.15286 | 14.7% | 1.96 [1.45, 2.64] |
| Cohen 2022 | 0.615186 | 0.0714 | 17.0% | 1.85 [1.61, 2.13] |
| Wan 2023a, UK cohort | 0.47 | 0.13678 | 15.3% | 1.60 [1.22, 2.09] |
| **Subtotal (95% CI)** | | | **47.0%** | **1.82 [1.62, 2.04]** |
| Heterogeneity: Tau² = 0.00; Chi² = 1.17, df = 2 (P = 0.56); I² = 0% | | | | |
| Test for overall effect: Z = 10.21 (P < 0.00001) | | | | |
| **Total (95% CI)** | | | **100.0%** | **1.61 [1.22, 2.12]** |
| Heterogeneity: Tau² = 0.11; Chi² = 291.13, df = 5 (P < 0.00001); I² = 98% | | | | |
| Test for overall effect: Z = 3.37 (P = 0.0007) | | | | |
| Test for subgroup differences: Chi² = 1.11, df = 1 (P = 0.29), I² = 10.0% | | | | |

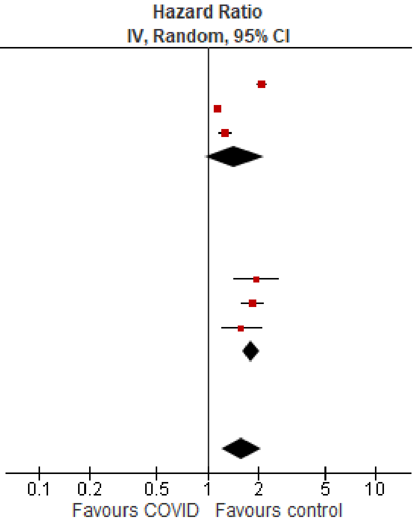

##### *Diabetes, overall*

#### Outpatient/mixed, <18 y

| Study or Subgroup | log[Hazard Ratio] | SE | Weight | Hazard Ratio IV, Random, 95% CI |
|---|---|---|---|---|
| **5.1.1 Tested +ve** | | | | |
| Gordon 2024, 0-1 y | 0.920402 | 0.730235 | 1.0% | 2.51 [0.60, 10.50] |
| Gordon 2024, 12-17 y | 0.177811 | 0.101203 | 15.7% | 1.19 [0.98, 1.46] |
| Gordon 2024, 2-4 y | 0.828814 | 0.385061 | 3.3% | 2.29 [1.08, 4.87] |
| Gordon 2024, 5-11 y | 0.362418 | 0.159062 | 11.2% | 1.44 [1.05, 1.96] |
| Mizrahi 2023, 12-18 y | 0.431782 | 0.485066 | 2.2% | 1.54 [0.60, 3.98] |
| Noorzae 2023, 0-4 y | 0.086178 | 0.28434 | 5.4% | 1.09 [0.62, 1.90] |
| Noorzae 2023, 11-17 y | -0.27444 | 0.138961 | 12.6% | 0.76 [0.58, 1.00] |
| Noorzae 2023, 5-10 y | -0.09431 | 0.159087 | 11.2% | 0.91 [0.67, 1.24] |
| Tartof 2022 | -0.35667 | 0.215354 | 8.0% | 0.70 [0.46, 1.07] |
| **Subtotal (95% CI)** | | | **70.5%** | **1.09 [0.87, 1.37]** |
| Heterogeneity: Tau² = 0.06; Chi² = 21.42, df = 8 (P = 0.006); I² = 63% | | | | |
| Test for overall effect: Z = 0.74 (P = 0.46) | | | | |
| **5.1.2 Other** | | | | |
| Kompaniyets 2022 | 0.171211 | 0.029592 | 20.9% | 1.19 [1.12, 1.26] |
| Pietropaolo 2022 | 0.188465 | 0.202801 | 8.6% | 1.21 [0.81, 1.80] |
| **Subtotal (95% CI)** | | | **29.5%** | **1.19 [1.12, 1.26]** |
| Heterogeneity: Tau² = 0.00; Chi² = 0.01, df = 1 (P = 0.93); I² = 0% | | | | |
| Test for overall effect: Z = 5.86 (P < 0.00001) | | | | |
| **Total (95% CI)** | | | **100.0%** | **1.10 [0.95, 1.28]** |
| Heterogeneity: Tau² = 0.03; Chi² = 24.24, df = 10 (P = 0.007); I² = 59% | | | | |
| Test for overall effect: Z = 1.28 (P = 0.20) | | | | |
| Test for subgroup differences: Chi² = 0.53, df = 1 (P = 0.47), I² = 0% | | | | |

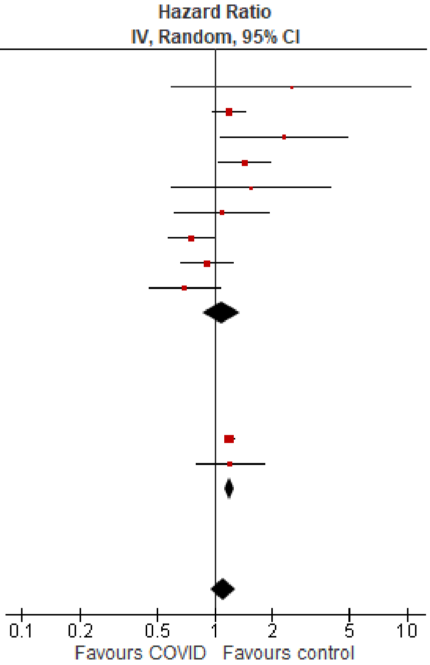

#### Outpatient/mixed, 18-64 y

| Study or Subgroup | log[Hazard Ratio] | SE | Weight | Hazard Ratio IV, Random, 95% CI |
|---|---|---|---|---|
| **5.2.1 Tested +ve** | | | | |
| Chang 2023, 18-40 y | 1.022451 | 0.056719 | 6.0% | 2.78 [2.49, 3.11] |
| Chang 2023, 41-64 y | 0.951658 | 0.050104 | 6.0% | 2.59 [2.35, 2.86] |
| Mizrahi 2023, 19-40 y | 0.301483 | 0.296006 | 3.5% | 1.35 [0.76, 2.41] |
| Mizrahi 2023, 41-60 y | 0.115372 | 0.125237 | 5.4% | 1.12 [0.88, 1.43] |
| Mizrahi 2023, 60+ y | 0.05005 | 0.372991 | 2.8% | 1.05 [0.51, 2.18] |
| Naveed 2023 | 0.157004 | 0.04811 | 6.0% | 1.17 [1.06, 1.29] |
| Xie 2022a | 0.307485 | 0.018766 | 6.1% | 1.36 [1.31, 1.41] |
| Zang 2023, INSIGHT cohort | 1.308333 | 0.041054 | 6.0% | 3.70 [3.41, 4.01] |
| **Subtotal (95% CI)** | | | **41.8%** | **1.75 [1.21, 2.53]** |
| Heterogeneity: Tau² = 0.26; Chi² = 719.17, df = 7 (P < 0.00001); I² = 99% | | | | |
| Test for overall effect: Z = 2.97 (P = 0.003) | | | | |
| **5.2.2 Other** | | | | |
| Choi 2023 | 0.431782 | 0.021478 | 6.1% | 1.54 [1.48, 1.61] |
| Daugherty 2021 | 0.604316 | 0.127551 | 5.4% | 1.83 [1.43, 2.35] |
| Pietropaolo 2022 | -0.07035 | 0.152822 | 5.1% | 0.93 [0.69, 1.26] |
| Rezel-Potts 2022 | 0.110593 | 0.048347 | 6.0% | 1.12 [1.02, 1.23] |
| Taylor 2024, pre-vaccination cohort, 18-39 y | 0.395044 | 0.075306 | 5.8% | 1.48 [1.28, 1.72] |
| Taylor 2024, pre-vaccination cohort, 40-59 y | 0.212603 | 0.035455 | 6.0% | 1.24 [1.15, 1.33] |
| Taylor 2024, pre-vaccination cohort, 60-79 y | 0.252617 | 0.087333 | 5.7% | 1.29 [1.08, 1.53] |
| Taylor 2024, unvaccinated cohort, 40-59 y | 0.604316 | 0.153076 | 5.1% | 1.83 [1.36, 2.47] |
| Taylor 2024, unvaccinated cohort, 60-79 y | 0.883768 | 0.54832 | 1.7% | 2.42 [0.83, 7.09] |
| Taylor 2024, vaccinated cohort, 40-59 y | 0.173953 | 0.059796 | 5.9% | 1.19 [1.06, 1.34] |
| Taylor 2024, vaccinated cohort, 60-79 y | 0.223144 | 0.146779 | 5.2% | 1.25 [0.94, 1.67] |
| **Subtotal (95% CI)** | | | **58.2%** | **1.34 [1.20, 1.50]** |
| Heterogeneity: Tau² = 0.02; Chi² = 78.61, df = 10 (P < 0.00001); I² = 87% | | | | |
| Test for overall effect: Z = 5.17 (P < 0.00001) | | | | |
| **Total (95% CI)** | | | **100.0%** | **1.53 [1.29, 1.81]** |
| Heterogeneity: Tau² = 0.12; Chi² = 878.07, df = 18 (P < 0.00001); I² = 98% | | | | |
| Test for overall effect: Z = 4.95 (P < 0.00001) | | | | |
| Test for subgroup differences: Chi² = 1.81, df = 1 (P = 0.18), I² = 44.9% | | | | |

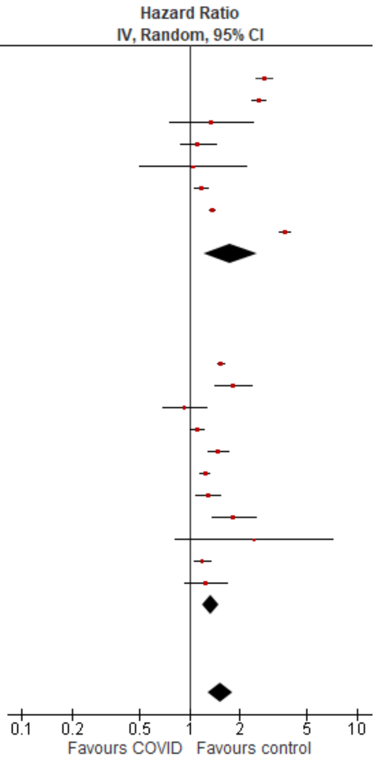

#### Outpatient/mixed ≥65 y

| Study or Subgroup | log[Hazard Ratio] | SE | Weight | Hazard Ratio IV, Random, 95% CI |
|---|---|---|---|---|
| **5.3.1 Tested +ve** | | | | |
| Chang 2023 | 0 | 0 | | Not estimable |
| Mizrahi 2023, 60+ y | 0.05005 | 0.837214 | 0.2% | 1.05 [0.20, 5.42] |
| Xie 2022a | 0.357674 | 0.02142 | 23.5% | 1.43 [1.37, 1.49] |
| Zang 2023, INSIGHT cohort | 0.401926 | 0.037497 | 20.7% | 1.49 [1.39, 1.61] |
| **Subtotal (95% CI)** | | | **44.4%** | **1.45 [1.39, 1.50]** |
| Heterogeneity: Tau² = 0.00; Chi² = 1.19, df = 2 (P = 0.55); I² = 0% | | | | |
| Test for overall effect: Z = 19.81 (P < 0.00001) | | | | |
| **5.3.2 Other** | | | | |
| Cohen 2022 | 0.672944 | 0.103435 | 9.4% | 1.96 [1.60, 2.40] |
| Taylor 2024, pre-vaccination cohort, 60-79 y | 0.252617 | 0.050422 | 18.1% | 1.29 [1.17, 1.42] |
| Taylor 2024, pre-vaccination cohort, 80-110 y | 0.240626 | 0.093384 | 10.7% | 1.27 [1.06, 1.53] |
| Taylor 2024, unvaccinated cohort, 60-79 y | 0.883768 | 0.316573 | 1.5% | 2.42 [1.30, 4.50] |
| Taylor 2024, vaccinated cohort, 60-79 y | 0.223144 | 0.084743 | 11.9% | 1.25 [1.06, 1.48] |
| Taylor 2024, vaccination cohort, 80-110 y | 0.350657 | 0.184439 | 4.0% | 1.42 [0.99, 2.04] |
| **Subtotal (95% CI)** | | | **55.6%** | **1.44 [1.23, 1.70]** |
| Heterogeneity: Tau² = 0.03; Chi² = 18.54, df = 5 (P = 0.002); I² = 73% | | | | |
| Test for overall effect: Z = 4.46 (P < 0.00001) | | | | |
| **Total (95% CI)** | | | **100.0%** | **1.43 [1.32, 1.54]** |
| Heterogeneity: Tau² = 0.01; Chi² = 22.00, df = 8 (P = 0.005); I² = 64% | | | | |
| Test for overall effect: Z = 8.86 (P < 0.00001) | | | | |
| Test for subgroup differences: Chi² = 0.00, df = 1 (P = 1.00), I² = 0% | | | | |

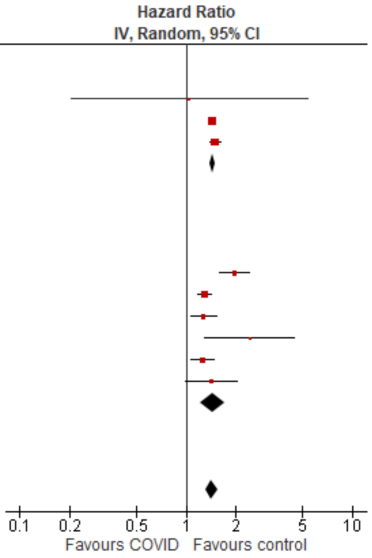

#### *Mental illness, overall*

<u>Outpatient/mixed, <18 y</u>

| Study or Subgroup | log[Hazard Ratio] | SE | Weight | Hazard Ratio IV, Random, 95% CI |
|---|---|---|---|---|
| **6.1.1 Tested +ve** | | | | |
| Gordon 2024, 0-1 y | -0.19237 | 0.234921 | 4.1% | 0.83 [0.52, 1.31] |
| Gordon 2024, 12-17 y | -0.11649 | 0.021231 | 7.3% | 0.89 [0.85, 0.93] |
| Gordon 2024, 2-4 y | 0.016261 | 0.080112 | 6.7% | 1.02 [0.87, 1.19] |
| Gordon 2024, 5-11 y | -0.03881 | 0.03265 | 7.2% | 0.96 [0.90, 1.03] |
| Mizrahi 2023, 12-18 y | -0.10932 | 0.200457 | 4.6% | 0.90 [0.61, 1.33] |
| Nielsen 2024 | -0.31471 | 0.027984 | 7.2% | 0.73 [0.69, 0.77] |
| Rao 2022 | 0.482426 | 0.059359 | 6.9% | 1.62 [1.44, 1.82] |
| Roessler 2022 | 0.360624 | 0.096104 | 6.4% | 1.43 [1.19, 1.73] |
| Taquet 2022 | -0.16626 | 0.039868 | 7.1% | 0.85 [0.78, 0.92] |
| Tartof 2022 | -0.1568 | 0.313754 | 3.0% | 0.85 [0.46, 1.58] |
| Valdivieso-Martinez 2023, unvaccinated cohort | -0.24484 | 0.597572 | 1.2% | 0.78 [0.24, 2.53] |
| Valdivieso-Martinez 2023, vaccinated cohort | -0.01384 | 0.427767 | 2.0% | 0.99 [0.43, 2.28] |
| **Subtotal (95% CI)** | | | **63.7%** | **0.99 [0.86, 1.14]** |
| Heterogeneity: Tau² = 0.04; Chi² = 186.66, df = 11 (P < 0.00001); I² = 94% | | | | |
| Test for overall effect: Z = 0.14 (P = 0.89) | | | | |
| **6.1.2 Other** | | | | |
| Donnachie 2022, 0-11 y | 0.144966 | 0.037248 | 7.2% | 1.16 [1.07, 1.24] |
| Donnachie 2022, 12-17 y | -0.23067 | 0.023763 | 7.2% | 0.79 [0.76, 0.83] |
| Kompaniyets 2022 | -0.1726 | 0.010074 | 7.3% | 0.84 [0.83, 0.86] |
| Zhang-James 2024, 13-18 y | 0.04879 | 0.009719 | 7.3% | 1.05 [1.03, 1.07] |
| Zhang-James 2024, under 13 y | 0.451076 | 0.013002 | 7.3% | 1.57 [1.53, 1.61] |
| **Subtotal (95% CI)** | | | **36.3%** | **1.05 [0.82, 1.34]** |
| Heterogeneity: Tau² = 0.08; Chi² = 1583.44, df = 4 (P < 0.00001); I² = 100% | | | | |
| Test for overall effect: Z = 0.39 (P = 0.70) | | | | |
| **Total (95% CI)** | | | **100.0%** | **1.01 [0.88, 1.16]** |
| Heterogeneity: Tau² = 0.07; Chi² = 1879.94, df = 16 (P < 0.00001); I² = 99% | | | | |
| Test for overall effect: Z = 0.12 (P = 0.90) | | | | |
| Test for subgroup differences: Chi² = 0.16, df = 1 (P = 0.69), I² = 0% | | | | |

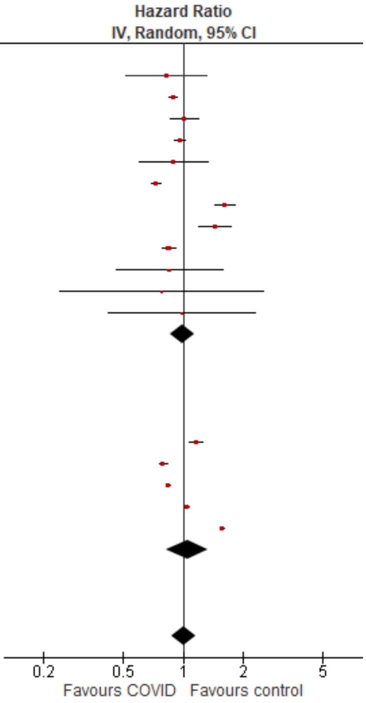

#### Outpatient/mixed, 18-64 y

| Study or Subgroup | log[Hazard Ratio] | SE | Weight | Hazard Ratio IV, Random, 95% CI |
|---|---|---|---|---|
| **6.2.1 Tested +ve** | | | | |
| Abel 2021, 16-24 y | 0.24686 | 0.109916 | 3.3% | 1.28 [1.03, 1.59] |
| Abel 2021, 25-34 y | 0.463734 | 0.120405 | 3.2% | 1.59 [1.26, 2.01] |
| Abel 2021, 35-49 y | 0.693147 | 0.102396 | 3.4% | 2.00 [1.64, 2.44] |
| Abel 2021, 50-59 y | 0.806476 | 0.127085 | 3.1% | 2.24 [1.75, 2.87] |
| Abel 2021, 60-69 y | 1.128171 | 0.236563 | 2.0% | 3.09 [1.94, 4.91] |
| Donnachie 2022, 18-39y | 0.325556 | 0.008473 | 4.1% | 1.38 [1.36, 1.41] |
| Donnachie 2022, 40-59y | -0.00321 | 0.009212 | 4.1% | 1.00 [0.98, 1.01] |
| Donnachie 2022, 60+ y | 0.094219 | 0.037287 | 4.0% | 1.10 [1.02, 1.18] |
| Lam 2023 | 0.523995 | 0.307968 | 1.5% | 1.69 [0.92, 3.09] |
| Mizrahi 2023, 19-40 y | 0.028888 | 0.134651 | 3.0% | 1.03 [0.79, 1.34] |
| Mizrahi 2023, 41-60 y | 0.091665 | 0.170862 | 2.6% | 1.10 [0.78, 1.53] |
| Mizrahi 2023, 60+ y | 0.062342 | 0.503706 | 0.7% | 1.06 [0.40, 2.86] |
| Nielsen 2024, 18-39 y | -0.27444 | 0.02015 | 4.1% | 0.76 [0.73, 0.79] |
| Nielsen 2024, 40-64 y | -0.34249 | 0.028774 | 4.0% | 0.71 [0.67, 0.75] |
| Taquet 2022 | -0.01706 | 0.008532 | 4.1% | 0.98 [0.97, 1.00] |
| Xu 2022 | 0.425268 | 0.024948 | 4.0% | 1.53 [1.46, 1.61] |
| Zang 2023, INSIGHT cohort | 0.121421 | 0.033472 | 4.0% | 1.13 [1.06, 1.21] |
| **Subtotal (95% CI)** | | | **55.3%** | **1.24 [1.10, 1.40]** |
| Heterogeneity: Tau² = 0.05; Chi² = 1866.84, df = 16 (P < 0.00001); I² = 99% | | | | |
| Test for overall effect: Z = 3.46 (P = 0.0005) | | | | |
| **6.2.2 Other** | | | | |
| Daugherty 2021 | 0.329304 | 0.036505 | 4.0% | 1.39 [1.29, 1.49] |
| Jacob 2022, 18-30 y | -0.01855 | 0.095097 | 3.5% | 0.98 [0.81, 1.18] |
| Jacob 2022, 31-40 y | -0.05372 | 0.107087 | 3.4% | 0.95 [0.77, 1.17] |
| Jacob 2022, 41-50 y | -0.00605 | 0.103521 | 3.4% | 0.99 [0.81, 1.22] |
| Jacob 2022, 51-60 y | -0.05027 | 0.086349 | 3.6% | 0.95 [0.80, 1.13] |
| Jacob 2022, 61-70 y | 0.122487 | 0.191274 | 2.4% | 1.13 [0.78, 1.64] |
| Park 2021, 20-39 y | 0.470004 | 0.083952 | 3.6% | 1.60 [1.36, 1.89] |
| Park 2021, 40-59 y | 0.81978 | 0.066238 | 3.8% | 2.27 [1.99, 2.58] |
| Smith 2024, 16-30 y | 0.157004 | 0.076811 | 3.7% | 1.17 [1.01, 1.36] |
| Smith 2024, 31-40 y | 0.254642 | 0.078027 | 3.7% | 1.29 [1.11, 1.50] |
| Smith 2024, 41-50 y | 0.231112 | 0.078648 | 3.7% | 1.26 [1.08, 1.47] |
| Smith 2024, 51-60 y | 0.182322 | 0.074001 | 3.7% | 1.20 [1.04, 1.39] |
| Smith 2024, 60+ y | 0.223144 | 0.196996 | 2.4% | 1.25 [0.85, 1.84] |
| **Subtotal (95% CI)** | | | **44.7%** | **1.24 [1.07, 1.42]** |
| Heterogeneity: Tau² = 0.06; Chi² = 120.74, df = 12 (P < 0.00001); I² = 90% | | | | |
| Test for overall effect: Z = 2.97 (P = 0.003) | | | | |
| **Total (95% CI)** | | | **100.0%** | **1.24 [1.13, 1.36]** |
| Heterogeneity: Tau² = 0.05; Chi² = 2064.61, df = 29 (P < 0.00001); I² = 99% | | | | |
| Test for overall effect: Z = 4.59 (P < 0.00001) | | | | |
| Test for subgroup differences: Chi² = 0.00, df = 1 (P = 0.98), I² = 0% | | | | |

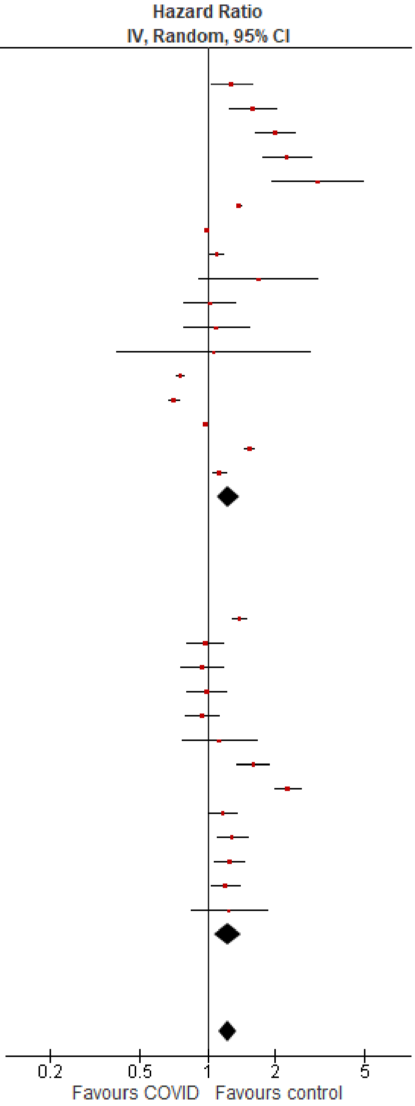

#### Outpatient/mixed ≥65 y

| Study or Subgroup | log[Hazard Ratio] | SE | Weight | Hazard Ratio IV, Random, 95% CI |
|---|---|---|---|---|
| **6.3.1 Tested +ve** | | | | |
| Abel 2021, 60-69 y | 1.128171 | 0.259142 | 3.0% | 3.09 [1.86, 5.14] |
| Abel 2021, 70-79 y | 1.297463 | 0.307646 | 2.5% | 3.66 [2.00, 6.69] |
| Abel 2021, 80+ y | 1.427916 | 0.228145 | 3.5% | 4.17 [2.67, 6.52] |
| Donnachie 2022, 60+ y | 0.094219 | 0.014441 | 7.0% | 1.10 [1.07, 1.13] |
| Mizrahi 2023, 60+ y | -0.31714 | 0.219835 | 3.6% | 0.73 [0.47, 1.12] |
| Murata 2024, alpha variant | -0.71335 | 0.689518 | 0.7% | 0.49 [0.13, 1.89] |
| Murata 2024, delta variant | 0.086178 | 0.339601 | 2.2% | 1.09 [0.56, 2.12] |
| Murata 2024, omicron BA.1/BA.2 variant | -0.52763 | 0.131419 | 5.2% | 0.59 [0.46, 0.76] |
| Murata 2024, omicron BA.5 variant | -0.79851 | 0.101126 | 5.8% | 0.45 [0.37, 0.55] |
| Nielsen 2024 | -0.30111 | 0.055374 | 6.6% | 0.74 [0.66, 0.82] |
| Taquet 2022 | 0.045458 | 0.015112 | 7.0% | 1.05 [1.02, 1.08] |
| Xu 2022 | 0.270027 | 0.02524 | 6.9% | 1.31 [1.25, 1.38] |
| Zang 2023, INSIGHT cohort | 0.18938 | 0.052341 | 6.7% | 1.21 [1.09, 1.34] |
| **Subtotal (95% CI)** | | | **60.6%** | **1.06 [0.93, 1.21]** |
| Heterogeneity: Tau² = 0.03; Chi² = 282.72, df = 12 (P < 0.00001); I² = 96% | | | | |
| Test for overall effect: Z = 0.90 (P = 0.37) | | | | |
| **6.3.2 Other** | | | | |
| Cohen 2022 | 0.565314 | 0.056278 | 6.6% | 1.76 [1.58, 1.97] |
| Jacob 2022, 61-70 y | 0.122487 | 0.174608 | 4.4% | 1.13 [0.80, 1.59] |
| Jacob 2022, 70+ y | 0.084079 | 0.145272 | 5.0% | 1.09 [0.82, 1.45] |
| Park 2021, 60+ y | 0.620576 | 0.070584 | 6.4% | 1.86 [1.62, 2.14] |
| Rahman 2024, 60+ y | 0.844623 | 0.096388 | 5.9% | 2.33 [1.93, 2.81] |
| Smith 2024, 60+ y | 0.223144 | 0.085976 | 6.1% | 1.25 [1.06, 1.48] |
| Wan 2023a, UK cohort | 0.53487 | 0.14634 | 4.9% | 1.71 [1.28, 2.27] |
| **Subtotal (95% CI)** | | | **39.4%** | **1.57 [1.30, 1.89]** |
| Heterogeneity: Tau² = 0.05; Chi² = 40.12, df = 6 (P < 0.00001); I² = 85% | | | | |
| Test for overall effect: Z = 4.77 (P < 0.00001) | | | | |
| **Total (95% CI)** | | | **100.0%** | **1.25 [1.11, 1.41]** |
| Heterogeneity: Tau² = 0.05; Chi² = 475.96, df = 19 (P < 0.00001); I² = 96% | | | | |
| Test for overall effect: Z = 3.76 (P = 0.0002) | | | | |
| Test for subgroup differences: Chi² = 11.51, df = 1 (P = 0.0007), I² = 91.3% | | | | |

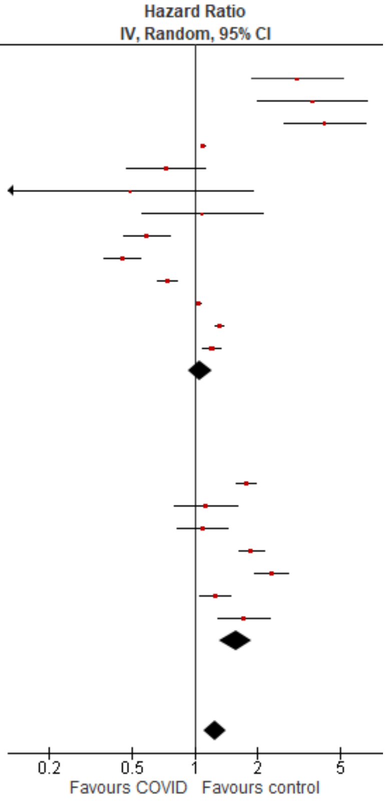

#### Inpatients, ≥65 y

| Study or Subgroup | log[Hazard Ratio] | SE | Weight | Hazard Ratio IV, Random, 95% CI |
|---|---|---|---|---|
| **6.4.1 Tested +ve** | | | | |
| Murata 2024, alpha variant | -0.08338 | 0.211931 | 14.2% | 0.92 [0.61, 1.39] |
| Murata 2024, delta variant | 0.173953 | 0.160164 | 15.0% | 1.19 [0.87, 1.63] |
| Murata 2024, omicron BA.1/BA.2 variant | -0.18633 | 0.092095 | 15.8% | 0.83 [0.69, 0.99] |
| Murata 2024, omicron BA.5 variant | -0.19845 | 0.104701 | 15.7% | 0.82 [0.67, 1.01] |
| Nersejan 2023, 50-69 y | -0.94161 | 0.590548 | 7.6% | 0.39 [0.12, 1.24] |
| **Subtotal (95% CI)** | | | **68.2%** | **0.88 [0.75, 1.05]** |
| Heterogeneity: Tau² = 0.01; Chi² = 6.32, df = 4 (P = 0.18); I² = 37% | | | | |
| Test for overall effect: Z = 1.44 (P = 0.15) | | | | |
| **6.4.2 Other** | | | | |
| Cohen 2022 | 0.804107 | 0.041652 | 16.1% | 2.23 [2.06, 2.42] |
| Rahman 2024, 60+ y | 0.844623 | 0.099187 | 15.7% | 2.33 [1.92, 2.83] |
| **Subtotal (95% CI)** | | | **31.8%** | **2.25 [2.09, 2.42]** |
| Heterogeneity: Tau² = 0.00; Chi² = 0.14, df = 1 (P = 0.71); I² = 0% | | | | |
| Test for overall effect: Z = 21.10 (P < 0.00001) | | | | |
| **Total (95% CI)** | | | **100.0%** | **1.16 [0.74, 1.79]** |
| Heterogeneity: Tau² = 0.31; Chi² = 181.75, df = 6 (P < 0.00001); I² = 97% | | | | |
| Test for overall effect: Z = 0.64 (P = 0.52) | | | | |
| Test for subgroup differences: Chi² = 98.86, df = 1 (P < 0.00001), I² = 99.0% | | | | |

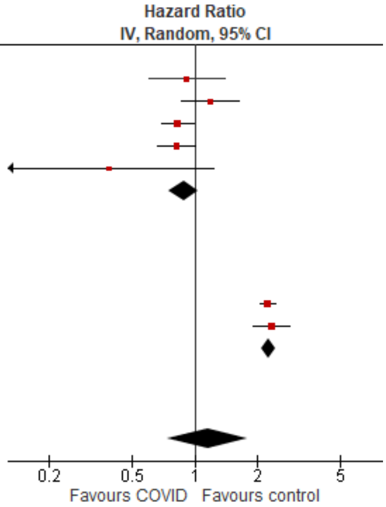

##### *Mental illness, anxiety/anxiety disorders*

###### Outpatient/mixed, 18-64 y

| Study or Subgroup | log[Hazard Ratio] | SE | Weight | Hazard Ratio IV, Random, 95% CI |
|---|---|---|---|---|
| **7.1.1 Tested +ve** | | | | |
| Lam 2023 | 0.29267 | 0.140606 | 7.7% | 1.34 [1.02, 1.77] |
| Mizrahi 2023, 19-40 y | 0.041275 | 0.085815 | 10.3% | 1.04 [0.88, 1.23] |
| Mizrahi 2023, 41-60 y | 0.118999 | 0.113247 | 8.9% | 1.13 [0.90, 1.41] |
| Mizrahi 2023, 60+ y | 0.22156 | 0.57783 | 1.0% | 1.25 [0.40, 3.87] |
| Taquet 2022 | -0.02922 | 0.003546 | 12.8% | 0.97 [0.96, 0.98] |
| Zang 2023, INSIGHT cohort | 0.121421 | 0.033472 | 12.4% | 1.13 [1.06, 1.21] |
| **Subtotal (95% CI)** | | | **53.1%** | **1.08 [0.97, 1.21]** |
| Heterogeneity: Tau² = 0.01; Chi² = 27.70, df = 5 (P < 0.0001); I² = 82% | | | | |
| Test for overall effect: Z = 1.44 (P = 0.15) | | | | |
| **7.1.2 Other** | | | | |
| Daugherty 2021 | 0.350657 | 0.037662 | 12.3% | 1.42 [1.32, 1.53] |
| Jacob 2022, 18-30 y | -0.10536 | 0.13104 | 8.1% | 0.90 [0.70, 1.16] |
| Jacob 2022, 31-40 y | -0.26136 | 0.153403 | 7.1% | 0.77 [0.57, 1.04] |
| Jacob 2022, 41-50 y | 0.067659 | 0.149946 | 7.3% | 1.07 [0.80, 1.44] |
| Jacob 2022, 51-60 y | 0.067659 | 0.121884 | 8.5% | 1.07 [0.84, 1.36] |
| Jacob 2022, 61-70 y | 0.04879 | 0.274293 | 3.6% | 1.05 [0.61, 1.80] |
| **Subtotal (95% CI)** | | | **46.9%** | **1.04 [0.83, 1.32]** |
| Heterogeneity: Tau² = 0.06; Chi² = 29.83, df = 5 (P < 0.0001); I² = 83% | | | | |
| Test for overall effect: Z = 0.36 (P = 0.72) | | | | |
| **Total (95% CI)** | | | **100.0%** | **1.08 [0.96, 1.22]** |
| Heterogeneity: Tau² = 0.03; Chi² = 131.22, df = 11 (P < 0.00001); I² = 92% | | | | |
| Test for overall effect: Z = 1.29 (P = 0.20) | | | | |
| Test for subgroup differences: Chi² = 0.07, df = 1 (P = 0.78), I² = 0% | | | | |

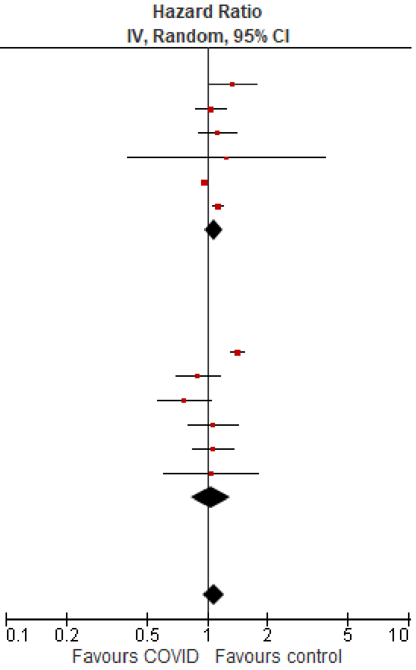

###### Outpatient/mixed ≥65 y

| Study or Subgroup | log[Hazard Ratio] | SE | Weight | Hazard Ratio IV, Random, 95% CI |
|---|---|---|---|---|
| **7.2.1 Tested +ve** | | | | |
| Mizrahi 2023, 60+ y | 0.22156 | 0.252186 | 8.8% | 1.25 [0.76, 2.05] |
| Murata 2024, alpha variant | 0.131028 | 0.574672 | 3.6% | 1.14 [0.37, 3.52] |
| Murata 2024, delta variant | -0.4943 | 0.572963 | 3.6% | 0.61 [0.20, 1.88] |
| Murata 2024, omicron BA.1/BA.2 variant | -1.13943 | 0.210887 | 9.8% | 0.32 [0.21, 0.48] |
| Murata 2024, omicron BA.5 variant | -1.10866 | 0.04262 | 13.3% | 0.33 [0.30, 0.36] |
| Taquet 2022 | 0.013706 | 0.008429 | 13.5% | 1.01 [1.00, 1.03] |
| Zang 2023, INSIGHT cohort | 0.18938 | 0.052341 | 13.2% | 1.21 [1.09, 1.34] |
| **Subtotal (95% CI)** | | | **65.7%** | **0.72 [0.43, 1.19]** |
| Heterogeneity: Tau² = 0.39; Chi² = 713.97, df = 6 (P < 0.00001); I² = 99% | | | | |
| Test for overall effect: Z = 1.29 (P = 0.20) | | | | |
| **7.2.2 Other** | | | | |
| Jacob 2022, 61-70 y | -0.06188 | 0.019888 | 13.4% | 0.94 [0.90, 0.98] |
| Jacob 2022, 70+ y | 0.04879 | 0.211401 | 9.8% | 1.05 [0.69, 1.59] |
| Wan 2023a, UK cohort | 0.530628 | 0.16273 | 11.0% | 1.70 [1.24, 2.34] |
| **Subtotal (95% CI)** | | | **34.3%** | **1.17 [0.80, 1.72]** |
| Heterogeneity: Tau² = 0.10; Chi² = 13.29, df = 2 (P = 0.001); I² = 85% | | | | |
| Test for overall effect: Z = 0.80 (P = 0.42) | | | | |
| **Total (95% CI)** | | | **100.0%** | **0.84 [0.66, 1.08]** |
| Heterogeneity: Tau² = 0.12; Chi² = 728.94, df = 9 (P < 0.00001); I² = 99% | | | | |
| Test for overall effect: Z = 1.34 (P = 0.18) | | | | |
| Test for subgroup differences: Chi² = 2.27, df = 1 (P = 0.13), I² = 55.9% | | | | |

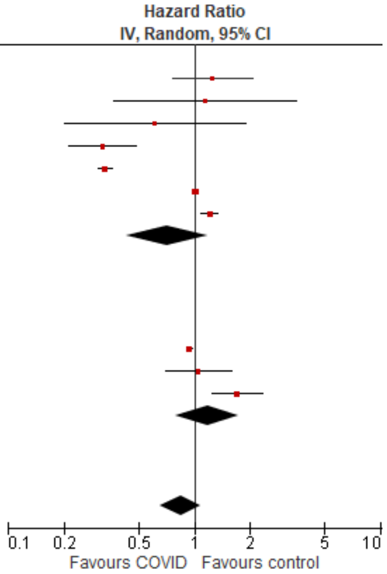

#### *Mental illness, depression & mood disorders*

##### Outpatient/mixed, <18 y

| Study or Subgroup | log[Hazard Ratio] | SE | Weight | Hazard Ratio IV, Random, 95% CI |
|---|---|---|---|---|
| **8.1.1 Tested +ve** | | | | |
| Mizrahi 2023, 12-18 y | -0.44514 | 0.21327 | 13.1% | 0.64 [0.42, 0.97] |
| Roessler 2022 | 0.371564 | 0.130768 | 13.9% | 1.45 [1.12, 1.87] |
| Taquet 2022 | -0.05933 | 0.012802 | 14.5% | 0.94 [0.92, 0.97] |
| Valdivieso-Martinez 2023, unvaccinated cohort | 1.098612 | 0.864466 | 5.3% | 3.00 [0.55, 16.33] |
| Valdivieso-Martinez 2023, vaccinated cohort | -0.48548 | 0.448891 | 9.8% | 0.62 [0.26, 1.48] |
| **Subtotal (95% CI)** | | | **56.6%** | **0.98 [0.71, 1.35]** |
| Heterogeneity: Tau² = 0.07; Chi² = 16.78, df = 4 (P = 0.002); I² = 76% | | | | |
| Test for overall effect: Z = 0.14 (P = 0.89) | | | | |
| **8.1.2 Other** | | | | |
| Kompaniyets 2022 | -0.24846136 | 0.01646391 | 14.5% | 0.78 [0.76, 0.81] |
| Zhang-James 2024, 13-18 y | 1.435085 | 0.030047 | 14.5% | 4.20 [3.96, 4.45] |
| Zhang-James 2024, under 13 y | 1.029619 | 0.054875 | 14.4% | 2.80 [2.51, 3.12] |
| **Subtotal (95% CI)** | | | **43.4%** | **2.09 [0.61, 7.23]** |
| Heterogeneity: Tau² = 1.20; Chi² = 2660.14, df = 2 (P < 0.00001); I² = 100% | | | | |
| Test for overall effect: Z = 1.17 (P = 0.24) | | | | |
| **Total (95% CI)** | | | **100.0%** | **1.37 [0.84, 2.23]** |
| Heterogeneity: Tau² = 0.43; Chi² = 2858.70, df = 7 (P < 0.00001); I² = 100% | | | | |
| Test for overall effect: Z = 1.27 (P = 0.20) | | | | |
| Test for subgroup differences: Chi² = 1.36, df = 1 (P = 0.24), I² = 26.3% | | | | |

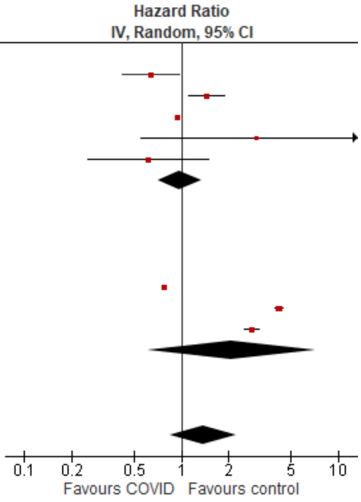

##### Outpatient/mixed, 18-64 y

| Study or Subgroup | log[Hazard Ratio] | SE | Weight | Hazard Ratio IV, Random, 95% CI |
|---|---|---|---|---|
| **8.2.1 Tested +ve** | | | | |
| Mizrahi 2023, 19-40 y | 0.062594 | 0.120899 | 5.2% | 1.06 [0.84, 1.35] |
| Mizrahi 2023, 41-60 y | 0.111062 | 0.1459 | 4.1% | 1.12 [0.84, 1.49] |
| Mizrahi 2023, 60+ y | 0.183801 | 0.592558 | 0.4% | 1.20 [0.38, 3.84] |
| Taquet 2022 | 0.006976 | 0.004509 | 12.4% | 1.01 [1.00, 1.02] |
| **Subtotal (95% CI)** | | | **22.2%** | **1.01 [1.00, 1.02]** |
| Heterogeneity: Tau² = 0.00; Chi² = 0.81, df = 3 (P = 0.85); I² = 0% | | | | |
| Test for overall effect: Z = 1.59 (P = 0.11) | | | | |
| **8.2.2 Other** | | | | |
| Daugherty 2021 | 0.239017 | 0.050181 | 10.1% | 1.27 [1.15, 1.40] |
| Jacob 2022, 18-30 y | 0.019803 | 0.087103 | 7.3% | 1.02 [0.86, 1.21] |
| Jacob 2022, 31-40 y | 0.019803 | 0.091285 | 7.0% | 1.02 [0.85, 1.22] |
| Jacob 2022, 41-50 y | -0.03046 | 0.086279 | 7.3% | 0.97 [0.82, 1.15] |
| Jacob 2022, 51-60 y | -0.06188 | 0.075739 | 8.1% | 0.94 [0.81, 1.09] |
| Jacob 2022, 61-70 y | 0.14842 | 0.162712 | 3.6% | 1.16 [0.84, 1.60] |
| Smith 2024, 16-30 y | 0.157004 | 0.076811 | 8.0% | 1.17 [1.01, 1.36] |
| Smith 2024, 31-40 y | 0.254642 | 0.078027 | 7.9% | 1.29 [1.11, 1.50] |
| Smith 2024, 41-50 y | 0.231112 | 0.078648 | 7.9% | 1.26 [1.08, 1.47] |
| Smith 2024, 51-60 y | 0.182322 | 0.074001 | 8.2% | 1.20 [1.04, 1.39] |
| Smith 2024, 60+ y | 0.223144 | 0.196996 | 2.7% | 1.25 [0.85, 1.84] |
| **Subtotal (95% CI)** | | | **77.8%** | **1.13 [1.05, 1.22]** |
| Heterogeneity: Tau² = 0.01; Chi² = 22.59, df = 10 (P = 0.01); I² = 56% | | | | |
| Test for overall effect: Z = 3.24 (P = 0.001) | | | | |
| **Total (95% CI)** | | | **100.0%** | **1.11 [1.04, 1.19]** |
| Heterogeneity: Tau² = 0.01; Chi² = 51.77, df = 14 (P < 0.00001); I² = 73% | | | | |
| Test for overall effect: Z = 2.92 (P = 0.003) | | | | |
| Test for subgroup differences: Chi² = 9.21, df = 1 (P = 0.002), I² = 89.1% | | | | |

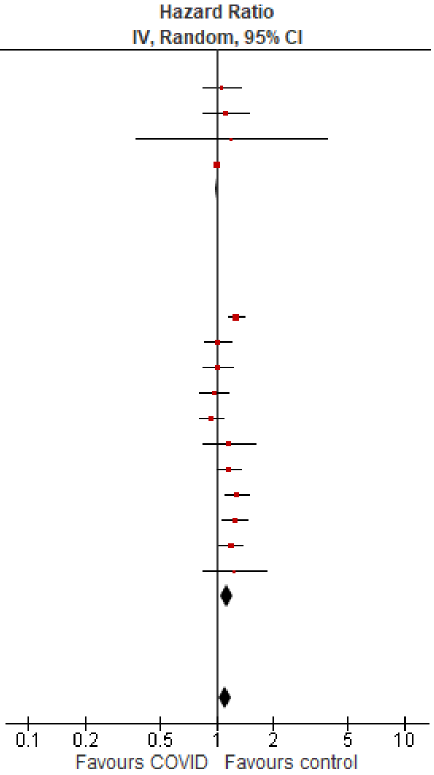

#### Outpatient/mixed ≥65 y

| Study or Subgroup | log[Hazard Ratio] | SE | Weight | Hazard Ratio IV, Random, 95% CI |
|---|---|---|---|---|
| **8.3.1 Tested +ve** | | | | |
| Mizrahi 2023, 60+ y | 0.183801 | 0.258614 | 0.1% | 1.20 [0.72, 2.00] |
| Murata 2024, alpha variant | 0.81093 | 0.727984 | 0.0% | 2.25 [0.54, 9.37] |
| Murata 2024, delta variant | -0.19845 | 1.016738 | 0.0% | 0.82 [0.11, 6.02] |
| Murata 2024, omicron BA.1/BA.2 variant | 0.139762 | 0.263712 | 0.1% | 1.15 [0.69, 1.93] |
| Murata 2024, omicron BA.5 variant | -0.15082 | 0.22717 | 0.2% | 0.86 [0.55, 1.34] |
| Taquet 2022 | 0.062881 | 0.009486 | 97.4% | 1.06 [1.05, 1.08] |
| **Subtotal (95% CI)** | | | **97.8%** | **1.06 [1.05, 1.08]** |
| Heterogeneity: Tau² = 0.00; Chi² = 2.31, df = 5 (P = 0.80); I² = 0% | | | | |
| Test for overall effect: Z = 6.64 (P < 0.00001) | | | | |
| **8.3.2 Other** | | | | |
| Jacob 2022, 61-70 y | 0.14842 | 0.148535 | 0.4% | 1.16 [0.87, 1.55] |
| Jacob 2022, 70+ y | 0.131028 | 0.119899 | 0.6% | 1.14 [0.90, 1.44] |
| Smith 2024, 60+ y | 0.223144 | 0.085976 | 1.2% | 1.25 [1.06, 1.48] |
| **Subtotal (95% CI)** | | | **2.2%** | **1.20 [1.06, 1.36]** |
| Heterogeneity: Tau² = 0.00; Chi² = 0.46, df = 2 (P = 0.79); I² = 0% | | | | |
| Test for overall effect: Z = 2.91 (P = 0.004) | | | | |
| **Total (95% CI)** | | | **100.0%** | **1.07 [1.05, 1.09]** |
| Heterogeneity: Tau² = 0.00; Chi² = 6.36, df = 8 (P = 0.61); I² = 0% | | | | |
| Test for overall effect: Z = 7.00 (P < 0.00001) | | | | |
| Test for subgroup differences: Chi² = 3.59, df = 1 (P = 0.06), I² = 72.1% | | | | |

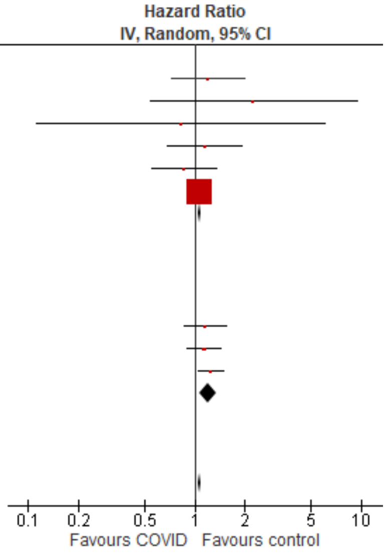

#### *Neurological disorders, overall*

#### Outpatient/mixed, ≥65 y

| Study or Subgroup | log[Hazard Ratio] | SE | Weight | Hazard Ratio IV, Random, 95% CI |
|---|---|---|---|---|
| **9.1.1 Tested +ve** | | | | |
| Donnachie 2022, 60+ y | 0.135997 | 0.049883 | 6.1% | 1.15 [1.04, 1.26] |
| Gronkjaer 2023, 60-69 y | 0.113329 | 0.094061 | 6.0% | 1.12 [0.93, 1.35] |
| Gronkjaer 2023, 70-79 y | 0.131028 | 0.060436 | 6.1% | 1.14 [1.01, 1.28] |
| Gronkjaer 2023, 80+ y | 0.182322 | 0.070304 | 6.1% | 1.20 [1.05, 1.38] |
| Mizrahi 2023, 60+ y | 0.023683 | 0.405044 | 4.3% | 1.02 [0.46, 2.26] |
| Nielsen 2024 | 0.052315 | 0.148244 | 5.8% | 1.05 [0.79, 1.41] |
| Taquet 2022 | 0.091728 | 0.045347 | 6.1% | 1.10 [1.00, 1.20] |
| Wang 2022a | 0.524729 | 0.029861 | 6.2% | 1.69 [1.59, 1.79] |
| Westman 2022, 100+ y | 0.004689 | 1.997988 | 0.5% | 1.00 [0.02, 50.43] |
| Westman 2022, 61-80y | 0.24686 | 0.118502 | 5.9% | 1.28 [1.01, 1.61] |
| Westman 2022, 81-100 y | 0.29267 | 0.163323 | 5.8% | 1.34 [0.97, 1.85] |
| Xu 2022 | 0.34359 | 0.023621 | 6.2% | 1.41 [1.35, 1.48] |
| Zang 2023, INSIGHT cohort | 0.320368 | 0.078479 | 6.1% | 1.38 [1.18, 1.61] |
| Zang 2023, OneFlorida+ cohort | 2.734368 | 0.062372 | 6.1% | 15.40 [13.63, 17.40] |
| Zarifkar 2022, 60-79 y | 0.779905 | 0.582952 | 3.2% | 2.18 [0.70, 6.84] |
| Zarifkar 2022, 80+ y | 0.267583 | 0.688153 | 2.7% | 1.31 [0.34, 5.03] |
| **Subtotal (95% CI)** | | | **83.3%** | **1.52 [1.09, 2.11]** |
| Heterogeneity: Tau² = 0.39; Chi² = 1539.73, df = 15 (P < 0.00001); I² = 99% | | | | |
| Test for overall effect: Z = 2.47 (P = 0.01) | | | | |
| **9.1.2 Other** | | | | |
| Cohen 2022 | 0.793707 | 0.232087 | 5.4% | 2.21 [1.40, 3.49] |
| Gollop 2023 | 0.029559 | 0.106419 | 6.0% | 1.03 [0.84, 1.27] |
| Wan 2023a, UK cohort | 0.09531 | 0.240453 | 5.3% | 1.10 [0.69, 1.76] |
| **Subtotal (95% CI)** | | | **16.7%** | **1.32 [0.84, 2.09]** |
| Heterogeneity: Tau² = 0.13; Chi² = 9.02, df = 2 (P = 0.01); I² = 78% | | | | |
| Test for overall effect: Z = 1.21 (P = 0.23) | | | | |
| **Total (95% CI)** | | | **100.0%** | **1.49 [1.10, 2.01]** |
| Heterogeneity: Tau² = 0.38; Chi² = 1558.58, df = 18 (P < 0.00001); I² = 99% | | | | |
| Test for overall effect: Z = 2.61 (P = 0.009) | | | | |
| Test for subgroup differences: Chi² = 0.22, df = 1 (P = 0.64), I² = 0% | | | | |

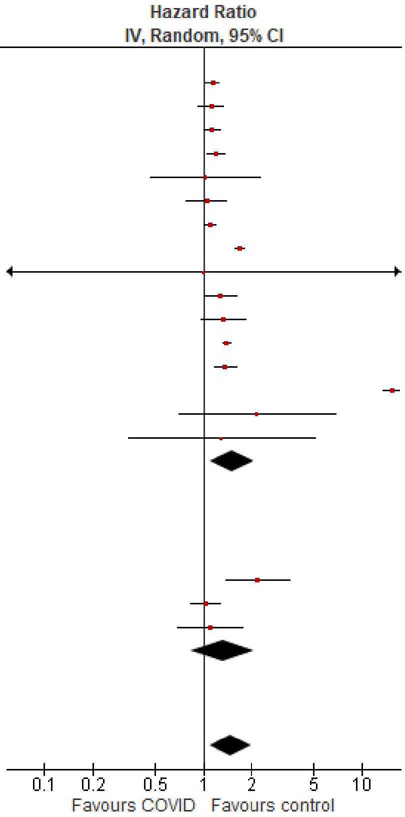

#### *Neurological disorders, dementia/mild cognitive impairment*

##### Outpatient/mixed ≥65 y

| Study or Subgroup | log[Hazard Ratio] | SE | Weight | Hazard Ratio IV, Random, 95% CI |
|---|---|---|---|---|
| **10.1.1 Tested +ve** | | | | |
| Donnachie 2022, 60+ y | 0.166107 | 0.060209 | 8.2% | 1.18 [1.05, 1.33] |
| Murata 2024, alpha variant | -0.31471 | 1.389474 | 0.3% | 0.73 [0.05, 11.12] |
| Murata 2024, delta variant | 1.811562 | 0.422699 | 2.2% | 6.12 [2.67, 14.01] |
| Murata 2024, omicron BA.1/BA.2 variant | 0.71295 | 0.200632 | 5.3% | 2.04 [1.38, 3.02] |
| Murata 2024, omicron BA.5 variant | 0.029559 | 0.183813 | 5.6% | 1.03 [0.72, 1.48] |
| Nielsen 2024 | -0.31471 | 0.049074 | 8.4% | 0.73 [0.66, 0.80] |
| Taquet 2022 | 0.289231 | 0.014765 | 8.7% | 1.34 [1.30, 1.37] |
| Wang 2022a | 0.524729 | 0.029861 | 8.6% | 1.69 [1.59, 1.79] |
| Xu 2022 | 0.512824 | 0.048739 | 8.4% | 1.67 [1.52, 1.84] |
| Zang 2023, INSIGHT cohort | 0.360747 | 0.053668 | 8.3% | 1.43 [1.29, 1.59] |
| Zang 2023, OneFlorida+ cohort | 0.352838 | 0.065038 | 8.1% | 1.42 [1.25, 1.62] |
| Zarifkar 2022, 60-79 y | 0.832909 | 0.334979 | 3.1% | 2.30 [1.19, 4.43] |
| Zarifkar 2022, 80+ y | 0.262364 | 0.203701 | 5.2% | 1.30 [0.87, 1.94] |
| **Subtotal (95% CI)** | | | **80.3%** | **1.41 [1.21, 1.64]** |
| Heterogeneity: Tau² = 0.06; Chi² = 261.67, df = 12 (P < 0.00001); I² = 95% | | | | |
| Test for overall effect: Z = 4.35 (P < 0.0001) | | | | |
| **10.1.2 Other** | | | | |
| Cohen 2022 | 0.81978 | 0.081838 | 7.8% | 2.27 [1.93, 2.66] |
| Gollop 2023 | 0.029559 | 0.106419 | 7.3% | 1.03 [0.84, 1.27] |
| Wan 2023a, UK cohort | 0.09531 | 0.240453 | 4.5% | 1.10 [0.69, 1.76] |
| **Subtotal (95% CI)** | | | **19.7%** | **1.39 [0.76, 2.55]** |
| Heterogeneity: Tau² = 0.26; Chi² = 37.64, df = 2 (P < 0.00001); I² = 95% | | | | |
| Test for overall effect: Z = 1.07 (P = 0.29) | | | | |
| **Total (95% CI)** | | | **100.0%** | **1.42 [1.23, 1.63]** |
| Heterogeneity: Tau² = 0.06; Chi² = 308.26, df = 15 (P < 0.00001); I² = 95% | | | | |
| Test for overall effect: Z = 4.76 (P < 0.00001) | | | | |
| Test for subgroup differences: Chi² = 0.00, df = 1 (P = 0.97), I² = 0% | | | | |

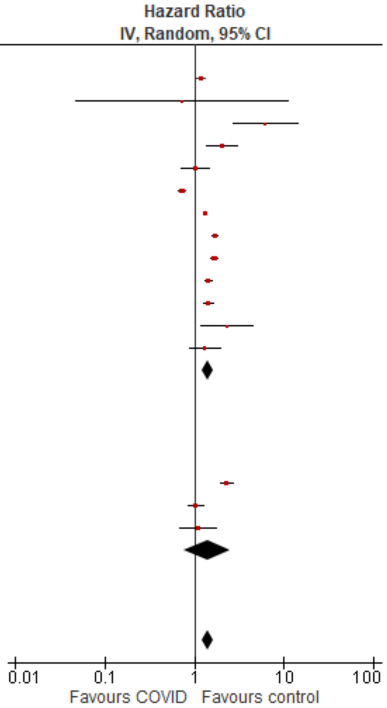

#### *Respiratory disorders, overall*

##### Outpatient/mixed, 18-64 y

| Study or Subgroup | log[Hazard Ratio] | SE | Weight | Hazard Ratio IV, Random, 95% CI |
|---|---|---|---|---|
| **11.1.1 Tested +ve** | | | | |
| Mizrahi 2023, 19-40 y | -0.4943 | 0.543782 | 4.7% | 0.61 [0.21, 1.77] |
| Mizrahi 2023, 41-60 y | 0.193874 | 0.247303 | 9.7% | 1.21 [0.75, 1.97] |
| Mizrahi 2023, 60+ y | -0.01137 | 1.620559 | 0.8% | 0.99 [0.04, 23.69] |
| Oh 2024, Japan cohort, 20-39 y | 1.068153 | 0.039324 | 13.4% | 2.91 [2.69, 3.14] |
| Oh 2024, Japan cohort, 40-59 y | 0.947789 | 0.031681 | 13.4% | 2.58 [2.42, 2.75] |
| Oh 2024, Japan cohort, 60+ y | 0.727549 | 0.178137 | 11.2% | 2.07 [1.46, 2.93] |
| Robertson 2024, BMI 20 to 25 | 0.038949 | 0.162655 | 11.6% | 1.04 [0.76, 1.43] |
| Zang 2023, INSIGHT cohort | 0.173953 | 0.069947 | 13.1% | 1.19 [1.04, 1.36] |
| **Subtotal (95% CI)** | | | **77.9%** | **1.63 [1.20, 2.21]** |
| Heterogeneity: Tau² = 0.14; Chi² = 171.19, df = 7 (P < 0.00001); I² = 96% | | | | |
| Test for overall effect: Z = 3.12 (P = 0.002) | | | | |
| **11.1.2 Other** | | | | |
| Daugherty 2021 | 2.190555 | 0.254895 | 9.6% | 8.94 [5.42, 14.73] |
| Meng 2024 | 0.410333 | 0.110206 | 12.5% | 1.51 [1.21, 1.87] |
| **Subtotal (95% CI)** | | | **22.1%** | **3.62 [0.63, 20.70]** |
| Heterogeneity: Tau² = 1.55; Chi² = 41.10, df = 1 (P < 0.00001); I² = 98% | | | | |
| Test for overall effect: Z = 1.44 (P = 0.15) | | | | |
| **Total (95% CI)** | | | **100.0%** | **1.89 [1.42, 2.51]** |
| Heterogeneity: Tau² = 0.16; Chi² = 215.47, df = 9 (P < 0.00001); I² = 96% | | | | |
| Test for overall effect: Z = 4.37 (P < 0.0001) | | | | |
| Test for subgroup differences: Chi² = 0.78, df = 1 (P = 0.38), I² = 0% | | | | |

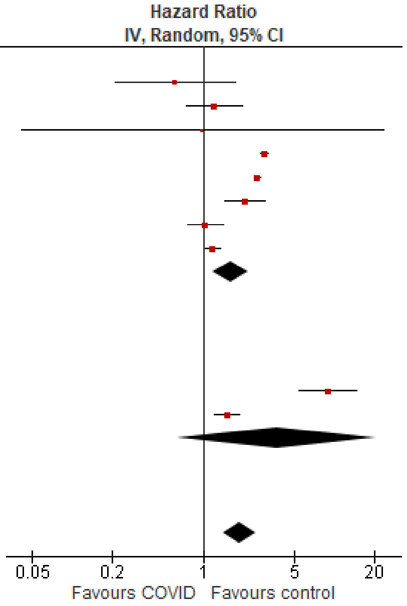

#### Outpatient/mixed ≥65 y

| Study or Subgroup | log[Hazard Ratio] | SE | Weight | Hazard Ratio IV, Random, 95% CI |
|---|---|---|---|---|
| **11.2.1 Tested +ve** | | | | |
| Mizrahi 2023, 60+ y | 0.00878 | 0.387936 | 9.8% | 1.01 [0.47, 2.16] |
| Oh 2024, Japan cohort | 0.530628 | 0.206854 | 15.3% | 1.70 [1.13, 2.55] |
| Oh 2024, South Korea cohort | 0.727549 | 0.077745 | 18.9% | 2.07 [1.78, 2.41] |
| Zang 2023, INSIGHT cohort | 0.280733 | 0.050088 | 19.3% | 1.32 [1.20, 1.46] |
| **Subtotal (95% CI)** | | | **63.3%** | **1.57 [1.13, 2.16]** |
| Heterogeneity: $Tau^2 = 0.08$; $Chi^2 = 24.75$, df = 3 (P < 0.0001); $I^2 = 88\%$ | | | | |
| Test for overall effect: Z = 2.73 (P = 0.006) | | | | |
| **11.2.2 Other** | | | | |
| Cohen 2022 | 1.2947 | 0.108294 | 18.2% | 3.65 [2.95, 4.51] |
| Meng 2024 | 0.521952 | 0.097869 | 18.5% | 1.69 [1.39, 2.04] |
| **Subtotal (95% CI)** | | | **36.7%** | **2.48 [1.16, 5.28]** |
| Heterogeneity: $Tau^2 = 0.29$; $Chi^2 = 28.03$, df = 1 (P < 0.00001); $I^2 = 96\%$ | | | | |
| Test for overall effect: Z = 2.35 (P = 0.02) | | | | |
| **Total (95% CI)** | | | **100.0%** | **1.83 [1.31, 2.56]** |
| Heterogeneity: $Tau^2 = 0.15$; $Chi^2 = 82.86$, df = 5 (P < 0.00001); $I^2 = 94\%$ | | | | |
| Test for overall effect: Z = 3.54 (P = 0.0004) | | | | |
| Test for subgroup differences: $Chi^2 = 1.19$, df = 1 (P = 0.27), $I^2 = 16.2\%$ | | | | |

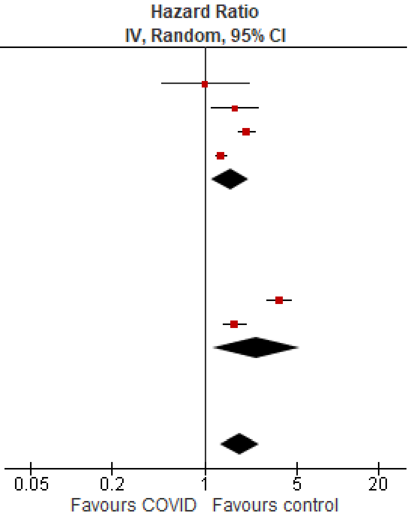

##### *Sleep disorders, overall*

#### Outpatient/mixed ≥65 y

| Study or Subgroup | log[Hazard Ratio] | SE | Weight | Hazard Ratio IV, Random, 95% CI |
|---|---|---|---|---|
| **12.2.1 Tested +ve** | | | | |
| Lin 2024 | 0.405465 | 0.008533 | 16.4% | 1.50 [1.48, 1.53] |
| Mizrahi 2023, 60+ y | -1.20397 | 0.724736 | 1.2% | 0.30 [0.07, 1.24] |
| Murata 2024, alpha variant | -0.82098 | 1.002182 | 0.6% | 0.44 [0.06, 3.14] |
| Murata 2024, delta variant | -0.37106 | 0.166409 | 9.7% | 0.69 [0.50, 0.96] |
| Murata 2024, omicron BA.1/BA.2 variant | -0.54473 | 0.126888 | 11.7% | 0.58 [0.45, 0.74] |
| Murata 2024, omicron BA.5 variant | 0.322083 | 0.451043 | 2.7% | 1.38 [0.57, 3.34] |
| Taquet 2022 | 0.14842 | 0.026413 | 16.2% | 1.16 [1.10, 1.22] |
| Zang 2023, INSIGHT cohort | 0.359142 | 0.046804 | 15.6% | 1.43 [1.31, 1.57] |
| Zarifkar 2022, 60-79 y | 1.196767 | 0.933663 | 0.7% | 3.31 [0.53, 20.63] |
| Zarifkar 2022, 80+ y | 0.989169 | 0.979509 | 0.7% | 2.69 [0.39, 18.34] |
| **Subtotal (95% CI)** | | | **75.5%** | **1.07 [0.88, 1.29]** |
| Heterogeneity: $Tau^2 = 0.04$; $Chi^2 = 166.44$, df = 9 (P < 0.00001); $I^2 = 95\%$ | | | | |
| Test for overall effect: Z = 0.66 (P = 0.51) | | | | |
| **12.2.2 Other** | | | | |
| Cohen 2022 | 0.405465 | 0.094435 | 13.4% | 1.50 [1.25, 1.80] |
| Wan 2023a, UK cohort | 0.336472 | 0.139424 | 11.1% | 1.40 [1.07, 1.84] |
| **Subtotal (95% CI)** | | | **24.5%** | **1.47 [1.26, 1.71]** |
| Heterogeneity: $Tau^2 = 0.00$; $Chi^2 = 0.17$, df = 1 (P = 0.68); $I^2 = 0\%$ | | | | |
| Test for overall effect: Z = 4.91 (P < 0.00001) | | | | |
| **Total (95% CI)** | | | **100.0%** | **1.15 [0.98, 1.35]** |
| Heterogeneity: $Tau^2 = 0.04$; $Chi^2 = 166.62$, df = 11 (P < 0.00001); $I^2 = 93\%$ | | | | |
| Test for overall effect: Z = 1.76 (P = 0.08) | | | | |
| Test for subgroup differences: $Chi^2 = 6.61$, df = 1 (P = 0.01), $I^2 = 84.9\%$ | | | | |

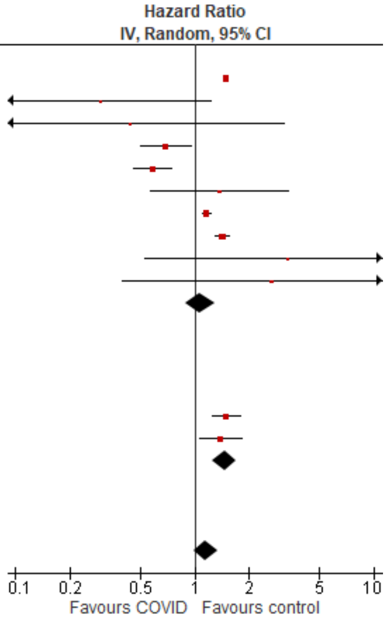

*Stroke, overall*

Outpatient/mixed, 18-64 y

| Study or Subgroup | log[Hazard Ratio] | SE | Weight | Hazard Ratio IV, Random, 95% CI |
|---|---|---|---|---|
| **13.1.1 Tested +ve** | | | | |
| Lam 2023 | -0.24846 | 0.391448 | 2.7% | 0.78 [0.36, 1.68] |
| Lim 2023 | 0.48858 | 0.135964 | 5.8% | 1.63 [1.25, 2.13] |
| Mizrahi 2023, 19-40 y | -1.34707 | 0.772582 | 1.0% | 0.26 [0.06, 1.18] |
| Mizrahi 2023, 41-60 y | -0.04602 | 0.413093 | 2.5% | 0.96 [0.42, 2.15] |
| Mizrahi 2023, 60+ y | 0.198851 | 0.918382 | 0.7% | 1.22 [0.20, 7.38] |
| Nielsen 2024, 18-39 y | 0.19062 | 0.123234 | 6.0% | 1.21 [0.95, 1.54] |
| Nielsen 2024, 40-64 y | -0.21072 | 0.037399 | 6.8% | 0.81 [0.75, 0.87] |
| Taquet 2022 | -0.10536 | 0.014369 | 6.8% | 0.90 [0.88, 0.93] |
| Valasquez Garcia 2024 | 0.323025 | 0.121986 | 6.0% | 1.38 [1.09, 1.75] |
| Wang 2022b, 20-44y | 0.267734 | 0.064357 | 6.6% | 1.31 [1.15, 1.48] |
| Wang 2022b, 45-64y | 0.402126 | 0.039324 | 6.7% | 1.49 [1.38, 1.61] |
| Xie 2022b | 0.364643 | 0.054939 | 6.7% | 1.44 [1.29, 1.60] |
| Xu 2022 | 0.470004 | 0.035132 | 6.8% | 1.60 [1.49, 1.71] |
| Zappacosta 2022, 18-30 y | 1.530395 | 1.116326 | 0.5% | 4.62 [0.52, 41.20] |
| Zappacosta 2022, 31-40 y | 0.00995 | 0.537656 | 1.8% | 1.01 [0.35, 2.90] |
| Zappacosta 2022, 41-50 y | -0.30111 | 0.396695 | 2.7% | 0.74 [0.34, 1.61] |
| Zappacosta 2022, 51-60 y | 0.139762 | 0.252027 | 4.2% | 1.15 [0.70, 1.88] |
| Zappacosta 2022, 61-70 y | -0.10536 | 0.71835 | 1.1% | 0.90 [0.22, 3.68] |
| Zarifkar 2022, 18-39 y | -1.87015 | 1.003109 | 0.6% | 0.15 [0.02, 1.10] |
| Zarifkar 2022, 40-59 y | -0.69315 | 0.216147 | 4.7% | 0.50 [0.33, 0.76] |
| Zarifkar 2022, 60-79 y | -0.35667 | 0.188849 | 5.1% | 0.70 [0.48, 1.01] |
| **Subtotal (95% CI)** | | | **85.7%** | **1.08 [0.90, 1.28]** |
| Heterogeneity: Tau² = 0.10; Chi² = 471.43, df = 20 (P < 0.00001); I² = 96% | | | | |
| Test for overall effect: Z = 0.83 (P = 0.41) | | | | |
| **13.1.2 Other** | | | | |
| Batistoni 2024 | 0.494696 | 0.382586 | 2.8% | 1.64 [0.77, 3.47] |
| Daugherty 2021 | 0.900161 | 0.157049 | 5.5% | 2.46 [1.81, 3.35] |
| Rezel-Potts 2022 | -0.26487 | 0.118513 | 6.0% | 0.77 [0.61, 0.97] |
| **Subtotal (95% CI)** | | | **14.3%** | **1.44 [0.60, 3.47]** |
| Heterogeneity: Tau² = 0.55; Chi² = 35.79, df = 2 (P < 0.00001); I² = 94% | | | | |
| Test for overall effect: Z = 0.81 (P = 0.42) | | | | |
| **Total (95% CI)** | | | **100.0%** | **1.11 [0.95, 1.31]** |
| Heterogeneity: Tau² = 0.10; Chi² = 509.93, df = 23 (P < 0.00001); I² = 95% | | | | |
| Test for overall effect: Z = 1.30 (P = 0.19) | | | | |
| Test for subgroup differences: Chi² = 0.41, df = 1 (P = 0.52), I² = 0% | | | | |

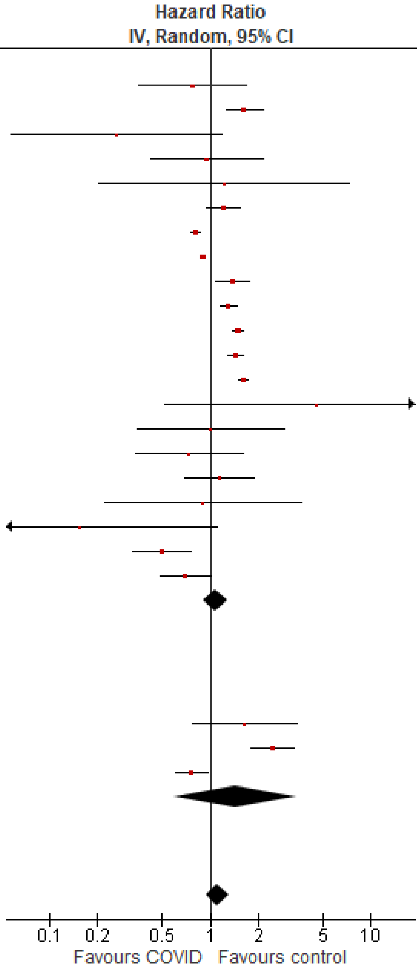

Outpatient/mixed ≥65 y

| Study or Subgroup | log[Hazard Ratio] | SE | Weight | Hazard Ratio IV, Random, 95% CI |
|---|---|---|---|---|
| **13.2.1 Tested +ve** | | | | |
| Mizrahi 2023, 60+ y | 0.410713 | 0.918382 | 0.8% | 1.51 [0.25, 9.12] |
| Nielsen 2024 | -0.17435 | 0.033256 | 9.1% | 0.84 [0.79, 0.90] |
| Taquet 2022 | 0.070552 | 0.014475 | 9.2% | 1.07 [1.04, 1.10] |
| Wang 2022b | 0.535908 | 0.033565 | 9.1% | 1.71 [1.60, 1.83] |
| Xie 2022b | 0.425268 | 0.031617 | 9.1% | 1.53 [1.44, 1.63] |
| Xu 2022 | 0.425268 | 0.058421 | 8.8% | 1.53 [1.36, 1.72] |
| Yang 2024 | 0.002934 | 0.051066 | 8.9% | 1.00 [0.91, 1.11] |
| Zappacosta 2022, 61-70 y | -0.10536 | 0.378604 | 3.3% | 0.90 [0.43, 1.89] |
| Zappacosta 2022, 70+ y | 0.398776 | 0.223997 | 5.6% | 1.49 [0.96, 2.31] |
| Zarifkar 2022, 60-79 y | -0.35667 | 0.119439 | 7.8% | 0.70 [0.55, 0.88] |
| Zarifkar 2022, 80+ y | -0.51083 | 0.119899 | 7.8% | 0.60 [0.47, 0.76] |
| **Subtotal (95% CI)** | | | **79.2%** | **1.09 [0.91, 1.31]** |
| Heterogeneity: Tau² = 0.08; Chi² = 412.48, df = 10 (P < 0.00001); I² = 98% | | | | |
| Test for overall effect: Z = 0.95 (P = 0.34) | | | | |
| **13.2.2 Other** | | | | |
| Batistoni 2024 | 0.463734 | 0.201786 | 6.1% | 1.59 [1.07, 2.36] |
| Cohen 2022 | 0.652325 | 0.076443 | 8.6% | 1.92 [1.65, 2.23] |
| Wan 2023a, UK cohort | 0.09531 | 0.197698 | 6.1% | 1.10 [0.75, 1.62] |
| **Subtotal (95% CI)** | | | **20.8%** | **1.55 [1.11, 2.17]** |
| Heterogeneity: Tau² = 0.06; Chi² = 7.20, df = 2 (P = 0.03); I² = 72% | | | | |
| Test for overall effect: Z = 2.56 (P = 0.01) | | | | |
| **Total (95% CI)** | | | **100.0%** | **1.17 [0.99, 1.39]** |
| Heterogeneity: Tau² = 0.08; Chi² = 459.52, df = 13 (P < 0.00001); I² = 97% | | | | |
| Test for overall effect: Z = 1.88 (P = 0.06) | | | | |
| Test for subgroup differences: Chi² = 3.21, df = 1 (P = 0.07), I² = 68.9% | | | | |

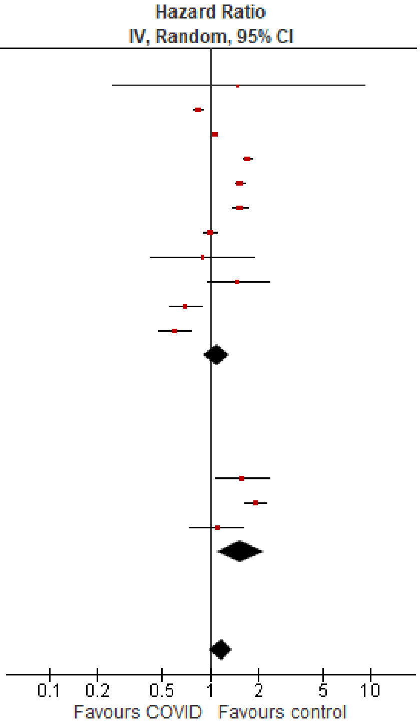

#### Appendix E. Stratified analysis of new diagnoses of chronic conditions after SARS-CoV-2 infection: studies with less than 12 months follow-up vs. ≥12 months

**Summary:** With the exception of psychosis/psychotic disorders in outpatients/mixed patients <18y, no differences in association by follow-up timing were observed in outcomes with at least 2 studies reporting for each sub group. Only outcomes with ≥2 studies contributing to each subgroup are presented.

| Group<br>No. studies | Subgroup<br>No. Studies; weight | Relative findings<br>HR (95% CI) | $I^2$ | P-value for subgroup differences |
|---|---|---|---|---|
| **Autoimmune disorders, overall** | | | | |
| Outpatients/mixed, 18-64 y | Overall, 10 studies | 1.51 (1.31 to 1.75) | 98% | P=0.43 |
| | <12 mos follow-up<br>5 studies; 30.2% | 1.38 (1.06 to 1.80) | 88% | |
| | ≥12 mos follow-up<br>5 studies; 69.8% | 1.57 (1.31 to 1.88) | 99% | |
| Outpatients/mixed, ≥65 y | Overall, 9 studies | 1.44 (1.23 to 1.69) | 89% | P=0.86 |
| | <12 mos follow-up<br>4 studies; 33.9% | 1.47 (1.03 to 2.09) | 85% | |
| | ≥12 mos follow-up<br>5 studies; 66.1% | 1.42 (1.16 to 1.72) | 91% | |
| **Cardiovascular disorders, overall** | | | | |
| Outpatients/mixed, <18 y | Overall, 7 studies | 1.17 (1.13 to 1.21) | 0% | P=0.86 |
| | <12 mos follow-up<br>4 studies; 1.9% | 1.25 (0.98 to 1.60) | 0% | |
| | ≥12 mos follow-up<br>3 studies; 98.1% | 1.29 (1.04 to 1.59) | 66% | |
| Outpatients/mixed, 18-64 y | Overall, 12 studies | 1.32 (1.12 to 1.55) | 92% | P=0.36 |
| | <12 mos follow-up<br>8 studies; 53.1 % | 1.41 (1.17 to 1.71) | 77% | |
| | ≥12 mos follow-up<br>4 studies; 46.9% | 1.21 (0.93 to 1.58) | 96% | |
| Outpatients/mixed, ≥65 y | Overall, 10 studies | 1.33 (1.14 to 1.54) | 96% | P=0.71 |
| | <12 mos follow-up<br>6 studies; 50.4% | 1.36 (1.12 to 1.66) | 84% | |
| | ≥12 mos follow-up<br>4 studies; 49.6% | 1.29 (1.01 to 1.64) | 98% | |
| **Cardiovascular disorders, acute coronary disease** | | | | |
| Outpatients/mixed, <18y | Overall, 4 studies | 1.45 (1.17 to 1.79) | 0% | P=0.31 |
| | <12 mos follow-up<br>2 studies; 75.0% | 1.36 (1.06 to 1.74) | 0% | |
| | ≥12 mos follow-up<br>2 studies; 25.0% | 1.75 (1.14 to 2.69) | 0% | |
| Outpatients/mixed, 18-64 y | Overall, 9 studies | 1.43 (1.14 to 1.81) | 86% | P=0.91 |
| | <12 mos follow-up<br>6 studies; 59.7% | 1.46 (1.00 to 2.12) | 84% | |
| | ≥12 mos follow-up<br>3 studies; 40.3% | 1.42 (1.03 to 1.96) | 91% | |
| Outpatients/mixed, ≥65 y | Overall, 7 studies | 1.55 (1.27 to 1.89) | 94% | P=0.75 |
| | <12 mos follow-up<br>4 studies; 48.7% | 1.64 (1.01 to 2.65) | 91% | |
| | ≥12 mos follow-up<br>3 studies; 51.3% | 1.50 (1.19 to 1.90) | 96% | |
| **Cardiovascular disorders, arrhythmias/dysrhythmias** | | | | |
| Outpatients/mixed, <18y | Overall, 7 studies | 1.28 (1.17 to 1.41) | 67% | P=0.85 |
| | <12 mos follow-up<br>4 studies; 59.2% | 1.28 (1.10 to 1.49) | 62% | |
| | ≥12 mos follow-up<br>3 studies; 40.8% | 1.31 (1.08 to 1.59) | 77% | |
| Outpatients/mixed, 18-64 y | Overall, 8 studies | 1.56 (1.38 to 1.77) | 87% | P=0.69 |

| Group<br>No. studies | Subgroup<br>No. Studies; weight | Relative findings<br>HR (95% CI) | $I^2$ | P-value for subgroup differences |
|---|---|---|---|---|
| | <12 mos follow-up<br>5 studies; 50.2% | 1.47 (1.06 to 2.03) | 86% | |
| | ≥12 mos follow-up<br>3 studies; 49.8% | 1.57 (1.41 to 1.76) | 87% | |
| Outpatients/mixed, ≥65 y | Overall, 8 studies | 1.43 (1.18 to 1.73) | 97% | P=0.35 |
| | <12 mos follow-up<br>5 studies; 55.9% | 1.31 (0.99 to 1.74) | 87% | |
| | ≥12 mos follow-up<br>3 studies; 44.1% | 1.59 (1.18 to 2.15) | 99% | |
| **Cardiovascular disease, heart failure** | | | | |
| Outpatients/mixed, 18-64 y | Overall, 7 studies | 1.81 (1.40 to 2.34) | 94% | P=0.80 |
| | <12 mos follow-up<br>5 studies; 60.0% | 1.84 (1.26 to 2.67) | 90% | |
| | ≥12 mos follow-up<br>2 studies; 40.0% | 1.94 (1.54 to 2.44) | 86% | |
| Outpatients/mixed, ≥65 y | Overall, 6 studies | 1.61 (1.22 to 2.12) | 98% | P=0.89 |
| | <12 mos follow-up<br>4 studies; 64.6% | 1.62 (1.27 to 2.08) | 88% | |
| | ≥12 mos follow-up<br>2 studies; 35.4% | 1.55 (0.86 to 2.81) | 100% | |
| **Diabetes, overall** | | | | |
| Outpatients/mixed, <18y | Overall, 6 studies | 1.10 (0.95 to 1.28) | 59% | P=0.25 |
| | <12 mos follow-up<br>3 studies; 41.4% | 1.26 (0.94 to 1.70) | 57% | |
| | ≥12 mos follow-up<br>3 studies; 58.6% | 1.02 (0.82 to 1.26) | 67% | |
| Outpatients/mixed, 18-64 y | Overall, 10 studies* | 1.53 (1.29 to 1.81) | 98% | P=0.10 |
| | <12 mos follow-up<br>6 studies; 59.1% | 1.71 (1.25 to 2.35) | 98% | |
| | ≥12 mos follow-up<br>5 studies; 40.9% | 1.30 (1.18 to 1.43) | 91% | |
| Outpatients/mixed, ≥65 y | Overall, 6 studies* | 1.61 (1.36 to 1.92) | 94% | P=0.10 |
| | <12 mos follow-up<br>5 studies; 62.6% | 1.80 (1.30 to 2.51) | 95% | |
| | ≥12 mos follow-up<br>2 studies; 37.4% | 1.36 (1.24 to 1.48) | 58% | |
| **Diabetes, type 2** | | | | |
| Outpatients/mixed, 18-64 y | Overall, 3 studies* | 1.35 (1.18 to 1.55) | 85% | P=0.16 |
| | <12 mos follow-up<br>2 studies; 46.7% | 1.52 (1.18 to 1.98) | 85% | |
| | ≥12 mos follow-up<br>2 studies; 53.3% | 1.23 (1.07 to 1.43) | 81% | |
| **Mental illness, overall** | | | | |
| Outpatients/mixed, <18y | Overall, 11 studies | 1.01 (0.88 to 1.16) | 99% | P=0.49 |
| | <12 mos follow-up<br>6 studies; 50.1% | 0.97 (0.82 to 1.14) | 94% | |
| | ≥12 mos follow-up<br>5 studies; 49.9% | 1.06 (0.86 to 1.30) | 100% | |
| Outpatients/mixed, 18-64 y | Overall, 11 studies | 1.24 (1.13 to 1.36) | 99% | P=0.57 |
| | <12 mos follow-up<br>8 studies; 62.7% | 1.28 (1.08 to 1.52) | 97% | |
| | ≥12 mos follow-up<br>4 studies; 37.3% | 1.20 (1.06 to 1.37) | 99% | |
| Outpatients/mixed, ≥65 y | Overall, 13 studies | 1.20 (1.07 to 1.35) | 96% | P=0.55 |
| | <12 mos follow-up<br>9 studies; 70.7% | 1.27 (0.96 to 1.68) | 96% | |
| | ≥12 mos follow-up<br>4 studies; 29.3% | 1.16 (1.05 to 1.28) | 95% | |
| **Mental illness, anxiety/anxiety disorders** | | | | |

| Group<br>No. studies | Subgroup<br>No. Studies; weight | Relative findings<br>HR (95% CI) | $I^2$ | P-value for subgroup differences |
|---|---|---|---|---|
| Outpatients/mixed, <18y | Overall, 6 studies | 0.87 (0.81 to 0.92) | 86% | P=0.11 |
| | <12 mos follow-up<br>3 studies; 25.2% | 0.94 (0.84 to 1.06 | 0% | |
| | ≥12 mos follow-up<br>3 studies; 74.8% | 0.84 (0.78 to 0.91) | 96% | |
| **Mental illness, depression & mood disorders** | | | | |
| Outpatients/mixed, <18y | Overall, 6 studies | 1.37 (0.84 to 2.23) | 100% | P=0.06 |
| | <12 mos follow-up<br>2 studies; 28.2% | 0.75 (0.41 to 1.34) | 35% | |
| | ≥12 mos follow-up<br>4 studies; 71.8% | 1.66 (0.93 to 2.95) | 100% | |
| Outpatients/mixed, 18-64 y | Overall, 5 studies | 1.11 (1.04 to 1.19) | 73% | P=0.22 |
| | <12 mos follow-up<br>3 studies; 52.9% | 1.07 (0.97 to 1.17) | 51% | |
| | ≥12 mos follow-up<br>2 studies; 47.1% | 1.17 (1.04 to 1.33) | 82% | |
| Outpatients/mixed, ≥65 y | Overall, 5 studies | 1.07 (1.05 to 1.09) | 0% | P=0.96 |
| | <12 mos follow-up<br>3 studies; 1.5% | 1.12 (0.96 to 1.30) | 0% | |
| | ≥12 mos follow-up<br>2 studies; 98.5% | 1.13 (0.97 to 1.31) | 71% | |
| **Mental illness, psychosis/psychotic disorders** | | | | |
| Outpatients/mixed, <18y | Overall, 4 studies | 1.23 (0.70 to 2.16) | 89% | P=0.03 |
| | <12 mos follow-up<br>2 studies; 31.0% | 0.42 (0.10 to 1.75) | 77% | |
| | ≥12 mos follow-up<br>2 studies; 69.0% | 2.14 (1.46 to 3.14) | 77% | |
| **Neurological disorders, overall** | | | | |
| Outpatients/mixed, <18y | Overall, 10 studies | 1.24 (1.04 to 1.47) | 86% | P=0.35 |
| | <12 mos follow-up<br>6 studies; 64.0% | 1.35 (1.03 to 1.77) | 82% | |
| | ≥12 mos follow-up<br>4 studies; 36.0% | 1.10 (0.78 to 1.54) | 80% | |
| Outpatients/mixed, 18-64 y | Overall, 10 studies | 1.16 (1.00 to 1.35) | 96% | P=0.68 |
| | <12 mos follow-up<br>5 studies; 56.3% | 1.13 (1.04 to 1.23) | 61% | |
| | ≥12 mos follow-up<br>5 studies; 43.7% | 1.20 (0.90 to 1.60) | 98% | |
| Outpatients/mixed, ≥65 y | Overall, 14 studies | 1.55 (1.18 to 2.04) | 99% | P=0.39 |
| | <12 mos follow-up<br>7 studies; 58.1% | 1.73 (0.90 to 3.34) | 99% | |
| | ≥12 mos follow-up<br>7 studies; 41.9% | 1.29 (1.12 to 1.48) | 90% | |
| **Neurological disorders, communication & motor disorders** | | | | |
| Outpatients/mixed, <18y | Overall, 4 studies | 1.31 (1.06 to 1.61) | 48% | P=0.54 |
| | <12 mos follow-up<br>2 studies; 61.8% | 1.59 (0.78 to 3.27) | 80% | |
| | ≥12 mos follow-up<br>2 studies; 38.2% | 1.26 (1.00 to 1.59 | 0% | |
| Outpatients/mixed, 18-64 y | Overall, 6 studies | 1.46 (0.72 to 2.96) | 99% | P=0.22 |
| | <12 mos follow-up<br>3 studies; 65.4% | 2.02 (0.85 to 4.80) | 97% | |
| | ≥12 mos follow-up<br>3 studies; 34.6% | 0.80 (0.24 to 2.66) | 99% | |
| Outpatients/mixed, ≥65 y | Overall, 7 studies | 1.31 (0.99 to 1.74) | 91% | P=0.35 |
| | <12 mos follow-up<br>4 studies; 50.7% | 1.60 (0.84 to 3.05) | 86% | |
| | ≥12 mos follow-up<br>3 studies; 49.3% | 1.12 (0.78 to 1.61) | 94% | |

| Group<br>No. studies | Subgroup<br>No. Studies; weight | Relative findings<br>HR (95% CI) | $I^2$ | P-value for subgroup differences |
|---|---|---|---|---|
| **Neurological disorders, dementia/mild cognitive impairment** | | | | |
| Outpatients/mixed, 18-64 y | Overall, 7 studies | 1.42 (1.05 to 1.92) | 97% | P=0.13 |
| | <12 mos follow-up<br>3 studies; 40.8% | 1.07 (0.66 to 1.72) | 88% | |
| | ≥12 mos follow-up<br>4 studies; 59.2% | 1.73 (1.16 to 2.56) | 98% | |
| Outpatients/mixed, ≥65 y | Overall, 11 studies | 1.42 (1.23 to 1.63) | 95% | P=0.72 |
| | <12 mos follow-up<br>5 studies; 50.5% | 1.50 (1.06 to 2.10) | 96% | |
| | ≥12 mos follow-up<br>6 studies; 49.5% | 1.40 (1.22 to 1.61) | 93% | |
| **Neurological disorders, epilepsy** | | | | |
| Outpatients/mixed, <18y | Overall, 4 studies | 1.09 (0.61 to 1.95) | 95% | P=0.48 |
| | <12 mos follow-up<br>2 studies; 61.2% | 0.90 (0.58 to 1.40) | 48% | |
| | ≥12 mos follow-up<br>2 studies; 38.8% | 1.31 (0.51 to 3.34) | 98% | |
| Outpatients/mixed, 18-64 y | Overall, 5 studies | 1.05 (0.94 to 1.17) | 89% | P=0.80 |
| | <12 mos follow-up<br>3 studies; 24.8% | 1.08 (0.78 to 1.51) | 55% | |
| | ≥12 mos follow-up<br>2 studies; 75.2% | 1.04 (0.92 to 1.17) | 96% | |
| Outpatients/mixed, ≥65 y | Overall, 5 studies | 1.26 (1.04 to 1.53) | 68% | P=0.72 |
| | <12 mos follow-up<br>3 studies; 41.4% | 1.22 (0.74 to 2.01) | 78% | |
| | ≥12 mos follow-up<br>2 studies; 58.6% | 1.11 (1.07 to 1.16) | 0% | |
| **Neurological disorders, Guillain-Barre syndrome** | | | | |
| Outpatients/mixed, 18-64 y | Overall, 4 studies | 1.18 (1.04 to 1.33) | 0% | P=0.62 |
| | <12 mos follow-up<br>2 studies; 4.8% | 1.52 (0.55 to 4.20) | 44% | |
| | ≥12 mos follow-up<br>2 studies; 95.2% | 1.17 (1.03 to 1.33) | 0% | |
| **Neurological disorders, nerve disorders** | | | | |
| Outpatients/mixed, <18 y | Overall, 4 studies | 1.10 (0.76 to 1.61) | 76% | P=0.70 |
| | <12 mos follow-up<br>2 studies; 57.4% | 0.96 (0.67 to 1.38) | 47% | |
| | ≥12 mos follow-up<br>2 studies 42.6% | 1.17 (0.44 to 3.11) | 42% | |
| Outpatients/mixed, 18-64 y | Overall, 6 studies | 1.08 (0.97 to 1.20) | 94% | P=0.61 |
| | <12 mos follow-up<br>4 studies; 73.0% | 1.06 (0.94 to 1.20) | 84% | |
| | ≥12 mos follow-up<br>2 studies; 27.0% | 1.14 (0.87 to 1.51) | 99% | |
| Outpatients/mixed, ≥65 y | Overall, 6 studies | 1.06 (0.93 to 1.22) | 94% | P=0.43 |
| | <12 mos follow-up<br>4 studies; 61.7% | 1.01 (0.86 to 1.20) | 72% | |
| | ≥12 mos follow-up<br>2 studies; 38.3% | 1.15 (0.89 to 1.48) | 99% | |
| **Respiratory disorders, overall** | | | | |
| Outpatients/mixed, 18-64 y | Overall, 6 studies | 1.89 (1.42 to 2.51) | 96% | P=0.27 |
| | <12 mos follow-up<br>4 studies; 49.4% | 1.56 (0.82 to 2.97) | 92% | |
| | ≥12 mos follow-up<br>2 studies; 50.6% | 2.28 (1.87 to 2.79) | 91% | |
| Outpatients/mixed, ≥65 y | Overall, 5 studies | 1.83 (1.31 to 2.56) | 94% | P=0.89 |
| | <12 mos follow-up<br>3 studies; 47.4% | 1.76 (0.78 to 3.99) | 97% | |

| Group<br>No. studies | Subgroup<br>No. Studies; weight | Relative findings<br>HR (95% CI) | $I^2$ | P-value for subgroup differences |
|---|---|---|---|---|
| | ≥12 mos follow-up<br>2 studies; 52.6% | 1.87 (1.60 to 2.17) | 33% | |
| **Sleep disorders, overall** | | | | |
| Outpatients/mixed, <18y | Overall, 6 studies | 0.97 (0.91 to 1.04) | 81% | P=0.49 |
| | <12 mos follow-up<br>3 studies; 5.1% | 1.12 (0.75 to 1.68) | 18% | |
| | ≥12 mos follow-up<br>3 studies; 94.9% | 0.97 (0.91 to 1.04) | 89% | |
| Outpatients/mixed, 18-64 y | Overall, 6 studies | 1.52 (1.23 to 1.87) | 88% | P=0.45 |
| | <12 mos follow-up<br>4 studies; 97.5% | 1.54 (1.24 to 1.91) | 93% | |
| | ≥12 mos follow-up<br>2 studies; 2.5% | 0.92 (0.24 to 3.45) | 0% | |
| Outpatients/mixed, ≥65 y | Overall, 8 studies | 1.15 (0.98 to 1.35) | 93% | P=0.71 |
| | <12 mos follow-up<br>6 studies; 82.5% | 1.12 (0.92 to 1.36) | 91% | |
| | ≥12 mos follow-up<br>2 studies; 17.5% | 1.16 (1.10 to 1.22) | 0% | |
| **Stroke, overall** | | | | |
| Outpatients/mixed, <18y | Overall, 5 studies | 1.14 (0.99 to 1.30) | 0% | P=0.30 |
| | <12 mos follow-up<br>2 studies; 3.7% | 0.79 (0.40 to 1.59) | 0% | |
| | ≥12 mos follow-up<br>3 studies; 96.3% | 1.15 (1.01 to 1.32) | 0% | |
| Outpatients/mixed, 18-64 y | Overall, 14 studies | 1.11 (0.95 to 1.31) | 95% | P=0.64 |
| | <12 mos follow-up<br>8 studies; 45.8% | 1.16 (0.87 to 1.54) | 89% | |
| | ≥12 mos follow-up<br>6 studies; 54.2% | 1.06 (0.84 to 1.34) | 97% | |
| Outpatients/mixed, ≥65 y | Overall, 12 studies | 1.17 (0.99 to 1.39) | 97% | P=0.73 |
| | <12 mos follow-up<br>6 studies; 39.5% | 1.22 (0.89 to 1.68) | 95% | |
| | ≥12 mos follow-up<br>6 studies; 60.5% | 1.15 (0.92 to 1.42) | 98% | |
| **Stroke, ischemic stroke** | | | | |
| Outpatients/mixed, ≥65 y | Overall, 4 studies | 0.98 (0.76 to 1.28) | 96% | P=0.15 |
| | <12 mos follow-up<br>2 studies; 41.1% | 1.37 (0.71 to 2.64) | 98% | |
| | ≥12 mos follow-up<br>2 studies; 58.9% | 0.78 (0.52 to 1.17) | 94% | |

* includes study contributing to more than one subgroup

#### Forest Plots for stratified analysis by follow-up duration (<12 mos vs. ≥12 mos)

##### *Autoimmune disorders, overall*

###### Outpatients/mixed, 18-64 y

| Study or Subgroup | log[Hazard Ratio] | SE | Weight | Hazard Ratio IV, Random, 95% CI |
|---|---|---|---|---|
| **1.2.1 Autoimmune disorders, overall (outpatients/mixed, 18-64 y), <12 mo follow-up** | | | | |
| Chang 2023, 18-40 y | 0.656266 | 0.123921 | 4.7% | 1.93 [1.51, 2.46] |
| Chang 2023, 41-64 y | 0.946803 | 0.098617 | 5.0% | 2.58 [2.12, 3.13] |
| Daugherty 2021 | 0.215111 | 0.10761 | 4.9% | 1.24 [1.00, 1.53] |
| Lim 2023 | 0.04697 | 0.219831 | 3.7% | 1.05 [0.68, 1.61] |
| Mizrahi 2023, 19-40 y | -0.26379 | 0.240527 | 3.5% | 0.77 [0.48, 1.23] |
| Mizrahi 2023, 41-60 y | 0.128753 | 0.302499 | 2.9% | 1.14 [0.63, 2.06] |
| Mizrahi 2023, 60+ y | -0.82098055 | 1.764631 | 0.2% | 0.44 [0.01, 13.98] |
| Schmitt 2024 | 0.262364 | 0.031437 | 5.4% | 1.30 [1.22, 1.38] |
| **Subtotal (95% CI)** | | | **30.2%** | **1.38 [1.06, 1.80]** |
| Heterogeneity: Tau² = 0.10; Chi² = 60.32, df = 7 (P < 0.00001); I² = 88% | | | | |
| Test for overall effect: Z = 2.38 (P = 0.02) | | | | |
| **1.2.2 Autoimmune disorders, overall (outpatients/mixed, 18-64 y), 12+ mo follow-up** | | | | |
| Huang 2024 | 0.678034 | 0.013018 | 5.4% | 1.97 [1.92, 2.02] |
| Kim 2024, 20-39 y | 0.058269 | 0.040807 | 5.3% | 1.06 [0.98, 1.15] |
| Kim 2024, 40-59 y | 0.139762 | 0.019888 | 5.4% | 1.15 [1.11, 1.20] |
| Kyung 2024, Japan cohort, 20-39 y | 0.300105 | 0.078828 | 5.1% | 1.35 [1.16, 1.58] |
| Kyung 2024, Japan cohort, 40-59 y | 0.329304 | 0.060195 | 5.2% | 1.39 [1.24, 1.56] |
| Kyung 2024, South Korea cohort, 20-39 y | 0.405465 | 0.16005 | 4.3% | 1.50 [1.10, 2.05] |
| Kyung 2024, South Korea cohort, 40-59y | 0.756122 | 0.126909 | 4.7% | 2.13 [1.66, 2.73] |
| Oh 2024, Japan cohort, 20-39 y | 0.95958 | 0.032756 | 5.4% | 2.61 [2.45, 2.78] |
| Oh 2024, Japan cohort, 40-59 y | 0.945678 | 0.025499 | 5.4% | 2.57 [2.45, 2.71] |
| Oh 2024, Japan cohort, 60+ y | 0.747603 | 0.118029 | 4.8% | 2.11 [1.68, 2.66] |
| Oh 2024, South Korea cohort, 20-39 y | 0.211086 | 0.129947 | 4.7% | 1.24 [0.96, 1.59] |
| Oh 2024, South Korea cohort, 40-59 y | 0.18091 | 0.094441 | 5.0% | 1.20 [1.00, 1.44] |
| Oh 2024, South Korea cohort, 60+ y | 0.183624 | 0.217209 | 3.7% | 1.20 [0.78, 1.84] |
| Tesch 2023 | 0.364643 | 0.023126 | 5.4% | 1.44 [1.38, 1.51] |
| **Subtotal (95% CI)** | | | **69.8%** | **1.57 [1.31, 1.88]** |
| Heterogeneity: Tau² = 0.11; Chi² = 1183.00, df = 13 (P < 0.00001); I² = 99% | | | | |
| Test for overall effect: Z = 4.94 (P < 0.00001) | | | | |
| **Total (95% CI)** | | | **100.0%** | **1.51 [1.31, 1.75]** |
| Heterogeneity: Tau² = 0.10; Chi² = 1303.50, df = 21 (P < 0.00001); I² = 98% | | | | |
| Test for overall effect: Z = 5.50 (P < 0.00001) | | | | |
| Test for subgroup differences: Chi² = 0.63, df = 1 (P = 0.43), I² = 0% | | | | |

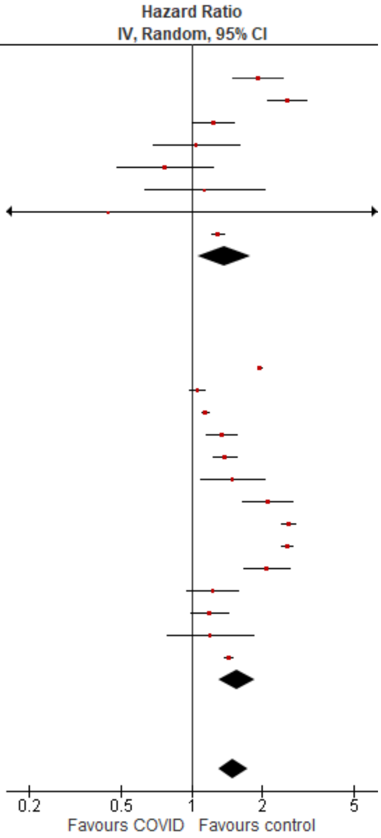

###### Outpatients/mixed, ≥65 y

| Study or Subgroup | log[Hazard Ratio] | SE | Weight | Hazard Ratio IV, Random, 95% CI |
|---|---|---|---|---|
| **1.3.1 Autoimmune disorders, overall (outpatients/mixed, ≥65 y), <12 mo follow-up** | | | | |
| Chang 2023 | 1.070937 | 0.158296 | 7.6% | 2.92 [2.14, 3.98] |
| Cohen 2022 | -0.03046 | 0.180547 | 7.0% | 0.97 [0.68, 1.38] |
| Mizrahi 2023, 60+ y | -0.82098 | 0.770148 | 1.0% | 0.44 [0.10, 1.99] |
| Schmitt 2024, 65-79 y | 0.350657 | 0.093141 | 9.3% | 1.42 [1.18, 1.70] |
| Schmitt 2024, 80+ y | 0.371564 | 0.104872 | 9.0% | 1.45 [1.18, 1.78] |
| **Subtotal (95% CI)** | | | **33.9%** | **1.47 [1.03, 2.09]** |
| Heterogeneity: Tau² = 0.12; Chi² = 26.48, df = 4 (P < 0.0001); I² = 85% | | | | |
| Test for overall effect: Z = 2.13 (P = 0.03) | | | | |
| **1.3.2 Autoimmune disorders, overall (outpatients/mixed, ≥65 y), 12+ mo follow-up** | | | | |
| Huang 2024 | 0.779325 | 0.191731 | 6.7% | 2.18 [1.50, 3.17] |
| Kim 2024 | 0.14842 | 0.044135 | 10.2% | 1.16 [1.06, 1.26] |
| Kyung 2024, Japan cohort | 0.322083 | 0.17357 | 7.2% | 1.38 [0.98, 1.94] |
| Kyung 2024, South Korea cohort | -0.56212 | 0.390963 | 3.1% | 0.57 [0.26, 1.23] |
| Oh 2024, Japan cohort | 0.747603 | 0.057592 | 10.0% | 2.11 [1.89, 2.36] |
| Oh 2024, South Korea cohort | 0.183624 | 0.105987 | 9.0% | 1.20 [0.98, 1.48] |
| Tesch 2023, 65-79 y | 0.350657 | 0.052096 | 10.1% | 1.42 [1.28, 1.57] |
| Tesch 2023, 80+ y | 0.300105 | 0.067926 | 9.8% | 1.35 [1.18, 1.54] |
| **Subtotal (95% CI)** | | | **66.1%** | **1.42 [1.16, 1.72]** |
| Heterogeneity: Tau² = 0.06; Chi² = 82.00, df = 7 (P < 0.00001); I² = 91% | | | | |
| Test for overall effect: Z = 3.48 (P = 0.0005) | | | | |
| **Total (95% CI)** | | | **100.0%** | **1.44 [1.23, 1.69]** |
| Heterogeneity: Tau² = 0.06; Chi² = 109.46, df = 12 (P < 0.00001); I² = 89% | | | | |
| Test for overall effect: Z = 4.45 (P < 0.00001) | | | | |
| Test for subgroup differences: Chi² = 0.03, df = 1 (P = 0.86), I² = 0% | | | | |

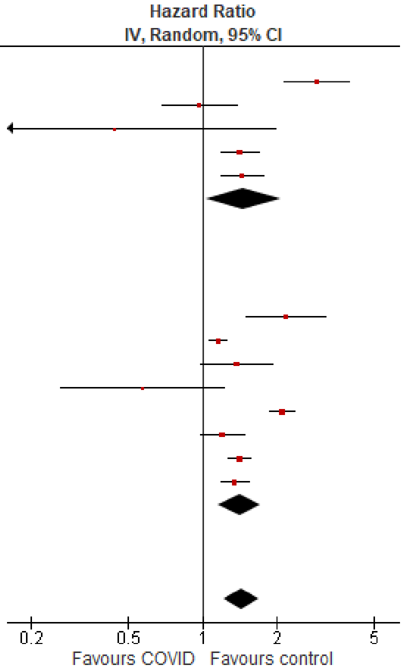

*Cardiovascular disorders, overall*

#### Outpatients/mixed, <18 y

| Study or Subgroup | log[Hazard Ratio] | SE | Weight | Hazard Ratio IV, Random, 95% CI |
|---|---|---|---|---|
| **2.1.1 Cardiovascular disorders, overall (outpatients/mixed, <18 y), <12 mo follow-up** | | | | |
| Gordon 2024, 0-1 y | 0.071479 | 0.21927 | 0.6% | 1.07 [0.70, 1.65] |
| Gordon 2024, 12-17 y | 0.237643 | 2.813738 | 0.0% | 1.27 [0.01, 314.97] |
| Gordon 2024, 2-4 y | 0.36921 | 2.813829 | 0.0% | 1.45 [0.01, 359.32] |
| Gordon 2024, 5-11 y | 0.321619 | 0.173146 | 1.0% | 1.38 [0.98, 1.94] |
| Mizrahi 2023, 12-18 y | -0.05374 | 0.386854 | 0.2% | 0.95 [0.44, 2.02] |
| Tartof 2022 | 0.576613 | 0.624046 | 0.1% | 1.78 [0.52, 6.05] |
| Valdivieso-Martinez 2023, unvaccinated cohort | 1.098612 | 1.154739 | 0.0% | 3.00 [0.31, 28.84] |
| Valdivieso-Martinez 2023, vaccinated cohort | 1.94591 | 3.75576 | 0.0% | 7.00 [0.00, 11015.56] |
| **Subtotal (95% CI)** | | | **1.9%** | **1.25 [0.98, 1.60]** |
| Heterogeneity: Tau² = 0.00; Chi² = 2.42, df = 7 (P = 0.93); I² = 0% | | | | |
| Test for overall effect: Z = 1.79 (P = 0.07) | | | | |
| **2.1.2 Cardiovascular disorders, overall (outpatients/mixed, <18 y), 12+ mo follow-up** | | | | |
| Kompaniyets 2022 | 0.14842 | 0.0176 | 95.2% | 1.16 [1.12, 1.20] |
| Roessler 2022 | 0.355874 | 0.624046 | 0.1% | 1.43 [0.42, 4.85] |
| Zhang 2024 | 0.398109 | 0.102964 | 2.8% | 1.49 [1.22, 1.82] |
| **Subtotal (95% CI)** | | | **98.1%** | **1.29 [1.04, 1.59]** |
| Heterogeneity: Tau² = 0.02; Chi² = 5.82, df = 2 (P = 0.05); I² = 66% | | | | |
| Test for overall effect: Z = 2.30 (P = 0.02) | | | | |
| **Total (95% CI)** | | | **100.0%** | **1.17 [1.13, 1.21]** |
| Heterogeneity: Tau² = 0.00; Chi² = 8.53, df = 10 (P = 0.58); I² = 0% | | | | |
| Test for overall effect: Z = 9.14 (P < 0.00001) | | | | |
| Test for subgroup differences: Chi² = 0.03, df = 1 (P = 0.86), I² = 0% | | | | |

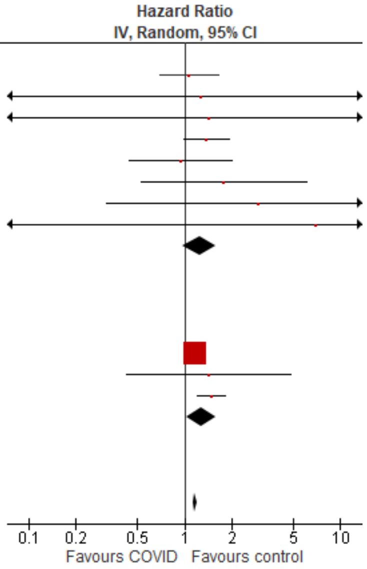

#### Outpatients/mixed, 18-64 y

| Study or Subgroup | log[Hazard Ratio] | SE | Weight | Hazard Ratio IV, Random, 95% CI |
|---|---|---|---|---|
| **2.2.1 Cardiovascular disorders, overall (outpatients/mixed, 18-64 y), <12 mo follow-up** | | | | |
| Batistoni 2024 | 0.836308 | 0.216484 | 4.5% | 2.31 [1.51, 3.53] |
| Daugherty 2021 | 0.715483 | 0.094995 | 5.9% | 2.05 [1.70, 2.46] |
| Katsoularis 2023, 60+ y | 0.227813 | 0.25172 | 4.0% | 1.26 [0.77, 2.06] |
| Katsoularis 2023, 60 y & under | 0.348127 | 0.183357 | 4.9% | 1.42 [0.99, 2.03] |
| Lam 2023 | 0.385263 | 0.163468 | 5.1% | 1.47 [1.07, 2.03] |
| Lim 2023 | 0.690556 | 0.220029 | 4.4% | 1.99 [1.30, 3.07] |
| Mizrahi 2023, 19-40 y | 0.074487 | 0.201878 | 4.6% | 1.08 [0.73, 1.60] |
| Mizrahi 2023, 41-60 y | 0.044255 | 0.158501 | 5.2% | 1.05 [0.77, 1.43] |
| Mizrahi 2023, 60+ y | -0.13266 | 0.359911 | 2.9% | 0.88 [0.43, 1.77] |
| Valasquez Garcia 2024 | 0.329304 | 0.1379 | 5.4% | 1.39 [1.06, 1.82] |
| Zang 2023, INSIGHT cohort | 0.110557 | 0.063558 | 6.2% | 1.12 [0.99, 1.27] |
| **Subtotal (95% CI)** | | | **53.1%** | **1.41 [1.17, 1.71]** |
| Heterogeneity: Tau² = 0.07; Chi² = 43.03, df = 10 (P < 0.00001); I² = 77% | | | | |
| Test for overall effect: Z = 3.57 (P = 0.0004) | | | | |
| **2.2.2 Cardiovascular disorders, overall (outpatients/mixed, 18-64 y), 12+ mo follow-up** | | | | |
| Rezel-Potts 2022 | -0.05033 | 0.054413 | 6.2% | 0.95 [0.85, 1.06] |
| Wang 2022b, 20-44y | 0.527832 | 0.083008 | 6.0% | 1.70 [1.44, 1.99] |
| Wang 2022b, 45-64y | 0.721112 | 0.045725 | 6.3% | 2.06 [1.88, 2.25] |
| Xie 2022b | 0.51721819 | 0.043625 | 6.3% | 1.68 [1.54, 1.83] |
| Zappacosta 2022, 18-30 y | 0.350657 | 0.37779 | 2.8% | 1.42 [0.68, 2.98] |
| Zappacosta 2022, 31-40 y | 0.29267 | 0.254071 | 4.0% | 1.34 [0.81, 2.20] |
| Zappacosta 2022, 41-50 y | -0.57982 | 0.168459 | 5.1% | 0.56 [0.40, 0.78] |
| Zappacosta 2022, 51-60 y | -0.08338 | 0.131688 | 5.5% | 0.92 [0.71, 1.19] |
| Zappacosta 2022, 61-70 y | -0.0202 | 0.186986 | 4.8% | 0.98 [0.68, 1.41] |
| **Subtotal (95% CI)** | | | **46.9%** | **1.21 [0.93, 1.58]** |
| Heterogeneity: Tau² = 0.14; Chi² = 179.94, df = 8 (P < 0.00001); I² = 96% | | | | |
| Test for overall effect: Z = 1.43 (P = 0.15) | | | | |
| **Total (95% CI)** | | | **100.0%** | **1.32 [1.12, 1.55]** |
| Heterogeneity: Tau² = 0.11; Chi² = 227.01, df = 19 (P < 0.00001); I² = 92% | | | | |
| Test for overall effect: Z = 3.30 (P = 0.0010) | | | | |
| Test for subgroup differences: Chi² = 0.83, df = 1 (P = 0.36), I² = 0% | | | | |

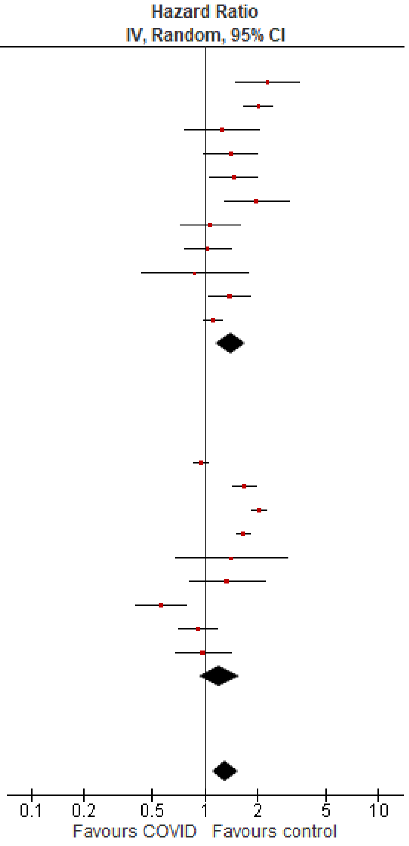

Outpatients/mixed, ≥65 y

| Study or Subgroup | log[Hazard Ratio] | SE | Weight | Hazard Ratio IV, Random, 95% CI |
|---|---|---|---|---|
| **2.3.1 Cardiovascular disorders, overall (outpatients/mixed, ≥65 y), <12 mo follow-up** | | | | |
| Batistoni 2024 | 0.43628 | 0.1502 | 7.8% | 1.55 [1.15, 2.08] |
| Cohen 2022 | 0.61252 | 0.0677 | 10.1% | 1.85 [1.62, 2.11] |
| Katsoularis 2023, 60+ y | 0.22781 | 0.25172 | 5.1% | 1.26 [0.77, 2.06] |
| Mizrahi 2023, 60+ y | -0.1327 | 0.15708 | 7.6% | 0.88 [0.64, 1.19] |
| Wan 2023a, UK cohort | 0.34905 | 0.09979 | 9.3% | 1.42 [1.17, 1.72] |
| Zang 2023, INSIGHT cohort | 0.2389 | 0.0423 | 10.6% | 1.27 [1.17, 1.38] |
| **Subtotal (95% CI)** | | | **50.4%** | **1.36 [1.12, 1.66]** |
| Heterogeneity: Tau² = 0.04; Chi² = 31.42, df = 5 (P < 0.00001); I² = 84% | | | | |
| Test for overall effect: Z = 3.10 (P = 0.002) | | | | |
| **2.3.2 Cardiovascular disorders, overall (outpatients/mixed, ≥65 y), 12+ mo follow-up** | | | | |
| Wang 2022b | 0.715354 | 0.043015 | 10.6% | 2.04 [1.88, 2.22] |
| Xie 2022b | 0.494319 | 0.027525 | 10.8% | 1.64 [1.55, 1.73] |
| Yang 2024 | 0.21486 | 0.01727 | 10.9% | 1.24 [1.20, 1.28] |
| Zappacosta 2022, 61-70 y | -0.0202 | 0.15267 | 7.7% | 0.98 [0.73, 1.32] |
| Zappacosta 2022, 70+ y | -0.2485 | 0.08638 | 9.6% | 0.78 [0.66, 0.92] |
| **Subtotal (95% CI)** | | | **49.6%** | **1.29 [1.01, 1.64]** |
| Heterogeneity: Tau² = 0.07; Chi² = 209.93, df = 4 (P < 0.00001); I² = 98% | | | | |
| Test for overall effect: Z = 2.03 (P = 0.04) | | | | |
| **Total (95% CI)** | | | **100.0%** | **1.33 [1.14, 1.54]** |
| Heterogeneity: Tau² = 0.05; Chi² = 241.39, df = 10 (P < 0.00001); I² = 96% | | | | |
| Test for overall effect: Z = 3.64 (P = 0.0003) | | | | |
| Test for subgroup differences: Chi² = 0.14, df = 1 (P = 0.71), I² = 0% | | | | |

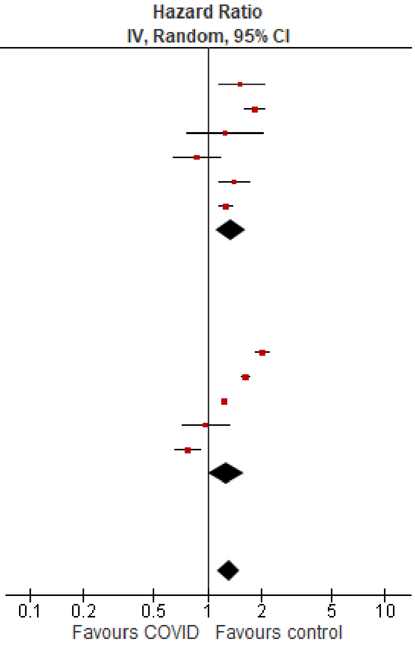

*Cardiovascular disorders, acute coronary disease*

Outpatients/mixed, <18 y

| Study or Subgroup | log[Hazard Ratio] | SE | Weight | Hazard Ratio IV, Random, 95% CI |
|---|---|---|---|---|
| **2.5.1 Acute coronary disease (outpatients/mixed, <18 y), <12 mo follow-up** | | | | |
| Gordon 2024, 0-1 y | 0.287682 | 0.381718 | 8.2% | 1.33 [0.63, 2.82] |
| Gordon 2024, 12-17 y | 0.267315 | 0.168705 | 42.2% | 1.31 [0.94, 1.82] |
| Gordon 2024, 2-4 y | 0.223143 | 0.387205 | 8.0% | 1.25 [0.59, 2.67] |
| Gordon 2024, 5-11 y | 0.476924 | 0.300027 | 13.4% | 1.61 [0.89, 2.90] |
| Mizrahi 2023, 12-18 y | 0.336472 | 0.614454 | 3.2% | 1.40 [0.42, 4.67] |
| **Subtotal (95% CI)** | | | **75.0%** | **1.36 [1.06, 1.74]** |
| Heterogeneity: Tau² = 0.00; Chi² = 0.43, df = 4 (P = 0.98); I² = 0% | | | | |
| Test for overall effect: Z = 2.41 (P = 0.02) | | | | |
| **2.5.2 Acute coronary disease (outpatients/mixed, <18 y), 12+ mo follow-up** | | | | |
| Roessler 2022 | 1.199965 | 1.05458 | 1.1% | 3.32 [0.42, 26.23] |
| Zhang 2024 | 0.530794 | 0.224256 | 23.9% | 1.70 [1.10, 2.64] |
| **Subtotal (95% CI)** | | | **25.0%** | **1.75 [1.14, 2.69]** |
| Heterogeneity: Tau² = 0.00; Chi² = 0.39, df = 1 (P = 0.53); I² = 0% | | | | |
| Test for overall effect: Z = 2.55 (P = 0.01) | | | | |
| **Total (95% CI)** | | | **100.0%** | **1.45 [1.17, 1.79]** |
| Heterogeneity: Tau² = 0.00; Chi² = 1.82, df = 6 (P = 0.94); I² = 0% | | | | |
| Test for overall effect: Z = 3.36 (P = 0.0008) | | | | |
| Test for subgroup differences: Chi² = 1.01, df = 1 (P = 0.31), I² = 1.1% | | | | |

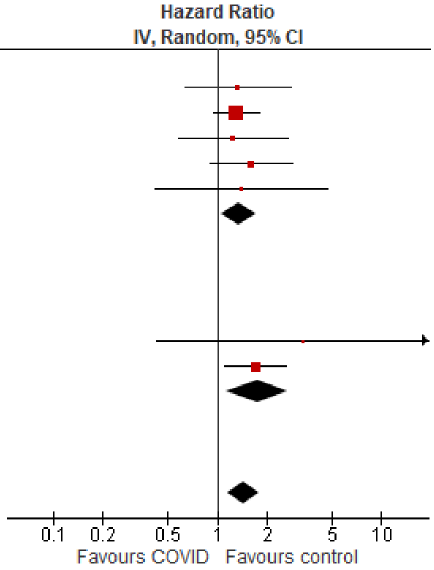

#### Outpatients/mixed, 18-64 y

| Study or Subgroup | log[Hazard Ratio] | SE | Weight | Hazard Ratio IV, Random, 95% CI |
|---|---|---|---|---|
| **2.6.1 Acute coronary disease (outpatients/mixed, 18-64 y), <12 mo follow-up** | | | | |
| Batistoni 2024 | 0.530628 | 0.281988 | 6.8% | 1.70 [0.98, 2.95] |
| Daugherty 2021 | 0.802002 | 0.156599 | 9.2% | 2.23 [1.64, 3.03] |
| Lam 2023 | 0.274072 | 0.188926 | 8.6% | 1.32 [0.91, 1.90] |
| Lim 2023 | 1.652497 | 0.377588 | 5.2% | 5.22 [2.49, 10.94] |
| Mizrahi 2023, 19-40 y | 0.250003 | 0.326955 | 6.0% | 1.28 [0.68, 2.44] |
| Mizrahi 2023, 41-60 y | -0.33604 | 0.147626 | 9.4% | 0.71 [0.54, 0.95] |
| Mizrahi 2023, 60+ y | -0.28462 | 0.384812 | 5.1% | 0.75 [0.35, 1.60] |
| Valasquez Garcia 2024 | 0.329304 | 0.1379 | 9.6% | 1.39 [1.06, 1.82] |
| **Subtotal (95% CI)** | | | **59.7%** | **1.46 [1.00, 2.12]** |
| Heterogeneity: Tau² = 0.23; Chi² = 44.73, df = 7 (P < 0.00001); I² = 84% | | | | |
| Test for overall effect: Z = 1.98 (P = 0.05) | | | | |
| **2.6.2 Acute coronary disease (outpatients/mixed, 18-64 y), 12+ mo follow-up** | | | | |
| Rezel-Potts 2022 | -0.17732 | 0.123467 | 9.8% | 0.84 [0.66, 1.07] |
| Wang 2022b, 20-44y | 0.372339 | 0.138893 | 9.5% | 1.45 [1.11, 1.91] |
| Wang 2022b, 45-64y | 0.662593 | 0.086399 | 10.4% | 1.94 [1.64, 2.30] |
| Xie 2022b | 0.506818 | 0.070291 | 10.6% | 1.66 [1.45, 1.91] |
| **Subtotal (95% CI)** | | | **40.3%** | **1.42 [1.03, 1.96]** |
| Heterogeneity: Tau² = 0.10; Chi² = 32.78, df = 3 (P < 0.00001); I² = 91% | | | | |
| Test for overall effect: Z = 2.14 (P = 0.03) | | | | |
| **Total (95% CI)** | | | **100.0%** | **1.43 [1.14, 1.81]** |
| Heterogeneity: Tau² = 0.13; Chi² = 80.19, df = 11 (P < 0.00001); I² = 86% | | | | |
| Test for overall effect: Z = 3.04 (P = 0.002) | | | | |
| Test for subgroup differences: Chi² = 0.01, df = 1 (P = 0.91), I² = 0% | | | | |

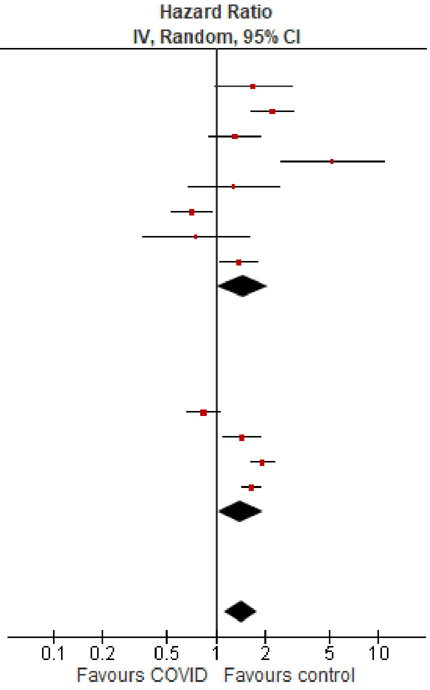

#### Outpatients/mixed, ≥65 y

| Study or Subgroup | log[Hazard Ratio] | SE | Weight | Hazard Ratio IV, Random, 95% CI |
|---|---|---|---|---|
| **2.7.1 Acute coronary disease (outpatients/mixed, ≥65 y), <12 mo follow-up** | | | | |
| Batistoni 2024 | 1.1474 | 0.29663 | 7.0% | 3.15 [1.76, 5.63] |
| Cohen 2022 | 0.732368 | 0.088939 | 15.8% | 2.08 [1.75, 2.48] |
| Mizrahi 2023, 60+ y | -0.2846 | 0.16795 | 12.0% | 0.75 [0.54, 1.05] |
| Wan 2023a, UK cohort | 0.48734 | 0.12869 | 13.9% | 1.63 [1.27, 2.10] |
| **Subtotal (95% CI)** | | | **48.7%** | **1.64 [1.01, 2.65]** |
| Heterogeneity: Tau² = 0.21; Chi² = 33.14, df = 3 (P < 0.00001); I² = 91% | | | | |
| Test for overall effect: Z = 2.01 (P = 0.04) | | | | |
| **2.7.2 Acute coronary disease (outpatients/mixed, ≥65 y), 12+ mo follow-up** | | | | |
| Wang 2022b | 0.589343 | 0.084561 | 16.0% | 1.80 [1.53, 2.13] |
| Xie 2022b | 0.470004 | 0.031731 | 17.7% | 1.60 [1.50, 1.70] |
| Yang 2024 | 0.18462 | 0.02869 | 17.7% | 1.20 [1.14, 1.27] |
| **Subtotal (95% CI)** | | | **51.3%** | **1.50 [1.19, 1.90]** |
| Heterogeneity: Tau² = 0.04; Chi² = 54.55, df = 2 (P < 0.00001); I² = 96% | | | | |
| Test for overall effect: Z = 3.38 (P = 0.0007) | | | | |
| **Total (95% CI)** | | | **100.0%** | **1.55 [1.27, 1.89]** |
| Heterogeneity: Tau² = 0.06; Chi² = 96.66, df = 6 (P < 0.00001); I² = 94% | | | | |
| Test for overall effect: Z = 4.37 (P < 0.0001) | | | | |
| Test for subgroup differences: Chi² = 0.10, df = 1 (P = 0.75), I² = 0% | | | | |

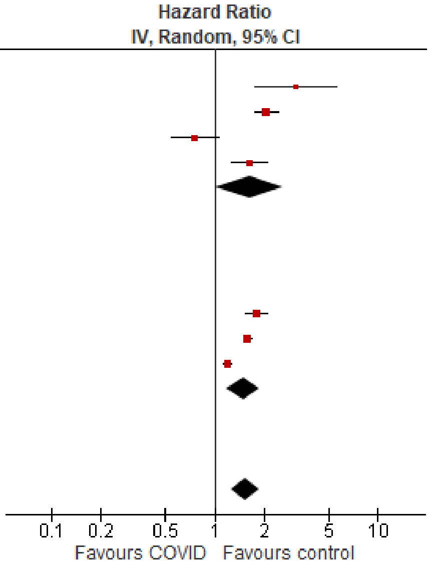

#### *Cardiovascular disorders, arrhythmias/dysrhythmias*

##### Outpatients/mixed, <18 y

| Study or Subgroup | log[Hazard Ratio] | SE | Weight | Hazard Ratio IV, Random, 95% CI |
|---|---|---|---|---|
| **2.8.1 Arrhythmia/dysrhythmia (outpatients/mixed, <18 y), <12 mo follow-up** | | | | |
| Gordon 2024, 0-1 y | 0.057432 | 0.097299 | 11.8% | 1.06 [0.88, 1.28] |
| Gordon 2024, 12-17 y | 0.207361 | 0.039932 | 19.3% | 1.23 [1.14, 1.33] |
| Gordon 2024, 2-4 y | 0.518563 | 0.124121 | 9.1% | 1.68 [1.32, 2.14] |
| Gordon 2024, 5-11 y | 0.310877 | 0.078909 | 14.0% | 1.36 [1.17, 1.59] |
| Mizrahi 2023, 12-18 y | -0.58949 | 0.340836 | 1.9% | 0.55 [0.28, 1.08] |
| Tartof 2022 | 0.576613 | 0.263687 | 3.0% | 1.78 [1.06, 2.98] |
| Valdivieso-Martinez 2023, unvaccinated cohort | 1.098612 | 1.154739 | 0.2% | 3.00 [0.31, 28.84] |
| Valdivieso-Martinez 2023, vaccinated cohort | 1.94591 | 3.75576 | 0.0% | 7.00 [0.00, 11015.56] |
| **Subtotal (95% CI)** | | | **59.2%** | **1.28 [1.10, 1.49]** |
| Heterogeneity: Tau² = 0.02; Chi² = 18.20, df = 7 (P = 0.01); I² = 62% | | | | |
| Test for overall effect: Z = 3.15 (P = 0.002) | | | | |
| **2.8.2 Arrhythmia/dysrhythmia (outpatients/mixed, <18 y), 12+ mo follow-up** | | | | |
| Kompaniyets 2022 | 0.14842 | 0.0176 | 21.5% | 1.16 [1.12, 1.20] |
| Roessler 2022 | 0.360835 | 0.161986 | 6.4% | 1.43 [1.04, 1.97] |
| Zhang 2024 | 0.385262 | 0.087884 | 12.9% | 1.47 [1.24, 1.75] |
| **Subtotal (95% CI)** | | | **40.8%** | **1.31 [1.08, 1.59]** |
| Heterogeneity: Tau² = 0.02; Chi² = 8.54, df = 2 (P = 0.01); I² = 77% | | | | |
| Test for overall effect: Z = 2.79 (P = 0.005) | | | | |
| **Total (95% CI)** | | | **100.0%** | **1.28 [1.17, 1.41]** |
| Heterogeneity: Tau² = 0.01; Chi² = 30.28, df = 10 (P = 0.0008); I² = 67% | | | | |
| Test for overall effect: Z = 5.14 (P < 0.00001) | | | | |
| Test for subgroup differences: Chi² = 0.04, df = 1 (P = 0.85), I² = 0% | | | | |

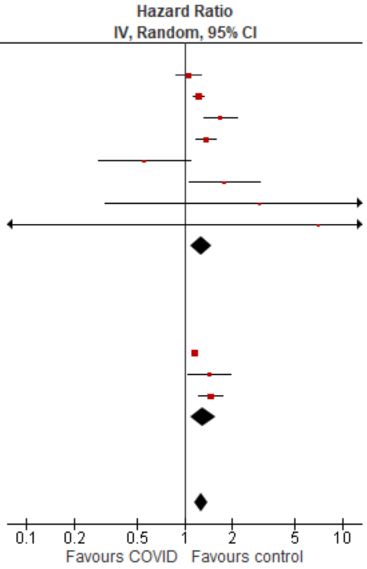

##### Outpatients/mixed, 18-64 y

| Study or Subgroup | log[Hazard Ratio] | SE | Weight | Hazard Ratio IV, Random, 95% CI |
|---|---|---|---|---|
| **2.9.1 Arrhythmia/dysrhythmia (outpatients/mixed, 18-64 y), <12 mo follow-up** | | | | |
| Batistoni 2024 | 0.916291 | 0.166762 | 7.2% | 2.50 [1.80, 3.47] |
| Daugherty 2021 | 0.916291 | 0.061931 | 12.5% | 2.50 [2.21, 2.82] |
| Katsoularis 2023, 60+ y | 0.227813 | 0.25172 | 4.4% | 1.26 [0.77, 2.06] |
| Katsoularis 2023, 60 y & under | 0.348127 | 0.183357 | 6.5% | 1.42 [0.99, 2.03] |
| Lam 2023 | 0.285179 | 0.217401 | 5.4% | 1.33 [0.87, 2.04] |
| Mizrahi 2023, 19-40 y | -0.07429 | 0.223295 | 5.2% | 0.93 [0.60, 1.44] |
| Mizrahi 2023, 41-60 y | 0.140799 | 0.174525 | 6.9% | 1.15 [0.82, 1.62] |
| Mizrahi 2023, 60+ y | 0.007484 | 0.410256 | 2.1% | 1.01 [0.45, 2.25] |
| **Subtotal (95% CI)** | | | **50.2%** | **1.47 [1.06, 2.03]** |
| Heterogeneity: Tau² = 0.17; Chi² = 49.86, df = 7 (P < 0.00001); I² = 86% | | | | |
| Test for overall effect: Z = 2.34 (P = 0.02) | | | | |
| **2.9.2 Arrhythmia/dysrhythmia (outpatients/mixed, 18-64 y), 12+ mo follow-up** | | | | |
| Rezel-Potts 2022 | 0.001457 | 0.129349 | 9.0% | 1.00 [0.78, 1.29] |
| Wang 2022b, 20-44y | 0.445361 | 0.038156 | 13.4% | 1.56 [1.45, 1.68] |
| Wang 2022b, 45-64y | 0.577963 | 0.030978 | 13.7% | 1.78 [1.68, 1.89] |
| Xie 2022b | 0.518794 | 0.027359 | 13.8% | 1.68 [1.59, 1.77] |
| **Subtotal (95% CI)** | | | **49.8%** | **1.57 [1.41, 1.76]** |
| Heterogeneity: Tau² = 0.01; Chi² = 23.22, df = 3 (P < 0.0001); I² = 87% | | | | |
| Test for overall effect: Z = 8.02 (P < 0.00001) | | | | |
| **Total (95% CI)** | | | **100.0%** | **1.56 [1.38, 1.77]** |
| Heterogeneity: Tau² = 0.03; Chi² = 85.99, df = 11 (P < 0.00001); I² = 87% | | | | |
| Test for overall effect: Z = 6.95 (P < 0.00001) | | | | |
| Test for subgroup differences: Chi² = 0.15, df = 1 (P = 0.69), I² = 0% | | | | |

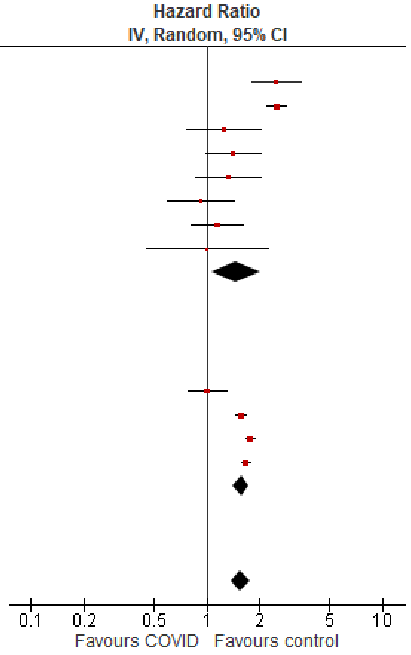

#### Outpatients/mixed, ≥65 y

| Study or Subgroup | log[Hazard Ratio] | SE | Weight | Hazard Ratio IV, Random, 95% CI |
|---|---|---|---|---|
| **2.10.1 Arrhythmia/dysrhythmia (outpatients/mixed, ≥65 y), <12 mo follow-up** | | | | |
| Batistoni 2024 | 0.3001 | 0.08647 | 13.3% | 1.35 [1.14, 1.60] |
| Cohen 2022 | 0.652325 | 0.056924 | 14.1% | 1.92 [1.72, 2.15] |
| Katsoularis 2023, 60+ y | 0.22781 | 0.25172 | 7.4% | 1.26 [0.77, 2.06] |
| Mizrahi 2023, 60+ y | 0.00748 | 0.17905 | 9.9% | 1.01 [0.71, 1.43] |
| Wan 2023a, UK cohort | 0.04038 | 0.14434 | 11.2% | 1.04 [0.78, 1.38] |
| **Subtotal (95% CI)** | | | **55.9%** | **1.31 [0.99, 1.74]** |
| Heterogeneity: Tau² = 0.08; Chi² = 30.52, df = 4 (P < 0.00001); I² = 87% | | | | |
| Test for overall effect: Z = 1.87 (P = 0.06) | | | | |
| **2.10.2 Arrhythmia/dysrhythmia (outpatients/mixed, ≥65 y), 12+ mo follow-up** | | | | |
| Wang 2022b | 0.665519 | 0.03084304 | 14.6% | 1.95 [1.83, 2.07] |
| Xie 2022b | 0.518794 | 0.031834 | 14.6% | 1.68 [1.58, 1.79] |
| Yang 2024 | 0.21486 | 0.01365 | 14.8% | 1.24 [1.21, 1.27] |
| **Subtotal (95% CI)** | | | **44.1%** | **1.59 [1.18, 2.15]** |
| Heterogeneity: Tau² = 0.07; Chi² = 223.82, df = 2 (P < 0.00001); I² = 99% | | | | |
| Test for overall effect: Z = 3.06 (P = 0.002) | | | | |
| **Total (95% CI)** | | | **100.0%** | **1.43 [1.18, 1.73]** |
| Heterogeneity: Tau² = 0.06; Chi² = 264.27, df = 7 (P < 0.00001); I² = 97% | | | | |
| Test for overall effect: Z = 3.69 (P = 0.0002) | | | | |
| Test for subgroup differences: Chi² = 0.87, df = 1 (P = 0.35), I² = 0% | | | | |

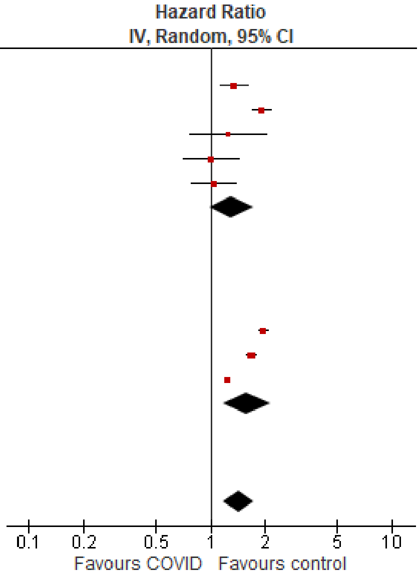

### *Cardiovascular disease, heart failure*

#### Outpatients/mixed, 18-64 y

| Study or Subgroup | log[Hazard Ratio] | SE | Weight | Hazard Ratio IV, Random, 95% CI |
|---|---|---|---|---|
| **2.14.1 Heart failure (outpatients/mixed, 18-64 y), <12 mo follow-up** | | | | |
| Batistoni 2024 | 0.947789 | 0.339347 | 7.6% | 2.58 [1.33, 5.02] |
| Daugherty 2021 | 0.963174 | 0.160982 | 12.4% | 2.62 [1.91, 3.59] |
| Lam 2023 | 0.582216 | 0.194005 | 11.4% | 1.79 [1.22, 2.62] |
| Lim 2023 | 0.615186 | 0.105693 | 13.9% | 1.85 [1.50, 2.28] |
| Zang 2023, INSIGHT cohort | 0.110557 | 0.063558 | 14.7% | 1.12 [0.99, 1.27] |
| **Subtotal (95% CI)** | | | **60.0%** | **1.84 [1.26, 2.67]** |
| Heterogeneity: Tau² = 0.15; Chi² = 39.53, df = 4 (P < 0.00001); I² = 90% | | | | |
| Test for overall effect: Z = 3.18 (P = 0.001) | | | | |
| **2.14.2 Heart failure (outpatients/mixed, 18-64 y), 12+ mo follow-up** | | | | |
| Rezel-Potts 2022 | -0.04229 | 0.237164 | 10.2% | 0.96 [0.60, 1.53] |
| Wang 2022b, 20-44y | 0.756122 | 0.063915 | 14.7% | 2.13 [1.88, 2.41] |
| Wang 2022b, 45-64y | 0.836381 | 0.035453 | 15.1% | 2.31 [2.15, 2.47] |
| **Subtotal (95% CI)** | | | **40.0%** | **1.94 [1.54, 2.44]** |
| Heterogeneity: Tau² = 0.03; Chi² = 14.13, df = 2 (P = 0.0009); I² = 86% | | | | |
| Test for overall effect: Z = 5.67 (P < 0.00001) | | | | |
| **Total (95% CI)** | | | **100.0%** | **1.81 [1.40, 2.34]** |
| Heterogeneity: Tau² = 0.11; Chi² = 114.74, df = 7 (P < 0.00001); I² = 94% | | | | |
| Test for overall effect: Z = 4.50 (P < 0.00001) | | | | |
| Test for subgroup differences: Chi² = 0.06, df = 1 (P = 0.80), I² = 0% | | | | |

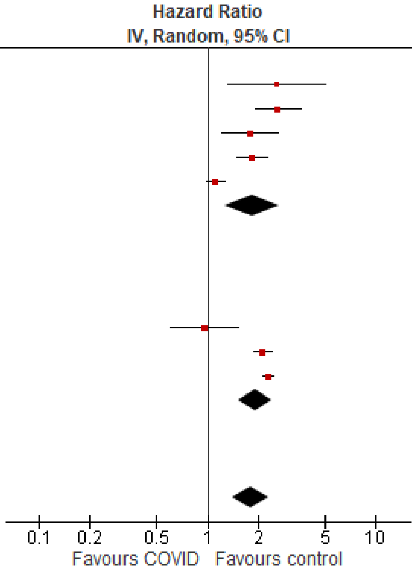

#### Outpatients/mixed, ≥65 y

| Study or Subgroup | log[Hazard Ratio] | SE | Weight | Hazard Ratio IV, Random, 95% CI |
|---|---|---|---|---|
| **2.15.1 Heart failure (outpatients/mixed, ≥65 y), <12 mo follow-up** | | | | |
| Batistoni 2024 | 0.67294 | 0.15286 | 14.7% | 1.96 [1.45, 2.64] |
| Cohen 2022 | 0.615186 | 0.0714 | 17.0% | 1.85 [1.61, 2.13] |
| Wan 2023a, UK cohort | 0.47 | 0.13678 | 15.3% | 1.60 [1.22, 2.09] |
| Zang 2023, INSIGHT cohort | 0.2389 | 0.0423 | 17.5% | 1.27 [1.17, 1.38] |
| **Subtotal (95% CI)** | | | **64.6%** | **1.62 [1.27, 2.08]** |
| Heterogeneity: Tau² = 0.05; Chi² = 25.80, df = 3 (P < 0.0001); I² = 88% | | | | |
| Test for overall effect: Z = 3.85 (P = 0.0001) | | | | |
| **2.15.2 Heart failure (outpatients/mixed, ≥65 y), 12+ mo follow-up** | | | | |
| Wang 2022b | 0.744315 | 0.030161 | 17.7% | 2.10 [1.98, 2.23] |
| Yang 2024 | 0.139 | 0.02173 | 17.8% | 1.15 [1.10, 1.20] |
| **Subtotal (95% CI)** | | | **35.4%** | **1.55 [0.86, 2.81]** |
| Heterogeneity: Tau² = 0.18; Chi² = 265.15, df = 1 (P < 0.00001); I² = 100% | | | | |
| Test for overall effect: Z = 1.46 (P = 0.14) | | | | |
| **Total (95% CI)** | | | **100.0%** | **1.61 [1.22, 2.12]** |
| Heterogeneity: Tau² = 0.11; Chi² = 291.13, df = 5 (P < 0.00001); I² = 98% | | | | |
| Test for overall effect: Z = 3.37 (P = 0.0007) | | | | |
| Test for subgroup differences: Chi² = 0.02, df = 1 (P = 0.89), I² = 0% | | | | |

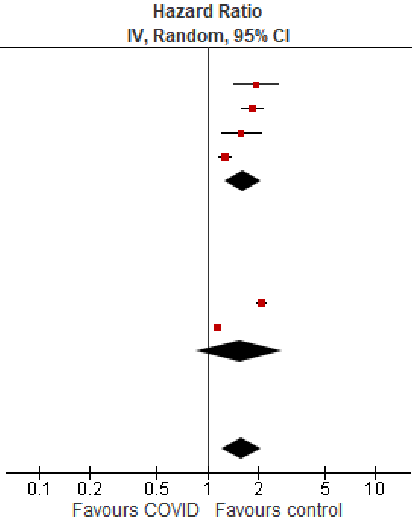

### *Diabetes, overall*

#### Outpatients/mixed, <18 y

| Study or Subgroup | log[Hazard Ratio] | SE | Weight | Hazard Ratio IV, Random, 95% CI |
|---|---|---|---|---|
| **3.1.1 Diabetes, overall (outpatients/mixed, <18 y), <12 mo follow-up** | | | | |
| Gordon 2024, 0-1 y | 0.920402 | 0.730235 | 1.0% | 2.51 [0.60, 10.50] |
| Gordon 2024, 12-17 y | 0.177811 | 0.101203 | 15.7% | 1.19 [0.98, 1.46] |
| Gordon 2024, 2-4 y | 0.828814 | 0.385061 | 3.3% | 2.29 [1.08, 4.87] |
| Gordon 2024, 5-11 y | 0.362418 | 0.159062 | 11.2% | 1.44 [1.05, 1.96] |
| Mizrahi 2023, 12-18 y | 0.431782 | 0.485066 | 2.2% | 1.54 [0.60, 3.98] |
| Tartof 2022 | -0.35667 | 0.215354 | 8.0% | 0.70 [0.46, 1.07] |
| **Subtotal (95% CI)** | | | **41.4%** | **1.26 [0.94, 1.70]** |
| Heterogeneity: Tau² = 0.06; Chi² = 11.63, df = 5 (P = 0.04); I² = 57% | | | | |
| Test for overall effect: Z = 1.53 (P = 0.12) | | | | |
| **3.1.2 Diabetes, overall (outpatients/mixed, <18 y), 12+ mo follow-up** | | | | |
| Kompaniyets 2022 | 0.171211 | 0.029592 | 20.9% | 1.19 [1.12, 1.26] |
| Noorzae 2023, 0-4 y | 0.086178 | 0.28434 | 5.4% | 1.09 [0.62, 1.90] |
| Noorzae 2023, 11-17 y | -0.09431 | 0.159087 | 11.2% | 0.91 [0.67, 1.24] |
| Noorzae 2023, 5-10 y | -0.27444 | 0.138961 | 12.6% | 0.76 [0.58, 1.00] |
| Pietropaolo 2022 | 0.188465 | 0.202801 | 8.6% | 1.21 [0.81, 1.80] |
| **Subtotal (95% CI)** | | | **58.6%** | **1.02 [0.82, 1.26]** |
| Heterogeneity: Tau² = 0.03; Chi² = 12.25, df = 4 (P = 0.02); I² = 67% | | | | |
| Test for overall effect: Z = 0.15 (P = 0.88) | | | | |
| **Total (95% CI)** | | | **100.0%** | **1.10 [0.95, 1.28]** |
| Heterogeneity: Tau² = 0.03; Chi² = 24.24, df = 10 (P = 0.007); I² = 59% | | | | |
| Test for overall effect: Z = 1.28 (P = 0.20) | | | | |
| Test for subgroup differences: Chi² = 1.35, df = 1 (P = 0.25), I² = 25.8% | | | | |

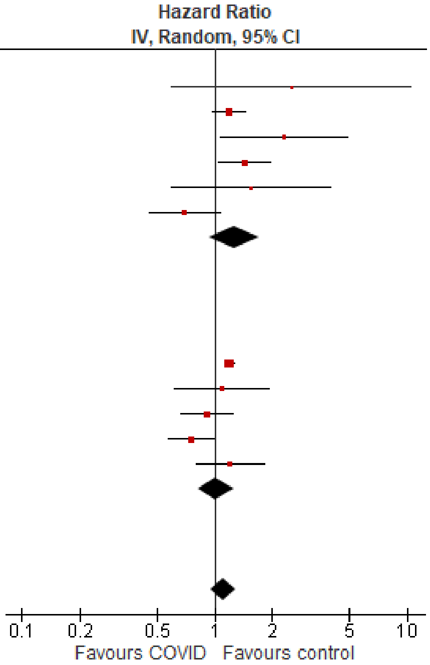

#### Outpatients/mixed, 18-64 y

| Study or Subgroup | log[Hazard Ratio] | SE | Weight | Hazard Ratio IV, Random, 95% CI |
|---|---|---|---|---|
| **3.2.1 Diabetes, overall (outpatients/mixed, 18-64 y), <12 mo follow-up** | | | | |
| Chang 2023, 18-40 y | 1.022451 | 0.056719 | 6.0% | 2.78 [2.49, 3.11] |
| Chang 2023, 41-64 y | 0.951658 | 0.050104 | 6.0% | 2.59 [2.35, 2.86] |
| Daugherty 2021 | 0.604316 | 0.127551 | 5.4% | 1.83 [1.43, 2.35] |
| Mizrahi 2023, 19-40 y | 0.301483 | 0.296006 | 3.5% | 1.35 [0.76, 2.41] |
| Mizrahi 2023, 41-60 y | 0.115372 | 0.125237 | 5.4% | 1.12 [0.88, 1.43] |
| Mizrahi 2023, 60+ y | 0.05005 | 0.372991 | 2.8% | 1.05 [0.51, 2.18] |
| Naveed 2023 | 0.157004 | 0.04811 | 6.0% | 1.17 [1.06, 1.29] |
| Taylor 2024, unvaccinated cohort, 40-59 y | 0.604316 | 0.153076 | 5.1% | 1.83 [1.36, 2.47] |
| Taylor 2024, unvaccinated cohort, 60-79 y | 0.883768 | 0.54832 | 1.7% | 2.42 [0.83, 7.09] |
| Taylor 2024, vaccinated cohort, 40-59 y | 0.173953 | 0.059796 | 5.9% | 1.19 [1.06, 1.34] |
| Taylor 2024, vaccinated cohort, 60-79 y | 0.223144 | 0.146779 | 5.2% | 1.25 [0.94, 1.67] |
| Zang 2023, INSIGHT cohort | 1.308333 | 0.041054 | 6.0% | 3.70 [3.41, 4.01] |
| **Subtotal (95% CI)** | | | **59.1%** | **1.71 [1.25, 2.35]** |
| Heterogeneity: Tau² = 0.27; Chi² = 515.56, df = 11 (P < 0.00001); I² = 98% | | | | |
| Test for overall effect: Z = 3.36 (P = 0.0008) | | | | |
| **3.2.2 Diabetes, overall (outpatients/mixed, 18-64 y), 12+ mo follow-up** | | | | |
| Choi 2023 | 0.431782 | 0.021478 | 6.1% | 1.54 [1.48, 1.61] |
| Pietropaolo 2022 | -0.07035 | 0.152822 | 5.1% | 0.93 [0.69, 1.26] |
| Rezel-Potts 2022 | 0.110593 | 0.048347 | 6.0% | 1.12 [1.02, 1.23] |
| Taylor 2024, pre-vaccination cohort, 18-39 y | 0.395044 | 0.075306 | 5.8% | 1.48 [1.28, 1.72] |
| Taylor 2024, pre-vaccination cohort, 40-59 y | 0.212603 | 0.035455 | 6.0% | 1.24 [1.15, 1.33] |
| Taylor 2024, pre-vaccination cohort, 60-79 y | 0.252617 | 0.087333 | 5.7% | 1.29 [1.08, 1.53] |
| Xie 2022a | 0.307485 | 0.018766 | 6.1% | 1.36 [1.31, 1.41] |
| **Subtotal (95% CI)** | | | **40.9%** | **1.30 [1.18, 1.43]** |
| Heterogeneity: Tau² = 0.01; Chi² = 63.52, df = 6 (P < 0.00001); I² = 91% | | | | |
| Test for overall effect: Z = 5.39 (P < 0.00001) | | | | |
| **Total (95% CI)** | | | **100.0%** | **1.53 [1.29, 1.81]** |
| Heterogeneity: Tau² = 0.12; Chi² = 878.07, df = 18 (P < 0.00001); I² = 98% | | | | |
| Test for overall effect: Z = 4.95 (P < 0.00001) | | | | |
| Test for subgroup differences: Chi² = 2.72, df = 1 (P = 0.10), I² = 63.3% | | | | |

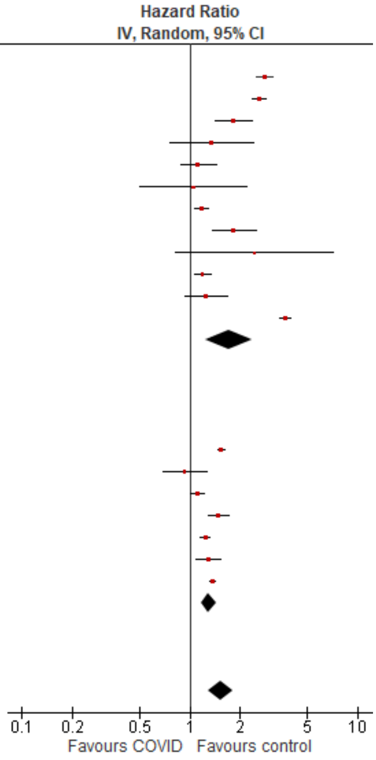

#### Outpatients/mixed, ≥65 y

| Study or Subgroup | log[Hazard Ratio] | SE | Weight | Hazard Ratio IV, Random, 95% CI |
|---|---|---|---|---|
| **3.3.1 Diabetes, overall (outpatients/mixed, ≥65 y), <12 mo follow-up** | | | | |
| Chang 2023 | 1.181727 | 0.070225 | 12.2% | 3.26 [2.84, 3.74] |
| Cohen 2022 | 0.672944 | 0.103435 | 11.2% | 1.96 [1.60, 2.40] |
| Mizrahi 2023, 60+ y | 0.05005 | 0.837214 | 1.0% | 1.05 [0.20, 5.42] |
| Taylor 2024, unvaccinated cohort, 60-79 y | 0.883768 | 0.316573 | 4.9% | 2.42 [1.30, 4.50] |
| Taylor 2024, vaccinated cohort, 60-79 y | 0.223144 | 0.084743 | 11.8% | 1.25 [1.06, 1.48] |
| Taylor 2024, vaccination cohort, 80-110 y | 0.350657 | 0.184439 | 8.4% | 1.42 [0.99, 2.04] |
| Zang 2023, INSIGHT cohort | 0.401926 | 0.037497 | 13.0% | 1.49 [1.39, 1.61] |
| **Subtotal (95% CI)** | | | **62.6%** | **1.80 [1.30, 2.51]** |
| Heterogeneity: Tau² = 0.16; Chi² = 115.27, df = 6 (P < 0.00001); I² = 95% | | | | |
| Test for overall effect: Z = 3.49 (P = 0.0005) | | | | |
| **3.3.2 Diabetes, overall (outpatients/mixed, ≥65 y), 12+ mo follow-up** | | | | |
| Taylor 2024, pre-vaccination cohort, 60-79 y | 0.252617 | 0.050422 | 12.7% | 1.29 [1.17, 1.42] |
| Taylor 2024, pre-vaccination cohort, 80-110 y | 0.240626 | 0.093384 | 11.6% | 1.27 [1.06, 1.53] |
| Xie 2022a | 0.357674 | 0.02142 | 13.2% | 1.43 [1.37, 1.49] |
| **Subtotal (95% CI)** | | | **37.4%** | **1.36 [1.24, 1.48]** |
| Heterogeneity: Tau² = 0.00; Chi² = 4.80, df = 2 (P = 0.09); I² = 58% | | | | |
| Test for overall effect: Z = 6.97 (P < 0.00001) | | | | |
| **Total (95% CI)** | | | **100.0%** | **1.61 [1.36, 1.92]** |
| Heterogeneity: Tau² = 0.06; Chi² = 153.17, df = 9 (P < 0.00001); I² = 94% | | | | |
| Test for overall effect: Z = 5.43 (P < 0.00001) | | | | |
| Test for subgroup differences: Chi² = 2.68, df = 1 (P = 0.10), I² = 62.6% | | | | |

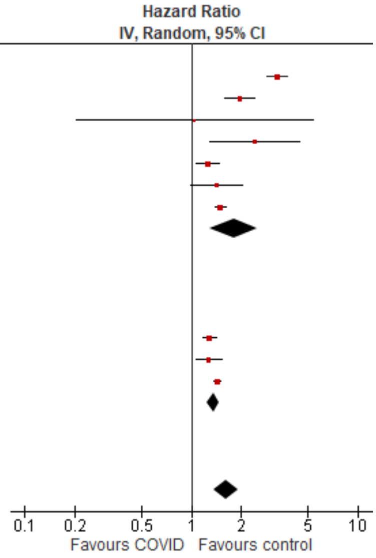

#### *Diabetes, Type 2*

##### Outpatients/mixed, 18-64 y

| Study or Subgroup | log[Hazard Ratio] | SE | Weight | Hazard Ratio IV, Random, 95% CI |
|---|---|---|---|---|
| **3.5.1 Type 2 (outpatients/mixed, 18-64 y), <12 mo follow-up** | | | | |
| Daugherty 2021 | 0.60431597 | 0.069371 | 13.4% | 1.83 [1.60, 2.10] |
| Taylor 2024, unvaccinated cohort, 40-59 y | 0.604316 | 0.153076 | 8.8% | 1.83 [1.36, 2.47] |
| Taylor 2024, unvaccinated cohort, 60-79 y | 0.883768 | 0.54832 | 1.5% | 2.42 [0.83, 7.09] |
| Taylor 2024, vaccinated cohort, 40-59 y | 0.173953 | 0.059796 | 13.9% | 1.19 [1.06, 1.34] |
| Taylor 2024, vaccinated cohort, 60-79 y | 0.223144 | 0.146779 | 9.1% | 1.25 [0.94, 1.67] |
| **Subtotal (95% CI)** | | | **46.7%** | **1.52 [1.18, 1.98]** |
| Heterogeneity: Tau² = 0.06; Chi² = 26.38, df = 4 (P < 0.0001); I² = 85% | | | | |
| Test for overall effect: Z = 3.18 (P = 0.001) | | | | |
| **3.5.2 Type 2 (outpatients/mixed, 18-64 y), 12+ mo follow-up** | | | | |
| Pietropaolo 2022 | -0.02224561 | 0.076557 | 13.0% | 0.98 [0.84, 1.14] |
| Taylor 2024, pre-vaccination cohort, 18-39 y | 0.395044 | 0.075306 | 13.1% | 1.48 [1.28, 1.72] |
| Taylor 2024, pre-vaccination cohort, 40-59 y | 0.212603 | 0.035455 | 14.9% | 1.24 [1.15, 1.33] |
| Taylor 2024, pre-vaccination cohort, 60-79 y | 0.252617 | 0.087333 | 12.4% | 1.29 [1.08, 1.53] |
| **Subtotal (95% CI)** | | | **53.3%** | **1.23 [1.07, 1.43]** |
| Heterogeneity: Tau² = 0.02; Chi² = 15.48, df = 3 (P = 0.001); I² = 81% | | | | |
| Test for overall effect: Z = 2.81 (P = 0.005) | | | | |
| **Total (95% CI)** | | | **100.0%** | **1.35 [1.18, 1.55]** |
| Heterogeneity: Tau² = 0.03; Chi² = 51.72, df = 8 (P < 0.00001); I² = 85% | | | | |
| Test for overall effect: Z = 4.37 (P < 0.0001) | | | | |
| Test for subgroup differences: Chi² = 1.95, df = 1 (P = 0.16), I² = 48.6% | | | | |

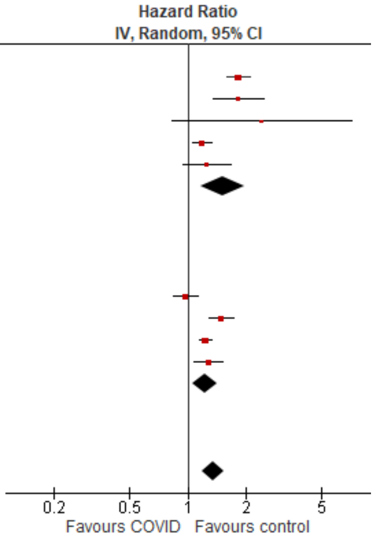

#### *Mental illness, overall*

##### Outpatients/mixed, <18 y

| Study or Subgroup | log[Hazard Ratio] | SE | Weight | Hazard Ratio IV, Random, 95% CI |
|---|---|---|---|---|
| **4.1.1 Mental illness, overall (outpatients/mixed, <18 y), <12 mo follow-up** | | | | |
| Gordon 2024, 0-1 y | -0.19237 | 0.234921 | 4.1% | 0.83 [0.52, 1.31] |
| Gordon 2024, 12-17 y | -0.11649 | 0.021231 | 7.3% | 0.89 [0.85, 0.93] |
| Gordon 2024, 2-4 y | 0.016261 | 0.080112 | 6.7% | 1.02 [0.87, 1.19] |
| Gordon 2024, 5-11 y | -0.03881 | 0.03265 | 7.2% | 0.96 [0.90, 1.03] |
| Mizrahi 2023, 12-18 y | -0.10932 | 0.200457 | 4.6% | 0.90 [0.61, 1.33] |
| Nielsen 2024 | -0.31471 | 0.027984 | 7.2% | 0.73 [0.69, 0.77] |
| Rao 2022 | 0.482426 | 0.059359 | 6.9% | 1.62 [1.44, 1.82] |
| Tartof 2022 | -0.1568 | 0.313754 | 3.0% | 0.85 [0.46, 1.58] |
| Valdivieso-Martinez 2023, unvaccinated cohort | -0.24484 | 0.597572 | 1.2% | 0.78 [0.24, 2.53] |
| Valdivieso-Martinez 2023, vaccinated cohort | -0.01384 | 0.427767 | 2.0% | 0.99 [0.43, 2.28] |
| **Subtotal (95% CI)** | | | **50.1%** | **0.97 [0.82, 1.14]** |
| Heterogeneity: Tau² = 0.04; Chi² = 160.59, df = 9 (P < 0.00001); I² = 94% | | | | |
| Test for overall effect: Z = 0.42 (P = 0.67) | | | | |
| **4.1.2 Mental illness, overall (outpatients/mixed, <18 y), 12+ mo follow-up** | | | | |
| Donnachie 2022, 0-11 y | 0.144966 | 0.037248 | 7.2% | 1.16 [1.07, 1.24] |
| Donnachie 2022, 12-17 y | -0.23067 | 0.023763 | 7.2% | 0.79 [0.76, 0.83] |
| Kompaniyets 2022 | -0.1726 | 0.010074 | 7.3% | 0.84 [0.83, 0.86] |
| Roessler 2022 | 0.360624 | 0.096104 | 6.4% | 1.43 [1.19, 1.73] |
| Taquet 2022 | -0.16626 | 0.039868 | 7.1% | 0.85 [0.78, 0.92] |
| Zhang-James 2024, 13-18 y | 0.04879 | 0.009719 | 7.3% | 1.05 [1.03, 1.07] |
| Zhang-James 2024, under 13 y | 0.451076 | 0.013002 | 7.3% | 1.57 [1.53, 1.61] |
| **Subtotal (95% CI)** | | | **49.9%** | **1.06 [0.86, 1.30]** |
| Heterogeneity: Tau² = 0.08; Chi² = 1621.20, df = 6 (P < 0.00001); I² = 100% | | | | |
| Test for overall effect: Z = 0.55 (P = 0.58) | | | | |
| **Total (95% CI)** | | | **100.0%** | **1.01 [0.88, 1.16]** |
| Heterogeneity: Tau² = 0.07; Chi² = 1879.94, df = 16 (P < 0.00001); I² = 99% | | | | |
| Test for overall effect: Z = 0.12 (P = 0.90) | | | | |
| Test for subgroup differences: Chi² = 0.48, df = 1 (P = 0.49), I² = 0% | | | | |

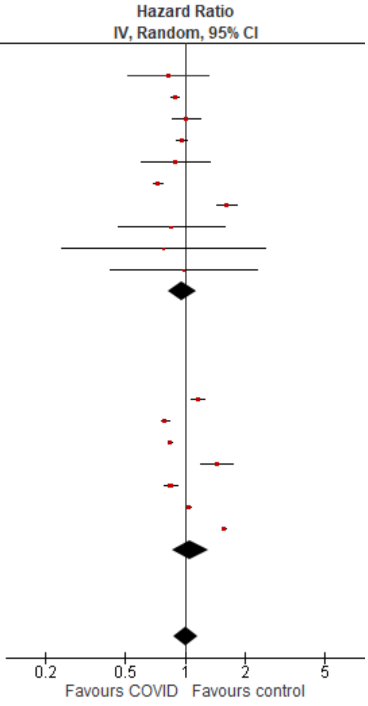

#### Outpatients/mixed, 18-64 y

| Study or Subgroup | log[Hazard Ratio] | SE | Weight | Hazard Ratio IV, Random, 95% CI |
|---|---|---|---|---|
| **4.2.1 Mental illness, overall (outpatients/mixed, 18-64 y), <12 mo follow-up** | | | | |
| Abel 2021, 16-24 y | 0.24686 | 0.109916 | 3.3% | 1.28 [1.03, 1.59] |
| Abel 2021, 25-34 y | 0.463734 | 0.120405 | 3.2% | 1.59 [1.26, 2.01] |
| Abel 2021, 35-49 y | 0.693147 | 0.102396 | 3.4% | 2.00 [1.64, 2.44] |
| Abel 2021, 50-59 y | 0.806476 | 0.127085 | 3.1% | 2.24 [1.75, 2.87] |
| Abel 2021, 60-69 y | 1.128171 | 0.236563 | 2.0% | 3.09 [1.94, 4.91] |
| Daugherty 2021 | 0.329304 | 0.036505 | 4.0% | 1.39 [1.29, 1.49] |
| Jacob 2022, 18-30 y | -0.01855 | 0.095097 | 3.5% | 0.98 [0.81, 1.18] |
| Jacob 2022, 31-40 y | -0.05372 | 0.107087 | 3.4% | 0.95 [0.77, 1.17] |
| Jacob 2022, 41-50 y | -0.00605 | 0.103521 | 3.4% | 0.99 [0.81, 1.22] |
| Jacob 2022, 51-60 y | -0.05027 | 0.086349 | 3.6% | 0.95 [0.80, 1.13] |
| Jacob 2022, 61-70 y | 0.122487 | 0.191274 | 2.4% | 1.13 [0.78, 1.64] |
| Lam 2023 | 0.523995 | 0.307968 | 1.5% | 1.69 [0.92, 3.09] |
| Mizrahi 2023, 19-40 y | 0.028888 | 0.134651 | 3.0% | 1.03 [0.79, 1.34] |
| Mizrahi 2023, 41-60 y | 0.091665 | 0.170862 | 2.6% | 1.10 [0.78, 1.53] |
| Mizrahi 2023, 60+ y | 0.062342 | 0.503706 | 0.7% | 1.06 [0.40, 2.86] |
| Nielsen 2024, 18-39 y | -0.27444 | 0.02015 | 4.1% | 0.76 [0.73, 0.79] |
| Nielsen 2024, 40-64 y | -0.34249 | 0.028774 | 4.0% | 0.71 [0.67, 0.75] |
| Park 2021, 20-39 y | 0.470004 | 0.083952 | 3.6% | 1.60 [1.36, 1.89] |
| Park 2021, 40-59 y | 0.81978 | 0.066238 | 3.8% | 2.27 [1.99, 2.58] |
| Zang 2023, INSIGHT cohort | 0.121421 | 0.033472 | 4.0% | 1.13 [1.06, 1.21] |
| **Subtotal (95% CI)** | | | **62.7%** | **1.28 [1.08, 1.52]** |
| Heterogeneity: Tau² = 0.14; Chi² = 726.23, df = 19 (P < 0.00001); I² = 97% | | | | |
| Test for overall effect: Z = 2.79 (P = 0.005) | | | | |
| **4.2.2 Mental illness, overall (outpatients/mixed, 18-64 y), 12+ mo follow-up** | | | | |
| Donnachie 2022, 18-39y | 0.325556 | 0.008473 | 4.1% | 1.38 [1.36, 1.41] |
| Donnachie 2022, 40-59y | -0.00321 | 0.009212 | 4.1% | 1.00 [0.98, 1.01] |
| Donnachie 2022, 60+ y | 0.094219 | 0.037287 | 4.0% | 1.10 [1.02, 1.18] |
| Smith 2024, 16-30 y | 0.157004 | 0.076811 | 3.7% | 1.17 [1.01, 1.36] |
| Smith 2024, 31-40 y | 0.254642 | 0.078027 | 3.7% | 1.29 [1.11, 1.50] |
| Smith 2024, 41-50 y | 0.231112 | 0.078648 | 3.7% | 1.26 [1.08, 1.47] |
| Smith 2024, 51-60 y | 0.182322 | 0.074001 | 3.7% | 1.20 [1.04, 1.39] |
| Smith 2024, 60+ y | 0.223144 | 0.196996 | 2.4% | 1.25 [0.85, 1.84] |
| Taquet 2022 | -0.01706 | 0.008532 | 4.1% | 0.98 [0.97, 1.00] |
| Xu 2022 | 0.425268 | 0.024948 | 4.0% | 1.53 [1.46, 1.61] |
| **Subtotal (95% CI)** | | | **37.3%** | **1.20 [1.06, 1.37]** |
| Heterogeneity: Tau² = 0.04; Chi² = 1181.81, df = 9 (P < 0.00001); I² = 99% | | | | |
| Test for overall effect: Z = 2.79 (P = 0.005) | | | | |
| **Total (95% CI)** | | | **100.0%** | **1.24 [1.13, 1.36]** |
| Heterogeneity: Tau² = 0.05; Chi² = 2064.61, df = 29 (P < 0.00001); I² = 99% | | | | |
| Test for overall effect: Z = 4.59 (P < 0.00001) | | | | |
| Test for subgroup differences: Chi² = 0.32, df = 1 (P = 0.57), I² = 0% | | | | |

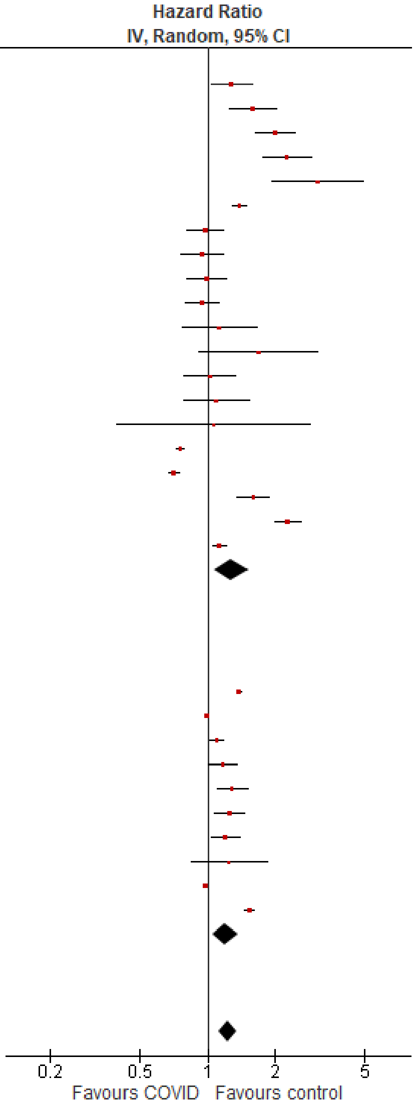

#### Outpatients/mixed, ≥65 y

| Study or Subgroup | log[Hazard Ratio] | SE | Weight | Hazard Ratio IV, Random, 95% CI |
|---|---|---|---|---|
| **4.3.1 Mental illness, overall (outpatients/mixed, ≥65 y), <12 mo follow-up** | | | | |
| Abel 2021, 60-69 y | 1.128171 | 0.259142 | 3.1% | 3.09 [1.86, 5.14] |
| Abel 2021, 70-79 y | 1.297463 | 0.307646 | 2.5% | 3.66 [2.00, 6.69] |
| Abel 2021, 80+ y | 1.427916 | 0.228145 | 3.6% | 4.17 [2.67, 6.52] |
| Cohen 2022 | 0.565314 | 0.056278 | 7.1% | 1.76 [1.58, 1.97] |
| Jacob 2022, 61-70 y | 0.122487 | 0.174608 | 4.6% | 1.13 [0.80, 1.59] |
| Jacob 2022, 70+ y | 0.084079 | 0.145272 | 5.2% | 1.09 [0.82, 1.45] |
| Mizrahi 2023, 60+ y | -0.31714 | 0.219835 | 3.7% | 0.73 [0.47, 1.12] |
| Murata 2024, alpha variant | -0.71335 | 0.689518 | 0.7% | 0.49 [0.13, 1.89] |
| Murata 2024, delta variant | 0.086178 | 0.339601 | 2.2% | 1.09 [0.56, 2.12] |
| Murata 2024, omicron BA.1/BA.2 variant | -0.52763 | 0.131419 | 5.5% | 0.59 [0.46, 0.76] |
| Murata 2024, omicron BA.5 variant | -0.79851 | 0.101126 | 6.2% | 0.45 [0.37, 0.55] |
| Nielsen 2024 | -0.30111 | 0.055374 | 7.2% | 0.74 [0.66, 0.82] |
| Park 2021, 60+ y | 0.620576 | 0.070584 | 6.9% | 1.86 [1.62, 2.14] |
| Wan 2023a, UK cohort | 0.53487 | 0.14634 | 5.2% | 1.71 [1.28, 2.27] |
| Zang 2023, INSIGHT cohort | 0.18938 | 0.052341 | 7.2% | 1.21 [1.09, 1.34] |
| **Subtotal (95% CI)** | | | **70.7%** | **1.27 [0.96, 1.68]** |
| Heterogeneity: Tau² = 0.26; Chi² = 353.08, df = 14 (P < 0.00001); I² = 96% | | | | |
| Test for overall effect: Z = 1.67 (P = 0.09) | | | | |
| **4.3.2 Mental illness, overall (outpatients/mixed, ≥65 y), 12+ mo follow-up** | | | | |
| Donnachie 2022, 60+ y | 0.094219 | 0.014441 | 7.6% | 1.10 [1.07, 1.13] |
| Smith 2024, 60+ y | 0.223144 | 0.085976 | 6.6% | 1.25 [1.06, 1.48] |
| Taquet 2022 | 0.045458 | 0.015112 | 7.6% | 1.05 [1.02, 1.08] |
| Xu 2022 | 0.270027 | 0.02524 | 7.5% | 1.31 [1.25, 1.38] |
| **Subtotal (95% CI)** | | | **29.3%** | **1.16 [1.05, 1.28]** |
| Heterogeneity: Tau² = 0.01; Chi² = 60.59, df = 3 (P < 0.00001); I² = 95% | | | | |
| Test for overall effect: Z = 2.97 (P = 0.003) | | | | |
| **Total (95% CI)** | | | **100.0%** | **1.20 [1.07, 1.35]** |
| Heterogeneity: Tau² = 0.05; Chi² = 418.18, df = 18 (P < 0.00001); I² = 96% | | | | |
| Test for overall effect: Z = 3.10 (P = 0.002) | | | | |
| Test for subgroup differences: Chi² = 0.35, df = 1 (P = 0.55), I² = 0% | | | | |

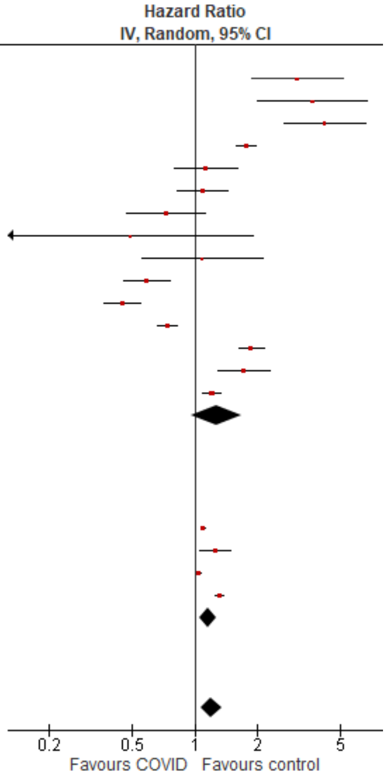

### *Mental illness, anxiety/anxiety disorders*

#### Outpatients/mixed, <18 y

| Study or Subgroup | log[Hazard Ratio] | SE | Weight | Hazard Ratio IV, Random, 95% CI |
|---|---|---|---|---|
| **4.6.1 Anxiety/anxiety disorders (outpatients/mixed, <18 y), <12 mo follow-up** | | | | |
| Mizrahi 2023, 0-4 y | -0.13926 | 0.451107 | 0.5% | 0.87 [0.36, 2.11] |
| Mizrahi 2023, 12-18 y | 0.020456 | 0.127184 | 5.8% | 1.02 [0.80, 1.31] |
| Mizrahi 2023, 5-11 y | 0.036664 | 0.151149 | 4.3% | 1.04 [0.77, 1.40] |
| Tartof 2022 | -0.12783 | 0.080929 | 11.7% | 0.88 [0.75, 1.03] |
| Valdivieso-Martinez 2023, unvaccinated cohort | -0.69315 | 0.499375 | 0.4% | 0.50 [0.19, 1.33] |
| Valdivieso-Martinez 2023, vaccinated cohort | 0.115113 | 0.21385 | 2.3% | 1.12 [0.74, 1.71] |
| **Subtotal (95% CI)** | | | **25.2%** | **0.94 [0.84, 1.06]** |
| Heterogeneity: Tau² = 0.00; Chi² = 3.82, df = 5 (P = 0.58); I² = 0% | | | | |
| Test for overall effect: Z = 1.03 (P = 0.30) | | | | |
| **4.6.2 Anxiety/anxiety disorders (outpatients/mixed, <18 y), 12+ mo follow-up** | | | | |
| Kompaniyets 2022 | -0.16252 | 0.006003 | 36.3% | 0.85 [0.84, 0.86] |
| Roessler 2022 | 0.409575 | 0.188676 | 2.9% | 1.51 [1.04, 2.18] |
| Taquet 2022 | -0.23383 | 0.009443 | 35.6% | 0.79 [0.78, 0.81] |
| **Subtotal (95% CI)** | | | **74.8%** | **0.84 [0.78, 0.91]** |
| Heterogeneity: Tau² = 0.00; Chi² = 50.47, df = 2 (P < 0.00001); I² = 96% | | | | |
| Test for overall effect: Z = 4.53 (P < 0.00001) | | | | |
| **Total (95% CI)** | | | **100.0%** | **0.87 [0.81, 0.92]** |
| Heterogeneity: Tau² = 0.00; Chi² = 58.55, df = 8 (P < 0.00001); I² = 86% | | | | |
| Test for overall effect: Z = 4.28 (P < 0.0001) | | | | |
| Test for subgroup differences: Chi² = 2.61, df = 1 (P = 0.11), I² = 61.7% | | | | |

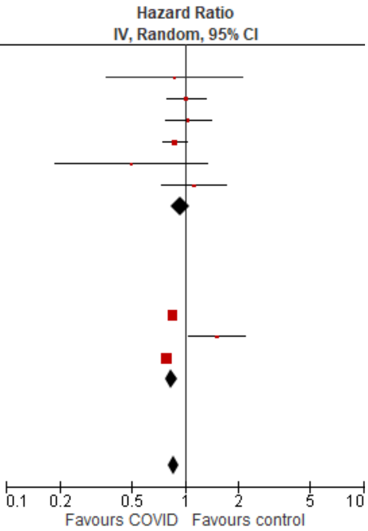

*Mental illness, depression & mood disorders*

<u>Outpatients/mixed, <18 y</u>

| Study or Subgroup | log[Hazard Ratio] | SE | Weight | Hazard Ratio IV, Random, 95% CI |
|---|---|---|---|---|
| **4.9.1 Depression & mood disorders (outpatients/mixed, <18 y), <12 mo follow-up** | | | | |
| Mizrahi 2023, 12-18 y | -0.44514 | 0.21327 | 13.1% | 0.64 [0.42, 0.97] |
| Valdivieso-Martinez 2023, unvaccinated cohort | 1.098612 | 0.864466 | 5.3% | 3.00 [0.55, 16.33] |
| Valdivieso-Martinez 2023, vaccinated cohort | -0.48548 | 0.448891 | 9.8% | 0.62 [0.26, 1.48] |
| **Subtotal (95% CI)** | | | **28.2%** | **0.75 [0.41, 1.34]** |
| Heterogeneity: Tau² = 0.10; Chi² = 3.07, df = 2 (P = 0.22); I² = 35% | | | | |
| Test for overall effect: Z = 0.98 (P = 0.33) | | | | |
| **4.9.2 Depression & mood disorders (outpatients/mixed, <18 y), 12+ mo follow-up** | | | | |
| Kompaniyets 2022 | -0.24846136 | 0.01646391 | 14.5% | 0.78 [0.76, 0.81] |
| Roessler 2022 | 0.371564 | 0.130768 | 13.9% | 1.45 [1.12, 1.87] |
| Taquet 2022 | -0.05933 | 0.012802 | 14.5% | 0.94 [0.92, 0.97] |
| Zhang-James 2024, 13-18 y | 1.435085 | 0.030047 | 14.5% | 4.20 [3.96, 4.45] |
| Zhang-James 2024, under 13 y | 1.029619 | 0.054875 | 14.4% | 2.80 [2.51, 3.12] |
| **Subtotal (95% CI)** | | | **71.8%** | **1.66 [0.93, 2.95]** |
| Heterogeneity: Tau² = 0.43; Chi² = 2850.19, df = 4 (P < 0.00001); I² = 100% | | | | |
| Test for overall effect: Z = 1.72 (P = 0.09) | | | | |
| **Total (95% CI)** | | | **100.0%** | **1.37 [0.84, 2.23]** |
| Heterogeneity: Tau² = 0.43; Chi² = 2858.70, df = 7 (P < 0.00001); I² = 100% | | | | |
| Test for overall effect: Z = 1.27 (P = 0.20) | | | | |
| Test for subgroup differences: Chi² = 3.63, df = 1 (P = 0.06), I² = 72.4% | | | | |

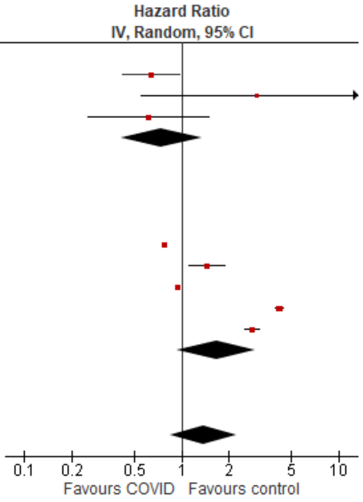

<u>Outpatients/mixed, 18-64 y</u>

| Study or Subgroup | log[Hazard Ratio] | SE | Weight | Hazard Ratio IV, Random, 95% CI |
|---|---|---|---|---|
| **4.10.1 Depression & mood disorders (outpatients/mixed, 18-64 y), <12 mo follow-up** | | | | |
| Daugherty 2021 | 0.239017 | 0.050181 | 10.1% | 1.27 [1.15, 1.40] |
| Jacob 2022, 18-30 y | 0.019803 | 0.087103 | 7.3% | 1.02 [0.86, 1.21] |
| Jacob 2022, 31-40 y | 0.019803 | 0.091285 | 7.0% | 1.02 [0.85, 1.22] |
| Jacob 2022, 41-50 y | -0.03046 | 0.086279 | 7.3% | 0.97 [0.82, 1.15] |
| Jacob 2022, 51-60 y | -0.06188 | 0.075739 | 8.1% | 0.94 [0.81, 1.09] |
| Jacob 2022, 61-70 y | 0.14842 | 0.162712 | 3.6% | 1.16 [0.84, 1.60] |
| Mizrahi 2023, 19-40 y | 0.062594 | 0.120899 | 5.2% | 1.06 [0.84, 1.35] |
| Mizrahi 2023, 41-60 y | 0.111062 | 0.1459 | 4.1% | 1.12 [0.84, 1.49] |
| Mizrahi 2023, 60+ y | 0.183801 | 0.592558 | 0.4% | 1.20 [0.38, 3.84] |
| **Subtotal (95% CI)** | | | **52.9%** | **1.07 [0.97, 1.17]** |
| Heterogeneity: Tau² = 0.01; Chi² = 16.26, df = 8 (P = 0.04); I² = 51% | | | | |
| Test for overall effect: Z = 1.33 (P = 0.19) | | | | |
| **4.10.2 Depression & mood disorders (outpatients/mixed, 18-64 y)** | | | | |
| Smith 2024, 16-30 y | 0.157004 | 0.076811 | 8.0% | 1.17 [1.01, 1.36] |
| Smith 2024, 31-40 y | 0.254642 | 0.078027 | 7.9% | 1.29 [1.11, 1.50] |
| Smith 2024, 41-50 y | 0.231112 | 0.078648 | 7.9% | 1.26 [1.08, 1.47] |
| Smith 2024, 51-60 y | 0.182322 | 0.074001 | 8.2% | 1.20 [1.04, 1.39] |
| Smith 2024, 60+ y | 0.223144 | 0.196996 | 2.7% | 1.25 [0.85, 1.84] |
| Taquet 2022 | 0.006976 | 0.004509 | 12.4% | 1.01 [1.00, 1.02] |
| **Subtotal (95% CI)** | | | **47.1%** | **1.17 [1.04, 1.33]** |
| Heterogeneity: Tau² = 0.02; Chi² = 28.44, df = 5 (P < 0.0001); I² = 82% | | | | |
| Test for overall effect: Z = 2.56 (P = 0.01) | | | | |
| **Total (95% CI)** | | | **100.0%** | **1.11 [1.04, 1.19]** |
| Heterogeneity: Tau² = 0.01; Chi² = 51.77, df = 14 (P < 0.00001); I² = 73% | | | | |
| Test for overall effect: Z = 2.92 (P = 0.003) | | | | |
| Test for subgroup differences: Chi² = 1.50, df = 1 (P = 0.22), I² = 33.4% | | | | |

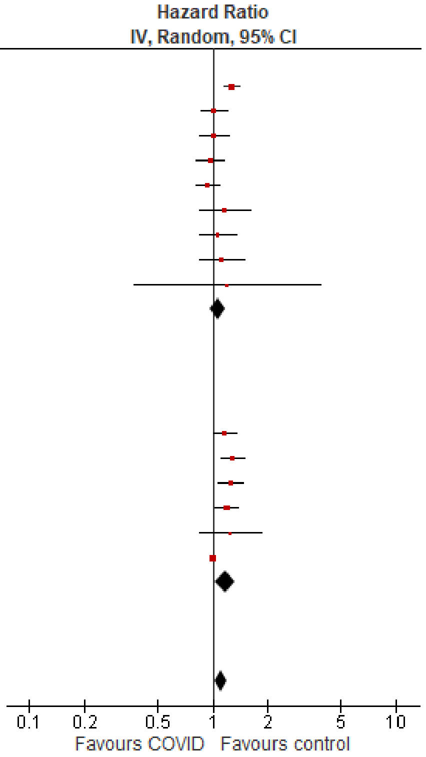

#### Outpatients/mixed, ≥65 y

| Study or Subgroup | log[Hazard Ratio] | SE | Weight | Hazard Ratio IV, Random, 95% CI |
|---|---|---|---|---|
| **4.11.1 Depression & mood disorders (outpatients/mixed, ≥65 y), <12 mo follow-up** | | | | |
| Jacob 2022, 61-70 y | 0.14842 | 0.148535 | 0.4% | 1.16 [0.87, 1.55] |
| Jacob 2022, 70+ y | 0.131028 | 0.119899 | 0.6% | 1.14 [0.90, 1.44] |
| Mizrahi 2023, 60+ y | 0.183801 | 0.258614 | 0.1% | 1.20 [0.72, 2.00] |
| Murata 2024, alpha variant | 0.81093 | 0.727984 | 0.0% | 2.25 [0.54, 9.37] |
| Murata 2024, delta variant | -0.19845 | 1.016738 | 0.0% | 0.82 [0.11, 6.02] |
| Murata 2024, omicron BA.1/BA.2 variant | 0.139762 | 0.263712 | 0.1% | 1.15 [0.69, 1.93] |
| Murata 2024, omicron BA.5 variant | -0.15082 | 0.22717 | 0.2% | 0.86 [0.55, 1.34] |
| **Subtotal (95% CI)** | | | **1.5%** | **1.12 [0.96, 1.30]** |
| Heterogeneity: Tau² = 0.00; Chi² = 2.53, df = 6 (P = 0.87); I² = 0% | | | | |
| Test for overall effect: Z = 1.47 (P = 0.14) | | | | |
| **4.11.2 Depression & mood disorders (outpatients/mixed, ≥65 y), 12+ mo follow-up** | | | | |
| Smith 2024, 60+ y | 0.223144 | 0.085976 | 1.2% | 1.25 [1.06, 1.48] |
| Taquet 2022 | 0.062881 | 0.009486 | 97.4% | 1.06 [1.05, 1.08] |
| **Subtotal (95% CI)** | | | **98.5%** | **1.13 [0.97, 1.31]** |
| Heterogeneity: Tau² = 0.01; Chi² = 3.43, df = 1 (P = 0.06); I² = 71% | | | | |
| Test for overall effect: Z = 1.57 (P = 0.12) | | | | |
| **Total (95% CI)** | | | **100.0%** | **1.07 [1.05, 1.09]** |
| Heterogeneity: Tau² = 0.00; Chi² = 6.36, df = 8 (P = 0.61); I² = 0% | | | | |
| Test for overall effect: Z = 7.00 (P < 0.00001) | | | | |
| Test for subgroup differences: Chi² = 0.00, df = 1 (P = 0.96), I² = 0% | | | | |

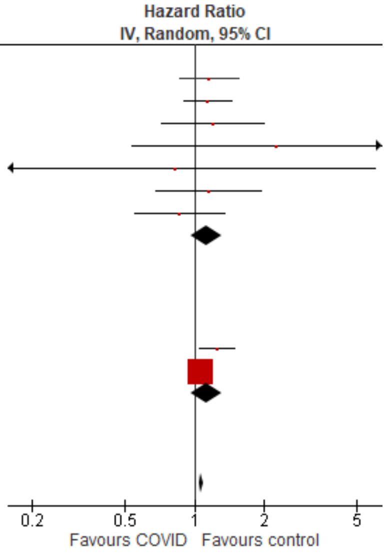

#### *Mental illness, psychosis/psychotic disorders*

#### Outpatients/mixed, <18 y

| Study or Subgroup | log[Hazard Ratio] | SE | Weight | Hazard Ratio IV, Random, 95% CI |
|---|---|---|---|---|
| **4.12.1 Psychosis/psychotic disorders (outpatients/mixed, <18 y), <12 mo follow-up** | | | | |
| Mizrahi 2023, 12-18 y | -0.2013 | 0.379172 | 18.3% | 0.82 [0.39, 1.72] |
| Tartof 2022 | -1.66073 | 0.583107 | 12.7% | 0.19 [0.06, 0.60] |
| **Subtotal (95% CI)** | | | **31.0%** | **0.42 [0.10, 1.75]** |
| Heterogeneity: Tau² = 0.82; Chi² = 4.40, df = 1 (P = 0.04); I² = 77% | | | | |
| Test for overall effect: Z = 1.19 (P = 0.23) | | | | |
| **4.12.2 Psychosis/psychotic disorders (outpatients/mixed, <18 y), 12+ mo follow-up** | | | | |
| Taquet 2022 | 1.048967 | 0.10743 | 25.9% | 2.85 [2.31, 3.52] |
| Zhang-James 2024, 13-18 y | 0.788457 | 0.118016 | 25.7% | 2.20 [1.75, 2.77] |
| Zhang-James 2024, under 13 y | -0.10536 | 0.410571 | 17.3% | 0.90 [0.40, 2.01] |
| **Subtotal (95% CI)** | | | **69.0%** | **2.14 [1.46, 3.14]** |
| Heterogeneity: Tau² = 0.08; Chi² = 8.81, df = 2 (P = 0.01); I² = 77% | | | | |
| Test for overall effect: Z = 3.89 (P = 0.0001) | | | | |
| **Total (95% CI)** | | | **100.0%** | **1.23 [0.70, 2.16]** |
| Heterogeneity: Tau² = 0.31; Chi² = 35.02, df = 4 (P < 0.00001); I² = 89% | | | | |
| Test for overall effect: Z = 0.72 (P = 0.47) | | | | |
| Test for subgroup differences: Chi² = 4.66, df = 1 (P = 0.03), I² = 78.5% | | | | |

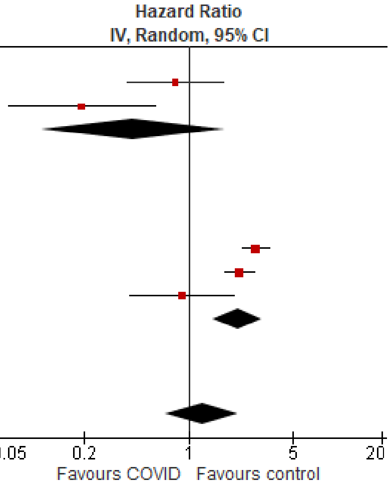

#### *Neurological disorders, overall*

##### Outpatients/mixed, <18 y

| Study or Subgroup | log[Hazard Ratio] | SE | Weight | Hazard Ratio IV, Random, 95% CI |
|---|---|---|---|---|
| **5.1.1 Neurological disorders, overall (outpatients/mixed, <18 y), <12 mo follow-up** | | | | |
| Donnachie 2022, 0-11 y | 1.029619 | 0.368361 | 4.2% | 2.80 [1.36, 5.76] |
| Donnachie 2022, 12-17 y | 0.897923 | 0.145685 | 10.5% | 2.45 [1.84, 3.27] |
| Gronkjaer 2023 | -0.11653 | 0.069693 | 13.5% | 0.89 [0.78, 1.02] |
| Mizrahi 2023, 0-4 y | 0.866777 | 0.464457 | 2.9% | 2.38 [0.96, 5.91] |
| Mizrahi 2023, 12-18 y | 0.312513 | 0.366435 | 4.2% | 1.37 [0.67, 2.80] |
| Mizrahi 2023, 5-11 y | -0.13955 | 0.325134 | 4.9% | 0.87 [0.46, 1.64] |
| Nielsen 2024 | 0.20678 | 0.32787 | 4.9% | 1.23 [0.65, 2.34] |
| Rao 2022 | 0.165514 | 0.05038 | 14.0% | 1.18 [1.07, 1.30] |
| Valdivieso-Martinez 2023, unvaccinated cohort | -0.28768 | 0.76335 | 1.2% | 0.75 [0.17, 3.35] |
| Valdivieso-Martinez 2023, vaccinated cohort | 0 | 0.40773 | 3.6% | 1.00 [0.45, 2.22] |
| **Subtotal (95% CI)** | | | **64.0%** | **1.35 [1.03, 1.77]** |
| Heterogeneity: Tau² = 0.10; Chi² = 50.59, df = 9 (P < 0.00001); I² = 82% | | | | |
| Test for overall effect: Z = 2.19 (P = 0.03) | | | | |
| **5.1.2 Neurological disorders, overall (outpatients/mixed, <18 y), 12+ mo follow-up** | | | | |
| Kompaniyets 2022 | -0.06188 | 0.008186 | 14.7% | 0.94 [0.93, 0.96] |
| Roessler 2022 | 0.195264 | 0.535629 | 2.3% | 1.22 [0.43, 3.47] |
| Taquet 2022 | 0.679838 | 0.201247 | 8.4% | 1.97 [1.33, 2.93] |
| Westman 2022 | -0.22314 | 0.143126 | 10.6% | 0.80 [0.60, 1.06] |
| **Subtotal (95% CI)** | | | **36.0%** | **1.10 [0.78, 1.54]** |
| Heterogeneity: Tau² = 0.08; Chi² = 15.08, df = 3 (P = 0.002); I² = 80% | | | | |
| Test for overall effect: Z = 0.55 (P = 0.58) | | | | |
| **Total (95% CI)** | | | **100.0%** | **1.24 [1.04, 1.47]** |
| Heterogeneity: Tau² = 0.05; Chi² = 92.67, df = 13 (P < 0.00001); I² = 86% | | | | |
| Test for overall effect: Z = 2.39 (P = 0.02) | | | | |
| Test for subgroup differences: Chi² = 0.88, df = 1 (P = 0.35), I² = 0% | | | | |

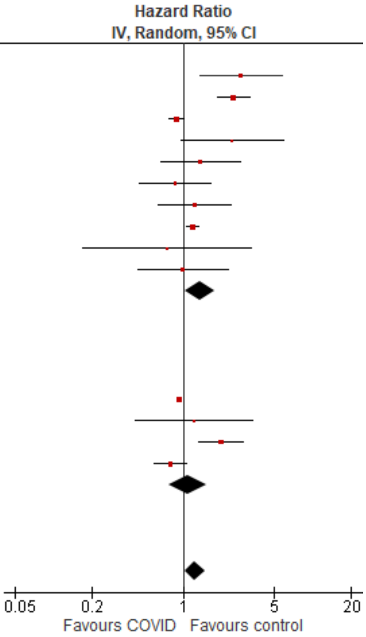

##### Outpatients/mixed, 18-64 y

| Study or Subgroup | log[Hazard Ratio] | SE | Weight | Hazard Ratio IV, Random, 95% CI |
|---|---|---|---|---|
| **5.2.1 Neurological disorders, overall (outpatients/mixed, 18-64 y), <12 mo follow-up** | | | | |
| Daugherty 2021 | 0.488405 | 0.235579 | 3.6% | 1.63 [1.03, 2.59] |
| Gronkjaer 2023, 20-29 y | 0.076961 | 0.059041 | 5.3% | 1.08 [0.96, 1.21] |
| Gronkjaer 2023, 30-39 y | 0.122218 | 0.058435 | 5.3% | 1.13 [1.01, 1.27] |
| Gronkjaer 2023, 40-49 y | 0.254642 | 0.049401 | 5.3% | 1.29 [1.17, 1.42] |
| Gronkjaer 2023, 50-59 y | 0.076961 | 0.04953 | 5.3% | 1.08 [0.98, 1.19] |
| Gronkjaer 2023, 60-69 y | 0.113329 | 0.084131 | 5.1% | 1.12 [0.95, 1.32] |
| Mizrahi 2023, 19-40 y | 0.017494 | 0.227602 | 3.7% | 1.02 [0.65, 1.59] |
| Mizrahi 2023, 41-60 y | 0.174803 | 0.313754 | 2.8% | 1.19 [0.64, 2.20] |
| Mizrahi 2023, 60+ y | -0.08154 | 0.76822 | 0.8% | 0.92 [0.20, 4.15] |
| Nielsen 2024, 18-39 y | 0.100848 | 0.170984 | 4.3% | 1.11 [0.79, 1.55] |
| Nielsen 2024, 40-64 y | 0.109452 | 0.080718 | 5.1% | 1.12 [0.95, 1.31] |
| Zang 2023, INSIGHT cohort | 0.320223 | 0.081779 | 5.1% | 1.38 [1.17, 1.62] |
| Zang 2023, OneFlorida+ cohort | -0.35667 | 0.1281 | 4.7% | 0.70 [0.54, 0.90] |
| **Subtotal (95% CI)** | | | **56.3%** | **1.13 [1.04, 1.23]** |
| Heterogeneity: Tau² = 0.01; Chi² = 30.93, df = 12 (P = 0.002); I² = 61% | | | | |
| Test for overall effect: Z = 2.85 (P = 0.004) | | | | |
| **5.2.2 Neurological disorders, overall (outpatients/mixed, 18-64 y), 12+ mo follow-up** | | | | |
| Donnachie 2022, 18-39y | 0.761549 | 0.077881 | 5.1% | 2.14 [1.84, 2.49] |
| Donnachie 2022, 40-59y | 1.134362 | 0.051255 | 5.3% | 3.11 [2.81, 3.44] |
| Donnachie 2022, 60+ y | 0.135997 | 0.126196 | 4.7% | 1.15 [0.89, 1.47] |
| Taquet 2022 | 0.029667 | 0.027151 | 5.4% | 1.03 [0.98, 1.09] |
| Westman 2022, 21-40 y | -0.52763 | 0.124706 | 4.7% | 0.59 [0.46, 0.75] |
| Westman 2022, 41-60 y | -0.43078 | 0.113063 | 4.8% | 0.65 [0.52, 0.81] |
| Westman 2022, 61-80y | 0.24686 | 0.187368 | 4.1% | 1.28 [0.89, 1.85] |
| Xu 2022 | 0.34359 | 0.023453 | 5.4% | 1.41 [1.35, 1.48] |
| Zarifkar 2022, 18-39 y | -0.3191 | 0.324026 | 2.7% | 0.73 [0.39, 1.37] |
| Zarifkar 2022, 40-59 y | -0.26227 | 0.914212 | 0.6% | 0.77 [0.13, 4.62] |
| Zarifkar 2022, 60-79 y | 0.760617 | 0.831084 | 0.7% | 2.14 [0.42, 10.91] |
| **Subtotal (95% CI)** | | | **43.7%** | **1.20 [0.90, 1.60]** |
| Heterogeneity: Tau² = 0.18; Chi² = 494.51, df = 10 (P < 0.00001); I² = 98% | | | | |
| Test for overall effect: Z = 1.25 (P = 0.21) | | | | |
| **Total (95% CI)** | | | **100.0%** | **1.16 [1.00, 1.35]** |
| Heterogeneity: Tau² = 0.11; Chi² = 559.38, df = 23 (P < 0.00001); I² = 96% | | | | |
| Test for overall effect: Z = 1.97 (P = 0.05) | | | | |
| Test for subgroup differences: Chi² = 0.17, df = 1 (P = 0.68), I² = 0% | | | | |

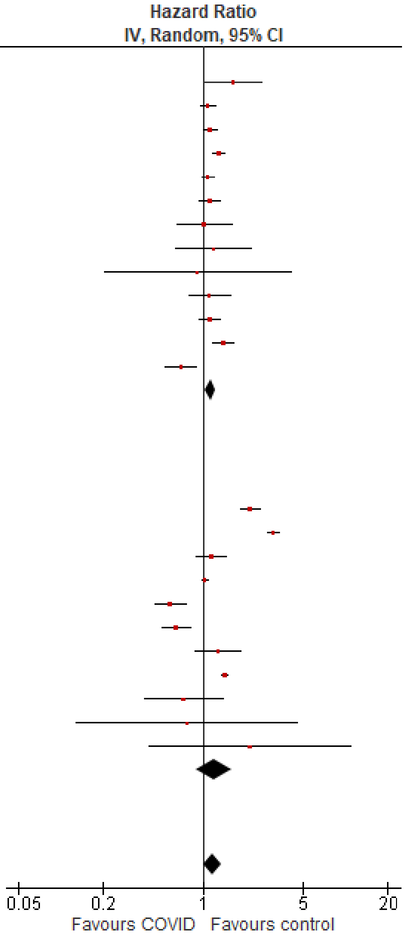

#### Outpatients/mixed, ≥65 y

| Study or Subgroup | log[Hazard Ratio] | SE | Weight | Hazard Ratio IV, Random, 95% CI |
|---|---|---|---|---|
| **5.3.1 Neurological disorders, overall (outpatients/mixed, ≥65 y), <12 mo follow-up** | | | | |
| Cohen 2022 | 0.793707 | 0.232087 | 4.6% | 2.21 [1.40, 3.49] |
| Gronkjaer 2023, 60-69 y | 0.113329 | 0.094061 | 5.2% | 1.12 [0.93, 1.35] |
| Gronkjaer 2023, 70-79 y | 0.131028 | 0.060436 | 5.3% | 1.14 [1.01, 1.28] |
| Gronkjaer 2023, 80+ y | 0.182322 | 0.070304 | 5.2% | 1.20 [1.05, 1.38] |
| Mizrahi 2023, 60+ y | 0.023683 | 0.405044 | 3.7% | 1.02 [0.46, 2.26] |
| Murata 2024, alpha variant | -0.31471 | 1.389474 | 0.9% | 0.73 [0.05, 11.12] |
| Murata 2024, delta variant | 1.811562 | 0.422699 | 3.6% | 6.12 [2.67, 14.01] |
| Murata 2024, omicron BA.1/BA.2 variant | 0.71295 | 0.200632 | 4.8% | 2.04 [1.38, 3.02] |
| Murata 2024, omicron BA.5 variant | 0.029559 | 0.183813 | 4.9% | 1.03 [0.72, 1.48] |
| Nielsen 2024 | 0.052315 | 0.148244 | 5.0% | 1.05 [0.79, 1.41] |
| Wan 2023a, UK cohort | 0.09531 | 0.240453 | 4.6% | 1.10 [0.69, 1.76] |
| Zang 2023, INSIGHT cohort | 0.320368 | 0.078479 | 5.2% | 1.38 [1.18, 1.61] |
| Zang 2023, OneFlorida+ cohort | 2.734368 | 0.062372 | 5.2% | 15.40 [13.63, 17.40] |
| **Subtotal (95% CI)** | | | **58.1%** | **1.73 [0.90, 3.34]** |
| Heterogeneity: Tau² = 1.35; Chi² = 1312.82, df = 12 (P < 0.00001); I² = 99% | | | | |
| Test for overall effect: Z = 1.63 (P = 0.10) | | | | |
| **5.3.2 Neurological disorders, overall (outpatients/mixed, ≥65 y), 12+ mo follow-up** | | | | |
| Donnachie 2022, 60+ y | 0.135997 | 0.049883 | 5.3% | 1.15 [1.04, 1.26] |
| Gollop 2023 | 0.029559 | 0.106419 | 5.1% | 1.03 [0.84, 1.27] |
| Taquet 2022 | 0.091728 | 0.045347 | 5.3% | 1.10 [1.00, 1.20] |
| Wang 2022a | 0.524729 | 0.029861 | 5.3% | 1.69 [1.59, 1.79] |
| Westman 2022, 100+ y | 0.004689 | 1.997988 | 0.5% | 1.00 [0.02, 50.43] |
| Westman 2022, 61-80y | 0.24686 | 0.118502 | 5.1% | 1.28 [1.01, 1.61] |
| Westman 2022, 81-100 y | 0.29267 | 0.163323 | 4.9% | 1.34 [0.97, 1.85] |
| Xu 2022 | 0.34359 | 0.023621 | 5.3% | 1.41 [1.35, 1.48] |
| Zarifkar 2022, 60-79 y | 0.779905 | 0.582952 | 2.8% | 2.18 [0.70, 6.84] |
| Zarifkar 2022, 80+ y | 0.267583 | 0.688153 | 2.3% | 1.31 [0.34, 5.03] |
| **Subtotal (95% CI)** | | | **41.9%** | **1.29 [1.12, 1.48]** |
| Heterogeneity: Tau² = 0.03; Chi² = 94.67, df = 9 (P < 0.00001); I² = 90% | | | | |
| Test for overall effect: Z = 3.49 (P = 0.0005) | | | | |
| **Total (95% CI)** | | | **100.0%** | **1.55 [1.18, 2.04]** |
| Heterogeneity: Tau² = 0.37; Chi² = 1576.28, df = 22 (P < 0.00001); I² = 99% | | | | |
| Test for overall effect: Z = 3.13 (P = 0.002) | | | | |
| Test for subgroup differences: Chi² = 0.75, df = 1 (P = 0.39), I² = 0% | | | | |

Hazard Ratio IV, Random, 95% CI

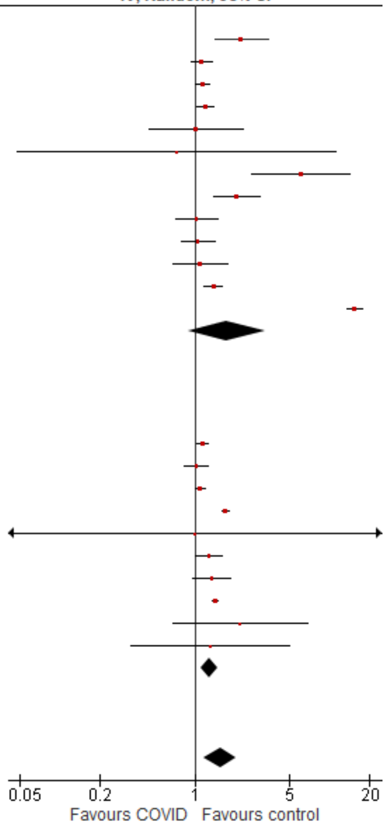

#### *Neurological disorders, communication & motor disorders*

#### Outpatients/mixed, <18 y

| Study or Subgroup | log[Hazard Ratio] | SE | Weight | Hazard Ratio IV, Random, 95% CI |
|---|---|---|---|---|
| **5.6.1 Communication & motor disorders (outpatients/mixed, <18 y), <12 mo follow-up** | | | | |
| Nielsen 2024 | 0.912283 | 0.334132 | 8.9% | 2.49 [1.29, 4.79] |
| Rao 2022 | 0.165514 | 0.05038 | 52.9% | 1.18 [1.07, 1.30] |
| **Subtotal (95% CI)** | | | **61.8%** | **1.59 [0.78, 3.27]** |
| Heterogeneity: Tau² = 0.22; Chi² = 4.88, df = 1 (P = 0.03); I² = 80% | | | | |
| Test for overall effect: Z = 1.27 (P = 0.20) | | | | |
| **5.6.2 Communication & motor disorders (outpatients/mixed, <18 y), 12+ mo follow-up** | | | | |
| Roessler 2022 | 0.211468 | 0.120209 | 34.4% | 1.24 [0.98, 1.56] |
| Taquet 2022 | 0.663519 | 0.537058 | 3.8% | 1.94 [0.68, 5.56] |
| **Subtotal (95% CI)** | | | **38.2%** | **1.26 [1.00, 1.59]** |
| Heterogeneity: Tau² = 0.00; Chi² = 0.67, df = 1 (P = 0.41); I² = 0% | | | | |
| Test for overall effect: Z = 1.99 (P = 0.05) | | | | |
| **Total (95% CI)** | | | **100.0%** | **1.31 [1.06, 1.61]** |
| Heterogeneity: Tau² = 0.02; Chi² = 5.72, df = 3 (P = 0.13); I² = 48% | | | | |
| Test for overall effect: Z = 2.47 (P = 0.01) | | | | |
| Test for subgroup differences: Chi² = 0.37, df = 1 (P = 0.54), I² = 0% | | | | |

Hazard Ratio IV, Random, 95% CI

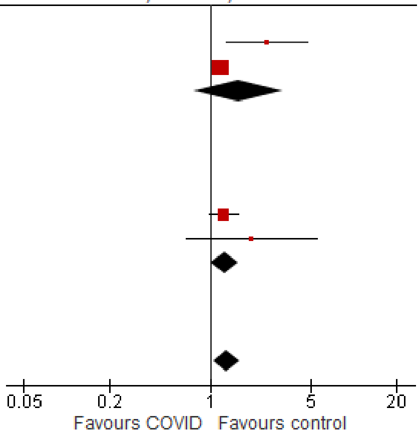

#### Outpatients/mixed, 18-64 y

| Study or Subgroup | log[Hazard Ratio] | SE | Weight | Hazard Ratio IV, Random, 95% CI |
|---|---|---|---|---|
| **5.7.1 Communication & motor disorders (outpatients/mixed, 18-64 y), <12 mo follow-up** | | | | |
| Gronkjaer 2023, 20-39 y | 1.191618 | 0.236124 | 10.8% | 3.29 [2.07, 5.23] |
| Gronkjaer 2023, 40-59 y | 2.123265 | 0.136175 | 11.1% | 8.36 [6.40, 10.92] |
| Gronkjaer 2023, 60-79 y | 1.290414 | 0.216985 | 10.9% | 3.63 [2.38, 5.56] |
| Nielsen 2024, 18-39 y | -0.36817 | 0.323975 | 10.4% | 0.69 [0.37, 1.31] |
| Nielsen 2024, 40-64 y | -0.06253 | 0.154557 | 11.1% | 0.94 [0.69, 1.27] |
| Zang 2023, INSIGHT cohort | -0.01228 | 0.130172 | 11.1% | 0.99 [0.77, 1.27] |
| **Subtotal (95% CI)** | | | **65.4%** | **2.02 [0.85, 4.80]** |
| Heterogeneity: Tau² = 1.13; Chi² = 185.17, df = 5 (P < 0.00001); I² = 97% | | | | |
| Test for overall effect: Z = 1.59 (P = 0.11) | | | | |
| **5.7.2 Communication & motor disorders (outpatients/mixed, 18-64 y), 12+ mo follow-up** | | | | |
| Taquet 2022 | -1.17054 | 0.051588 | 11.3% | 0.31 [0.28, 0.34] |
| Xu 2022 | 0.364643 | 0.037139 | 11.3% | 1.44 [1.34, 1.55] |
| Zarifkar 2022, 18-39 y | -0.31732 | 1.496508 | 3.8% | 0.73 [0.04, 13.68] |
| Zarifkar 2022, 40-59 y | 0.375821 | 0.911733 | 6.6% | 1.46 [0.24, 8.70] |
| Zarifkar 2022, 60-79 y | -0.03695 | 2.615193 | 1.6% | 0.96 [0.01, 162.18] |
| **Subtotal (95% CI)** | | | **34.6%** | **0.80 [0.24, 2.66]** |
| Heterogeneity: Tau² = 1.16; Chi² = 583.63, df = 4 (P < 0.00001); I² = 99% | | | | |
| Test for overall effect: Z = 0.37 (P = 0.71) | | | | |
| **Total (95% CI)** | | | **100.0%** | **1.46 [0.72, 2.96]** |
| Heterogeneity: Tau² = 1.15; Chi² = 916.60, df = 10 (P < 0.00001); I² = 99% | | | | |
| Test for overall effect: Z = 1.05 (P = 0.29) | | | | |
| Test for subgroup differences: Chi² = 1.50, df = 1 (P = 0.22), I² = 33.4% | | | | |

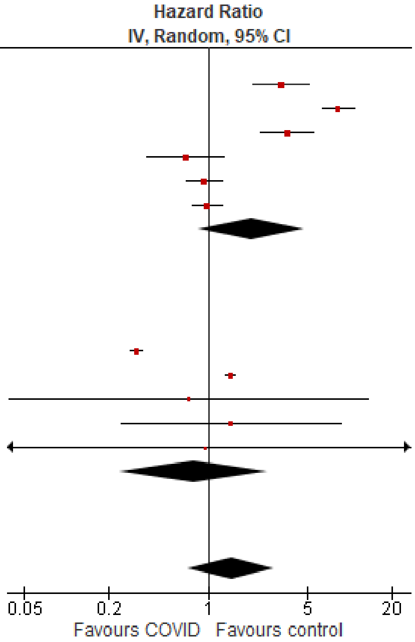

#### Outpatients/mixed, ≥65 y

| Study or Subgroup | log[Hazard Ratio] | SE | Weight | Hazard Ratio IV, Random, 95% CI |
|---|---|---|---|---|
| **5.8.1 Communication & motor disorders (outpatients/mixed, ≥65 y), <12 mo follow-up** | | | | |
| Gronkjaer 2023, 60-79 y | 1.673351 | 0.324743 | 10.4% | 5.33 [2.82, 10.07] |
| Mizrahi 2023, 60+ y | -0.08154 | 0.441947 | 7.2% | 0.92 [0.39, 2.19] |
| Nielsen 2024 | 0.053452 | 0.149542 | 17.7% | 1.05 [0.79, 1.41] |
| Zang 2023, INSIGHT cohort | 0.267505 | 0.200016 | 15.4% | 1.31 [0.88, 1.93] |
| **Subtotal (95% CI)** | | | **50.7%** | **1.60 [0.84, 3.05]** |
| Heterogeneity: Tau² = 0.35; Chi² = 21.37, df = 3 (P < 0.0001); I² = 86% | | | | |
| Test for overall effect: Z = 1.42 (P = 0.15) | | | | |
| **5.8.2 Communication & motor disorders (outpatients/mixed, ≥65 y), 12+ mo follow-up** | | | | |
| Taquet 2022 | -0.08921 | 0.028377 | 21.8% | 0.91 [0.87, 0.97] |
| Xu 2022 | 0.314811 | 0.050255 | 21.4% | 1.37 [1.24, 1.51] |
| Zarifkar 2022, 60-79 y | -0.03697 | 1.120579 | 1.5% | 0.96 [0.11, 8.67] |
| Zarifkar 2022, 80+ y | 0.173882 | 0.592904 | 4.6% | 1.19 [0.37, 3.80] |
| **Subtotal (95% CI)** | | | **49.3%** | **1.12 [0.78, 1.61]** |
| Heterogeneity: Tau² = 0.08; Chi² = 49.09, df = 3 (P < 0.00001); I² = 94% | | | | |
| Test for overall effect: Z = 0.60 (P = 0.55) | | | | |
| **Total (95% CI)** | | | **100.0%** | **1.31 [0.99, 1.74]** |
| Heterogeneity: Tau² = 0.09; Chi² = 76.84, df = 7 (P < 0.00001); I² = 91% | | | | |
| Test for overall effect: Z = 1.90 (P = 0.06) | | | | |
| Test for subgroup differences: Chi² = 0.89, df = 1 (P = 0.35), I² = 0% | | | | |

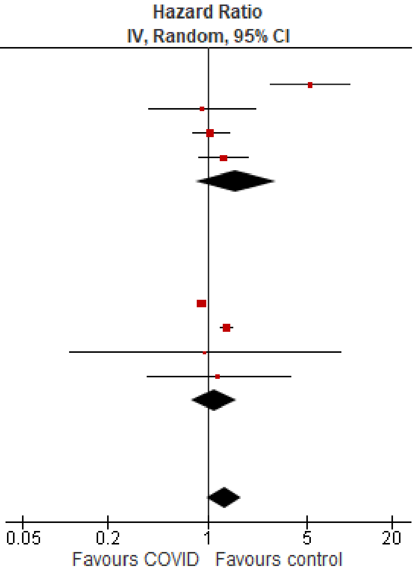

#### *Neurological disorders, dementia/mild cognitive impairment*

##### Outpatients/mixed, 18-64 y

| Study or Subgroup | log[Hazard Ratio] | SE | Weight | Hazard Ratio IV, Random, 95% CI |
|---|---|---|---|---|
| **5.9.1 Dementia/mild cognitive impairment (outpatients/mixed, 18-64 y), <12 mo follow-up** | | | | |
| Daugherty 2021 | 1.019158 | 0.540074 | 4.7% | 2.77 [0.96, 7.99] |
| Nielsen 2024, 18-39 y | 0.24686 | 0.486549 | 5.3% | 1.28 [0.49, 3.32] |
| Nielsen 2024, 40-64 y | -0.41552 | 0.130313 | 10.2% | 0.66 [0.51, 0.85] |
| Zang 2023, INSIGHT cohort | 0.435024 | 0.122723 | 10.3% | 1.55 [1.21, 1.97] |
| Zang 2023, OneFlorida+ cohort | -0.35667 | 0.1281 | 10.3% | 0.70 [0.54, 0.90] |
| **Subtotal (95% CI)** | | | **40.8%** | **1.07 [0.66, 1.72]** |
| Heterogeneity: Tau² = 0.22; Chi² = 33.46, df = 4 (P < 0.00001); I² = 88% | | | | |
| Test for overall effect: Z = 0.27 (P = 0.78) | | | | |
| **5.9.2 Dementia/mild cognitive impairment (outpatients/mixed, 18-64 y), 12+ mo follow-up** | | | | |
| Donnachie 2022, 18-39y | 0.773205 | 0.131887 | 10.2% | 2.17 [1.67, 2.81] |
| Donnachie 2022, 40-59y | 0.951388 | 0.078656 | 10.7% | 2.59 [2.22, 3.02] |
| Donnachie 2022, 60+ y | 0.166107 | 0.152318 | 10.0% | 1.18 [0.88, 1.59] |
| Taquet 2022 | -0.09519 | 0.033674 | 11.0% | 0.91 [0.85, 0.97] |
| Xu 2022 | 0.565314 | 0.030388 | 11.0% | 1.76 [1.66, 1.87] |
| Zarifkar 2022, 18-39 y | 3.049951 | 1.999941 | 0.6% | 21.11 [0.42, 1063.97] |
| Zarifkar 2022, 40-59 y | 1.844352 | 1.632594 | 0.8% | 6.32 [0.26, 155.12] |
| Zarifkar 2022, 60-79 y | 0.832909 | 0.511689 | 5.0% | 2.30 [0.84, 6.27] |
| **Subtotal (95% CI)** | | | **59.2%** | **1.73 [1.16, 2.56]** |
| Heterogeneity: Tau² = 0.22; Chi² = 297.93, df = 7 (P < 0.00001); I² = 98% | | | | |
| Test for overall effect: Z = 2.72 (P = 0.007) | | | | |
| **Total (95% CI)** | | | **100.0%** | **1.42 [1.05, 1.92]** |
| Heterogeneity: Tau² = 0.22; Chi² = 359.48, df = 12 (P < 0.00001); I² = 97% | | | | |
| Test for overall effect: Z = 2.26 (P = 0.02) | | | | |
| Test for subgroup differences: Chi² = 2.30, df = 1 (P = 0.13), I² = 56.6% | | | | |

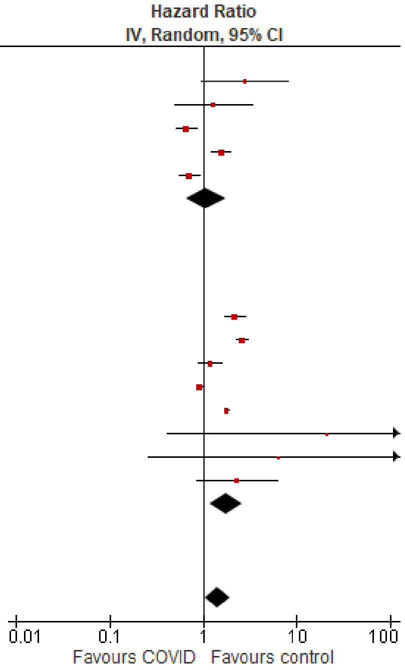

##### Outpatients/mixed, ≥65 y

| Study or Subgroup | log[Hazard Ratio] | SE | Weight | Hazard Ratio IV, Random, 95% CI |
|---|---|---|---|---|
| **5.10.1 Dementia/mild cognitive impairment (outpatients/mixed, ≥65 y), <12 mo follow-up** | | | | |
| Cohen 2022 | 0.81978 | 0.081838 | 7.8% | 2.27 [1.93, 2.66] |
| Murata 2024, alpha variant | -0.31471 | 1.389474 | 0.3% | 0.73 [0.05, 11.12] |
| Murata 2024, delta variant | 1.811562 | 0.422699 | 2.2% | 6.12 [2.67, 14.01] |
| Murata 2024, omicron BA.1/BA.2 variant | 0.71295 | 0.200632 | 5.3% | 2.04 [1.38, 3.02] |
| Murata 2024, omicron BA.5 variant | 0.029559 | 0.183813 | 5.6% | 1.03 [0.72, 1.48] |
| Nielsen 2024 | -0.31471 | 0.049074 | 8.4% | 0.73 [0.66, 0.80] |
| Wan 2023a, UK cohort | 0.09531 | 0.240453 | 4.5% | 1.10 [0.69, 1.76] |
| Zang 2023, INSIGHT cohort | 0.360747 | 0.053668 | 8.3% | 1.43 [1.29, 1.59] |
| Zang 2023, OneFlorida+ cohort | 0.352838 | 0.065038 | 8.1% | 1.42 [1.25, 1.62] |
| **Subtotal (95% CI)** | | | **50.5%** | **1.50 [1.06, 2.10]** |
| Heterogeneity: Tau² = 0.21; Chi² = 203.94, df = 8 (P < 0.00001); I² = 96% | | | | |
| Test for overall effect: Z = 2.32 (P = 0.02) | | | | |
| **5.10.2 Dementia/mild cognitive impairment (outpatients/mixed, ≥65 y), 12+ mo follow-up** | | | | |
| Donnachie 2022, 60+ y | 0.166107 | 0.060209 | 8.2% | 1.18 [1.05, 1.33] |
| Gollop 2023 | 0.029559 | 0.106419 | 7.3% | 1.03 [0.84, 1.27] |
| Taquet 2022 | 0.289231 | 0.014765 | 8.7% | 1.34 [1.30, 1.37] |
| Wang 2022a | 0.524729 | 0.029861 | 8.6% | 1.69 [1.59, 1.79] |
| Xu 2022 | 0.512824 | 0.048739 | 8.4% | 1.67 [1.52, 1.84] |
| Zarifkar 2022, 60-79 y | 0.832909 | 0.334979 | 3.1% | 2.30 [1.19, 4.43] |
| Zarifkar 2022, 80+ y | 0.262364 | 0.203701 | 5.2% | 1.30 [0.87, 1.94] |
| **Subtotal (95% CI)** | | | **49.5%** | **1.40 [1.22, 1.61]** |
| Heterogeneity: Tau² = 0.02; Chi² = 81.73, df = 6 (P < 0.00001); I² = 93% | | | | |
| Test for overall effect: Z = 4.78 (P < 0.00001) | | | | |
| **Total (95% CI)** | | | **100.0%** | **1.42 [1.23, 1.63]** |
| Heterogeneity: Tau² = 0.06; Chi² = 308.26, df = 15 (P < 0.00001); I² = 95% | | | | |
| Test for overall effect: Z = 4.76 (P < 0.00001) | | | | |
| Test for subgroup differences: Chi² = 0.12, df = 1 (P = 0.72), I² = 0% | | | | |

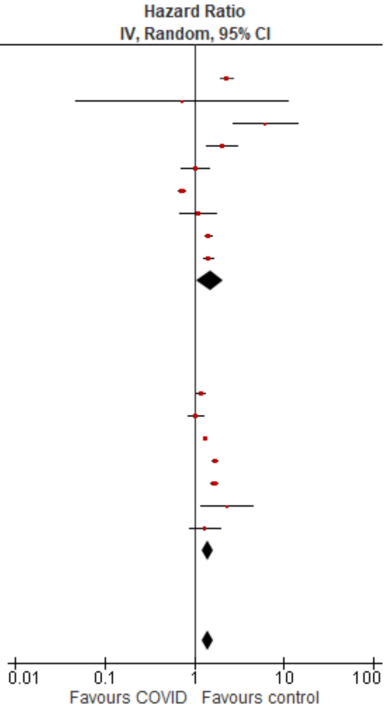

#### *Neurological disorders, epilepsy*

##### Outpatients/mixed, <18 y

| Study or Subgroup | log[Hazard Ratio] | SE | Weight | Hazard Ratio IV, Random, 95% CI |
|---|---|---|---|---|
| **5.13.1 Epilepsy (outpatients/mixed, <18 y), <12 mo follow-up** | | | | |
| Gronkjaer 2023 | -0.28768 | 0.147913 | 18.9% | 0.75 [0.56, 1.00] |
| Mizrahi 2023, 0-4 y | 0.866777 | 0.464457 | 13.4% | 2.38 [0.96, 5.91] |
| Mizrahi 2023, 12-18 y | -0.22314 | 0.50477 | 12.6% | 0.80 [0.30, 2.15] |
| Mizrahi 2023, 5-11 y | -0.33165 | 0.313322 | 16.3% | 0.72 [0.39, 1.33] |
| **Subtotal (95% CI)** | | | **61.2%** | **0.90 [0.58, 1.40]** |
| Heterogeneity: Tau² = 0.09; Chi² = 5.80, df = 3 (P = 0.12); I² = 48% | | | | |
| Test for overall effect: Z = 0.46 (P = 0.65) | | | | |
| **5.13.2 Epilepsy (outpatients/mixed, <18 y), 12+ mo follow-up** | | | | |
| Taquet 2022 | 0.736015 | 0.025306 | 19.8% | 2.09 [1.99, 2.19] |
| Westman 2022 | -0.22314 | 0.143126 | 19.0% | 0.80 [0.60, 1.06] |
| **Subtotal (95% CI)** | | | **38.8%** | **1.31 [0.51, 3.34]** |
| Heterogeneity: Tau² = 0.45; Chi² = 43.55, df = 1 (P < 0.00001); I² = 98% | | | | |
| Test for overall effect: Z = 0.56 (P = 0.58) | | | | |
| **Total (95% CI)** | | | **100.0%** | **1.09 [0.61, 1.95]** |
| Heterogeneity: Tau² = 0.45; Chi² = 101.19, df = 5 (P < 0.00001); I² = 95% | | | | |
| Test for overall effect: Z = 0.28 (P = 0.78) | | | | |
| Test for subgroup differences: Chi² = 0.49, df = 1 (P = 0.48), I² = 0% | | | | |

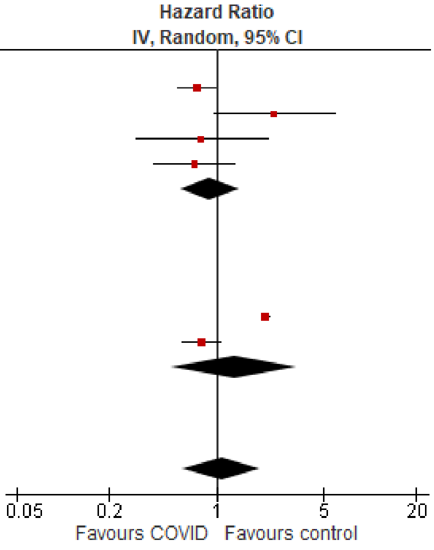

##### Outpatients/mixed, 18-64 y

| Study or Subgroup | log[Hazard Ratio] | SE | Weight | Hazard Ratio IV, Random, 95% CI |
|---|---|---|---|---|
| **5.14.1 Epilepsy (outpatients/mixed, 18-64 y), <12 mo follow-up** | | | | |
| Daugherty 2021 | 0.5988365 | 0.178809 | 7.0% | 1.82 [1.28, 2.58] |
| Gronkjaer 2023, 20-39 y | -0.19845 | 0.207918 | 5.6% | 0.82 [0.55, 1.23] |
| Gronkjaer 2023, 40-59 y | -0.16252 | 0.191771 | 6.3% | 0.85 [0.58, 1.24] |
| Gronkjaer 2023, 60-79 y | 0.058269 | 0.312814 | 2.8% | 1.06 [0.57, 1.96] |
| Mizrahi 2023, 19-40 y | 0.417722 | 0.389851 | 1.9% | 1.52 [0.71, 3.26] |
| Mizrahi 2023, 41-60 y | -0.3285 | 0.540475 | 1.0% | 0.72 [0.25, 2.08] |
| Mizrahi 2023, 60+ y | -0.60666 | 1.260595 | 0.2% | 0.55 [0.05, 6.45] |
| **Subtotal (95% CI)** | | | **24.8%** | **1.08 [0.78, 1.51]** |
| Heterogeneity: Tau² = 0.09; Chi² = 13.33, df = 6 (P = 0.04); I² = 55% | | | | |
| Test for overall effect: Z = 0.48 (P = 0.63) | | | | |
| **5.14.2 Epilepsy (outpatients/mixed, 18-64 y), 12+ mo follow-up** | | | | |
| Taquet 2022 | 0.263517 | 0.012367 | 25.7% | 1.30 [1.27, 1.33] |
| Westman 2022, 21-40 y | -0.52763274 | 0.124706 | 11.2% | 0.59 [0.46, 0.75] |
| Westman 2022, 41-60 y | -0.43078292 | 0.113063 | 12.5% | 0.65 [0.52, 0.81] |
| Westman 2022, 61-80y | 0.263517 | 0.01223 | 25.7% | 1.30 [1.27, 1.33] |
| **Subtotal (95% CI)** | | | **75.2%** | **1.04 [0.92, 1.17]** |
| Heterogeneity: Tau² = 0.01; Chi² = 77.13, df = 3 (P < 0.00001); I² = 96% | | | | |
| Test for overall effect: Z = 0.60 (P = 0.55) | | | | |
| **Total (95% CI)** | | | **100.0%** | **1.05 [0.94, 1.17]** |
| Heterogeneity: Tau² = 0.01; Chi² = 92.53, df = 10 (P < 0.00001); I² = 89% | | | | |
| Test for overall effect: Z = 0.86 (P = 0.39) | | | | |
| Test for subgroup differences: Chi² = 0.06, df = 1 (P = 0.80), I² = 0% | | | | |

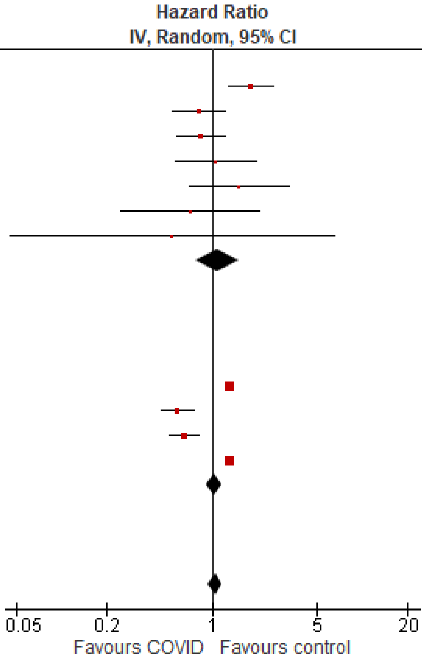

#### Outpatients/mixed, ≥65 y

| Study or Subgroup | log[Hazard Ratio] | SE | Weight | Hazard Ratio IV, Random, 95% CI |
|---|---|---|---|---|
| **5.15.1 Epilepsy (outpatients/mixed, ≥65 y), <12 mo follow-up** | | | | |
| Cohen 2022 | 0.774727 | 0.159395 | 15.3% | 2.17 [1.59, 2.97] |
| Gronkjaer 2023, 60-79 y | 0.058269 | 0.186942 | 13.3% | 1.06 [0.73, 1.53] |
| Gronkjaer 2023, 80+ y | 0.09531 | 0.245479 | 9.9% | 1.10 [0.68, 1.78] |
| Mizrahi 2023, 60+ y | -0.60666 | 0.550169 | 2.9% | 0.55 [0.19, 1.60] |
| **Subtotal (95% CI)** | | | **41.4%** | **1.22 [0.74, 2.01]** |
| Heterogeneity: Tau² = 0.18; Chi² = 13.62, df = 3 (P = 0.003); I² = 78% | | | | |
| Test for overall effect: Z = 0.79 (P = 0.43) | | | | |
| **5.15.2 Epilepsy (outpatients/mixed, ≥65 y), 12+ mo follow-up** | | | | |
| Taquet 2022 | 0.10084 | 0.020774 | 24.8% | 1.11 [1.06, 1.15] |
| Westman 2022, 100+ y | 0.004689 | 1.997988 | 0.2% | 1.00 [0.02, 50.43] |
| Westman 2022, 61-80y | 0.24686 | 0.118502 | 18.5% | 1.28 [1.01, 1.61] |
| Westman 2022, 81-100 y | 0.29267 | 0.163323 | 15.0% | 1.34 [0.97, 1.85] |
| **Subtotal (95% CI)** | | | **58.6%** | **1.11 [1.07, 1.16]** |
| Heterogeneity: Tau² = 0.00; Chi² = 2.77, df = 3 (P = 0.43); I² = 0% | | | | |
| Test for overall effect: Z = 5.32 (P < 0.00001) | | | | |
| **Total (95% CI)** | | | **100.0%** | **1.26 [1.04, 1.53]** |
| Heterogeneity: Tau² = 0.04; Chi² = 21.83, df = 7 (P = 0.003); I² = 68% | | | | |
| Test for overall effect: Z = 2.33 (P = 0.02) | | | | |
| Test for subgroup differences: Chi² = 0.13, df = 1 (P = 0.72), I² = 0% | | | | |

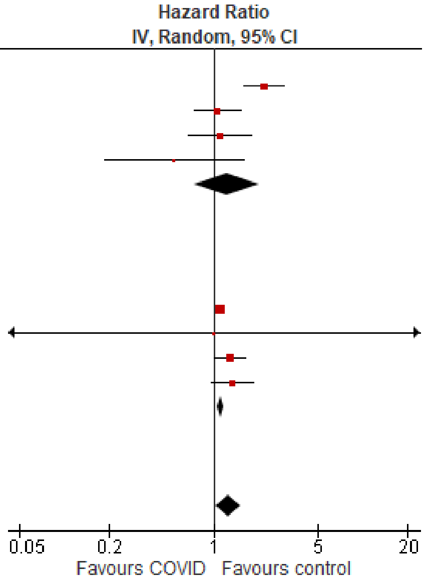

*Neurological disorders, Guillain-Barre syndrome*

#### Outpatients/mixed, 18-64 y

| Study or Subgroup | log[Hazard Ratio] | SE | Weight | Hazard Ratio IV, Random, 95% CI |
|---|---|---|---|---|
| **5.16.1 Guillain-Barré syndrome (outpatients/mixed, 18-64 y), <12 mo follow-up** | | | | |
| Daugherty 2021 | 1.211941 | 0.78529 | 0.7% | 3.36 [0.72, 15.66] |
| Mizrahi 2023, 41-60 y | 0.076961 | 0.313754 | 4.1% | 1.08 [0.58, 2.00] |
| **Subtotal (95% CI)** | | | **4.8%** | **1.52 [0.55, 4.20]** |
| Heterogeneity: Tau² = 0.29; Chi² = 1.80, df = 1 (P = 0.18); I² = 44% | | | | |
| Test for overall effect: Z = 0.80 (P = 0.42) | | | | |
| **5.16.2 Guillain-Barré syndrome (outpatients/mixed, 18-64 y), 12+ mo follow-up** | | | | |
| Taquet 2022 | 0.161523 | 0.065576 | 94.4% | 1.18 [1.03, 1.34] |
| Zarifkar 2022, 18-39 y | -0.31732 | 1.00442 | 0.4% | 0.73 [0.10, 5.21] |
| Zarifkar 2022, 40-59 y | -0.72052 | 1.435453 | 0.2% | 0.49 [0.03, 8.11] |
| Zarifkar 2022, 60-79 y | 0.503741 | 1.336515 | 0.2% | 1.65 [0.12, 22.72] |
| **Subtotal (95% CI)** | | | **95.2%** | **1.17 [1.03, 1.33]** |
| Heterogeneity: Tau² = 0.00; Chi² = 0.67, df = 3 (P = 0.88); I² = 0% | | | | |
| Test for overall effect: Z = 2.43 (P = 0.02) | | | | |
| **Total (95% CI)** | | | **100.0%** | **1.18 [1.04, 1.33]** |
| Heterogeneity: Tau² = 0.00; Chi² = 2.53, df = 5 (P = 0.77); I² = 0% | | | | |
| Test for overall effect: Z = 2.54 (P = 0.01) | | | | |
| Test for subgroup differences: Chi² = 0.24, df = 1 (P = 0.62), I² = 0% | | | | |

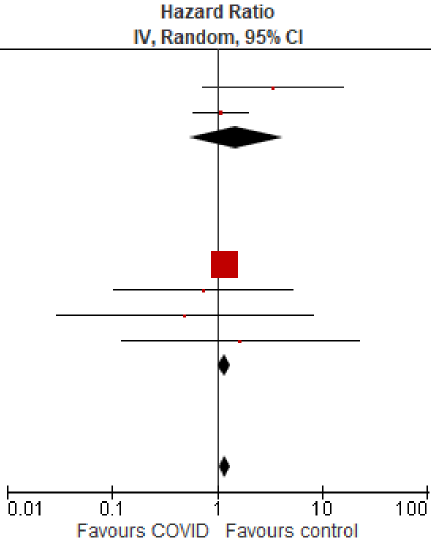

#### *Neurological disorders, nerve disorders*

##### Outpatients/mixed, <18 y

| Study or Subgroup | log[Hazard Ratio] | SE | Weight | Hazard Ratio IV, Random, 95% CI |
|---|---|---|---|---|
| **5.22.1 Nerve disorders (outpatients/mixed, <18 y), <12 mo follow-up** | | | | |
| Gronkjaer 2023 | -0.2627 | 0.22258 | 26.1% | 0.77 [0.50, 1.19] |
| Nielsen 2024 | 0.113329 | 0.160774 | 31.3% | 1.12 [0.82, 1.53] |
| **Subtotal (95% CI)** | | | **57.4%** | **0.96 [0.67, 1.38]** |
| Heterogeneity: Tau² = 0.03; Chi² = 1.88, df = 1 (P = 0.17); I² = 47% | | | | |
| Test for overall effect: Z = 0.23 (P = 0.82) | | | | |
| **5.22.2 Nerve disorders (outpatients/mixed, <18 y), 12+ mo follow-up** | | | | |
| Roessler 2022 | -0.79851 | 0.925426 | 3.9% | 0.45 [0.07, 2.76] |
| Taquet 2022 | 0.417328 | 0.058337 | 38.7% | 1.52 [1.35, 1.70] |
| **Subtotal (95% CI)** | | | **42.6%** | **1.17 [0.44, 3.11]** |
| Heterogeneity: Tau² = 0.31; Chi² = 1.72, df = 1 (P = 0.19); I² = 42% | | | | |
| Test for overall effect: Z = 0.32 (P = 0.75) | | | | |
| **Total (95% CI)** | | | **100.0%** | **1.10 [0.76, 1.61]** |
| Heterogeneity: Tau² = 0.09; Chi² = 12.59, df = 3 (P = 0.006); I² = 76% | | | | |
| Test for overall effect: Z = 0.51 (P = 0.61) | | | | |
| Test for subgroup differences: Chi² = 0.15, df = 1 (P = 0.70), I² = 0% | | | | |

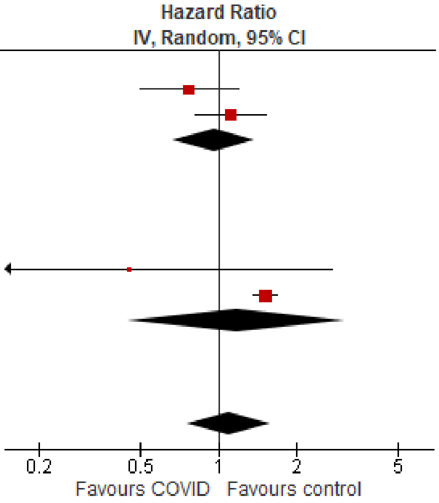

##### Outpatients/mixed, 18-64 y

| Study or Subgroup | log[Hazard Ratio] | SE | Weight | Hazard Ratio IV, Random, 95% CI |
|---|---|---|---|---|
| **5.23.1 Nerve disorders (outpatients/mixed, 18-64 y), <12 mo follow-up** | | | | |
| Daugherty 2021 | 0.71784 | 0.130313 | 7.5% | 2.05 [1.59, 2.65] |
| Gronkjaer 2023, 20-39 y | -0.20239 | 0.108552 | 8.7% | 0.82 [0.66, 1.01] |
| Gronkjaer 2023, 40-59 y | -0.01541 | 0.067573 | 11.2% | 0.98 [0.86, 1.12] |
| Gronkjaer 2023, 60-79 y | -0.1201 | 0.083145 | 10.3% | 0.89 [0.75, 1.04] |
| Nielsen 2024, 18-39 y | 0.086178 | 0.046511 | 12.4% | 1.09 [1.00, 1.19] |
| Nielsen 2024, 40-64 y | 0.019803 | 0.030047 | 13.2% | 1.02 [0.96, 1.08] |
| Zang 2023, INSIGHT cohort | 0.057231 | 0.092442 | 9.7% | 1.06 [0.88, 1.27] |
| **Subtotal (95% CI)** | | | **73.0%** | **1.06 [0.94, 1.20]** |
| Heterogeneity: Tau² = 0.02; Chi² = 37.79, df = 6 (P < 0.00001); I² = 84% | | | | |
| Test for overall effect: Z = 0.90 (P = 0.37) | | | | |
| **5.23.2 Nerve disorders (outpatients/mixed, 18-64 y), 12+ mo follow-up** | | | | |
| Taquet 2022 | -0.00441 | 0.005842 | 13.7% | 1.00 [0.98, 1.01] |
| Xu 2022 | 0.277632 | 0.027082 | 13.3% | 1.32 [1.25, 1.39] |
| **Subtotal (95% CI)** | | | **27.0%** | **1.14 [0.87, 1.51]** |
| Heterogeneity: Tau² = 0.04; Chi² = 103.64, df = 1 (P < 0.00001); I² = 99% | | | | |
| Test for overall effect: Z = 0.96 (P = 0.34) | | | | |
| **Total (95% CI)** | | | **100.0%** | **1.08 [0.97, 1.20]** |
| Heterogeneity: Tau² = 0.02; Chi² = 142.72, df = 8 (P < 0.00001); I² = 94% | | | | |
| Test for overall effect: Z = 1.46 (P = 0.14) | | | | |
| Test for subgroup differences: Chi² = 0.26, df = 1 (P = 0.61), I² = 0% | | | | |

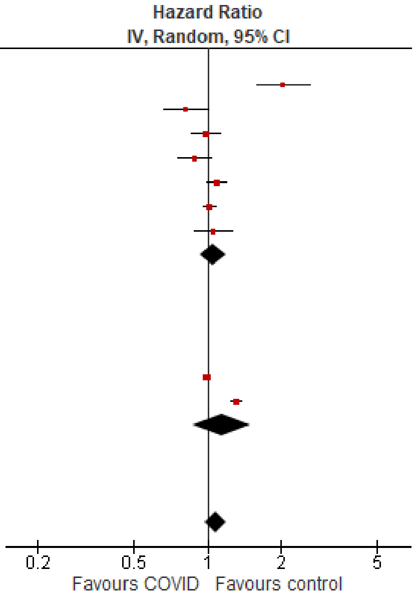

#### Outpatients/mixed, ≥65 y

| Study or Subgroup | log[Hazard Ratio] | SE | Weight | Hazard Ratio IV, Random, 95% CI |
|---|---|---|---|---|
| **5.24.1 Nerve disorders (outpatients/mixed, ≥65 y), <12 mo follow-up** | | | | |
| Cohen 2022 | 0.19062 | 0.175336 | 8.6% | 1.21 [0.86, 1.71] |
| Gronkjaer 2023, 60-79 y | -0.1201 | 0.099377 | 13.9% | 0.89 [0.73, 1.08] |
| Gronkjaer 2023, 80+ y | -0.45442 | 0.218488 | 6.6% | 0.63 [0.41, 0.97] |
| Nielsen 2024 | 0.165514 | 0.043342 | 18.1% | 1.18 [1.08, 1.28] |
| Zang 2023, INSIGHT cohort | 0.057231 | 0.089785 | 14.6% | 1.06 [0.89, 1.26] |
| **Subtotal (95% CI)** | | | **61.7%** | **1.01 [0.86, 1.20]** |
| Heterogeneity: $Tau^2 = 0.02$; $Chi^2 = 14.12$, df = 4 (P = 0.007); $I^2 = 72\%$ | | | | |
| Test for overall effect: Z = 0.16 (P = 0.87) | | | | |
| **5.24.2 Nerve disorders (outpatients/mixed, ≥65 y), 12+ mo follow-up** | | | | |
| Taquet 2022 | 0.008762 | 0.01024 | 19.4% | 1.01 [0.99, 1.03] |
| Xu 2022 | 0.270027 | 0.027289 | 18.9% | 1.31 [1.24, 1.38] |
| **Subtotal (95% CI)** | | | **38.3%** | **1.15 [0.89, 1.48]** |
| Heterogeneity: $Tau^2 = 0.03$; $Chi^2 = 80.35$, df = 1 (P < 0.00001); $I^2 = 99\%$ | | | | |
| Test for overall effect: Z = 1.06 (P = 0.29) | | | | |
| **Total (95% CI)** | | | **100.0%** | **1.06 [0.93, 1.22]** |
| Heterogeneity: $Tau^2 = 0.02$; $Chi^2 = 96.95$, df = 6 (P < 0.00001); $I^2 = 94\%$ | | | | |
| Test for overall effect: Z = 0.88 (P = 0.38) | | | | |
| Test for subgroup differences: $Chi^2 = 0.63$, df = 1 (P = 0.43), $I^2 = 0\%$ | | | | |

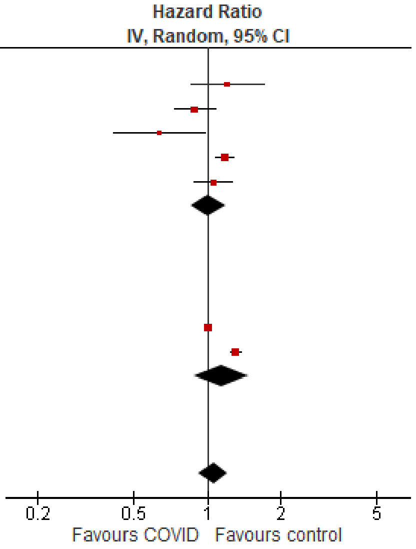

### *Respiratory disorders, overall*

#### Outpatients/mixed, 18-64 y

| Study or Subgroup | log[Hazard Ratio] | SE | Weight | Hazard Ratio IV, Random, 95% CI |
|---|---|---|---|---|
| **6.2.1 Repiratory disorders, overall (outpatients/mixed, 18-64 y), <12 mo follow-up** | | | | |
| Daugherty 2021 | 2.190555 | 0.254895 | 9.6% | 8.94 [5.42, 14.73] |
| Mizrahi 2023, 19-40 y | -0.4943 | 0.543782 | 4.7% | 0.61 [0.21, 1.77] |
| Mizrahi 2023, 41-60 y | 0.193874 | 0.247303 | 9.7% | 1.21 [0.75, 1.97] |
| Mizrahi 2023, 60+ y | -0.01137 | 1.620559 | 0.8% | 0.99 [0.04, 23.69] |
| Robertson 2024, BMI 20 to 25 | 0.038949 | 0.162655 | 11.6% | 1.04 [0.76, 1.43] |
| Zang 2023, INSIGHT cohort | 0.173953 | 0.069947 | 13.1% | 1.19 [1.04, 1.36] |
| **Subtotal (95% CI)** | | | **49.4%** | **1.56 [0.82, 2.97]** |
| Heterogeneity: $Tau^2 = 0.48$; $Chi^2 = 62.75$, df = 5 (P < 0.00001); $I^2 = 92\%$ | | | | |
| Test for overall effect: Z = 1.37 (P = 0.17) | | | | |
| **6.2.2 Repiratory disorders, overall (outpatients/mixed, 18-64 y), 12+ mo follow-up** | | | | |
| Meng 2024 | 0.410333 | 0.110206 | 12.5% | 1.51 [1.21, 1.87] |
| Oh 2024, Japan cohort, 20-39 y | 1.068153 | 0.039324 | 13.4% | 2.91 [2.69, 3.14] |
| Oh 2024, Japan cohort, 40-59 y | 0.947789 | 0.031681 | 13.4% | 2.58 [2.42, 2.75] |
| Oh 2024, Japan cohort, 60+ y | 0.727549 | 0.178137 | 11.2% | 2.07 [1.46, 2.93] |
| **Subtotal (95% CI)** | | | **50.6%** | **2.28 [1.87, 2.79]** |
| Heterogeneity: $Tau^2 = 0.03$; $Chi^2 = 34.28$, df = 3 (P < 0.00001); $I^2 = 91\%$ | | | | |
| Test for overall effect: Z = 8.08 (P < 0.00001) | | | | |
| **Total (95% CI)** | | | **100.0%** | **1.89 [1.42, 2.51]** |
| Heterogeneity: $Tau^2 = 0.16$; $Chi^2 = 215.47$, df = 9 (P < 0.00001); $I^2 = 96\%$ | | | | |
| Test for overall effect: Z = 4.37 (P < 0.0001) | | | | |
| Test for subgroup differences: $Chi^2 = 1.21$, df = 1 (P = 0.27), $I^2 = 17.4\%$ | | | | |

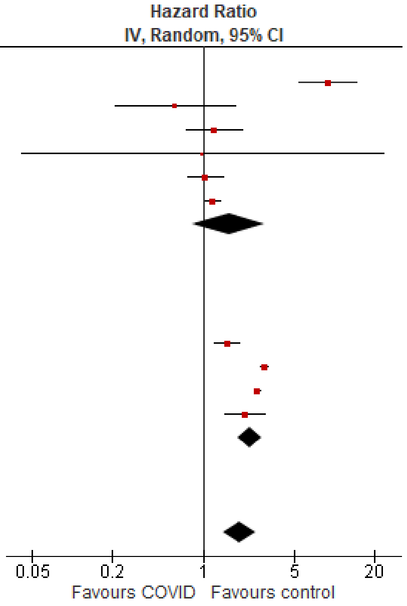

#### Outpatients/mixed, ≥65 y

| Study or Subgroup | log[Hazard Ratio] | SE | Weight | Hazard Ratio IV, Random, 95% CI |
|---|---|---|---|---|
| **6.3.1 Repiratory disorders, overall (outpatients/mixed, ≥65 y), <12 mo follow-up** | | | | |
| Cohen 2022 | 1.2947 | 0.108294 | 18.2% | 3.65 [2.95, 4.51] |
| Mizrahi 2023, 60+ y | 0.00878 | 0.387936 | 9.8% | 1.01 [0.47, 2.16] |
| Zang 2023, INSIGHT cohort | 0.280733 | 0.050088 | 19.3% | 1.32 [1.20, 1.46] |
| **Subtotal (95% CI)** | | | **47.4%** | **1.76 [0.78, 3.99]** |
| Heterogeneity: $Tau^2 = 0.47$; $Chi^2 = 73.55$, df = 2 (P < 0.00001); $I^2 = 97\%$ | | | | |
| Test for overall effect: Z = 1.37 (P = 0.17) | | | | |
| **6.3.2 Repiratory disorders, overall (outpatients/mixed, ≥65 y), 12+ mo follow-up** | | | | |
| Meng 2024 | 0.521952 | 0.097869 | 18.5% | 1.69 [1.39, 2.04] |
| Oh 2024, Japan cohort | 0.530628 | 0.206854 | 15.3% | 1.70 [1.13, 2.55] |
| Oh 2024, South Korea cohort | 0.727549 | 0.077745 | 18.9% | 2.07 [1.78, 2.41] |
| **Subtotal (95% CI)** | | | **52.6%** | **1.87 [1.60, 2.17]** |
| Heterogeneity: $Tau^2 = 0.01$; $Chi^2 = 3.00$, df = 2 (P = 0.22); $I^2 = 33\%$ | | | | |
| Test for overall effect: Z = 8.02 (P < 0.00001) | | | | |
| **Total (95% CI)** | | | **100.0%** | **1.83 [1.31, 2.56]** |
| Heterogeneity: $Tau^2 = 0.15$; $Chi^2 = 82.86$, df = 5 (P < 0.00001); $I^2 = 94\%$ | | | | |
| Test for overall effect: Z = 3.54 (P = 0.0004) | | | | |
| Test for subgroup differences: $Chi^2 = 0.02$, df = 1 (P = 0.89), $I^2 = 0\%$ | | | | |

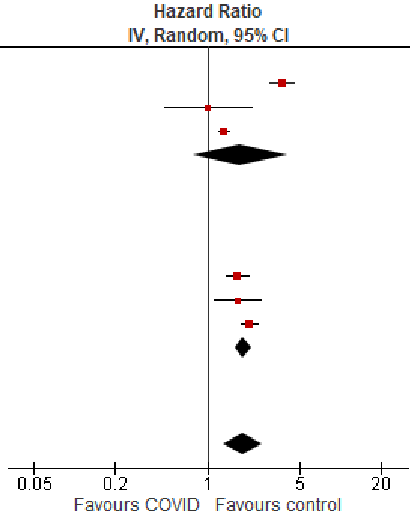

#### *Sleep disorders, overall*

#### Outpatients/mixed, <18 y

| Study or Subgroup | log[Hazard Ratio] | SE | Weight | Hazard Ratio IV, Random, 95% CI |
|---|---|---|---|---|
| **7.1.1 Sleep disorders, overall (outpatients/mixed, <18 y), <12 mo follow-up** | | | | |
| Mizrahi 2023, 0-4 y | 0.350657 | 0.446788 | 0.6% | 1.42 [0.59, 3.41] |
| Tartof 2022 | -0.09431 | 0.154972 | 4.0% | 0.91 [0.67, 1.23] |
| Valdivieso-Martinez 2023, unvaccinated cohort | 0.875469 | 0.53192 | 0.4% | 2.40 [0.85, 6.81] |
| Valdivieso-Martinez 2023, vaccinated cohort | 0 | 0.999776 | 0.1% | 1.00 [0.14, 7.10] |
| **Subtotal (95% CI)** | | | **5.1%** | **1.12 [0.75, 1.68]** |
| Heterogeneity: $Tau^2 = 0.04$; $Chi^2 = 3.68$, df = 3 (P = 0.30); $I^2 = 18\%$ | | | | |
| Test for overall effect: Z = 0.56 (P = 0.58) | | | | |
| **7.1.2 Sleep disorders, overall (outpatients/mixed, <18 y), 12+ mo follow-up** | | | | |
| Kompaniyets 2022, 12-17 y | -0.09431 | 0.013943 | 24.3% | 0.91 [0.89, 0.94] |
| Kompaniyets 2022, 2-4 y | -0.01005 | 0.033377 | 20.3% | 0.99 [0.93, 1.06] |
| Kompaniyets 2022, 5-11 y | -0.11653 | 0.017204 | 23.7% | 0.89 [0.86, 0.92] |
| Roessler 2022 | 0.398776 | 0.144739 | 4.5% | 1.49 [1.12, 1.98] |
| Taquet 2022 | 0.02557 | 0.026073 | 22.0% | 1.03 [0.97, 1.08] |
| **Subtotal (95% CI)** | | | **94.9%** | **0.97 [0.91, 1.04]** |
| Heterogeneity: $Tau^2 = 0.00$; $Chi^2 = 37.11$, df = 4 (P < 0.00001); $I^2 = 89\%$ | | | | |
| Test for overall effect: Z = 0.87 (P = 0.39) | | | | |
| **Total (95% CI)** | | | **100.0%** | **0.97 [0.91, 1.04]** |
| Heterogeneity: $Tau^2 = 0.00$; $Chi^2 = 41.24$, df = 8 (P < 0.00001); $I^2 = 81\%$ | | | | |
| Test for overall effect: Z = 0.80 (P = 0.42) | | | | |
| Test for subgroup differences: $Chi^2 = 0.48$, df = 1 (P = 0.49), $I^2 = 0\%$ | | | | |

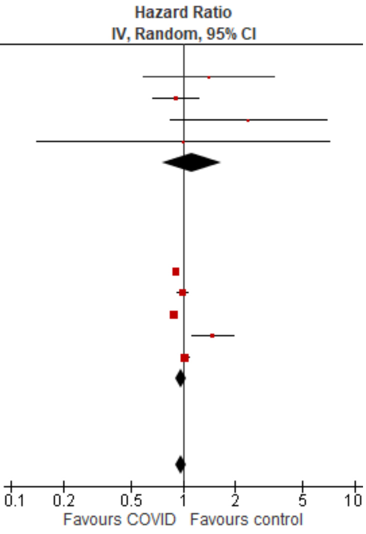

#### Outpatients/mixed, 18-64 y

| Study or Subgroup | log[Hazard Ratio] | SE | Weight | Hazard Ratio IV, Random, 95% CI |
|---|---|---|---|---|
| **7.2.1 Sleep disorders, overall (outpaient/mixed, 18-64 y), <12 mo follow-up** | | | | |
| Daugherty 2021 | 0.576613 | 0.062672 | 30.2% | 1.78 [1.57, 2.01] |
| Lin 2024 | 0.524729 | 0.013629 | 33.5% | 1.69 [1.65, 1.74] |
| Mizrahi 2023, 19-40 y | 0.487125 | 1.617151 | 0.4% | 1.63 [0.07, 38.73] |
| Mizrahi 2023, 41-60 y | 0.444686 | 1.581163 | 0.5% | 1.56 [0.07, 34.59] |
| Mizrahi 2023, 60+ y | -1.20397 | 1.620559 | 0.4% | 0.30 [0.01, 7.19] |
| Zang 2023, INSIGHT cohort | 0.218493 | 0.033683 | 32.6% | 1.24 [1.16, 1.33] |
| **Subtotal (95% CI)** | | | **97.5%** | **1.54 [1.24, 1.91]** |
| Heterogeneity: Tau² = 0.04; Chi² = 74.32, df = 5 (P < 0.00001); I² = 93% | | | | |
| Test for overall effect: Z = 3.89 (P = 0.0001) | | | | |
| **7.2.2 Sleep disorders, overall (outpaient/mixed, 18-64 y), 12+ mo follow-up** | | | | |
| Taquet 2022 | 0.030044 | 1.54256 | 0.5% | 1.03 [0.05, 21.19] |
| Zarifkar 2022, 18-39 y | -0.84188 | 1.42859 | 0.6% | 0.43 [0.03, 7.09] |
| Zarifkar 2022, 40-59 y | -0.27589 | 1.055883 | 1.0% | 0.76 [0.10, 6.01] |
| Zarifkar 2022, 60-79 y | 1.196767 | 1.617151 | 0.4% | 3.31 [0.14, 78.75] |
| **Subtotal (95% CI)** | | | **2.5%** | **0.92 [0.24, 3.45]** |
| Heterogeneity: Tau² = 0.00; Chi² = 0.95, df = 3 (P = 0.81); I² = 0% | | | | |
| Test for overall effect: Z = 0.13 (P = 0.90) | | | | |
| **Total (95% CI)** | | | **100.0%** | **1.52 [1.23, 1.87]** |
| Heterogeneity: Tau² = 0.03; Chi² = 75.99, df = 9 (P < 0.00001); I² = 88% | | | | |
| Test for overall effect: Z = 3.89 (P = 0.0001) | | | | |
| Test for subgroup differences: Chi² = 0.57, df = 1 (P = 0.45), I² = 0% | | | | |

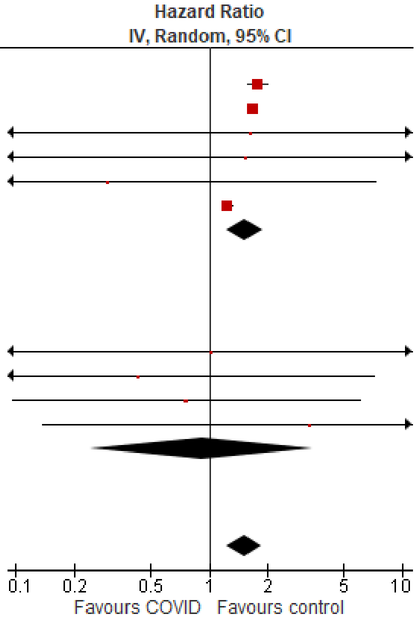

#### Outpatients/mixed, ≥65 y

| Study or Subgroup | log[Hazard Ratio] | SE | Weight | Hazard Ratio IV, Random, 95% CI |
|---|---|---|---|---|
| **7.3.1 Sleep disorders, overall (outpatients/mixed, ≥65 y), <12 mo follow-up** | | | | |
| Cohen 2022 | 0.405465 | 0.094435 | 13.4% | 1.50 [1.25, 1.80] |
| Lin 2024 | 0.405465 | 0.008533 | 16.4% | 1.50 [1.48, 1.53] |
| Mizrahi 2023, 60+ y | -1.20397 | 0.724736 | 1.2% | 0.30 [0.07, 1.24] |
| Murata 2024, alpha variant | -0.82098 | 1.002182 | 0.6% | 0.44 [0.06, 3.14] |
| Murata 2024, delta variant | -0.37106 | 0.166409 | 9.7% | 0.69 [0.50, 0.96] |
| Murata 2024, omicron BA.1/BA.2 variant | -0.54473 | 0.126888 | 11.7% | 0.58 [0.45, 0.74] |
| Murata 2024, omicron BA.5 variant | 0.322083 | 0.451043 | 2.7% | 1.38 [0.57, 3.34] |
| Wan 2023a, UK cohort | 0.336472 | 0.139424 | 11.1% | 1.40 [1.07, 1.84] |
| Zang 2023, INSIGHT cohort | 0.359142 | 0.046804 | 15.6% | 1.43 [1.31, 1.57] |
| **Subtotal (95% CI)** | | | **82.5%** | **1.12 [0.92, 1.36]** |
| Heterogeneity: Tau² = 0.05; Chi² = 84.60, df = 8 (P < 0.00001); I² = 91% | | | | |
| Test for overall effect: Z = 1.09 (P = 0.27) | | | | |
| **7.3.2 Sleep disorders, overall (outpatients/mixed, ≥65 y), 12+ mo follow-up** | | | | |
| Taquet 2022 | 0.14842 | 0.026413 | 16.2% | 1.16 [1.10, 1.22] |
| Zarifkar 2022, 60-79 y | 1.196767 | 0.933663 | 0.7% | 3.31 [0.53, 20.63] |
| Zarifkar 2022, 80+ y | 0.989169 | 0.979509 | 0.7% | 2.69 [0.39, 18.34] |
| **Subtotal (95% CI)** | | | **17.5%** | **1.16 [1.10, 1.22]** |
| Heterogeneity: Tau² = 0.00; Chi² = 1.99, df = 2 (P = 0.37); I² = 0% | | | | |
| Test for overall effect: Z = 5.68 (P < 0.00001) | | | | |
| **Total (95% CI)** | | | **100.0%** | **1.15 [0.98, 1.35]** |
| Heterogeneity: Tau² = 0.04; Chi² = 166.62, df = 11 (P < 0.00001); I² = 93% | | | | |
| Test for overall effect: Z = 1.76 (P = 0.08) | | | | |
| Test for subgroup differences: Chi² = 0.14, df = 1 (P = 0.71), I² = 0% | | | | |

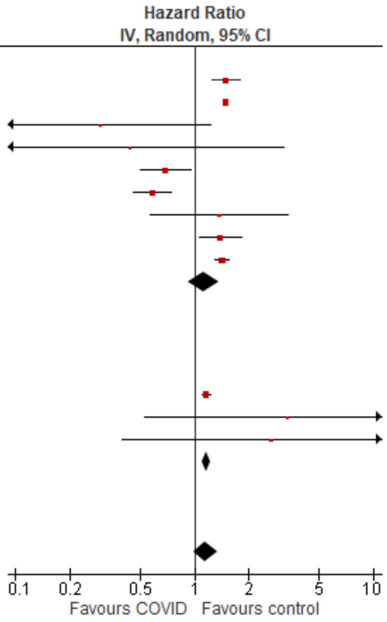

*Stroke, overall*

<u>Outpatients/mixed, <18 y</u>

| Study or Subgroup | log[Hazard Ratio] | SE | Weight | Hazard Ratio IV, Random, 95% CI |
|---|---|---|---|---|
| **8.1.1 Stroke, overall (outpatients/mixed, <18 y), <12 mo follow-up** | | | | |
| Nielsen 2024 | -0.10536 | 0.401708 | 2.9% | 0.90 [0.41, 1.98] |
| Tartof 2022 | -0.65393 | 0.740858 | 0.8% | 0.52 [0.12, 2.22] |
| **Subtotal (95% CI)** | | | **3.7%** | **0.79 [0.40, 1.59]** |
| Heterogeneity: Tau² = 0.00; Chi² = 0.42, df = 1 (P = 0.52); I² = 0% | | | | |
| Test for overall effect: Z = 0.65 (P = 0.51) | | | | |
| **8.1.2 Stroke, overall (outpatients/mixed, <18 y), 12+ mo follow-up** | | | | |
| Kompaniyets 2022 | 0.182322 | 0.094787 | 51.9% | 1.20 [1.00, 1.44] |
| Roessler 2022 | 0.223144 | 0.603648 | 1.3% | 1.25 [0.38, 4.08] |
| Taquet 2022 | 0.092397 | 0.104033 | 43.1% | 1.10 [0.89, 1.34] |
| **Subtotal (95% CI)** | | | **96.3%** | **1.15 [1.01, 1.32]** |
| Heterogeneity: Tau² = 0.00; Chi² = 0.43, df = 2 (P = 0.81); I² = 0% | | | | |
| Test for overall effect: Z = 2.05 (P = 0.04) | | | | |
| **Total (95% CI)** | | | **100.0%** | **1.14 [0.99, 1.30]** |
| Heterogeneity: Tau² = 0.00; Chi² = 1.92, df = 4 (P = 0.75); I² = 0% | | | | |
| Test for overall effect: Z = 1.88 (P = 0.06) | | | | |
| Test for subgroup differences: Chi² = 1.07, df = 1 (P = 0.30), I² = 6.7% | | | | |

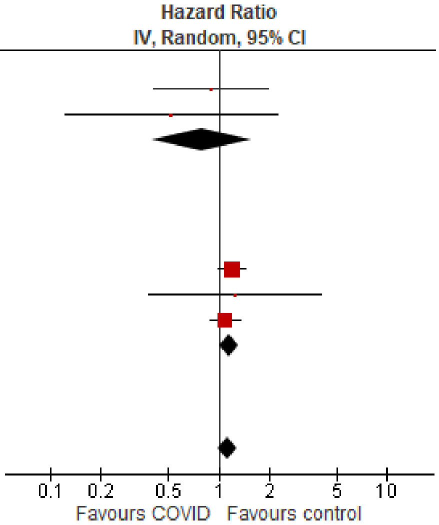

<u>Outpatients/mixed, 18-64 y</u>

| Study or Subgroup | log[Hazard Ratio] | SE | Weight | Hazard Ratio IV, Random, 95% CI |
|---|---|---|---|---|
| **8.2.1 Stroke, overall (outpatients/mixed, 18-64 y), <12 mo follow-up** | | | | |
| Batistoni 2024 | 0.494696 | 0.382586 | 2.8% | 1.64 [0.77, 3.47] |
| Daugherty 2021 | 0.900161 | 0.157049 | 5.5% | 2.46 [1.81, 3.35] |
| Lam 2023 | -0.24846 | 0.391448 | 2.7% | 0.78 [0.36, 1.68] |
| Lim 2023 | 0.48858 | 0.135964 | 5.8% | 1.63 [1.25, 2.13] |
| Mizrahi 2023, 19-40 y | -1.34707 | 0.772582 | 1.0% | 0.26 [0.06, 1.18] |
| Mizrahi 2023, 41-60 y | -0.04602 | 0.413093 | 2.5% | 0.96 [0.42, 2.15] |
| Mizrahi 2023, 60+ y | 0.198851 | 0.918382 | 0.7% | 1.22 [0.20, 7.38] |
| Nielsen 2024, 18-39 y | 0.19062 | 0.123234 | 6.0% | 1.21 [0.95, 1.54] |
| Nielsen 2024, 40-64 y | -0.21072 | 0.037399 | 6.8% | 0.81 [0.75, 0.87] |
| Rezel-Potts 2022 | -0.26487 | 0.118513 | 6.0% | 0.77 [0.61, 0.97] |
| Valasquez Garcia 2024 | 0.323025 | 0.121986 | 6.0% | 1.38 [1.09, 1.75] |
| **Subtotal (95% CI)** | | | **45.8%** | **1.16 [0.87, 1.54]** |
| Heterogeneity: Tau² = 0.16; Chi² = 91.96, df = 10 (P < 0.00001); I² = 89% | | | | |
| Test for overall effect: Z = 1.02 (P = 0.31) | | | | |
| **8.2.2 Stroke, overall (outpatients/mixed, 18-64 y), 12+ mo follow-up** | | | | |
| Taquet 2022 | -0.10536 | 0.014369 | 6.8% | 0.90 [0.88, 0.93] |
| Wang 2022b, 20-44y | 0.267734 | 0.064357 | 6.6% | 1.31 [1.15, 1.48] |
| Wang 2022b, 45-64y | 0.402126 | 0.039324 | 6.7% | 1.49 [1.38, 1.61] |
| Xie 2022b | 0.364643 | 0.054939 | 6.7% | 1.44 [1.29, 1.60] |
| Xu 2022 | 0.470004 | 0.035132 | 6.8% | 1.60 [1.49, 1.71] |
| Zappacosta 2022, 18-30 y | 1.530395 | 1.116326 | 0.5% | 4.62 [0.52, 41.20] |
| Zappacosta 2022, 31-40 y | 0.00995 | 0.537656 | 1.8% | 1.01 [0.35, 2.90] |
| Zappacosta 2022, 41-50 y | -0.30111 | 0.396695 | 2.7% | 0.74 [0.34, 1.61] |
| Zappacosta 2022, 51-60 y | 0.139762 | 0.252027 | 4.2% | 1.15 [0.70, 1.88] |
| Zappacosta 2022, 61-70 y | -0.10536 | 0.71835 | 1.1% | 0.90 [0.22, 3.68] |
| Zarifkar 2022, 18-39 y | -1.87015 | 1.003109 | 0.6% | 0.15 [0.02, 1.10] |
| Zarifkar 2022, 40-59 y | -0.69315 | 0.216147 | 4.7% | 0.50 [0.33, 0.76] |
| Zarifkar 2022, 60-79 y | -0.35667 | 0.188849 | 5.1% | 0.70 [0.48, 1.01] |
| **Subtotal (95% CI)** | | | **54.2%** | **1.06 [0.84, 1.34]** |
| Heterogeneity: Tau² = 0.12; Chi² = 406.63, df = 12 (P < 0.00001); I² = 97% | | | | |
| Test for overall effect: Z = 0.50 (P = 0.62) | | | | |
| **Total (95% CI)** | | | **100.0%** | **1.11 [0.95, 1.31]** |
| Heterogeneity: Tau² = 0.10; Chi² = 509.93, df = 23 (P < 0.00001); I² = 95% | | | | |
| Test for overall effect: Z = 1.30 (P = 0.19) | | | | |
| Test for subgroup differences: Chi² = 0.22, df = 1 (P = 0.64), I² = 0% | | | | |

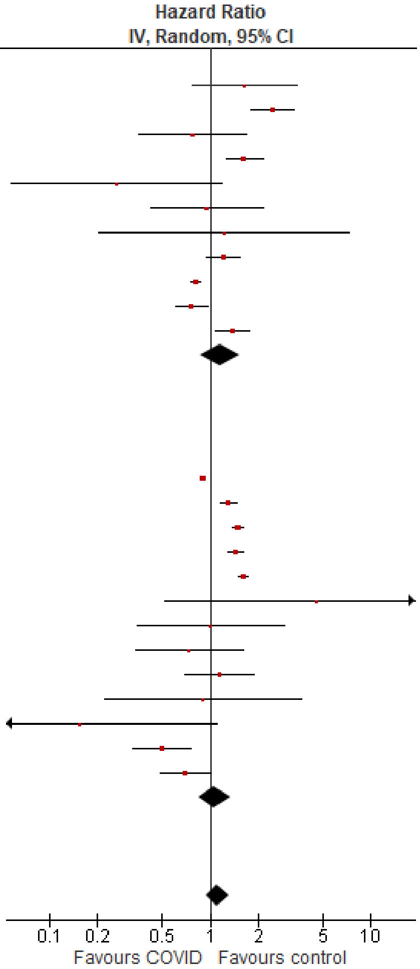

#### Outpatients/mixed, ≥65 y

| Study or Subgroup | log[Hazard Ratio] | SE | Weight | Hazard Ratio IV, Random, 95% CI |
|---|---|---|---|---|
| **8.3.1 Stroke, overall (outpatients/mixed, ≥65 y), <12 mo follow-up** | | | | |
| Batistoni 2024 | 0.463734 | 0.201786 | 6.1% | 1.59 [1.07, 2.36] |
| Cohen 2022 | 0.652325 | 0.076443 | 8.6% | 1.92 [1.65, 2.23] |
| Mizrahi 2023, 60+ y | 0.410713 | 0.918382 | 0.8% | 1.51 [0.25, 9.12] |
| Nielsen 2024 | -0.17435 | 0.033256 | 9.1% | 0.84 [0.79, 0.90] |
| Wan 2023a, UK cohort | 0.09531 | 0.197698 | 6.1% | 1.10 [0.75, 1.62] |
| Yang 2024 | 0.002934 | 0.051066 | 8.9% | 1.00 [0.91, 1.11] |
| **Subtotal (95% CI)** | | | **39.5%** | **1.22 [0.89, 1.68]** |
| Heterogeneity: Tau² = 0.12; Chi² = 105.40, df = 5 (P < 0.00001); I² = 95% | | | | |
| Test for overall effect: Z = 1.26 (P = 0.21) | | | | |
| **8.3.2 Stroke, overall (outpatients/mixed, ≥65 y), 12+ mo follow-up** | | | | |
| Taquet 2022 | 0.070552 | 0.014475 | 9.2% | 1.07 [1.04, 1.10] |
| Wang 2022b | 0.535908 | 0.033565 | 9.1% | 1.71 [1.60, 1.83] |
| Xie 2022b | 0.425268 | 0.031617 | 9.1% | 1.53 [1.44, 1.63] |
| Xu 2022 | 0.425268 | 0.058421 | 8.8% | 1.53 [1.36, 1.72] |
| Zappacosta 2022, 61-70 y | -0.10536 | 0.378604 | 3.3% | 0.90 [0.43, 1.89] |
| Zappacosta 2022, 70+ y | 0.398776 | 0.223997 | 5.6% | 1.49 [0.96, 2.31] |
| Zarifkar 2022, 60-79 y | -0.35667 | 0.119439 | 7.8% | 0.70 [0.55, 0.88] |
| Zarifkar 2022, 80+ y | -0.51083 | 0.119899 | 7.8% | 0.60 [0.47, 0.76] |
| **Subtotal (95% CI)** | | | **60.5%** | **1.15 [0.92, 1.42]** |
| Heterogeneity: Tau² = 0.08; Chi² = 302.21, df = 7 (P < 0.00001); I² = 98% | | | | |
| Test for overall effect: Z = 1.23 (P = 0.22) | | | | |
| **Total (95% CI)** | | | **100.0%** | **1.17 [0.99, 1.39]** |
| Heterogeneity: Tau² = 0.08; Chi² = 459.52, df = 13 (P < 0.00001); I² = 97% | | | | |
| Test for overall effect: Z = 1.88 (P = 0.06) | | | | |
| Test for subgroup differences: Chi² = 0.12, df = 1 (P = 0.73), I² = 0% | | | | |

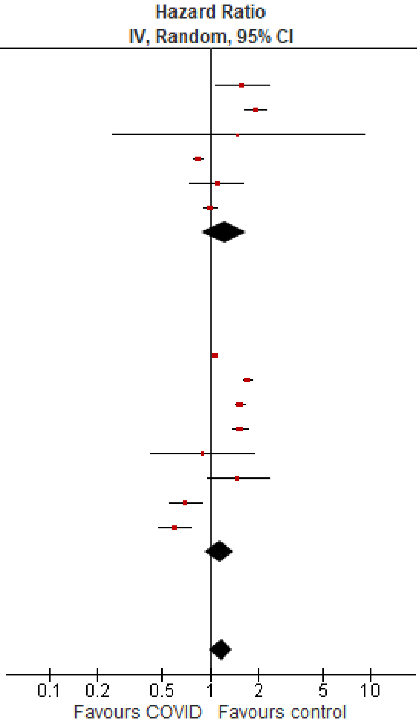

### *Stroke, ischemic stroke*

#### Outpatients/mixed, ≥65 y

| Study or Subgroup | log[Hazard Ratio] | SE | Weight | Hazard Ratio IV, Random, 95% CI |
|---|---|---|---|---|
| **8.7.1 Ischemic stroke (outpatients/mixed, ≥65 y), <12 mo follow-up** | | | | |
| Cohen 2022 | 0.652325 | 0.080672 | 20.2% | 1.92 [1.64, 2.25] |
| Yang 2024 | -0.01525 | 0.059333 | 20.9% | 0.98 [0.88, 1.11] |
| **Subtotal (95% CI)** | | | **41.1%** | **1.37 [0.71, 2.64]** |
| Heterogeneity: Tau² = 0.22; Chi² = 44.44, df = 1 (P < 0.00001); I² = 98% | | | | |
| Test for overall effect: Z = 0.95 (P = 0.34) | | | | |
| **8.7.3 Ischemic stroke (outpatients/mixed, ≥65 y), 12+ mo follow-up** | | | | |
| Taquet 2022 | 0.070552 | 0.014475 | 21.8% | 1.07 [1.04, 1.10] |
| Zarifkar 2022, 60-79 y | -0.51083 | 0.119439 | 18.6% | 0.60 [0.47, 0.76] |
| Zarifkar 2022, 80+ y | -0.35667 | 0.119899 | 18.5% | 0.70 [0.55, 0.89] |
| **Subtotal (95% CI)** | | | **58.9%** | **0.78 [0.52, 1.17]** |
| Heterogeneity: Tau² = 0.12; Chi² = 35.38, df = 2 (P < 0.00001); I² = 94% | | | | |
| Test for overall effect: Z = 1.21 (P = 0.23) | | | | |
| **Total (95% CI)** | | | **100.0%** | **0.98 [0.76, 1.28]** |
| Heterogeneity: Tau² = 0.08; Chi² = 90.48, df = 4 (P < 0.00001); I² = 96% | | | | |
| Test for overall effect: Z = 0.13 (P = 0.90) | | | | |
| Test for subgroup differences: Chi² = 2.09, df = 1 (P = 0.15), I² = 52.2% | | | | |

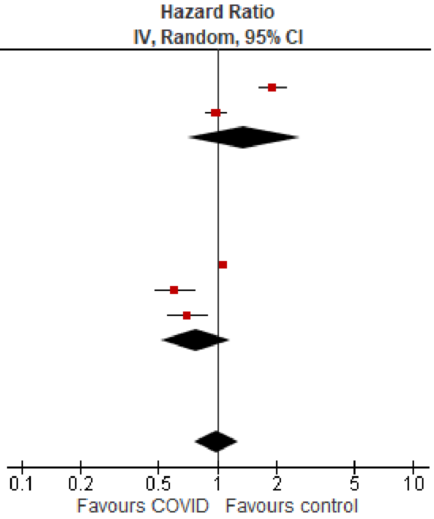
